# Risk of COVID-19 breakthrough infection and hospitalization in individuals with comorbidities

**DOI:** 10.1101/2022.04.26.22271727

**Authors:** Peter D Smits, Samuel Gratzl, Michael Simonov, Senthil K Nachimuthu, Brianna M Goodwin Cartwright, Michael D Wang, Charlotte Baker, Patricia Rodriguez, Mackenzie Bogiages, Benjamin M. Althouse, Nicholas L Stucky

## Abstract

**Background:** The successful development of multiple COVID-19 vaccines has led to a global vaccination effort to reduce severe COVID-19 infection and mortality. However, the effectiveness of the COVID-19 vaccines wane over time leading to breakthrough infections where vaccinated individuals experience a COVID-19 infection. Here we estimate the risks of break-through infection and subsequent hospitalization in individuals with common comorbidities who had completed an initial vaccination series.

**Methods:** Our study population included vaccinated patients between January 1, 2021 to March 31, 2022 who are present in the Truveta patient population. Models were developed to describe 1) time from completing primary vaccination series till breakthrough infection; and 2) if a patient was hospitalized within 14 days of breakthrough infection. We adjusted for age, race, ethnicity, sex, and year-month of vaccination.

**Results:** Of 1,192,135 patients in the Truveta Platform who had completed an initial vaccination sequence between January 1, 2021 and March 31, 2022, 2.84, 3.42, 2.76, and 2.89 percent of patients with CKD, chronic lung disease, diabetes, or are in an immunocompromised state experienced breakthrough infection, respectively, compared to 1.35 percent of the population without any of these four comorbidities. We found an increased risk of breakthrough infection and subsequent hospitalization in individuals with any of the four comorbidities when compared to individuals without these four comorbidities.

**Conclusions:** Vaccinated individuals with comorbidities experienced an increased risk of breakthrough COVID-19 infection and subsequent hospitalizations compared to the general population. Individuals with immunocompromising conditions and chronic lung disease were most at risk of breakthrough infection, while people with CKD were most at risk of hospitalization following breakthrough infection. Individuals with comorbidities should remain vigilant against infection even if vaccinated.

## 1 Introduction

The successful development of multiple COVID-19 vaccines has led to a global vaccination effort with the goal of reducing severe COVID-19 infection and mortality [1, 2]. The effectiveness of the COVID-19 vaccines, however, does wane over time and breakthrough infections have been reported since the beginning of the vaccination effort [3–10]. A breakthrough infection is defined as when an individual experiences a COVID-19 infection despite having completed their initial vaccination sequence (i.e., two doses plus an addition 2 weeks for an mRNA vaccine). This waning vaccine effectiveness, along with vaccine-variant mismatch, are the principal reasons behind the need for individuals to receive one or more booster doses of an mRNA vaccine [3, 11–14].

Prior studies of unvaccinated and vaccinated populations have shown more severe outcomes for COVID-19 infection for people with certain high-risk comorbidities such as diabetes, chronic kidney disease (CKD), lung disease, hypertension, or are immunocompromised (e.g., because of cancer, solid organ transplant, HIV, etc.) among many other conditions when compared to individuals without those conditions [6, 15–27]. However, most of the analyses surrounding breakthrough COVID-19 infection, and subsequent hospitalization, in vaccinated populations were not based on people in the United States (though see Embi et al. [24]), and instead focused on large populations in the United Kingdom [9, 19, 21].

Additionally, explicit interactions between comorbidities have not necessarily been analyzed, as previous work has tended to focus on one or two intrinsically related comorbidities [6, 7, 19, 24, 28, 29] or are full omnibus analyses which focus on incidence rates of breakthrough and hospitalization [5, 9, 10, 20, 21, 23, 30, 31].

In order to better understand the risk of breakthrough infection and severe outcomes in high-risk populations, we used Truveta data [32] to ask whether vaccinated patients with chronic kidney disease, chronic lung disease, diabetes, or those who have immunocompromising conditions have a greater risk of breakthrough COVID-19 infection and greater odds of hospitalization following breakthrough infection than in those vaccinated but without the studied comorbidities. We chose these comorbidities to study based on their prevalence in the US population, association with impaired immune function, as well as previous literature on risk factors for COVID hospitalization in unvaccinated and vaccinated populations. [9, 11, 15, 21, 33–35].

## 2 Methods

### 2.1 Study population

The study population included a subset of the Truveta patient population who received a complete initial series of an mRNA vaccine for COVID-19 patients present in Truveta between 2021-01-01 and 2022-03-31 [32]. We used the Truveta Studio to access the de-identified medical records used in this study on 2022-10-19. Truveta is a consortium of healthcare systems which have combined their electronic health record (EHR) data to enable medical research. Currently this consortium includes 25 members who provide patient care in over 20,000 clinics and 700 hospitals across 43 states. Updated data is provided daily to Truveta. Similar data fields across systems are mapped though syntactic normalization to a common schema referred to as the Truveta Data Model (TDM). Once organized into common fields, values are then semantically normalized to common ontologies such as ICD-10-CM, SNOMED-CT, LOINC, RxNorm, CVX, etc. These normalization procedures employ an expert-led, artificial intelligence driven process to accomplish high-confidence modeling at scale. The data are then de-identified by expert determination under the HIPAA Privacy Rule. Once de-identified, the data are then made available for analysis using Truveta Studio.

A patient was considered to have completed their primary vaccination series at two weeks after receiving a second mRNA vaccine dose (Moderna or Pfizer) based on the patient’s medical records. Patients were excluded from our study population if they were missing sex or age at time of vaccination fields, experienced a COVID-19 infection prior to being completing their initial mRNA COVID-19 vaccination sequence (i.e., 14 days post second vaccine dose), were missing their date of being fully vaccinated, and were under 18 years of age at time of first vaccine dose. All vaccination events generally consisted of vaccinations that took place within the health system as well as vaccination records actively pulled from the health system’s respective state’s Immunization Information System. See the Supplemental Material for a list of CVX codes corresponding to these vaccines.

Our four comorbidities of interest (chronic kidney disease, chronic lung disease, diabetes, and immunocompromised) were defined using Elixhauser comorbidity ICD-10-CM diagnostic codes and related SNOMED-CT diagnostic codes [36], and patients were identified as having one or more of these comorbidities based on the presence of these diagnostic codes in a patient’s medical record prior to their completion of a primary COVID-19 vaccination series. Patients who were diagnosed with a comorbidity after completing their initial dose sequence plus 14 days were excluded from analysis.

Our response variables of interest were 1) time from completing a COVID-19 primary vaccination series till breakthrough infection, and 2) if a patient who experienced a breakthrough infection was hospitalized within two-weeks of that infection. SARS-CoV-2 infection was defined as a patient’s first diagnosis of COVID-19 using either diagnosis codes or laboratory results.

The code lists associated with all studied conditions were initially based on code lists published to the National Institute of Health’s Value Set Authority Center website (https://vsac.nlm.nih.gov/). These value sets where then modified based on the expert opinion of the multiple clinical informaticists who are co-authors on this study. The complete lists of ICD-10-CM, SNOMED-CT, CVX, and LOINC codes for each of COVID-19 vaccination, COVID-19 diagnosis, COVID-19 test, and all considered comorbidities are presented in the Supplementary Material.

In addition to the four comorbidities stated above, we also included multiple demographic covariates in our models: race (White, Asian, Black or African American, American Indian or Alaska Native, Native Hawaiian or Other Pacific Islander, Unknown), ethnicity (Hispanic or Latino, Not Hispanic or Latino, and Unknown), sex, and person’s age in years at time of completing a COVID-19 primary vaccine sequence. In all analyses described below, the effect of age in years was modeled using a natural cubic spline with five degrees of freedom. We also included the year-month when a patient completed their primary vaccine sequence as a categorical covariate. We consider this variable as a proxy for differences associated with COVID-19 variant and transmission “environment” experienced by that patient.

### 2.2 Time from COVID-19 vaccination to breakthrough infection

We used a Cox proportional hazards model to describe the relationship between the time from completing a COVID-19 primary vaccination series till breakthrough infection and the comorbidities of interest and other covariates listed above. The response variable for our analysis of time from completing a COVID-19 primary vaccination series (i.e., time of second dose plus 14 days) till breakthrough infection was defined as the minimum time among three potential events: time of first COVID-19 infection, time of last encounter in EHR, and 180 days. If a patient did not experience a breakthrough infection they were considered right-censored at either 180 days or the time of their last encounter in the EHR, which ever was first. This censoring scheme assumes all censoring is uninformative and that any censored value less than 180 days indicates that a patient was lost to follow-up as of their last encounter in the EHR.

We hypothesized that, for patients with more than one of these four comorbidities, there may be an interaction effect influencing the chance of breakthrough infection. We developed four models of time till breakthrough infection, each allowing for a different degree of interactions. The base model assumes that the comorbidities have only an additive effect and there is no additional hazard associated having multiple comorbidities. We also considered all two-way interactions (i.e., any combination of 2 comorbidities), all two- and three-way interactions, and all two-, three-, and four-way interactions between the comorbidities.

We used AIC and AICc to compare the four different models of each outcome and then select the best model for each outcome. When comparing models of the same response, the model with the lowest AIC/AICc indicates which of those models does the best job balancing the complexity of the model and its likelihood [37–40]. This approach was chosen because we did not have a strong hypothesis as to the “correct” number of possible interactions, and we also wanted to balance the complexity of the model with the size of our data set.

The hazard ratio associated for individual comorbidities with no interaction terms is normally calculated as the exponentiated regression coefficients from the Cox regression model. However, as we are interested in the hazard ratios associated with a patient having one or more comorbidities in combination versus a patient with no comorbidities, we used the emmeans R package to calculate these hazard ratios [41].

### 2.3 Odds of hospitalization following breakthrough infection

We used a logistic regression model to describe the relationship between hospitalization following breakthrough COVID-19 infection and the comorbidities and other covariates listed above. Hospitalization following a breakthrough COVID-19 infection was defined as an inpatient encounter where the patient was hospitalized within 14 days of a positive SARS-CoV-2 test. We choose to analyze this outcome because we were specifically interested in the conversion probability from “infected with COVID-19” to “hospitalized” and not the time from vaccination till hospitalization nor time from breakthrough infection till hospitalization. This is a binary scenario describing a transition probability within the infected population for which this method is appropriate as we believe this analysis captures our research question well [42].

As with our analysis of time till breakthrough infection, we hypothesized that, for patients with more than one of these four comorbidities, there may be an interaction effect influencing the odds of hospitalization following breakthrough infection. We developed four models of hospitalization following breakthrough infection, each allowing for a different degree of interactions. The base model assumes that the comorbidities have only an additive effect and there is no additional hazard associated having multiple comorbidities. We also developed models which consisted of all two-way interactions (i.e., any combination of 2 comorbidities), all two- and three-way interactions, and all two-, three-, and four-way interactions. And as with the models of time till breakthrough infection, we used AIC and AICc to compare these models and select the best one among that group.

The odds ratio associated for individual comorbidities with no interaction terms is normally calculated as the exponentiated regression coefficients from the logistic regression model [42]. However, as we are interested in the odds ratios associated with a patient having one or more comorbidities in combination versus a patient with no comorbidities, we used the emmeans R package to calculate these odds ratios [41].

Analysis was done using the R programming language (4.1.1) [43] along with the following packages: `arrow` [44], `broom` [45], `dplyr` [46], `emmeans` [41], `ggplot2` [47], `janitor` [48], `purrr` [49], `rlang` [50], `stringr` [51], `survival` [52], `table1` [53], `tibble` [54], and `tidyr` [55].

The R code used to run the analyses presented in this study is available at https://github.com/Truveta/smits_et_al_covid_breakthrough_comorbidities.

## 3 Results

### 3.1 Study population

1,192,135 patients on Truveta met the study inclusion and exclusion criteria of having completed a primary vaccination sequence between 2021-01-01 and 2022-03-31. Of the patients in our study, 68,218 had chronic kidney disease, 138,752 had chronic lung disease, 133,829 had diabetes, 302,469 were considered immunocompromised, and 756,178 had none of these four comorbidities (Table 1). Note that patients can have more than one comorbidity, and thus the sum of patients with each comorbidity will exceed the total of patients with comorbidities.

**Table 1:** Overall summary statistics of our analyzed population of patients who have completed their primary COVID-19 vaccination sequence.

|  | Chronic kidney disease<br>(N=68218) | Chronic lung disease<br>(N=138752) | Diabetes<br>(N=133829) | Immunocompromised<br>(N=302469) | Comorbidity free<br>(N=756178) | Overall<br>(N=1192135) |
| --- | --- | --- | --- | --- | --- | --- |
| Sex |  |  |  |  |  |  |
| Female | 36215 (53.1%) | 89783 (64.7%) | 69772 (52.1%) | 190711 (63.1%) | 462136 (61.1%) | 726128 (60.9%) |
| Male | 32003 (46.9%) | 48969 (35.3%) | 64057 (47.9%) | 111758 (36.9%) | 294042 (38.9%) | 466007 (39.1%) |
| Race |  |  |  |  |  |  |
| White | 52245 (76.6%) | 109149 (78.7%) | 94270 (70.4%) | 238983 (79.0%) | 520728 (68.9%) | 855777 (71.8%) |
| Unknown | 2806 (4.1%) | 8139 (5.9%) | 10429 (7.8%) | 19073 (6.3%) | 126038 (16.7%) | 156050 (13.1%) |
| Black or African American | 10911 (16.0%) | 16442 (11.9%) | 21093 (15.8%) | 30649 (10.1%) | 58856 (7.8%) | 108672 (9.1%) |
| Asian | 1944 (2.8%) | 3947 (2.8%) | 6945 (5.2%) | 11869 (3.9%) | 46302 (6.1%) | 64462 (5.4%) |
| American Indian or Alaska Native | 177 (0.3%) | 737 (0.5%) | 591 (0.4%) | 1227 (0.4%) | 2297 (0.3%) | 4087 (0.3%) |
| Native Hawaiian or Other Pacific Islander | 135 (0.2%) | 338 (0.2%) | 501 (0.4%) | 668 (0.2%) | 1957 (0.3%) | 3087 (0.3%) |
| Ethnicity |  |  |  |  |  |  |
| Not Hispanic or Latino | 63629 (93.3%) | 127182 (91.7%) | 117799 (88.0%) | 275847 (91.2%) | 604305 (79.9%) | 997342 (83.7%) |
| Hispanic or Latino | 3234 (4.7%) | 8308 (6.0%) | 12509 (9.3%) | 18759 (6.2%) | 83532 (11.0%) | 114298 (9.6%) |
| Unknown | 1355 (2.0%) | 3262 (2.4%) | 3521 (2.6%) | 7863 (2.6%) | 68341 (9.0%) | 80495 (6.8%) |
| Age (years) |  |  |  |  |  |  |
| Mean (SD) | 72.2 (12.6) | 59.4 (18.0) | 64.7 (13.6) | 59.9 (17.0) | 49.8 (17.7) | 53.6 (18.1) |
| Median [Min, Max] | 73.7 [18.2, 99.0] | 62.4 [18.1, 99.0] | 66.2 [18.1, 98.7] | 62.4 [18.1, 99.0] | 49.7 [18.1, 98.9] | 54.9 [18.1, 99.0] |
| Age bracket |  |  |  |  |  |  |
| [18,20) | 26 (0.0%) | 1843 (1.3%) | 152 (0.1%) | 2786 (0.9%) | 16940 (2.2%) | 20824 (1.7%) |
| [20,25) | 105 (0.2%) | 4843 (3.5%) | 651 (0.5%) | 7656 (2.5%) | 49101 (6.5%) | 59909 (5.0%) |
| [25,30) | 230 (0.3%) | 5020 (3.6%) | 1161 (0.9%) | 8721 (2.9%) | 58187 (7.7%) | 70527 (5.9%) |
| [30,35) | 416 (0.6%) | 5949 (4.3%) | 1941 (1.5%) | 11646 (3.9%) | 65588 (8.7%) | 81839 (6.9%) |
| [35,45) | 1670 (2.4%) | 13896 (10.0%) | 8029 (6.0%) | 31491 (10.4%) | 129616 (17.1%) | 173002 (14.5%) |
| [45,55) | 4087 (6.0%) | 18621 (13.4%) | 17985 (13.4%) | 44299 (14.6%) | 128948 (17.1%) | 192017 (16.1%) |
| [55,65) | 9887 (14.5%) | 26980 (19.4%) | 32107 (24.0%) | 61975 (20.5%) | 131675 (17.4%) | 222898 (18.7%) |
| [65,75) | 21163 (31.0%) | 33668 (24.3%) | 41118 (30.7%) | 75695 (25.0%) | 115717 (15.3%) | 226075 (19.0%) |
| [75,85) | 20998 (30.8%) | 21137 (15.2%) | 24266 (18.1%) | 44578 (14.7%) | 47863 (6.3%) | 112383 (9.4%) |
| [85,Inf) | 9636 (14.1%) | 6795 (4.9%) | 6419 (4.8%) | 13622 (4.5%) | 12543 (1.7%) | 32661 (2.7%) |
| Breakthrough COVID-19 infection |  |  |  |  |  |  |
| Breakthrough COVID | 1936 (2.8%) | 4740 (3.4%) | 3695 (2.8%) | 8749 (2.9%) | 10229 (1.4%) | 22087 (1.9%) |
| No breakthrough | 66282 (97.2%) | 134012 (96.6%) | 130134 (97.2%) | 293720 (97.1%) | 745949 (98.6%) | 1170048 (98.1%) |
| Hospitalization following breakthrough |  |  |  |  |  |  |
| Hospitalized | 842 (1.2%) | 1213 (0.9%) | 1040 (0.8%) | 1912 (0.6%) | 775 (0.1%) | 3233 (0.3%) |
| Not hospitalized | 67376 (98.8%) | 137539 (99.1%) | 132789 (99.2%) | 300557 (99.4%) | 755403 (99.9%) | 1188902 (99.7%) |

### 3.2 Time from COVID-19 vaccination to breakthrough infection

The model of time from COVID-19 primary sequence till breakthrough infection with a maximum of four-way interactions between the comorbidities was considered “best” among the candidate models (Table 2). This means that specific interaction effects (e.g., changes to hazard ratios associated with specific combinations of comorbidities) are estimated up to the maximum possible four-way interaction among the comorbidities. This result means that we have found evidence that while patients with one of these four comorbities had an increased risk of breakthrough COVID-19 infection compared to individuals without any of these comorbidities, patients with two or more of the comorbidities have further increased risk than would be expected by the independent effects of the comorbidities on odds of breakthrough infection.

**Table 2:** Comparison between four candidate models of time till breakthrough COVID-19 infection each with varying degrees of interaction between comorbidities.

| Model complexity | AIC | delta_AIC | AICc | delta_AICc |
| --- | --- | --- | --- | --- |
| Four-way | 601933.00 | 0.00 | 601933.32 | 0.00 |
| Three-way | 601936.72 | 3.73 | 601937.03 | 3.71 |
| Two-way | 601935.42 | 2.42 | 601935.67 | 2.35 |
| Base | 602032.80 | 99.80 | 602032.98 | 99.66 |

Presented here (Fig.1) are the hazard ratios of breakthrough COVID-19 infection for a patient having one or more comorbidities versus a patient with none of the comorbidities. Our selected model included up to four-way interactions between comorbidities.

**Figure 1:**
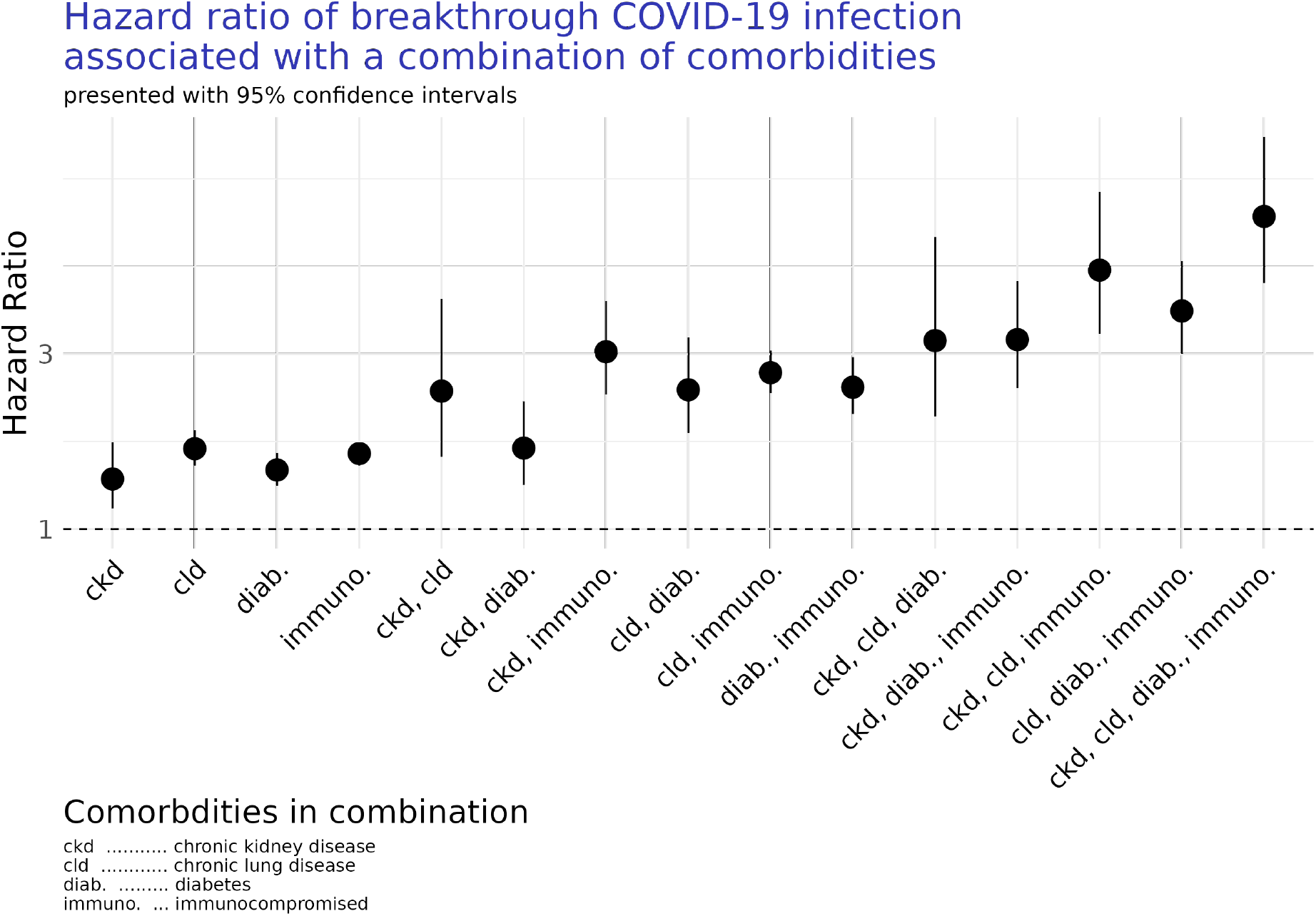
Estimated hazards-ratios of breakthrough COVID-19 infection associated with one or more comorbidity versus being comorbidity free. Hazards-ratios are estimated from a model which considers up to four-way interactions between comorbidities.

We find that persons with any of the studied comorbidities, in any combination, were associated with a greater risk of breakthrough COVID-19 infection than those persons without any comorbidities after adjustment (Fig. 1; CKD HR 1.57 [CI 1.24, 2.00]; immunocompromised HR 1.86 [CI 1.75, 1.99]; diabetes HR 1.67 [CI 1.50, 1.87]; chronic lung disease HR 1.91 [CI 1.73, 2.13]). See Supplemental Material for a full breakdown of the selected model’s parameter estimates.

### 3.3 Odds of hospitalization following breakthrough infection

The model of hospitalization following breakthrough COVID-19 infection with no interaction terms between the comorbidities was considered “best” among the candidate models (Table 3). While we are able to calculate the odds ratio for an arbitrary number of interactions, our model included no interaction effects among the comorbidities so the presented values are based on additive effects alone. While individuals with multiple of the comorbidities have an increased risk of hospitalization following breakthrough infection, our model selection results are consistent with the comorbidities having independent additive effects on the odds of hospitalization.

**Table 3:** Comparison between four candidate models of probability of hospitalization following breakthrough COVID-19 infection each with varying degrees of interaction between comorbidities.

| Model complexity | AIC | ΔAIC | AICc | ΔAICc |
| --- | --- | --- | --- | --- |
| Four-way | 17752.96 | 3.63 | 17753.28 | 3.78 |
| Three-way | 17751.35 | 2.02 | 17751.65 | 2.15 |
| Two-way | 17759.26 | 9.94 | 17759.52 | 10.01 |
| Base | 17749.33 | 0.00 | 17749.51 | 0.00 |

Presented here (Fig.2) are the odds ratio of hospitalization following a breakthrough COVID-19 infection for a patient having one or more comorbidities versus a patient with none of the comorbidities. Our selected model does not include any interaction effects among the comorbidities, though we can calculate these odds ratios for any combination of comorbidities.

**Figure 2:**
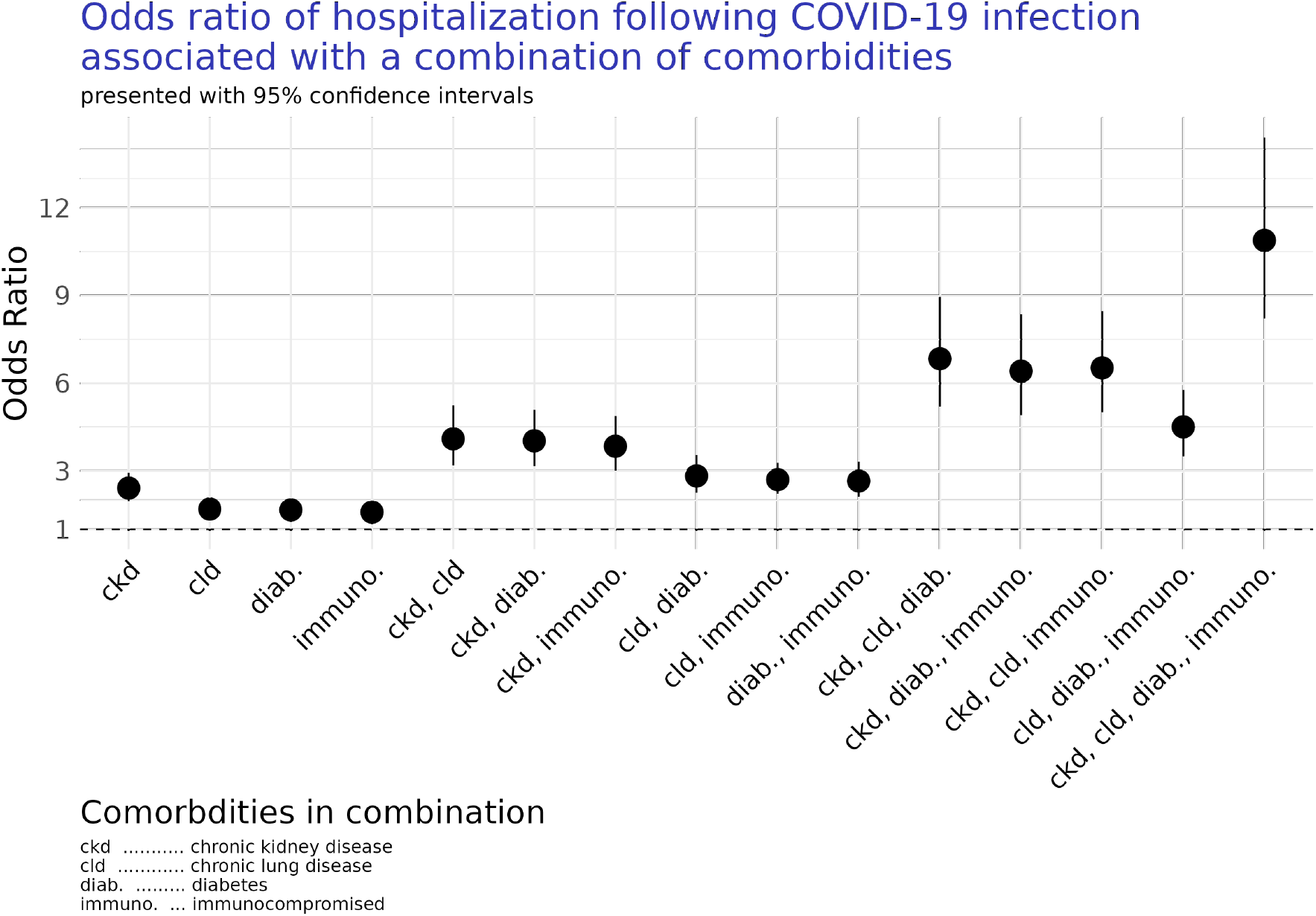
Estimated odds-ratios of breakthrough COVID-19 infection associated with one or more comorbidity versus being comorbidity free. Odds-ratios are estimated from a model which considers no interactions between comorbidities, and their combined effects do not reflect any explicit interaction effects between comorbidities.

We find that all of the comorbidities were associated with an increased risk of hospitalization following breakthrough COVID-19 infection when compared to patients without any of the comorbidities of interest after adjustment (Fig. 2; CKD OR 2.41 [CI 1.97, 2.95]; immunocompromised OR 1.59 [CI 1.37, 1.85]; diabetes OR 1.66 [CI 1.41, 1.98]; chronic lung disease OR 1.70 [CI 1.45, 1.99]). See Supplemental Material for a full breakdown of the selected model’s parameter estimates.

## 4 Discussion

Here we found that the risk of SARS-CoV-2 breakthrough infection and odds of subsequent hospitalization following breakthrough infection were greater among vaccinated patients with diabetes, chronic lung disease, CKD, and with immunocompromising conditions when compared to the vaccinated individuals without these conditions after adjusting for age, sex, race, ethnicity, and year-month of vaccination (Fig. 1). This is consistent with studies in unvaccinated and vaccinated people showing higher risk of infection and hospitalization in people with any of these comorbidities or similar high-risk conditions [5–7, 9, 10, 19–21, 23, 24, 26, 28–31]. We also found that while patients with one of these four comorbities had an increased risk of breakthrough COVID-19 infection compared to individuals without any of these comorbidities, patients with two or more of the comorbidities have further increased risk than would be expected by the independent effects of the comorbidities on odds of breakthrough infection (Fig. 1, Table 2). In contrast, while individuals with multiple of the comorbidities have an increased risk of hospitalization following breakthrough infection, our model selection results are consistent with the comorbidities having independent effects on odds of hospitalization (Fig. 2, Table 3).

We identified chronic kidney disease as the highest risk individual comorbidity for hospitalization after adjustment for age and other demographic factors (Fig. 2). This result is consistent with previous work examining differences in COVID-19 breakthrough infection incident rates in the United Kingdom [19, 21]. These studies found that the effectiveness of the COVID-19 vaccines against breakthrough infection and subsequent hospitalization varied with the severity of chronic kidney disease. In contrast, a large study in male U.S. veterans did not show an elevated risk of severe outcomes in breakthrough infections in patients with diabetes, chronic lung disease, or CKD [27]. This discrepancy was possibly due to differences in study design where patients were matched by comorbidity burden which may reduces differences between health status and demographics between groups and may limit the generalizability between patients in the Veterans Health Administration population and the other sample populations [56]. In contrast our study compared patients with any of the identified comorbidities in any combination as well as with patients who had none of these comorbidities, and did not exclude patients with multiple comorbidities.

Like all studies of EHR data, ours is subject to a variety of known limitations [57–62]. We are only able to identify events that are captured by the constituent health care systems that are a part of the Truveta member system. This means we will not capture COVID-19 infections which were reported or diagnosed by a health care system that is not a part of the Truveta. Similarly, we will not capture COVID-19 infections which were never reported to a health care system. This limitation means we patients with a precedent COVID-19 infection may be missed as part of our inclusion and exclusion criteria. Another example limitation is that a patient’s COVID-19 vaccination status may not captured in our data because only a limited number of member HCS reconcile their records with state health registries and other locations where many patients may have been vaccinated. Finally, a patients’ comorbidity status may be misclassified in our data set because their comorbidity status is captured in a different, non-member HCS or they are classified in the EHR using codes that were not present in our codesets. These are common and well understood limitations associated with using this kind of data. In the context of this study these inherent limitations will most likely lead to an underestimation of the size of the vaccinated population which will most likely lead to an underestimation of the effects of the comorbidities on risk of breakthrough COVID-19 infection and subsequent odds of hospitalization following breakthrough infection, especially in combination.

In addition to the limitations inherent in retrospective analysis of EHR data, there are other limitations associated with our study. For example, we did not include certain risk factors such as hypertension and smoking status due to limitations of the Truveta Platform at the time of analysis. Additionally, we do not consider alternate outcomes or competing risks in our analysis of time from COVID-19 vaccination till breakthrough infection, and instead consider all censoring uninformative. A follow-up analysis should consider a large suite of demographic features and risk factors as well as more varied outcomes and the potential for competing risks.

Future work should also consider the effect of booster doses on time till breakthrough COVID-19 infection and odds of hospitalization following breakthrough infection. The timing of the booster dose would most likely have to be accounted for as a time-varying covariate to account for the variation in time from completion of primary dose sequence till time of a booster dose. However, many individuals in our population were vaccinated well before booster doses were made available, meaning that booster doses may not be captured by the 180-day follow-up period. Similarly, additional subgroup analyses of differences breakthrough infection and hospitalization associated with severity level of CKD is warranted as recent analysis have found substantial differences in COVID-19 outcomes associated with severity of CKD [19, 21].

Overall, these findings complement prior studies which have shown worse outcomes following COVID-19 infection in people who are have diabetes, CKD, chronic lung disease, or immunocompromising conditions [5–7, 9, 10, 15–17, 19–21, 23, 24, 28–31]. These results add additional support to the recommendation of booster vaccines for those with high-risk conditions given that these groups continue to fare worse than the general vaccinated population in terms of breakthrough infection and subsequent hospitalization rates. Those with comorbidities will most likely benefit from booster vaccinations to increase and improve their immune response to infection, as has been observed in people with chronic kidney disease [19].

As vaccinated people continue to make decisions about booster vaccinations, they will be looking for information regarding their personal risk of breakthrough COVID-19 infection and severe outcomes like hospitalization. The FDA and CDC have both made recommendations that people belonging to high-risk groups, such as those with immunocompromising conditions, should receive additional doses of the COVID-19 vaccines and at a faster rate than the general population. The findings of this study improve the evidence and support recommendations for people with comorbidities such as chronic kidney disease, chronic lung disease, diabetes or who have immunocompromising conditions to receive booster vaccinations.

## Data Availability

All data produced in the present study are available with subscription to the Truveta Platform.

https://github.com/Truveta/smits_et_al_covid_breakthrough_comorbidities

## Acknowledgements

We thank Ari Robicsek, Sarah Gilson, and Ryan H Lee for help with data definitions, as well as helpful comments which improved this analysis and manuscript. We would also thank our engineering partners at Truveta for assisting us with enabling this research, in particular Ian Davies, Grace Turner, David Nguyen, Mohan Dharmarajan, and George Joy. Finally, we thank the editor and three anonymous reviewers whose comments greatly improved the analysis and manuscript.

## References

[1] Fernando P. Polack, Stephen J. Thomas, Nicholas Kitchin, Judith Absalon, Alejandra Gurtman, Stephen Lockhart, John L. Perez, Gonzalo Pérez Marc, Edson D. Moreira, Cristiano Zerbini, Ruth Bailey, Kena A. Swanson, Satrajit Roychoudhury, Kenneth Koury, Ping Li,Warren V. Kalina, David Cooper, Robert W. Frenck, Laura L. Hammitt, Özlem Türeci, Haylene Nell, Axel Schaefer, Serhat Ünal, Dina B. Tresnan, Susan Mather, Philip R. Dormitzer, Uğur Şahin, Kathrin U. Jansen, and William C. Gruber. Safety and efficacy of the BNT162b2 mRNA covid-19 vaccine. New England Journal of Medicine, 383(27):2603–2615, dec 2020. doi: 10.1056/nejmoa2034577. URL https://doi.org/10.1056%2Fnejmoa2034577.

[2] Lindsey R. Baden, Hana M. El Sahly, Brandon Essink, Karen Kotloff, Sharon Frey, Rick Novak, David Diemert, Stephen A. Spector, Nadine Rouphael, C. Buddy Creech, John McGettigan, Shishir Khetan, Nathan Segall, Joel Solis, Adam Brosz, Carlos Fierro, Howard Schwartz, Kathleen Neuzil, Lawrence Corey, Peter Gilbert, Holly Janes, Dean Follmann, Mary Marovich, John Mascola, Laura Polakowski, Julie Ledgerwood, Barney S. Graham, Hamilton Bennett, Rolando Pajon, Conor Knightly, Brett Leav, Weiping Deng, Honghong Zhou, Shu Han, Melanie Ivarsson, Jacqueline Miller, and Tal Zaks. Efficacy and safety of the mRNA-1273 SARS-CoV-2 vaccine. New England Journal of Medicine, 384(5):403–416, feb 2021. doi: 10.1056/nejmoa2035389. URL https://doi.org/10.1056%2Fnejmoa2035389.

[3] Philip R Krause, Thomas R Fleming, Richard Peto, Ira M Longini, J Peter Figueroa, Jonathan A C Sterne, Alejandro Cravioto, Helen Rees, Julian P T Higgins, Isabelle Boutron, Hongchao Pan, Marion F Gruber, Narendra Arora, Fatema Kazi, Rogerio Gaspar, Soumya Swaminathan, Michael J Ryan, and Ana-Maria Henao-Restrepo. Considerations in boosting covid-19 vaccine immune responses. The Lancet, 398(10308):1377–1380, 2021. doi: 10.1016/s0140-6736(21)02046-8.

[4] Timothy A. Bates, Savannah K. McBride, Bradie Winders, Devin Schoen, Lydie Trautmann, Marcel E. CURLin, and Fikadu G. Tafesse. Antibody response and variant cross-neutralization after sars-cov-2 breakthrough infection. JAMA, 327(2):179–181, 2022. doi: 10.1001/jama.2021.22898.

[5] Prerak V Juthani, Akash Gupta, Kelly A Borges, Christina C Price, Alfred I Lee, Christine H Won, and Hyung J Chun. Hospitalisation among vaccine breakthrough covid-19 infections. The Lancet, 21(11):1485–1486, 2021. doi: 10.1016/s1473-3099(21)00558-2.

[6] A.L. Schmidt, C. Labaki, C.-Y. Hsu, Z. Bakouny, N. Balanchivadze, S.A. Berg, S. Blau, A. Daher, T. El Zarif, C.R. Friese, E.A. Griffiths, J.E. Hawley, B. Hayes-Lattin, V. Karivedu, T. Latif, B.H. Mavromatis, R.R. McKay, G. Nagaraj, R.H. Nguyen, O.A. Panagiotou, A.J. Portuguese, M. Puc, M. Santos Dutra, B.A. Schroeder, A. Thakkar, E.M. Wulff-Burchfield, S. Mishra, D. Farmakiotis, Yu Shyr, J.L. Warner, T.K. Choueiri, T.K. Choueiri, N. Duma, D. Farmakiotis, P. Grivas, G. de Lima Lopes, C.A. Painter, S. Peters, B.I. Rini, D.P. Shah, M.A. Thompson, and J.L. Warner. Covid-19 vaccination and breakthrough infections in patients with cancer. Annals of Oncology, 33(3):340–346, 2022. doi: 10.1016/j.annonc.2021.12.006.

[7] Norbert Stefan. Metabolic disorders, covid-19 and vaccine-breakthrough infections. Nature Reviews Endocrinology, 18:75–76, 2022. doi: 10.1038/s41574-021-00608-9.

[8] Noa Dagan, Noam Barda, Eldad Kepten, Oren Miron, Shay Perchik, Mark A. Katz, Miguel A. Hernán, Marc Lipsitch, Ben Reis, and Ran D. Balicer. Bnt162b2 mrna covid-19 vaccine in a nationwide mass vaccination setting. The New England Journal of Medicine, 384:1412–1423, 2021. doi: 10.1056/nejmoa2101765.

[9] Utkarsh Agrawal, Srinivasa Vittal Katikireddi, Colin McCowan, Rachel H Mulholland, Amaya Azcoaga-Lorenzo, Sarah Amele, Adeniyi Francis Fagbamigbe, Eleftheria Vasileiou, Zoe Grange, Ting Shi, Steven Kerr, Emily Moore, Josephine L K Murray, Syed Ahmar Shah, Lewis Ritchie, Dermot O’Reilly, Sarah J Stock, Jillian Beggs, Antony Chuter, Fatemah Torabi, Ashley Akbari, Stuart Bedston, Jim McMenamin, Rachael Wood, Ruby S M Tang, Simon de Lusignan, F D Richard Hobbs, Mark Woolhouse, Colin R Simpson, Chris Robertson, and Aziz Sheikh. Covid-19 hospital admissions and deaths after bnt162b2 and chadox1 ncov-19 vaccinations in 2.57 million people in scotland (eave ii): a prospective cohort study. The Lancet Respiratory Medicine, 9(12):1439–1449, 2021. doi: 10.1016/s2213-2600(21)00380-5.

[10] Veerle Stouten, Pierre Hubin, Freek Haarhuis, Joris A. F. van Loenhout, Matthieu Billuart, Ruben Brondeel, Toon Braeye, Herman Van Oyen, Chloé Wyndham-Thomas, and Lucy Catteau. Incidence and risk factors of covid-19 vaccine breakthrough infections: A prospective cohort study in belgium. Viruses, 14(4), 2022. ISSN 1999-4915. doi: 10.3390/v14040802. URL https://www.mdpi.com/1999-4915/14/4/802.

[11] Kathleen Dooling. Evidence to Recommendation Framework: Moderna & Janssen COVID-19 Vaccine Booster Dose, 2021. URL https://www.cdc.gov/vaccines/acip/meetings/downloads/slides-2021-10-20-21/11-COVID-Dooling-508.pdf.

[12] Sara Oliver. Evidence to Recommendation Framework: Pfizer-BioNTech COVID-19 Booster Dose, 2021. URL https://www.cdc.gov/vaccines/acip/meetings/downloads/slides-2021-9-23/03-COVID-Oliver.pdf.

[13] Arjun Puranik, Patrick J. Lenehan, Eli Silvert, Michiel J.M. Niesen, Juan Corchado-Garcia, John C. O’Horo, Abinash Virk, Melanie D. Swift, John Halamka, Andrew D. Badley, A.J. Venkatakrishnan, and Venky Soundararajan. Comparison of two highly-effective mRNA vaccines for COVID-19 during periods of Alpha and Delta variant prevalence. medRxiv, page 2021.08.06.21261707, January 2021. doi: 10.1101/2021.08.06.21261707. URL http://medrxiv.org/content/early/2021/08/21/2021.08.06.21261707.abstract.

[14] Jocelyn Keehner, Lucy E. Horton, Nancy J. Binkin, Louise C. Laurent, David Pride, Christopher A. Longhurst, Shira R. Abeles, and Francesca J. Torriani. Resurgence of SARS-CoV-2 Infection in a Highly Vaccinated Health System Workforce. The New England journal of medicine, 385(14):1330–1332, September 2021. ISSN 1533-4406 0028-4793. doi: 10.1056/NEJMc2112981.

[15] Michael W Fried, Julie M Crawford, Andrea R Mospan, Stephanie E Watkins, Breda Munoz, Richard C Zink, Sherry Elliott, Kyle BURLeson, Charles Landis, K Rajender Reddy, and Robert S Brown. Patient Characteristics and Outcomes of 11 721 Patients With Coronavirus Disease 2019 (COVID-19) Hospitalized Across the United States. Clinical Infectious Diseases, 72(10):e558–e565, May 2021. ISSN 1058-4838, 1537-6591. doi: 10.1093/cid/ciaa1268. URL https://academic.oup.com/cid/article/72/10/e558/5898276.

[16] Arthur Eumann Mesas, Iván Cavero-Redondo, Celia Álvarez Bueno, Marcos Aparecido Sarriá Cabrera, Selma Maffei de Andrade, Irene Sequí-Dominguez, and Vicente Martínez-Vizcaíno. Predictors of in-hospital COVID-19 mortality: A comprehensive systematic review and meta-analysis exploring differences by age, sex and health conditions. PLOS ONE, 15(11): e0241742, November 2020. ISSN 1932-6203. doi: 10.1371/journal.pone.0241742. URL https://dx.plos.org/10.1371/journal.pone.0241742.

[17] Ya Gao, Yamin Chen, Ming Liu, Shuzhen Shi, and Jinhui Tian. Impacts of immunosuppression and immunodeficiency on COVID-19: A systematic review and meta-analysis. The Journal of Infection, 81(2):e93–e95, August 2020. ISSN 1532-2742. doi: 10.1016/j.jinf.2020.05.017.

[18] Kamal S. Saini, Marco Tagliamento, Matteo Lambertini, Richard McNally, Marco Romano, Manuela Leone, Giuseppe Curigliano, and Evandro de Azambuja. Mortality in patients with cancer and coronavirus disease 2019: A systematic review and pooled analysis of 52 studies. European Journal of Cancer (Oxford, England: 1990), 139:43–50, November 2020. ISSN 1879-0852. doi: 10.1016/j.ejca.2020.08.011.

[19] Edward PK Parker, Elsie MF Horne, William J Hulme, John Tazare, Bang Zheng, Edward J Carr, Fiona Loud, Susan Lyon, Viyaasan Mahalingasivam, Brian MacKenna, Amir Mehrkar, Miranda Scanlon, Shalini Santhakumaran, Retha Steenkamp, Ben Goldacre, Jonathan AC Sterne, Dorothea Nitsch, and Laurie A Tomlinson. Comparative effectiveness of two- and three-dose schedules involving azd1222 and bnt162b2 in people with kidney disease: a linked opensafely and uk renal registry cohort study. medRxiv, 2022. doi: 10.1101/2022.11.16.22282396. URL https://www.medrxiv.org/content/early/2022/11/18/2022.11.16.22282396.

[20] Elizabeth J. Williamson, Alex J. Walker, Krishnan Bhaskaran, Seb Bacon, Chris Bates, Caroline E. Morton, Helen J. Curtis, Amir Mehrkar, David Evans, Peter Inglesby, Jonathan Cockburn, Helen I. McDonald, Brian MacKenna, Laurie Tomlinson, Ian J. Douglas, Christopher T. Rentsch, Rohini Mathur, Angel Y. S. Wong, Richard Grieve, David Harrison, Harriet Forbes, Anna Schultze, Richard Croker, John Parry, Frank Hester, Sam Harper, Rafael Perera, Stephen J. W. Evans, Liam Smeeth, and Ben Goldacre. Factors associated with COVID-19-related death using OpenSAFELY. Nature, 584(7821):430–436, jul 2020. doi: 10.1038/s41586-020-2521-4. URL https://doi.org/10.1038%2Fs41586-020-2521-4.

[21] Amelia Green, Helen Curtis, William Hulme, Elizabeth Williamson, Helen McDonald, Krishnan Bhaskaran, Christopher Rentsch, Anna Schultze, Brian MacKenna, Viyaasan Mahalingasivam, Laurie Tomlinson, Alex Walker, Louis Fisher, Jon Massey, Colm Andrews, Lisa Hopcroft, Caroline Morton, Richard Croker, Jessica Morley, Amir Mehrkar, Seb Bacon, David Evans, Peter Inglesby, George Hickman, Tom Ward, Simon Davy, Rohini Mathur, John Tazare, Rosalind Eggo, Kevin Wing, Angel Wong, Harriet Forbes, Chris Bates, Jonathan Cockburn, John Parry, Frank Hester, Sam Harper, Ian Douglas, Stephen Evans, Liam Smeeth, Ben Goldacre, and The OpenSAFELY Collaborative. Describing the population experiencing COVID-19 vaccine breakthrough following second vaccination in England: a cohort study from OpenSAFELY. BMC Medicine, 20(1):243, July 2022. ISSN 1741-7015. doi: 10.1186/s12916-022-02422-0. URL https://doi.org/10.1186/s12916-022-02422-0.

[22] Dave Singh, Alexander G. Mathioudakis, and Andrew Higham. Chronic obstructive pulmonary disease and covid-19: interrelationships. Current Opinion in Pulmonary Medicine, 28(2):76–83, 2022. doi: 10.1097/mcp.0000000000000834.

[23] Krishnan Bhaskaran, Christopher T. Rentsch, George Hickman, William J. Hulme, Anna Schultze, Helen J. Curtis, Kevin Wing, Charlotte Warren-Gash, Laurie Tomlinson, Chris J. Bates, Rohini Mathur, Brian MacKenna, Viyaasan Mahalingasivam, Angel Wong, Alex J. Walker, Caroline E. Morton, Daniel Grint, Amir Mehrkar, Rosalind M. Eggo, Peter Inglesby, Ian J. Douglas, Helen I. McDonald, Jonathan Cockburn, Elizabeth J. Williamson, David Evans, John Parry, Frank Hester, Sam Harper, Stephen JW Evans, Sebastian Bacon, Liam Smeeth, and Ben Goldacre. Overall and cause-specific hospitalisation and death after COVID-19 hospitalisation in england: A cohort study using linked primary care, secondary care, and death registration data in the OpenSAFELY platform. PLOS Medicine, 19(1):e1003871, jan 2022. doi: 10.1371/journal.pmed.1003871. URL https://doi.org/10.1371%2Fjournal.pmed.1003871.

[24] Peter J. Embi, Matthew E. Levy, Allison L. Naleway, Palak Patel, Manjusha Gaglani, Karthik Natarajan, Kristin Dascomb, Toan C. Ong, Nicola P. Klein, I-Chia Liao, Shaun J. Grannis, Jungmi Han, Edward Stenehjem, Margaret M. Dunne, Ned Lewis, Stephanie A. Irving, Suchitra Rao, Charlene McEvoy, Catherine H. Bozio, Kempapura Murthy, Brian E. Dixon, Nancy Grisel, Duck-Hye Yang, Kristin Goddard, Anupam B. Kharbanda, Sue Reynolds, Chandni Raiyani, William F. Fadel, Julie Arndorfer, Elizabeth A. Rowley, Bruce Fireman, Jill Ferdinands, Nimish R. Valvi, Sarah W. Ball, Ousseny Zerbo, Eric P. Griggs, Patrick K. Mitchell, Rachael M. Porter, Salome A. Kiduko, Lenee Blanton, Yan Zhuang, Andrea Steffens, Sarah E. Reese, Natalie Olson, Jeremiah Williams, Monica Dickerson, Meredith McMorrow, Stephanie J. Schrag, Jennifer R. Verani, Alicia M. Fry, Eduardo Azziz-Baumgartner, Michelle A. Barron, Mark G. Thompson, and Malini B. DeSilva. Effectiveness of 2-dose vaccination with mRNA COVID-19 vaccines against COVID-19–associated hospitalizations among immunocompromised adults — nine states, january–september 2021. MMWR. Morbidity and Mortality Weekly Report, 70(44):1553–1559, nov 2021. doi: 10.15585/mmwr.mm7044e3. URL https://doi.org/10.15585%2Fmmwr.mm7044e3.

[25] Florian B. Mayr, Victor B. Talisa, Alexander D. Castro, Obaid S. Shaikh, Saad B. Omer, and Adeel A. Butt. COVID-19 disease severity in US veterans infected during omicron and delta variant predominant periods. Nature Communications, 13(1), jun 2022. doi: 10.1038/s41467-022-31402-4. URL https://doi.org/10.1038%2Fs41467-022-31402-4.

[26] Adeel A. Butt, Saad B. Omer, Peng Yan, Obaid S. Shaikh, and Florian B. Mayr. SARS-CoV-2 Vaccine Effectiveness in a High-Risk National Population in a Real-World Setting. Annals of Internal Medicine, 174(10):1404–1408, October 2021. ISSN 1539-3704. doi: 10.7326/M21-1577.

[27] Adeel A. Butt, Peng Yan, Obaid S. Shaikh, and Florian B. Mayr. Outcomes among patients with breakthrough SARS-CoV-2 infection after vaccination in a high-risk national population. EClinicalMedicine, 40:101117, October 2021. ISSN 25895370. doi: 10.1016/j.eclinm.2021.101117. URL https://linkinghub.elsevier.com/retrieve/pii/S2589537021003977.

[28] Brian MacKenna, Nicholas A Kennedy, Amir Mehrkar, Anna Rowan, James Galloway, Julian Matthewman, Kathryn E Mansfield, Katie Bechman, Mark Yates, Jeremy Brown, Anna Schultze, Sam Norton, Alex J Walker, Caroline E Morton, David Harrison, Krishnan Bhaskaran, Christopher T Rentsch, Elizabeth Williamson, Richard Croker, Seb Bacon, George Hickman, Tom Ward, Simon Davy, Amelia Green, Louis Fisher, William Hulme, Chris Bates, Helen J Curtis, John Tazare, Rosalind M Eggo, David Evans, Peter Inglesby, Jonathan Cockburn, Helen I McDonald, Laurie A Tomlinson, Rohini Mathur, Angel Y S Wong, Harriet Forbes, John Parry, Frank Hester, Sam Harper, Ian J Douglas, Liam Smeeth, Charlie W Lees, Stephen J W Evans, Ben Goldacre, Catherine H Smith, and Sinéad M Langan. Risk of severe COVID-19 outcomes associated with immune-mediated inflammatory diseases and immune-modifying therapies: a nationwide cohort study in the OpenSAFELY platform. The Lancet Rheumatology, 4(7):e490–e506, jul 2022. doi: 10.1016/s2665-9913(22)00098-4. URL https://doi.org/10.1016%2Fs2665-9913%2822%2900098-4.

[29] Anna Schultze, Alex J Walker, Brian MacKenna, Caroline E Morton, Krishnan Bhaskaran, Jeremy P Brown, Christopher T Rentsch, Elizabeth Williamson, Henry Drysdale, Richard Croker, Seb Bacon, William Hulme, Chris Bates, Helen J Curtis, Amir Mehrkar, David Evans, Peter Inglesby, Jonathan Cockburn, Helen I McDonald, Laurie Tomlinson, Rohini Mathur, Kevin Wing, Angel Y S Wong, Harriet Forbes, John Parry, Frank Hester, Sam Harper, Stephen J W Evans, Jennifer Quint, Liam Smeeth, Ian J Douglas, and Ben Goldacre. Risk of COVID-19-related death among patients with chronic obstructive pulmonary disease or asthma prescribed inhaled corticosteroids: an observational cohort study using the OpenSAFELY platform. The Lancet Respiratory Medicine, 8(11):1106–1120, nov 2020. doi: 10.1016/s2213-2600(20)30415-x. URL https://doi.org/10.1016%2Fs2213-2600%2820%2930415-x.

[30] Linda Nab, Edward PK Parker, Colm D Andrews, William J Hulme, Louis Fisher, Jessica Morley, Amir Mehrkar, Brian MacKenna, Peter Inglesby, Caroline E Morton, Sebastian CJ Bacon, George Hickman, David Evans, Tom Ward, Rebecca M Smith, Simon Davey, Iain Dillingham, Steven Maude, Ben FC Butler-Cole, Thomas O’Dwyer, Catherine L Stables, Lucy Bridges, Christopher Bates, Jonathan Cockburn, John Parry, Frank Hester, Sam Harper, Bang Zheng, Elizabeth J Williamson, Rosalind M Eggo, Stephen Evans, Ben Goldacre, Laurie A Tomlinson, and Alex J Walker. Changes in covid-19-related mortality across key demographic and clinical subgroups: an observational cohort study using the opensafely platform on 18 million adults in england. medRxiv, 2022. doi: 10.1101/2022.07.30.22278161. URL https://www.medrxiv.org/content/early/2022/08/02/2022.07.30.22278161.

[31] Krishnan Bhaskaran, Sebastian Bacon, Stephen JW Evans, Chris J Bates, Christopher T Rentsch, Brian MacKenna, Laurie Tomlinson, Alex J Walker, Anna Schultze, Caroline E Morton, Daniel Grint, Amir Mehrkar, Rosalind M Eggo, Peter Inglesby, Ian J Douglas, Helen I McDonald, Jonathan Cockburn, Elizabeth J Williamson, David Evans, Helen J Curtis, William J Hulme, John Parry, Frank Hester, Sam Harper, David Spiegelhalter, Liam Smeeth, and Ben Goldacre. Factors associated with deaths due to COVID-19 versus other causes: population-based cohort analysis of UK primary care data and linked national death registrations within the OpenSAFELY platform. The Lancet Regional Health - Europe, 6:100109, jul 2021. doi: 10.1016/j.lanepe.2021.100109. URL https://doi.org/10.1016%2Fj.lanepe.2021.100109.

[32] Truveta. Truveta Platform, 2022. URL https://www.truveta.com/.

[33] M. P. Moutschen, A. J. Scheen, and P. J. Lefebvre. Impaired immune responses in diabetes mellitus: analysis of the factors and mechanisms involved. Relevance to the increased susceptibility of diabetic patients to specific infections. Diabete & Metabolisme, 18(3):187–201, June 1992. ISSN 0338-1684.

[34] David N. O’Dwyer, Robert P. Dickson, and Bethany B. Moore. The Lung Microbiome, Immunity, and the Pathogenesis of Chronic Lung Disease. Journal of Immunology (Baltimore, Md.: 1950), 196(12):4839–4847, June 2016. ISSN 1550-6606. doi: 10.4049/jimmunol.1600279.

[35] Nosratola D. Vaziri, Madeleine V. Pahl, Albert Crum, and Keith Norris. Effect of uremia on structure and function of immune system. Journal of Renal Nutrition: The Official Journal of the Council on Renal Nutrition of the National Kidney Foundation, 22(1):149–156, January 2012. ISSN 1532-8503. doi: 10.1053/j.jrn.2011.10.020.

[36] A. Elixhauser, C. Steiner, D. R. Harris, and R. M. Coffey. Comorbidity measures for use with administrative data. Medical Care, 36(1):8–27, January 1998. ISSN 0025-7079. doi: 10.1097/00005650-199801000-00004.

[37] Hirotugu Akaike. A new look at the statistical model identification. IEEE transactions on automatic control, 19(6):716–723, 1974.

[38] Joseph E Cavanaugh. Unifying the derivations for the akaike and corrected akaike information criteria. Statistics & Probability Letters, 33(2):201–208, 1997.

[39] Kenneth P. Burnham and David R. Anderson. Model Selection and Multimodel Inference. Springer, 2007. ISBN 978-0-387-22456-5.

[40] Mark J Brewer, Adam Butler, and Susan L Cooksley. The relative performance of aic, aicc and bic in the presence of unobserved heterogeneity. Methods in Ecology and Evolution, 7(6): 679–692, 2016.

[41] Russell V. Lenth, Paul Buerkner, Maxime Herve, Maarten Jung, Jonathon Love, Fernando Miguez, Hannes Riebl, and Henrik Singmann. emmeans: Estimated Marginal Means, aka Least-Squares Means, September 2022. URL https://CRAN.R-project.org/package=emmeans.

[42] Andrew Gelman, Jennifer Hill, and Aki Vehtari. Regression and other stories. Analytical methods for social research. Cambridge University Press, Cambridge New York, NY Port Melbourne, VIC New Delhi Singapore, 2021. ISBN 978-1-107-67651-0978-1-107-02398-7. doi: 10.1017/9781139161879.

[43] R Core Team. R: A Language and Environment for Statistical Computing. R Foundation for Statistical Computing, Vienna, Austria, 2022. URL https://www.R-project.org/.

[44] Neal Richardson, Ian Cook, Nic Crane, Dewey Dunnington, Romain François, Jonathan Keane, Dragos Moldovan-Grünfeld, Jeroen Ooms, Javier Luraschi, Karl Dunkle Werner, Jeffrey Wong, and Apache Arrow. arrow: Integration to ‘Apache’ ‘Arrow’, October 2022. URL https://CRAN.R-project.org/package=arrow.

[45] David Robinson, Alex Hayes, Simon Couch [aut, cre, RStudio, Indrajeet Patil, Derek Chiu, Matthieu Gomez, Boris Demeshev, Dieter Menne, Benjamin Nutter, Luke Johnston, Ben Bolker, Francois Briatte, Jeffrey Arnold, Jonah Gabry, Luciano Selzer, Gavin Simpson, Jens Preussner, Jay Hesselberth, Hadley Wickham, Matthew Lincoln, Alessandro Gasparini, Lukasz Komsta, Frederick Novometsky, Wilson Freitas, Michelle Evans, Jason Cory Brunson, Simon Jackson, Ben Whalley, Karissa Whiting, Yves Rosseel, Michael Kuehn, Jorge Cimentada, Erle Holgersen, Karl Dunkle Werner, Ethan Christensen, Steven Pav, Paul PJ, Ben Schneider, Patrick Kennedy Medina, Jason Muhlenkamp, Matt Lehman, Bill Denney, Nic Crane, Andrew Bates, Vincent Arel-Bundock, Hideaki Hayashi, Luis Tobalina, Annie Wang, Wei Yang Tham, Clara Wang, Abby Smith, Jasper Cooper, E. Auden Krauska, Alex Wang, Malcolm Barrett, Charles Gray, Jared Wilber, Vilmantas Gegzna, Eduard Szoecs, Frederik Aust, Angus Moore, Nick Williams, Marius Barth, Bruna Wundervald, Joyce Cahoon, Grant McDermott, Kevin Zarca, Shiro Kuriwaki, Lukas Wallrich, James Martherus, Chuliang Xiao, Joseph Larmarange, Max Kuhn, Michal Bojanowski, Hakon Malmedal, Clara Wang, Sergio Oller, Luke Sonnet, Jim Hester, Cory Brunson, Ben Schneider, Bernie Gray, Mara Averick, Aaron Jacobs, Andreas Bender, Sven Templer, Paul-Christian Buerkner, Matthew Kay, Erwan Le Pennec, Johan Junkka, Hao Zhu, Benjamin Soltoff, Zoe Wilkinson Saldana, Tyler Littlefield, Charles T. Gray, Shabbh E. Banks, Serina Robinson, Roger Bivand, Riinu Ots, Nicholas Williams, Nina Jakobsen, Michael Weylandt, Lisa Lendway, Karl Hailperin, Josue Rodriguez, Jenny Bryan, Chris Jarvis, Greg Macfarlane, Brian Mannakee, Drew Tyre, Shreyas Singh, Laurens Geffert, Hong Ooi, Henrik Bengtsson, Eduard Szocs, David Hugh-Jones, Matthieu Stigler, Hugo Tavares, R. Willem Vervoort, Brenton M. Wiernik, Josh Yamamoto, Jasme Lee Sanders, Daniel D. Sjoberg, and Alex Reinhart. broom: Convert Statistical Objects into Tidy Tibbles, August 2022. URL https://CRAN.R-project.org/package=broom.

[46] Hadley Wickham, Romain François, Lionel Henry, and Kirill Müller. dplyr: A Grammar of Data Manipulation, 2022. URL https://CRAN.R-project.org/package=dplyr.

[47] Hadley Wickham. ggplot2: Elegant Graphics for Data Analysis, 2016. URL https://ggplot 2.tidyverse.org.

[48] Sam Firke, Bill Denney, Chris Haid, Ryan Knight, Malte Grosser, and Jonathan Zadra. janitor: Simple Tools for Examining and Cleaning Dirty Data, January 2021. URL https://CRAN.R-project.org/package=janitor.

[49] Lionel Henry, Hadley Wickham, and RStudio. purrr: Functional Programming Tools, October 2022. URL https://CRAN.R-project.org/package=purrr.

[50] Lionel Henry, Hadley Wickham, mikefc (Hash implementation based on Mike’s xxhashlite), Yann Collet (Author of the embedded xxHash library), and RStudio. rlang: Functions for Base Types and Core R and ‘Tidyverse’ Features, September 2022. URL https://CRAN.R-project.org/package=rlang.

[51] Hadley Wickham and RStudio. stringr: Simple, Consistent Wrappers for Common String Operations, August 2022. URL https://CRAN.R-project.org/package=stringr.

[52] Terry M. Therneau, Thomas Lumley (original S.->R port and R. maintainer until 2009), Atkinson Elizabeth, and Crowson Cynthia. survival: Survival Analysis, August 2022. URL https://CRAN.R-project.org/package=survival.

[53] Benjamin Rich. table1: Tables of Descriptive Statistics in HTML, 2021. URL https://CRAN.R-project.org/package=table1. R package version 1.4.2.

[54] Kirill Müller, Hadley Wickham, Romain Francois, Jennifer Bryan, and RStudio. tibble: Simple Data Frames, July 2022. URL https://CRAN.R-project.org/package=tibble.

[55] Hadley Wickham, Maximilian Girlich, and RStudio. tidyr: Tidy Messy Data, September 2022. URL https://CRAN.R-project.org/package=tidyr.

[56] Edwin S. Wong, Virginia Wang, Chuan-Fen Liu, Paul L. Hebert, and Matthew L. Maciejewski. Do veterans health administration enrollees generalize to other populations? Medical Care Research and Review, 2015. doi: 10.1177/1077558715617382.

[57] William R. Hersh, Mark G. Weiner, Peter J. Embi, Judith R. Logan, Philip R.O. Payne, Elmer V. Bernstam, Harold P. Lehmann, George Hripcsak, Timothy H. Hartzog, James J. Cimino, and Joel H. Saltz. Caveats for the use of operational electronic health record data in comparative effectiveness research. Medical Care, 51:S30–S37, 2013. doi: 10.1097/mlr.0b013e31829b1dbd.

[58] J Marc Overhage and Lauren M Overhage. Sensible use of observational clinical data. Statistical Methods in Medical Research, 22(1):7–13, 2013. doi: 10.1177/0962280211403598.

[59] Clemens Scott Kruse, Anna Stein, Heather Thomas, and Harmander Kaur. The use of electronic health records to support population health: A systematic review of the literature. Journal of Medical Systems, 42:214, 2018. doi: 10.1007/s10916-018-1075-6.

[60] Peter B. Jensen, Lars J. Jensen, and Søren Brunak. Mining electronic health records: towards better research applications and clinical care. Nature Reviews Genetics, 13:395–405, 2012. doi: 10.1038/nrg3208.

[61] Suchitra Kataria and Vinod Ravindran. Electronic health records: A critical appraisal of strengths and limitations. Journal of the Royal College of Physicians of Edinburgh, 50(3):262– 268, 2020. doi: 10.4997/jrcpe.2020.309. URL https://doi.org/10.4997/jrcpe.2020.309. PMID: 32936099.

[62] JANE M. Carrington and JUDITH A. Effken. Strengths and limitations of the electronic health record for documenting clinical events. CIN: Computers, Informatics, Nursing, 29(6):360–367, 2011. doi: 10.1097/ncn.0b013e3181fc4139.

