## Supplement Material for "Risk of COVID-19 breakthrough infection and hospitalization in individuals with comorbidities"

### Supplemental Material for: Risk of COVID-19 breakthrough infection and hospitalization in individuals with comorbidities

Peter D Smits, PhD[1], Samuel Gratzl, PhD[1], Michael Simonov, MD[1,2], Senthil K Nachimuthu, MD, PhD[1], Brianna M Goodwin Cartwright, MS[1], Michael D Wang, MD[1], Charlotte Baker, DrPH, MPH, CPH[1], Patricia Rodriguez, PhD[1], Mackenzie Bogiages[1], Benjamin M. Althouse, PhD, ScM[1,3,4], and Nicholas L Stucky, MD, PhD[1]

[1]Truveta, Inc, Bellevue, WA
[2]Yale School of Medicine, New Haven, CT
[3]University of Washington, Seattle, Washington
[4]New Mexico State University, Las Cruces, New Mexico
*

5.2 Logistic regression model of hospitalization following breakthrough COVID-19 infection239

### 1 CVX codes for COVID-19 vaccination

Table 1: Concept codes used to define vaccination for COVID-19.

| CodeSystem | ConceptCode | ConceptName | Manufacturer |
|---|---|---|---|
| CVX | 208 | SARS-COV-2 (COVID-19) vaccine, mRNA, spike protein, LNP, preservative free, 30 mcg/0.3mL dose | Pfizer |
| CVX | 217 | SARS-COV-2 (COVID-19) vaccine, mRNA, spike protein, LNP, preservative free, 30 mcg/0.3 mL dose, tris-sucrose formulation | Pfizer |
| CVX | 218 | SARS-COV-2 (COVID-19) vaccine, mRNA, spike protein, LNP, preservative free, 10 mcg/0.2 mL dose, tris-sucrose formulation | Pfizer |
| CVX | 219 | SARS-COV-2 (COVID-19) vaccine, mRNA, spike protein, LNP, preservative free, 3 mcg/0.2 mL dose, tris-sucrose formulation | Pfizer |
| CVX | 300 | SARS-COV-2 (COVID-19) vaccine, mRNA, spike protein, LNP, bivalent booster, preservative free, 30 mcg/0.3mL dose, tris-sucrose formulation | Pfizer |
| CVX | 301 | SARS-COV-2 (COVID-19) vaccine, mRNA, spike protein, LNP, bivalent booster, preservative free, 10 mcg/0.2 mL dose, tris-sucrose formulation | Pfizer |
| CVX | 207 | SARS-COV-2 (COVID-19) vaccine, mRNA, spike protein, LNP, preservative free, 100 mcg/0.5mL dose | Moderna |
| CVX | 221 | COVID-19, mRNA, LNP-S, PF, 50 mcg/0.5 mL dose | Moderna |
| CVX | 228 | COVID-19, mRNA, LNP-S, PF, pediatric 25 mcg/0.25 mL dose | Moderna |
| CVX | 229 | SARS-COV-2 (COVID-19) vaccine, mRNA, spike protein, LNP, bivalent booster, preservative free, 50 mcg/0.5 mL or 25mcg/0.25mL dose | Moderna |

### 2 ICD-10CM, SNOMEDCT, and LOINC codes for COVID-19 positive

#### 2.1 COVID-19 antigen or PCR test codes

Table 2: Concept codes used to define a positive COVID-19 test.

| CodeSystem | ConceptCode | ConceptName |
|---|---|---|
| LOINC | 99596-9 | SARS-CoV-2 (COVID-19) N protein IgG Ab [Presence] in Serum or Plasma by Immunoassay |
| LOINC | 99597-7 | SARS-CoV-2 (COVID-19) S protein IgG Ab [Presence] in Serum or Plasma by Immunoassay |
| LOINC | 99772-6 | SARS-CoV-2 stimulated gamma interferon [Interpretation] in Blood Qualitative |
| SNOMED CT | 871555000 | Detection of ribonucleic acid of severe acute respiratory syndrome coronavirus 2 |
| SNOMED CT | 871556004 | Detection of ribonucleic acid of severe acute respiratory syndrome coronavirus 2 in nasopharyngeal swab |
| SNOMED CT | 871557008 | Detection of ribonucleic acid of severe acute respiratory syndrome coronavirus 2 in oropharyngeal swab |
| SNOMED CT | 871558003 | Detection of ribonucleic acid of severe acute respiratory syndrome coronavirus 2 in sputum |
| SNOMED CT | 871559006 | Detection of ribonucleic acid of severe acute respiratory syndrome coronavirus 2 in bronchoalveolar lavage fluid |
| SNOMED CT | 871560001 | Detection of ribonucleic acid of severe acute respiratory syndrome coronavirus 2 using polymerase chain reaction |
| SNOMED CT | 871562009 | Detection of severe acute respiratory syndrome coronavirus 2 |
| SNOMED CT | 1240471000000102 | Measurement of severe acute respiratory syndrome coronavirus 2 antigen |
| LOINC | 94306-8 | SARS-CoV-2 (COVID-19) RNA panel - Specimen by NAA with probe detection |
| LOINC | 94307-6 | SARS-CoV-2 (COVID-19) N gene [Presence] in Specimen by Nucleic acid amplification using CDC primer-probe set N1 |
| LOINC | 94308-4 | SARS-CoV-2 (COVID-19) N gene [Presence] in Specimen by Nucleic acid amplification using CDC primer-probe set N2 |
| LOINC | 94309-2 | SARS-CoV-2 (COVID-19) RNA [Presence] in Specimen by NAA with probe detection |
| LOINC | 94314-2 | SARS-CoV-2 (COVID-19) RdRp gene [Presence] in Specimen by NAA with probe detection |
| LOINC | 94315-9 | SARS-related coronavirus E gene [Presence] in Specimen by NAA with probe detection |

Continued on next page

**Table 2 – continued from previous page**

| CodeSystem | ConceptCode | ConceptName |
|---|---|---|
| LOINC | 94316-7 | SARS-CoV-2 (COVID-19) N gene [Presence] in Specimen by NAA with probe detection |
| LOINC | 94500-6 | SARS-CoV-2 (COVID-19) RNA [Presence] in Respiratory specimen by NAA with probe detection |
| LOINC | 94507-1 | SARS-CoV-2 (COVID-19) IgG Ab [Presence] in Serum, Plasma or Blood by Rapid immunoassay |
| LOINC | 94508-9 | SARS-CoV-2 (COVID-19) IgM Ab [Presence] in Serum, Plasma or Blood by Rapid immunoassay |
| LOINC | 94531-1 | SARS-CoV-2 (COVID-19) RNA panel - Respiratory specimen by NAA with probe detection |
| LOINC | 94533-7 | SARS-CoV-2 (COVID-19) N gene [Presence] in Respiratory specimen by NAA with probe detection |
| LOINC | 94534-5 | SARS-CoV-2 (COVID-19) RdRp gene [Presence] in Respiratory specimen by NAA with probe detection |
| LOINC | 94547-7 | SARS-CoV-2 (COVID-19) IgG+IgM Ab [Presence] in Serum or Plasma by Immunoassay |
| LOINC | 94558-4 | SARS-CoV-2 (COVID-19) Ag [Presence] in Respiratory specimen by Rapid immunoassay |
| LOINC | 94559-2 | SARS-CoV-2 (COVID-19) ORF1ab region [Presence] in Respiratory specimen by NAA with probe detection |
| LOINC | 94562-6 | SARS-CoV-2 (COVID-19) IgA Ab [Presence] in Serum or Plasma by Immunoassay |
| LOINC | 94563-4 | SARS-CoV-2 (COVID-19) IgG Ab [Presence] in Serum or Plasma by Immunoassay |
| LOINC | 94564-2 | SARS-CoV-2 (COVID-19) IgM Ab [Presence] in Serum or Plasma by Immunoassay |
| LOINC | 94565-9 | SARS-CoV-2 (COVID-19) RNA [Presence] in Nasopharynx by NAA with non-probe detection |
| LOINC | 94639-2 | SARS-CoV-2 (COVID-19) ORF1ab region [Presence] in Specimen by NAA with probe detection |
| LOINC | 94640-0 | SARS-CoV-2 (COVID-19) S gene [Presence] in Respiratory specimen by NAA with probe detection |
| LOINC | 94641-8 | SARS-CoV-2 (COVID-19) S gene [Presence] in Specimen by NAA with probe detection |
| LOINC | 94660-8 | SARS-CoV-2 (COVID-19) RNA [Presence] in Serum or Plasma by NAA with probe detection |
| LOINC | 94756-4 | SARS-CoV-2 (COVID-19) N gene [Presence] in Respiratory specimen by Nucleic acid amplification using CDC primer-probe set N1 |
| LOINC | 94757-2 | SARS-CoV-2 (COVID-19) N gene [Presence] in Respiratory specimen by Nucleic acid amplification using CDC primer-probe set N2 |

Continued on next page

**Table 2 – continued from previous page**

| CodeSystem | ConceptCode | ConceptName |
|---|---|---|
| LOINC | 94759-8 | SARS-CoV-2 (COVID-19) RNA [Presence] in Nasopharynx by NAA with probe detection |
| LOINC | 94760-6 | SARS-CoV-2 (COVID-19) N gene [Presence] in Nasopharynx by NAA with probe detection |
| LOINC | 94761-4 | SARS-CoV-2 (COVID-19) IgG Ab [Presence] in DBS by Immunoassay |
| LOINC | 94762-2 | SARS-CoV-2 (COVID-19) Ab [Presence] in Serum or Plasma by Immunoassay |
| LOINC | 94763-0 | SARS-CoV-2 (COVID-19) [Presence] in Specimen by Organism specific culture |
| LOINC | 94766-3 | SARS-CoV-2 (COVID-19) N gene [Presence] in Serum or Plasma by NAA with probe detection |
| LOINC | 94767-1 | SARS-CoV-2 (COVID-19) S gene [Presence] in Serum or Plasma by NAA with probe detection |
| LOINC | 94768-9 | SARS-CoV-2 (COVID-19) IgA Ab [Presence] in Serum, Plasma or Blood by Rapid immunoassay |
| LOINC | 94822-4 | SARS-CoV-2 (COVID-19) RNA [Presence] in Saliva (oral fluid) by Sequencing |
| LOINC | 94845-5 | SARS-CoV-2 (COVID-19) RNA [Presence] in Saliva (oral fluid) by NAA with probe detection |
| LOINC | 95125-1 | SARS-CoV-2 (COVID-19) IgA+IgM [Presence] in Serum or Plasma by Immunoassay |
| LOINC | 95209-3 | SARS-CoV+SARS-CoV-2 (COVID-19) Ag [Presence] in Respiratory specimen by Rapid immunoassay |
| LOINC | 95406-5 | SARS-CoV-2 (COVID-19) RNA [Presence] in Nose by NAA with probe detection |
| LOINC | 95409-9 | SARS-CoV-2 (COVID-19) N gene [Presence] in Nose by NAA with probe detection |
| LOINC | 95411-5 | SARS-CoV-2 (COVID-19) neutralizing antibody [Presence] in Serum by pVNT |
| LOINC | 95416-4 | SARS-CoV-2 (COVID-19) IgM Ab [Presence] in DBS by Immunoassay |
| LOINC | 95424-8 | SARS-CoV-2 (COVID-19) RNA [Presence] in Respiratory specimen by Sequencing |
| LOINC | 95425-5 | SARS-CoV-2 (COVID-19) N gene [Presence] in Saliva (oral fluid) by NAA with probe detection |
| LOINC | 95542-7 | SARS-CoV-2 (COVID-19) IgG+IgM Ab [Presence] in Serum, Plasma or Blood by Rapid immunoassay |
| LOINC | 95608-6 | SARS-CoV-2 (COVID-19) RNA [Presence] in Respiratory specimen by NAA with non-probe detection |
| LOINC | 95609-4 | SARS-CoV-2 (COVID-19) S gene [Presence] in Respiratory specimen by Sequencing |

Continued on next page

**Table 2 – continued from previous page**

| CodeSystem | ConceptCode | ConceptName |
|---|---|---|
| LOINC | 95823-1 | SARS-related coronavirus E gene [Presence] in Saliva (oral fluid) by NAA with probe detection |
| LOINC | 95824-9 | SARS-CoV-2 (COVID-19) ORF1ab region [Presence] in Saliva (oral fluid) by NAA with probe detection |
| LOINC | 95825-6 | SARS-CoV-2 (COVID-19) Ab [Presence] in DBS by Immunoassay |
| LOINC | 95970-0 | SARS-CoV-2 (COVID-19) specific TCRB gene rearrangements [Presence] in Blood by Sequencing |
| LOINC | 95971-8 | SARS CoV-2 stimulated gamma interferon [Presence] in Blood |
| LOINC | 96091-4 | SARS-CoV-2 (COVID-19) RdRp gene [Presence] in Saliva (oral fluid) by NAA with probe detection |
| LOINC | 96119-3 | SARS-CoV-2 (COVID-19) Ag [Presence] in Upper respiratory specimen by Immunoassay |
| LOINC | 96120-1 | SARS-CoV-2 (COVID-19) RdRp gene [Presence] in Lower respiratory specimen by NAA with probe detection |
| LOINC | 96121-9 | SARS-related coronavirus E gene [Presence] in Lower respiratory specimen by NAA with probe detection |
| LOINC | 96122-7 | SARS-related coronavirus E gene [Presence] in Upper respiratory specimen by NAA with probe detection |
| LOINC | 96123-5 | SARS-CoV-2 (COVID-19) RdRp gene [Presence] in Upper respiratory specimen by NAA with probe detection |
| LOINC | 96448-6 | SARS-CoV-2 (COVID-19) N gene [Presence] in Saliva (oral fluid) by Nucleic acid amplification using CDC primer-probe set N1 |
| LOINC | 96603-6 | SARS-CoV-2 (COVID-19) S protein RBD neutralizing antibody [Presence] in Serum or Plasma by Immunoassay |
| LOINC | 98069-8 | SARS-CoV-2 (COVID-19) Ab [Presence] in Saliva (oral fluid) by Rapid immunoassay |
| LOINC | 98131-6 | SARS-CoV-2 (COVID-19) ORF1b region [Presence] in Respiratory specimen by NAA with probe detection |
| LOINC | 98132-4 | SARS-CoV-2 (COVID-19) ORF1a region [Presence] in Respiratory specimen by NAA with probe detection |
| LOINC | 98493-0 | SARS-CoV-2 (COVID-19) ORF1b region [Presence] in Saliva (oral fluid) by NAA with probe detection |
| LOINC | 98494-8 | SARS-CoV-2 (COVID-19) ORF1a region [Presence] in Saliva (oral fluid) by NAA with probe detection |
| LOINC | 96752-1 | SARS-CoV-2 (COVID-19) S gene mutation [Presence] in Specimen by Molecular genetics method |
| LOINC | 96763-8 | SARS-CoV-2 (COVID-19) E gene [Presence] in Respiratory specimen by NAA with probe detection |
| LOINC | 96765-3 | SARS-CoV-2 (COVID-19) S gene [Presence] in Saliva (oral fluid) by NAA with probe detection |

Continued on next page

**Table 2 – continued from previous page**

| CodeSystem | ConceptCode | ConceptName |
|---|---|---|
| LOINC | 96797-6 | SARS-CoV-2 (COVID-19) RNA [Presence] in Oropharyngeal wash by NAA with probe detection |
| LOINC | 96829-7 | SARS-CoV-2 (COVID-19) RNA [Presence] in Specimen from Donor by NAA with probe detection |
| LOINC | 96957-6 | SARS-CoV-2 (COVID-19) M gene [Presence] in Upper respiratory specimen by NAA with probe detection |
| LOINC | 96958-4 | SARS-CoV-2 (COVID-19) N gene [Presence] in Saliva (oral fluid) by Nucleic acid amplification using CDC primer-probe set N2 |
| LOINC | 96986-5 | SARS-CoV-2 (COVID-19) N gene [Presence] in Nose by NAA with non-probe detection |
| LOINC | 97097-0 | SARS-CoV-2 (COVID-19) Ag [Presence] in Upper respiratory specimen by Rapid immunoassay |
| LOINC | 97098-8 | SARS-CoV-2 (COVID-19) Nsp2 gene [Presence] in Upper respiratory specimen by NAA with probe detection |
| SNOMED CT | 871553007 | Detection of severe acute respiratory syndrome coronavirus 2 antigen |
| LOINC | 94310-0 | SARS-like coronavirus N gene [Presence] in Specimen by NAA with probe detection |
| LOINC | 94311-8 | SARS-CoV-2 (COVID-19) N gene [Cycle Threshold #] in Specimen by Nucleic acid amplification using CDC primer-probe set N1 |
| LOINC | 94312-6 | SARS-CoV-2 (COVID-19) N gene [Cycle Threshold #] in Specimen by Nucleic acid amplification using CDC primer-probe set N2 |
| LOINC | 94313-4 | SARS-like coronavirus N gene [Cycle Threshold #] in Specimen by NAA with probe detection |
| LOINC | 94502-2 | SARS-related coronavirus RNA [Presence] in Respiratory specimen by NAA with probe detection |
| LOINC | 94509-7 | SARS-related coronavirus E gene [Cycle Threshold #] in Specimen by NAA with probe detection |
| LOINC | 94510-5 | SARS-CoV-2 (COVID-19) N gene [Cycle Threshold #] in Specimen by NAA with probe detection |
| LOINC | 94511-3 | SARS-CoV-2 (COVID-19) ORF1ab region [Cycle Threshold #] in Specimen by NAA with probe detection |
| LOINC | 94532-9 | SARS-related coronavirus+MERS coronavirus RNA [Presence] in Respiratory specimen by NAA with probe detection |
| LOINC | 94642-6 | SARS-CoV-2 (COVID-19) S gene [Cycle Threshold #] in Respiratory specimen by NAA with probe detection |
| LOINC | 94643-4 | SARS-CoV-2 (COVID-19) S gene [Cycle Threshold #] in Specimen by NAA with probe detection |
| LOINC | 94644-2 | SARS-CoV-2 (COVID-19) ORF1ab region [Cycle Threshold #] in Respiratory specimen by NAA with probe detection |

Continued on next page

**Table 2 – continued from previous page**

| CodeSystem | ConceptCode | ConceptName |
|---|---|---|
| LOINC | 94645-9 | SARS-CoV-2 (COVID-19) RdRp gene [Cycle Threshold #] in Specimen by NAA with probe detection |
| LOINC | 94646-7 | SARS-CoV-2 (COVID-19) RdRp gene [Cycle Threshold #] in Respiratory specimen by NAA with probe detection |
| LOINC | 94647-5 | SARS-related coronavirus RNA [Presence] in Specimen by NAA with probe detection |
| LOINC | 94745-7 | SARS-CoV-2 (COVID-19) RNA [Cycle Threshold #] in Respiratory specimen by NAA with probe detection |
| LOINC | 94746-5 | SARS-CoV-2 (COVID-19) RNA [Cycle Threshold #] in Specimen by NAA with probe detection |
| LOINC | 94758-0 | SARS-related coronavirus E gene [Presence] in Respiratory specimen by NAA with probe detection |
| LOINC | 94819-0 | SARS-CoV-2 (COVID-19) RNA [Log #/volume] (viral load) in Specimen by NAA with probe detection |
| LOINC | 95521-1 | SARS-CoV-2 (COVID-19) N gene [#/volume] (viral load) in Respiratory specimen by NAA with probe detection |
| LOINC | 95522-9 | SARS-CoV-2 (COVID-19) N gene [Log #/volume] (viral load) in Respiratory specimen by NAA with probe detection |
| LOINC | 96764-6 | SARS-CoV-2 (COVID-19) E gene [Cycle Threshold #] in Respiratory specimen by NAA with probe detection |
| LOINC | 97099-6 | Influenza virus A and B and SARS-CoV-2 (COVID-19) Ag panel - Upper respiratory specimen by Rapid immunoassay |

#### 2.2 Diagnosis codes

Table 3: Concept codes used to define a positive COVID-19 diagnosis.

| CodeSystem | ConceptCode | ConceptName |
|---|---|---|
| LOINC | 99596-9 | SARS-CoV-2 (COVID-19) N protein IgG Ab [Presence] in Serum or Plasma by Immunoassay |
| LOINC | 99597-7 | SARS-CoV-2 (COVID-19) S protein IgG Ab [Presence] in Serum or Plasma by Immunoassay |
| LOINC | 99772-6 | SARS-CoV-2 stimulated gamma interferon [Interpretation] in Blood Qualitative |
| SNOMED CT | 871555000 | Detection of ribonucleic acid of severe acute respiratory syndrome coronavirus 2 |
| SNOMED CT | 871556004 | Detection of ribonucleic acid of severe acute respiratory syndrome coronavirus 2 in nasopharyngeal swab |
| SNOMED CT | 871557008 | Detection of ribonucleic acid of severe acute respiratory syndrome coronavirus 2 in oropharyngeal swab |
| SNOMED CT | 871558003 | Detection of ribonucleic acid of severe acute respiratory syndrome coronavirus 2 in sputum |
| SNOMED CT | 871559006 | Detection of ribonucleic acid of severe acute respiratory syndrome coronavirus 2 in bronchoalveolar lavage fluid |
| SNOMED CT | 871560001 | Detection of ribonucleic acid of severe acute respiratory syndrome coronavirus 2 using polymerase chain reaction |
| SNOMED CT | 871562009 | Detection of severe acute respiratory syndrome coronavirus 2 |
| SNOMED CT | 1240471000000102 | Measurement of severe acute respiratory syndrome coronavirus 2 antigen |
| LOINC | 94306-8 | SARS-CoV-2 (COVID-19) RNA panel - Specimen by NAA with probe detection |
| LOINC | 94307-6 | SARS-CoV-2 (COVID-19) N gene [Presence] in Specimen by Nucleic acid amplification using CDC primer-probe set N1 |
| LOINC | 94308-4 | SARS-CoV-2 (COVID-19) N gene [Presence] in Specimen by Nucleic acid amplification using CDC primer-probe set N2 |
| LOINC | 94309-2 | SARS-CoV-2 (COVID-19) RNA [Presence] in Specimen by NAA with probe detection |
| LOINC | 94314-2 | SARS-CoV-2 (COVID-19) RdRp gene [Presence] in Specimen by NAA with probe detection |
| LOINC | 94315-9 | SARS-related coronavirus E gene [Presence] in Specimen by NAA with probe detection |
| LOINC | 94316-7 | SARS-CoV-2 (COVID-19) N gene [Presence] in Specimen by NAA with probe detection |

Continued on next page

**Table 3 – continued from previous page**

| CodeSystem | ConceptCode | ConceptName |
|---|---|---|
| LOINC | 94500-6 | SARS-CoV-2 (COVID-19) RNA [Presence] in Respiratory specimen by NAA with probe detection |
| LOINC | 94507-1 | SARS-CoV-2 (COVID-19) IgG Ab [Presence] in Serum, Plasma or Blood by Rapid immunoassay |
| LOINC | 94508-9 | SARS-CoV-2 (COVID-19) IgM Ab [Presence] in Serum, Plasma or Blood by Rapid immunoassay |
| LOINC | 94531-1 | SARS-CoV-2 (COVID-19) RNA panel - Respiratory specimen by NAA with probe detection |
| LOINC | 94533-7 | SARS-CoV-2 (COVID-19) N gene [Presence] in Respiratory specimen by NAA with probe detection |
| LOINC | 94534-5 | SARS-CoV-2 (COVID-19) RdRp gene [Presence] in Respiratory specimen by NAA with probe detection |
| LOINC | 94547-7 | SARS-CoV-2 (COVID-19) IgG+IgM Ab [Presence] in Serum or Plasma by Immunoassay |
| LOINC | 94558-4 | SARS-CoV-2 (COVID-19) Ag [Presence] in Respiratory specimen by Rapid immunoassay |
| LOINC | 94559-2 | SARS-CoV-2 (COVID-19) ORF1ab region [Presence] in Respiratory specimen by NAA with probe detection |
| LOINC | 94562-6 | SARS-CoV-2 (COVID-19) IgA Ab [Presence] in Serum or Plasma by Immunoassay |
| LOINC | 94563-4 | SARS-CoV-2 (COVID-19) IgG Ab [Presence] in Serum or Plasma by Immunoassay |
| LOINC | 94564-2 | SARS-CoV-2 (COVID-19) IgM Ab [Presence] in Serum or Plasma by Immunoassay |
| LOINC | 94565-9 | SARS-CoV-2 (COVID-19) RNA [Presence] in Nasopharynx by NAA with non-probe detection |
| LOINC | 94639-2 | SARS-CoV-2 (COVID-19) ORF1ab region [Presence] in Specimen by NAA with probe detection |
| LOINC | 94640-0 | SARS-CoV-2 (COVID-19) S gene [Presence] in Respiratory specimen by NAA with probe detection |
| LOINC | 94641-8 | SARS-CoV-2 (COVID-19) S gene [Presence] in Specimen by NAA with probe detection |
| LOINC | 94660-8 | SARS-CoV-2 (COVID-19) RNA [Presence] in Serum or Plasma by NAA with probe detection |
| LOINC | 94756-4 | SARS-CoV-2 (COVID-19) N gene [Presence] in Respiratory specimen by Nucleic acid amplification using CDC primer-probe set N1 |
| LOINC | 94757-2 | SARS-CoV-2 (COVID-19) N gene [Presence] in Respiratory specimen by Nucleic acid amplification using CDC primer-probe set N2 |
| LOINC | 94759-8 | SARS-CoV-2 (COVID-19) RNA [Presence] in Nasopharynx by NAA with probe detection |

Continued on next page

**Table 3 – continued from previous page**

| CodeSystem | ConceptCode | ConceptName |
|---|---|---|
| LOINC | 94760-6 | SARS-CoV-2 (COVID-19) N gene [Presence] in Nasopharynx by NAA with probe detection |
| LOINC | 94761-4 | SARS-CoV-2 (COVID-19) IgG Ab [Presence] in DBS by Immunoassay |
| LOINC | 94762-2 | SARS-CoV-2 (COVID-19) Ab [Presence] in Serum or Plasma by Immunoassay |
| LOINC | 94763-0 | SARS-CoV-2 (COVID-19) [Presence] in Specimen by Organism specific culture |
| LOINC | 94766-3 | SARS-CoV-2 (COVID-19) N gene [Presence] in Serum or Plasma by NAA with probe detection |
| LOINC | 94767-1 | SARS-CoV-2 (COVID-19) S gene [Presence] in Serum or Plasma by NAA with probe detection |
| LOINC | 94768-9 | SARS-CoV-2 (COVID-19) IgA Ab [Presence] in Serum, Plasma or Blood by Rapid immunoassay |
| LOINC | 94822-4 | SARS-CoV-2 (COVID-19) RNA [Presence] in Saliva (oral fluid) by Sequencing |
| LOINC | 94845-5 | SARS-CoV-2 (COVID-19) RNA [Presence] in Saliva (oral fluid) by NAA with probe detection |
| LOINC | 95125-1 | SARS-CoV-2 (COVID-19) IgA+IgM [Presence] in Serum or Plasma by Immunoassay |
| LOINC | 95209-3 | SARS-CoV+SARS-CoV-2 (COVID-19) Ag [Presence] in Respiratory specimen by Rapid immunoassay |
| LOINC | 95406-5 | SARS-CoV-2 (COVID-19) RNA [Presence] in Nose by NAA with probe detection |
| LOINC | 95409-9 | SARS-CoV-2 (COVID-19) N gene [Presence] in Nose by NAA with probe detection |
| LOINC | 95411-5 | SARS-CoV-2 (COVID-19) neutralizing antibody [Presence] in Serum by pVNT |
| LOINC | 95416-4 | SARS-CoV-2 (COVID-19) IgM Ab [Presence] in DBS by Immunoassay |
| LOINC | 95424-8 | SARS-CoV-2 (COVID-19) RNA [Presence] in Respiratory specimen by Sequencing |
| LOINC | 95425-5 | SARS-CoV-2 (COVID-19) N gene [Presence] in Saliva (oral fluid) by NAA with probe detection |
| LOINC | 95542-7 | SARS-CoV-2 (COVID-19) IgG+IgM Ab [Presence] in Serum, Plasma or Blood by Rapid immunoassay |
| LOINC | 95608-6 | SARS-CoV-2 (COVID-19) RNA [Presence] in Respiratory specimen by NAA with non-probe detection |
| LOINC | 95609-4 | SARS-CoV-2 (COVID-19) S gene [Presence] in Respiratory specimen by Sequencing |
| LOINC | 95823-1 | SARS-related coronavirus E gene [Presence] in Saliva (oral fluid) by NAA with probe detection |

Continued on next page

**Table 3 – continued from previous page**

| CodeSystem | ConceptCode | ConceptName |
|---|---|---|
| LOINC | 95824-9 | SARS-CoV-2 (COVID-19) ORF1ab region [Presence] in Saliva (oral fluid) by NAA with probe detection |
| LOINC | 95825-6 | SARS-CoV-2 (COVID-19) Ab [Presence] in DBS by Immunoassay |
| LOINC | 95970-0 | SARS-CoV-2 (COVID-19) specific TCRB gene rearrangements [Presence] in Blood by Sequencing |
| LOINC | 95971-8 | SARS CoV-2 stimulated gamma interferon [Presence] in Blood |
| LOINC | 96091-4 | SARS-CoV-2 (COVID-19) RdRp gene [Presence] in Saliva (oral fluid) by NAA with probe detection |
| LOINC | 96119-3 | SARS-CoV-2 (COVID-19) Ag [Presence] in Upper respiratory specimen by Immunoassay |
| LOINC | 96120-1 | SARS-CoV-2 (COVID-19) RdRp gene [Presence] in Lower respiratory specimen by NAA with probe detection |
| LOINC | 96121-9 | SARS-related coronavirus E gene [Presence] in Lower respiratory specimen by NAA with probe detection |
| LOINC | 96122-7 | SARS-related coronavirus E gene [Presence] in Upper respiratory specimen by NAA with probe detection |
| LOINC | 96123-5 | SARS-CoV-2 (COVID-19) RdRp gene [Presence] in Upper respiratory specimen by NAA with probe detection |
| LOINC | 96448-6 | SARS-CoV-2 (COVID-19) N gene [Presence] in Saliva (oral fluid) by Nucleic acid amplification using CDC primer-probe set N1 |
| LOINC | 96603-6 | SARS-CoV-2 (COVID-19) S protein RBD neutralizing antibody [Presence] in Serum or Plasma by Immunoassay |
| LOINC | 98069-8 | SARS-CoV-2 (COVID-19) Ab [Presence] in Saliva (oral fluid) by Rapid immunoassay |
| LOINC | 98131-6 | SARS-CoV-2 (COVID-19) ORF1b region [Presence] in Respiratory specimen by NAA with probe detection |
| LOINC | 98132-4 | SARS-CoV-2 (COVID-19) ORF1a region [Presence] in Respiratory specimen by NAA with probe detection |
| LOINC | 98493-0 | SARS-CoV-2 (COVID-19) ORF1b region [Presence] in Saliva (oral fluid) by NAA with probe detection |
| LOINC | 98494-8 | SARS-CoV-2 (COVID-19) ORF1a region [Presence] in Saliva (oral fluid) by NAA with probe detection |
| LOINC | 96752-1 | SARS-CoV-2 (COVID-19) S gene mutation [Presence] in Specimen by Molecular genetics method |
| LOINC | 96763-8 | SARS-CoV-2 (COVID-19) E gene [Presence] in Respiratory specimen by NAA with probe detection |
| LOINC | 96765-3 | SARS-CoV-2 (COVID-19) S gene [Presence] in Saliva (oral fluid) by NAA with probe detection |
| LOINC | 96797-6 | SARS-CoV-2 (COVID-19) RNA [Presence] in Oropharyngeal wash by NAA with probe detection |

Continued on next page

**Table 3 – continued from previous page**

| CodeSystem | ConceptCode | ConceptName |
|---|---|---|
| LOINC | 96829-7 | SARS-CoV-2 (COVID-19) RNA [Presence] in Specimen from Donor by NAA with probe detection |
| LOINC | 96957-6 | SARS-CoV-2 (COVID-19) M gene [Presence] in Upper respiratory specimen by NAA with probe detection |
| LOINC | 96958-4 | SARS-CoV-2 (COVID-19) N gene [Presence] in Saliva (oral fluid) by Nucleic acid amplification using CDC primer-probe set N2 |
| LOINC | 96986-5 | SARS-CoV-2 (COVID-19) N gene [Presence] in Nose by NAA with non-probe detection |
| LOINC | 97097-0 | SARS-CoV-2 (COVID-19) Ag [Presence] in Upper respiratory specimen by Rapid immunoassay |
| LOINC | 97098-8 | SARS-CoV-2 (COVID-19) Nsp2 gene [Presence] in Upper respiratory specimen by NAA with probe detection |
| SNOMED CT | 871553007 | Detection of severe acute respiratory syndrome coronavirus 2 antigen |
| LOINC | 94310-0 | SARS-like coronavirus N gene [Presence] in Specimen by NAA with probe detection |
| LOINC | 94311-8 | SARS-CoV-2 (COVID-19) N gene [Cycle Threshold #] in Specimen by Nucleic acid amplification using CDC primer-probe set N1 |
| LOINC | 94312-6 | SARS-CoV-2 (COVID-19) N gene [Cycle Threshold #] in Specimen by Nucleic acid amplification using CDC primer-probe set N2 |
| LOINC | 94313-4 | SARS-like coronavirus N gene [Cycle Threshold #] in Specimen by NAA with probe detection |
| LOINC | 94502-2 | SARS-related coronavirus RNA [Presence] in Respiratory specimen by NAA with probe detection |
| LOINC | 94509-7 | SARS-related coronavirus E gene [Cycle Threshold #] in Specimen by NAA with probe detection |
| LOINC | 94510-5 | SARS-CoV-2 (COVID-19) N gene [Cycle Threshold #] in Specimen by NAA with probe detection |
| LOINC | 94511-3 | SARS-CoV-2 (COVID-19) ORF1ab region [Cycle Threshold #] in Specimen by NAA with probe detection |
| LOINC | 94532-9 | SARS-related coronavirus+MERS coronavirus RNA [Presence] in Respiratory specimen by NAA with probe detection |
| LOINC | 94642-6 | SARS-CoV-2 (COVID-19) S gene [Cycle Threshold #] in Respiratory specimen by NAA with probe detection |
| LOINC | 94643-4 | SARS-CoV-2 (COVID-19) S gene [Cycle Threshold #] in Specimen by NAA with probe detection |
| LOINC | 94644-2 | SARS-CoV-2 (COVID-19) ORF1ab region [Cycle Threshold #] in Respiratory specimen by NAA with probe detection |
| LOINC | 94645-9 | SARS-CoV-2 (COVID-19) RdRp gene [Cycle Threshold #] in Specimen by NAA with probe detection |

Continued on next page

**Table 3 – continued from previous page**

| CodeSystem | ConceptCode | ConceptName |
|---|---|---|
| LOINC | 94646-7 | SARS-CoV-2 (COVID-19) RdRp gene [Cycle Threshold #] in Respiratory specimen by NAA with probe detection |
| LOINC | 94647-5 | SARS-related coronavirus RNA [Presence] in Specimen by NAA with probe detection |
| LOINC | 94745-7 | SARS-CoV-2 (COVID-19) RNA [Cycle Threshold #] in Respiratory specimen by NAA with probe detection |
| LOINC | 94746-5 | SARS-CoV-2 (COVID-19) RNA [Cycle Threshold #] in Specimen by NAA with probe detection |
| LOINC | 94758-0 | SARS-related coronavirus E gene [Presence] in Respiratory specimen by NAA with probe detection |
| LOINC | 94819-0 | SARS-CoV-2 (COVID-19) RNA [Log #/volume] (viral load) in Specimen by NAA with probe detection |
| LOINC | 95521-1 | SARS-CoV-2 (COVID-19) N gene [#/volume] (viral load) in Respiratory specimen by NAA with probe detection |
| LOINC | 95522-9 | SARS-CoV-2 (COVID-19) N gene [Log #/volume] (viral load) in Respiratory specimen by NAA with probe detection |
| LOINC | 96764-6 | SARS-CoV-2 (COVID-19) E gene [Cycle Threshold #] in Respiratory specimen by NAA with probe detection |
| LOINC | 97099-6 | Influenza virus A and B and SARS-CoV-2 (COVID-19) Ag panel - Upper respiratory specimen by Rapid immunoassay |

### 3 ICD-10CM and SNOMEDCT codes for comorbidities

#### 3.1 Chronic kidney disease

Table 4: Concept codes used to define chronic kidney disease.

| CodeSystem | ConceptCode | ConceptName |
|---|---|---|
| ICD10CM | D63.1 | Anemia in chronic kidney disease |
| ICD10CM | E08.22 | Diabetes mellitus due to underlying condition with diabetic chronic kidney disease |
| ICD10CM | E09.22 | Drug or chemical induced diabetes mellitus with diabetic chronic kidney disease |
| ICD10CM | E13.22 | Other specified diabetes mellitus with diabetic chronic kidney disease |
| ICD10CM | I12 | Hypertensive chronic kidney disease |
| ICD10CM | I12.0 | Hypertensive chronic kidney disease with stage 5 chronic kidney disease or end stage renal disease |
| ICD10CM | I12.9 | Hypertensive chronic kidney disease with stage 1 through stage 4 chronic kidney disease, or unspecified chronic kidney disease |
| ICD10CM | I13 | Hypertensive heart and chronic kidney disease |
| ICD10CM | I13.0 | Hypertensive heart and chronic kidney disease with heart failure and stage 1 through stage 4 chronic kidney disease, or unspecified chronic kidney disease |
| ICD10CM | I13.1 | Hypertensive heart and chronic kidney disease without heart failure |
| ICD10CM | I13.10 | Hypertensive heart and chronic kidney disease without heart failure, with stage 1 through stage 4 chronic kidney disease, or unspecified chronic kidney disease |
| ICD10CM | I13.11 | Hypertensive heart and chronic kidney disease without heart failure, with stage 5 chronic kidney disease, or end stage renal disease |
| ICD10CM | I13.2 | Hypertensive heart and chronic kidney disease with heart failure and with stage 5 chronic kidney disease, or end stage renal disease |
| ICD10CM | N18 | Chronic kidney disease (CKD) |
| ICD10CM | O10.211 | Pre-existing hypertensive chronic kidney disease complicating pregnancy, first trimester |
| ICD10CM | O10.212 | Pre-existing hypertensive chronic kidney disease complicating pregnancy, second trimester |
| ICD10CM | O10.213 | Pre-existing hypertensive chronic kidney disease complicating pregnancy, third trimester |
| ICD10CM | O10.219 | Pre-existing hypertensive chronic kidney disease complicating pregnancy, unspecified trimester |

Continued on next page

**Table 4 – continued from previous page**

| CodeSystem | ConceptCode | ConceptName |
|---|---|---|
| ICD10CM | O10.22 | Pre-existing hypertensive chronic kidney disease complicating childbirth |
| ICD10CM | O10.31 | Pre-existing hypertensive heart and chronic kidney disease complicating pregnancy |
| ICD10CM | O10.311 | Pre-existing hypertensive heart and chronic kidney disease complicating pregnancy, first trimester |
| ICD10CM | O10.312 | Pre-existing hypertensive heart and chronic kidney disease complicating pregnancy, second trimester |
| ICD10CM | O10.313 | Pre-existing hypertensive heart and chronic kidney disease complicating pregnancy, third trimester |
| ICD10CM | O10.319 | Pre-existing hypertensive heart and chronic kidney disease complicating pregnancy, unspecified trimester |
| ICD10CM | O10.32 | Pre-existing hypertensive heart and chronic kidney disease complicating childbirth |
| ICD10CM | O10.33 | Pre-existing hypertensive heart and chronic kidney disease complicating the puerperium |
| ICD10CM | Q61.2 | Polycystic kidney, adult type |
| ICD10CM | Q61.3 | Polycystic kidney, unspecified |
| ICD10CM | Q61.8 | Other cystic kidney diseases |
| ICD10CM | Z94.0 | Kidney transplant status |
| ICD10CM | N18.1 | Chronic kidney disease, stage 1 |
| ICD10CM | N18.2 | Chronic kidney disease, stage 2 (mild) |
| ICD10CM | N18.3 | Chronic kidney disease, stage 3 (moderate) |
| ICD10CM | N18.4 | Chronic kidney disease, stage 4 (severe) |
| ICD10CM | N18.5 | Chronic kidney disease, stage 5 |
| ICD10CM | N18.6 | End stage renal disease |
| ICD10CM | N18.9 | Chronic kidney disease, unspecified |
| ICD10CM | N18.30 | Chronic kidney disease, stage 3 unspecified |
| ICD10CM | N18.31 | Chronic kidney disease, stage 3a |
| ICD10CM | N18.32 | Chronic kidney disease, stage 3b |
| SNOMED CT | 709044004 | Chronic kidney disease |
| SNOMED CT | 16726004 | Renal osteodystrophy |
| SNOMED CT | 46177005 | End-stage renal disease |
| SNOMED CT | 49708008 | Anemia of chronic renal failure |
| SNOMED CT | 57557005 | Chronic milk alkali syndrome |
| SNOMED CT | 90688005 | Chronic renal failure |
| SNOMED CT | 236433006 | Acute-on-chronic renal failure |
| SNOMED CT | 236434000 | End stage renal failure untreated by renal replacement therapy |
| SNOMED CT | 236435004 | End stage renal failure on dialysis |
| SNOMED CT | 236436003 | End stage renal failure with renal transplant |
| SNOMED CT | 236552002 | Adynamic bone disease |
| SNOMED CT | 425369003 | Chronic progressive renal failure |

Continued on next page

**Table 4 – continued from previous page**

| CodeSystem | ConceptCode | ConceptName |
|---|---|---|
| SNOMED CT | 431855005 | Chronic kidney disease stage 1 |
| SNOMED CT | 431856006 | Chronic kidney disease stage 2 |
| SNOMED CT | 431857002 | Chronic kidney disease stage 4 |
| SNOMED CT | 433144002 | Chronic kidney disease stage 3 |
| SNOMED CT | 433146000 | Chronic kidney disease stage 5 |
| SNOMED CT | 700378005 | Chronic kidney disease stage 3A |
| SNOMED CT | 700379002 | Chronic kidney disease stage 3B |
| SNOMED CT | 704667004 | Hypertension concurrent and due to end stage renal disease on dialysis |
| SNOMED CT | 707324008 | Anemia in end stage renal disease |
| SNOMED CT | 708975004 | Mixed renal osteodystrophy |
| SNOMED CT | 712487000 | End stage renal disease due to benign hypertension |
| SNOMED CT | 713313000 | Chronic kidney disease mineral and bone disorder |
| SNOMED CT | 714152005 | Chronic kidney disease stage 5 on dialysis |
| SNOMED CT | 714153000 | Chronic kidney disease stage 5 with transplant |
| SNOMED CT | 722098007 | Chronic kidney disease following donor nephrectomy |
| SNOMED CT | 722149000 | Chronic kidney disease due to and following excision of neoplasm of kidney |
| SNOMED CT | 722150000 | Chronic kidney disease due to systemic infection |
| SNOMED CT | 722467000 | Chronic kidney disease due to traumatic loss of kidney |
| SNOMED CT | 723190009 | Chronic renal insufficiency |
| SNOMED CT | 776416004 | Hyperuricemia, pulmonary hypertension, renal failure, alkalosis syndrome |
| SNOMED CT | 711000119100 | Chronic kidney disease stage 5 due to type 2 diabetes mellitus |
| SNOMED CT | 721000119107 | Chronic kidney disease stage 4 due to type 2 diabetes mellitus |
| SNOMED CT | 731000119105 | Chronic kidney disease stage 3 due to type 2 diabetes mellitus |
| SNOMED CT | 741000119101 | Chronic kidney disease stage 2 due to type 2 diabetes mellitus |
| SNOMED CT | 751000119104 | Chronic kidney disease stage 1 due to type 2 diabetes mellitus |
| SNOMED CT | 771000119108 | Chronic kidney disease due to type 2 diabetes mellitus |
| SNOMED CT | 1801000119106 | Anemia, pre-end stage renal disease on erythropoietin protocol |
| SNOMED CT | 8501000119104 | Hypertensive heart and chronic kidney disease |
| SNOMED CT | 71421000119105 | Hypertension in chronic kidney disease due to type 2 diabetes mellitus |
| SNOMED CT | 71701000119105 | Hypertension in chronic kidney disease due to type 1 diabetes mellitus |
| SNOMED CT | 90721000119101 | Chronic kidney disease stage 1 due to type 1 diabetes mellitus |

Continued on next page

**Table 4 – continued from previous page**

| CodeSystem | ConceptCode | ConceptName |
|---|---|---|
| SNOMED CT | 90731000119103 | Chronic kidney disease stage 2 due to type 1 diabetes mellitus |
| SNOMED CT | 90741000119107 | Chronic kidney disease stage 3 due to type 1 diabetes mellitus |
| SNOMED CT | 90751000119109 | Chronic kidney disease stage 4 due to type 1 diabetes mellitus |
| SNOMED CT | 90761000119106 | Chronic kidney disease stage 5 due to type 1 diabetes mellitus |
| SNOMED CT | 90771000119100 | End stage renal disease on dialysis due to type 1 diabetes mellitus |
| SNOMED CT | 90791000119104 | End stage renal disease on dialysis due to type 2 diabetes mellitus |
| SNOMED CT | 96441000119101 | Chronic kidney disease due to type 1 diabetes mellitus |
| SNOMED CT | 96701000119107 | Hypertensive heart AND chronic kidney disease on dialysis |
| SNOMED CT | 96711000119105 | Hypertensive heart AND chronic kidney disease stage 5 |
| SNOMED CT | 96721000119103 | Hypertensive heart AND chronic kidney disease stage 4 |
| SNOMED CT | 96731000119100 | Hypertensive heart AND chronic kidney disease stage 3 |
| SNOMED CT | 96741000119109 | Hypertensive heart AND chronic kidney disease stage 2 |
| SNOMED CT | 96751000119106 | Hypertensive heart AND chronic kidney disease stage 1 |
| SNOMED CT | 104931000119100 | Chronic kidney disease due to hypertension |
| SNOMED CT | 111411000119103 | End stage renal disease due to hypertension |
| SNOMED CT | 117681000119102 | Chronic kidney disease stage 1 due to hypertension |
| SNOMED CT | 118781000119108 | Pre-existing hypertensive chronic kidney disease in mother complicating pregnancy |
| SNOMED CT | 120261000119101 | Benign hypertensive heart disease and chronic renal disease stage 5 |
| SNOMED CT | 127991000119101 | Hypertension concurrent and due to end stage renal disease on dialysis due to type 2 diabetes mellitus |
| SNOMED CT | 128001000119105 | Hypertension concurrent and due to end stage renal disease on dialysis due to type 1 diabetes mellitus |
| SNOMED CT | 129151000119102 | Chronic kidney disease stage 4 due to hypertension |
| SNOMED CT | 129161000119100 | Chronic kidney disease stage 5 due to hypertension |
| SNOMED CT | 129171000119106 | Chronic kidney disease stage 3 due to hypertension |
| SNOMED CT | 129181000119109 | Chronic kidney disease stage 2 due to hypertension |
| SNOMED CT | 140101000119109 | Hypertension in chronic kidney disease stage 5 due to type 2 diabetes mellitus |
| SNOMED CT | 140111000119107 | Hypertension in chronic kidney disease stage 4 due to type 2 diabetes mellitus |
| SNOMED CT | 140121000119100 | Hypertension in chronic kidney disease stage 3 due to type 2 diabetes mellitus |
| SNOMED CT | 140131000119102 | Hypertension in chronic kidney disease stage 2 due to type 2 diabetes mellitus |
| SNOMED CT | 153851000119106 | Malignant hypertensive chronic kidney disease stage 5 |

Continued on next page

**Table 4 – continued from previous page**

| CodeSystem | ConceptCode | ConceptName |
|---|---|---|
| SNOMED CT | 153891000119101 | End stage renal disease on dialysis due to hypertension |
| SNOMED CT | 284961000119106 | Chronic kidney disease due to benign hypertension |
| SNOMED CT | 284971000119100 | Chronic kidney disease stage 1 due to benign hypertension |
| SNOMED CT | 284981000119102 | Chronic kidney disease stage 2 due to benign hypertension |
| SNOMED CT | 284991000119104 | Chronic kidney disease stage 3 due to benign hypertension |
| SNOMED CT | 285001000119105 | Chronic kidney disease stage 4 due to benign hypertension |
| SNOMED CT | 285011000119108 | Chronic kidney disease stage 5 due to benign hypertension |
| SNOMED CT | 285041000119107 | Benign hypertensive heart disease and chronic renal disease stage 1 |
| SNOMED CT | 285061000119106 | Benign hypertensive heart disease and chronic renal disease stage 2 |
| SNOMED CT | 285081000119102 | Benign hypertensive heart disease and chronic renal disease stage 3 |
| SNOMED CT | 285101000119109 | Benign hypertensive heart disease and chronic renal disease stage 4 |
| SNOMED CT | 285831000119108 | Malignant hypertensive chronic kidney disease |
| SNOMED CT | 285841000119104 | Malignant hypertensive end stage renal disease |
| SNOMED CT | 285851000119102 | Malignant hypertensive chronic kidney disease stage 1 |
| SNOMED CT | 285861000119100 | Malignant hypertensive chronic kidney disease stage 2 |
| SNOMED CT | 285871000119106 | Malignant hypertensive chronic kidney disease stage 3 |
| SNOMED CT | 285881000119109 | Malignant hypertensive chronic kidney disease stage 4 |
| SNOMED CT | 285911000119109 | Malignant hypertensive heart disease and chronic renal disease stage 3 |
| SNOMED CT | 285921000119102 | Malignant hypertensive heart disease and chronic renal disease stage 4 |
| SNOMED CT | 286371000119107 | Malignant hypertensive end stage renal disease on dialysis |
| SNOMED CT | 434431000124103 | Hypertensive end stage renal disease |
| SNOMED CT | 449631000124102 | Benign hypertensive heart disease and chronic renal disease |
| SNOMED CT | 691421000119108 | Anemia co-occurrent and due to chronic kidney disease stage 3 |
| SNOMED CT | 10757401000119104 | Pre-existing hypertensive heart and chronic kidney disease in mother complicating childbirth |
| SNOMED CT | 10757481000119107 | Pre-existing hypertensive heart and chronic kidney disease in mother complicating pregnancy |
| SNOMED CT | 368421000119108 | Chronic kidney disease stage 1 due to drug induced diabetes mellitus |
| SNOMED CT | 368431000119106 | Chronic kidney disease stage 2 due to drug induced diabetes mellitus |
| SNOMED CT | 368441000119102 | Chronic kidney disease stage 3 due to drug induced diabetes mellitus |
| SNOMED CT | 368451000119100 | Chronic kidney disease stage 4 due to drug induced diabetes mellitus |

Continued on next page

**Table 4 – continued from previous page**

| CodeSystem | ConceptCode | ConceptName |
|---|---|---|
| SNOMED CT | 368461000119103 | Chronic kidney disease stage 5 due to drug induced diabetes mellitus |
| SNOMED CT | 368471000119109 | End stage renal disease on dialysis due to drug induced diabetes mellitus |
| SNOMED CT | 897308007 | Chronic kidney disease with osteoporosis |
| SNOMED CT | 897310009 | Renal osteodystrophy with high bone turnover |
| SNOMED CT | 897311008 | Renal osteodystrophy with normal bone turnover |
| SNOMED CT | 897312001 | Renal osteodystrophy with low bone turnover |
| SNOMED CT | 1217070004 | Renal osteodystrophy due to hyperparathyroidism |
| SNOMED CT | 1208934006 | Sagliker syndrome |

#### 3.2 Chronic lung disease

Table 5: Concept codes used to define chronic lung disease.

| CodeSystem | ConceptCode | ConceptName |
|---|---|---|
| ICD10CM | I27.8 | Other specified pulmonary heart diseases |
| ICD10CM | I27.81 | Cor pulmonale (chronic) |
| ICD10CM | I27.82 | Chronic pulmonary embolism |
| ICD10CM | I27.83 | Eisenmenger's syndrome |
| ICD10CM | I27.89 | Other specified pulmonary heart diseases |
| ICD10CM | I27.9 | Pulmonary heart disease, unspecified |
| ICD10CM | J41 | Simple and mucopurulent chronic bronchitis |
| ICD10CM | J42 | Unspecified chronic bronchitis |
| ICD10CM | J43 | Emphysema |
| ICD10CM | J44 | Other chronic obstructive pulmonary disease |
| ICD10CM | J45 | Asthma |
| ICD10CM | J47 | Bronchiectasis |
| ICD10CM | J47.0 | Bronchiectasis with acute lower respiratory infection |
| ICD10CM | J47.1 | Bronchiectasis with (acute) exacerbation |
| ICD10CM | J47.9 | Bronchiectasis, uncomplicated |
| ICD10CM | J60 | Coalworker's pneumoconiosis |
| ICD10CM | J67 | Hypersensitivity pneumonitis due to organic dust |
| ICD10CM | J67.0 | Farmer's lung |
| ICD10CM | J67.1 | Bagassosis |
| ICD10CM | J67.2 | Bird fancier's lung |
| ICD10CM | J67.3 | Suberosis |
| ICD10CM | J67.4 | Maltworker's lung |
| ICD10CM | J67.5 | Mushroom-worker's lung |
| ICD10CM | J67.6 | Maple-bark-stripper's lung |
| ICD10CM | J67.7 | Air conditioner and humidifier lung |
| ICD10CM | J67.8 | Hypersensitivity pneumonitis due to other organic dusts |
| ICD10CM | J67.9 | Hypersensitivity pneumonitis due to unspecified organic dust |
| ICD10CM | J68.4 | Chronic respiratory conditions due to chemicals, gases, fumes and vapors |
| ICD10CM | J70.1 | Chronic and other pulmonary manifestations due to radiation |
| ICD10CM | J70.3 | Chronic drug-induced interstitial lung disorders |
| ICD10CM | J41.0 | Simple chronic bronchitis |
| ICD10CM | J41.1 | Mucopurulent chronic bronchitis |
| ICD10CM | J41.8 | Mixed simple and mucopurulent chronic bronchitis |
| ICD10CM | J43.0 | Unilateral pulmonary emphysema [MacLeod's syndrome] |
| ICD10CM | J43.1 | Panlobular emphysema |
| ICD10CM | J43.2 | Centrilobular emphysema |
| ICD10CM | J43.8 | Other emphysema |

Continued on next page

**Table 5 – continued from previous page**

| CodeSystem | ConceptCode | ConceptName |
|---|---|---|
| ICD10CM | J43.9 | Emphysema, unspecified |
| ICD10CM | J44.0 | Chronic obstructive pulmonary disease with (acute) lower respiratory infection |
| ICD10CM | J44.1 | Chronic obstructive pulmonary disease with (acute) exacerbation |
| ICD10CM | J44.9 | Chronic obstructive pulmonary disease, unspecified |
| ICD10CM | J45.2 | Mild intermittent asthma |
| ICD10CM | J45.20 | Mild intermittent asthma, uncomplicated |
| ICD10CM | J45.21 | Mild intermittent asthma with (acute) exacerbation |
| ICD10CM | J45.22 | Mild intermittent asthma with status asthmaticus |
| ICD10CM | J45.3 | Mild persistent asthma |
| ICD10CM | J45.30 | Mild persistent asthma, uncomplicated |
| ICD10CM | J45.31 | Mild persistent asthma with (acute) exacerbation |
| ICD10CM | J45.32 | Mild persistent asthma with status asthmaticus |
| ICD10CM | J45.4 | Moderate persistent asthma |
| ICD10CM | J45.40 | Moderate persistent asthma, uncomplicated |
| ICD10CM | J45.41 | Moderate persistent asthma with (acute) exacerbation |
| ICD10CM | J45.42 | Moderate persistent asthma with status asthmaticus |
| ICD10CM | J45.5 | Severe persistent asthma |
| ICD10CM | J45.50 | Severe persistent asthma, uncomplicated |
| ICD10CM | J45.51 | Severe persistent asthma with (acute) exacerbation |
| ICD10CM | J45.52 | Severe persistent asthma with status asthmaticus |
| ICD10CM | J45.9 | Other and unspecified asthma |
| ICD10CM | J45.90 | Unspecified asthma |
| ICD10CM | J45.901 | Unspecified asthma with (acute) exacerbation |
| ICD10CM | J45.902 | Unspecified asthma with status asthmaticus |
| ICD10CM | J45.909 | Unspecified asthma, uncomplicated |
| ICD10CM | J45.99 | Other asthma |
| ICD10CM | J45.990 | Exercise induced bronchospasm |
| ICD10CM | J45.991 | Cough variant asthma |
| ICD10CM | J45.998 | Other asthma |
| SNOMED CT | 13645005 | Chronic obstructive lung disease |
| SNOMED CT | 63480004 | Chronic bronchitis |
| SNOMED CT | 70995007 | Pulmonary hypertension |
| SNOMED CT | 195967001 | Asthma |
| SNOMED CT | 413839001 | Chronic lung disease |
| SNOMED CT | 2912004 | Cystic-bullous disease of the lung |
| SNOMED CT | 4981000 | Panacinar emphysema |
| SNOMED CT | 5499009 | Pulmonary hypertensive venous disease |
| SNOMED CT | 10964002 | Progressive pulmonary hypertension |
| SNOMED CT | 11399002 | Pulmonary arterial hypertension |
| SNOMED CT | 11641008 | Millers' asthma |
| SNOMED CT | 12428000 | Intrinsic asthma without status asthmaticus |

Continued on next page

**Table 5 – continued from previous page**

| CodeSystem | ConceptCode | ConceptName |
|---|---|---|
| SNOMED CT | 16003001 | Giant bullous emphysema |
| SNOMED CT | 16846004 | Obstructive emphysema |
| SNOMED CT | 18041002 | Printers' asthma |
| SNOMED CT | 18121009 | Chronic berylliosis |
| SNOMED CT | 18354001 | Chronic induration of lung |
| SNOMED CT | 19849005 | Meat-wrappers' asthma |
| SNOMED CT | 23958009 | Vanishing lung |
| SNOMED CT | 26427008 | Chronic pulmonary histoplasmosis |
| SNOMED CT | 31387002 | Exercise-induced asthma |
| SNOMED CT | 31898008 | Paraseptal emphysema |
| SNOMED CT | 33325001 | Compensatory emphysema |
| SNOMED CT | 34015007 | Bakers' asthma |
| SNOMED CT | 36599006 | Chronic fibrosis of lung |
| SNOMED CT | 37180002 | Chronic nonspecific lung disease |
| SNOMED CT | 37981002 | Allergic bronchopulmonary aspergillosis |
| SNOMED CT | 41553006 | Detergent asthma |
| SNOMED CT | 46847001 | Chronic pulmonary edema |
| SNOMED CT | 47895001 | Congenital emphysema |
| SNOMED CT | 47938003 | Chronic obliterative bronchiolitis |
| SNOMED CT | 49691004 | Occupational bronchitis |
| SNOMED CT | 51577008 | Wilson-Mikity syndrome |
| SNOMED CT | 52571006 | Chronic tracheobronchitis |
| SNOMED CT | 55570000 | Asthma without status asthmaticus |
| SNOMED CT | 56968009 | Asthma caused by wood dust |
| SNOMED CT | 57607007 | Occupational asthma |
| SNOMED CT | 57686001 | Emphysematous bleb of lung |
| SNOMED CT | 59786004 | Weavers' cough |
| SNOMED CT | 60805002 | Hemolytic anemia with emphysema AND cutis laxa |
| SNOMED CT | 61937009 | Simple chronic bronchitis |
| SNOMED CT | 63088003 | Allergic asthma without status asthmaticus |
| SNOMED CT | 63841001 | Chronic pulmonary congestion |
| SNOMED CT | 66987001 | Congenital lobar emphysema |
| SNOMED CT | 67569000 | Bronchopulmonary dysplasia of newborn |
| SNOMED CT | 68328006 | Centriacinar emphysema |
| SNOMED CT | 70756004 | Bronchial atresia with segmental pulmonary emphysema |
| SNOMED CT | 74417001 | Mucopurulent chronic bronchitis |
| SNOMED CT | 77690003 | Interstitial emphysema of lung |
| SNOMED CT | 84409004 | Fetid chronic bronchitis |
| SNOMED CT | 86680006 | Ruptured emphysematous bleb of lung |
| SNOMED CT | 87394009 | Episodic pulmonary hypertension |
| SNOMED CT | 87433001 | Pulmonary emphysema |
| SNOMED CT | 88223008 | Secondary pulmonary hypertension |
| SNOMED CT | 89099002 | Chronic allergic bronchitis |

Continued on next page

**Table 5 – continued from previous page**

| CodeSystem | ConceptCode | ConceptName |
|---|---|---|
| SNOMED CT | 89420002 | Pulmonary veno-occlusive disease |
| SNOMED CT | 90610005 | Interstitial pulmonary fibrosis of prematurity |
| SNOMED CT | 92807009 | Chemical-induced asthma |
| SNOMED CT | 93432008 | Drug-induced asthma |
| SNOMED CT | 123713005 | Chronic atelectasis |
| SNOMED CT | 135836000 | End stage chronic obstructive airways disease |
| SNOMED CT | 185086009 | Emphysematous bronchitis |
| SNOMED CT | 195949008 | Chronic asthmatic bronchitis |
| SNOMED CT | 195951007 | Acute exacerbation of chronic obstructive airways disease |
| SNOMED CT | 195953005 | Mixed simple and mucopurulent chronic bronchitis |
| SNOMED CT | 195957006 | Chronic bullous emphysema |
| SNOMED CT | 195958001 | Segmental bullous emphysema |
| SNOMED CT | 195959009 | Zonal bullous emphysema |
| SNOMED CT | 195977004 | Mixed asthma |
| SNOMED CT | 196001008 | Chronic obstructive pulmonary disease with acute lower respiratory infection |
| SNOMED CT | 196026004 | Chronic emphysema due to chemical fumes |
| SNOMED CT | 196028003 | Chronic pulmonary fibrosis due to chemical fumes |
| SNOMED CT | 196049002 | Chronic pulmonary radiation disease |
| SNOMED CT | 225057002 | Brittle asthma |
| SNOMED CT | 233615002 | Chronic pulmonary coccidioidomycosis |
| SNOMED CT | 233675009 | Toxic emphysema |
| SNOMED CT | 233677001 | Scar emphysema |
| SNOMED CT | 233678006 | Childhood asthma |
| SNOMED CT | 233679003 | Late onset asthma |
| SNOMED CT | 233683003 | Hay fever with asthma |
| SNOMED CT | 233687002 | Colophony asthma |
| SNOMED CT | 233688007 | Sulfite-induced asthma |
| SNOMED CT | 233691007 | Eosinophilic asthma |
| SNOMED CT | 233713004 | Seasonal cryptogenic organizing pneumonia with biochemical cholestasis |
| SNOMED CT | 233762004 | Chronic silicosis |
| SNOMED CT | 233815004 | Persistent pulmonary hypertension of the newborn |
| SNOMED CT | 233941006 | Solitary pulmonary hypertension |
| SNOMED CT | 233942004 | Small vessel pulmonary hypertension |
| SNOMED CT | 233943009 | Sporadic primary pulmonary hypertension |
| SNOMED CT | 233944003 | Familial primary pulmonary hypertension |
| SNOMED CT | 233945002 | Pulmonary arterial hypertension induced by drug |
| SNOMED CT | 233946001 | Large vessel pulmonary hypertension |
| SNOMED CT | 233947005 | Chronic thromboembolic pulmonary hypertension |
| SNOMED CT | 233949008 | Pulmonary capillary hemangiomatosis |
| SNOMED CT | 233950008 | Pulmonary hypertension associated with chronic underventilation |

Continued on next page

**Table 5 – continued from previous page**

| CodeSystem | ConceptCode | ConceptName |
|---|---|---|
| SNOMED CT | 233954004 | Pulmonary hypertension due to chronic exposure to high altitude |
| SNOMED CT | 234161007 | Familial pulmonary capillary hemangiomatosis |
| SNOMED CT | 240742009 | Chronic pulmonary African histoplasmosis |
| SNOMED CT | 240747003 | Chronic pulmonary blastomycosis |
| SNOMED CT | 266355005 | Bullous emphysema with collapse |
| SNOMED CT | 266356006 | Atrophic (senile) emphysema |
| SNOMED CT | 266361008 | Intrinsic asthma |
| SNOMED CT | 266404004 | Chronic chemical respiratory disease |
| SNOMED CT | 276537001 | Chronic pulmonary insufficiency of prematurity |
| SNOMED CT | 276792008 | Pulmonary hypertension with extreme obesity |
| SNOMED CT | 276793003 | Pulmonary hypertension with occult mitral stenosis |
| SNOMED CT | 276794009 | Facultative pulmonary hypertension with shunt at atrial level |
| SNOMED CT | 281239006 | Acute asthma |
| SNOMED CT | 285381006 | Acute infective exacerbation of chronic obstructive airways disease |
| SNOMED CT | 304527002 | Acute asthma |
| SNOMED CT | 313296004 | Mild chronic obstructive pulmonary disease |
| SNOMED CT | 313297008 | Moderate chronic obstructive pulmonary disease |
| SNOMED CT | 313299006 | Severe chronic obstructive pulmonary disease |
| SNOMED CT | 314954002 | Local recurrence of malignant tumor of lung |
| SNOMED CT | 360573001 | Postcapillary pulmonary hypertension |
| SNOMED CT | 360578005 | Pulmonary hypertension secondary to raised pulmonary vascular resistance |
| SNOMED CT | 361194002 | Perinatal pulmonary fibroplasia |
| SNOMED CT | 361195001 | Pulmonary fibroplasia |
| SNOMED CT | 370218001 | Mild asthma |
| SNOMED CT | 370219009 | Moderate asthma |
| SNOMED CT | 370220003 | Occasional asthma |
| SNOMED CT | 370221004 | Severe asthma |
| SNOMED CT | 389145006 | Allergic asthma |
| SNOMED CT | 404804003 | Platinum asthma |
| SNOMED CT | 404806001 | Cheese-makers' asthma |
| SNOMED CT | 404808000 | Isocyanate induced asthma |
| SNOMED CT | 405944004 | Asthmatic bronchitis |
| SNOMED CT | 407674008 | Aspirin-induced asthma |
| SNOMED CT | 409663006 | Cough variant asthma |
| SNOMED CT | 417688002 | Chronic progressive coccidioidal pneumonia |
| SNOMED CT | 418395004 | Tea-makers' asthma |
| SNOMED CT | 424199006 | Substance induced asthma |
| SNOMED CT | 424643009 | IgE-mediated allergic asthma |
| SNOMED CT | 425748003 | Acute exacerbation of chronic bronchitis |

Continued on next page

**Table 5 – continued from previous page**

| CodeSystem | ConceptCode | ConceptName |
|---|---|---|
| SNOMED CT | 425969006 | Exacerbation of intermittent asthma |
| SNOMED CT | 426437004 | Familial idiopathic pulmonary fibrosis |
| SNOMED CT | 426656000 | Severe persistent asthma |
| SNOMED CT | 426979002 | Mild persistent asthma |
| SNOMED CT | 427295004 | Moderate persistent asthma |
| SNOMED CT | 427603009 | Intermittent asthma |
| SNOMED CT | 427679007 | Mild intermittent asthma |
| SNOMED CT | 430476004 | Diffuse panbronchiolitis |
| SNOMED CT | 430969000 | Recurrent aspiration pneumonia |
| SNOMED CT | 432958009 | Chronic rejection of lung transplant |
| SNOMED CT | 442025000 | Acute exacerbation of chronic asthmatic bronchitis |
| SNOMED CT | 445237003 | Pulmonary arterial hypertension associated with portal hypertension |
| SNOMED CT | 445427006 | Seasonal asthma |
| SNOMED CT | 445928005 | Eisenmenger's syndrome |
| SNOMED CT | 447006007 | Relapse pulmonary tuberculosis |
| SNOMED CT | 472757001 | Pulmonary venous hypertension as complication of procedure |
| SNOMED CT | 472758006 | Pulmonary venous hypertension due to compression of pulmonary great vein |
| SNOMED CT | 472759003 | Pulmonary venous hypertension due to compression of pulmonary great vein by sclerosing mediastinitis |
| SNOMED CT | 472760008 | Pulmonary venous hypertension due to compression of pulmonary great vein by lymphadenopathy |
| SNOMED CT | 472761007 | Pulmonary venous hypertension due to compression of pulmonary great vein by neoplasm |
| SNOMED CT | 472790001 | Pulmonary hypertension due to left heart disease |
| SNOMED CT | 697896007 | Precapillary pulmonary hypertension |
| SNOMED CT | 697897003 | Heritable pulmonary arterial hypertension |
| SNOMED CT | 697898008 | Idiopathic pulmonary arterial hypertension |
| SNOMED CT | 697901009 | Pulmonary arterial hypertension induced by toxin |
| SNOMED CT | 697902002 | Associated pulmonary arterial hypertension |
| SNOMED CT | 697903007 | Pulmonary arterial hypertension associated with connective tissue disease |
| SNOMED CT | 697904001 | Pulmonary arterial hypertension associated with HIV infection |
| SNOMED CT | 697905000 | Pulmonary arterial hypertension associated with congenital heart disease |
| SNOMED CT | 697906004 | Pulmonary arterial hypertension associated with congenital systemic-to-pulmonary shunt |
| SNOMED CT | 697907008 | Pulmonary arterial hypertension associated with schistosomiasis |

Continued on next page

**Table 5 – continued from previous page**

| CodeSystem | ConceptCode | ConceptName |
|---|---|---|
| SNOMED CT | 697908003 | Pulmonary arterial hypertension associated with chronic hemolytic anemia |
| SNOMED CT | 697909006 | Pulmonary veno-occlusive disease and/or pulmonary capillary hemangiomatosis |
| SNOMED CT | 697910001 | Pulmonary hypertension due to lung disease and/or hypoxia |
| SNOMED CT | 697911002 | Pulmonary hypertension due to chronic obstructive pulmonary disease |
| SNOMED CT | 697912009 | Pulmonary hypertension due to interstitial lung disease |
| SNOMED CT | 697913004 | Pulmonary hypertension due to pulmonary disease with mixed restrictive and obstructive pattern |
| SNOMED CT | 697914005 | Pulmonary hypertension due to sleep-disordered breathing |
| SNOMED CT | 697915006 | Pulmonary hypertension due to alveolar hypoventilation disorder |
| SNOMED CT | 697916007 | Pulmonary hypertension due to developmental abnormality of the lung |
| SNOMED CT | 697917003 | Pulmonary hypertension due to hematological disorder |
| SNOMED CT | 697918008 | Pulmonary hypertension due to myeloproliferative disorder |
| SNOMED CT | 697919000 | Pulmonary hypertension due to post-splenectomy hematological disorder |
| SNOMED CT | 697920006 | Pulmonary hypertension in systemic disorder |
| SNOMED CT | 697921005 | Pulmonary hypertension in sarcoidosis |
| SNOMED CT | 697922003 | Pulmonary hypertension in Langerhans cell histiocytosis |
| SNOMED CT | 697923008 | Pulmonary hypertension in lymphangioleiomyomatosis |
| SNOMED CT | 697924002 | Pulmonary hypertension in neurofibromatosis |
| SNOMED CT | 697925001 | Pulmonary hypertension due to systolic systemic ventricular dysfunction |
| SNOMED CT | 697926000 | Pulmonary hypertension due to diastolic systemic ventricular dysfunction |
| SNOMED CT | 697927009 | Pulmonary hypertension due to left-sided valvular heart disease |
| SNOMED CT | 697928004 | Pulmonary venous hypertension due to congenital stenosis of pulmonary vein |
| SNOMED CT | 699014000 | Recurrent pneumonia |
| SNOMED CT | 700250006 | Idiopathic pulmonary fibrosis |
| SNOMED CT | 703355003 | Pulmonary hypertension due to vasculitis |
| SNOMED CT | 704345008 | Chronic interstitial pneumonia |
| SNOMED CT | 707444001 | Uncomplicated asthma |
| SNOMED CT | 707445000 | Exacerbation of mild persistent asthma |
| SNOMED CT | 707446004 | Exacerbation of moderate persistent asthma |
| SNOMED CT | 707447008 | Exacerbation of severe persistent asthma |
| SNOMED CT | 707511009 | Uncomplicated mild persistent asthma |
| SNOMED CT | 707512002 | Uncomplicated moderate persistent asthma |
| SNOMED CT | 707513007 | Uncomplicated severe persistent asthma |

Continued on next page

**Table 5 – continued from previous page**

| CodeSystem | ConceptCode | ConceptName |
|---|---|---|
| SNOMED CT | 707534000 | Chronic lung disease due to surfactant disorder |
| SNOMED CT | 707553005 | Chronic exogenous lipoid pneumonia |
| SNOMED CT | 707979007 | Acute severe exacerbation of severe persistent asthma |
| SNOMED CT | 707980005 | Acute severe exacerbation of moderate persistent asthma |
| SNOMED CT | 707981009 | Acute severe exacerbation of mild persistent asthma |
| SNOMED CT | 708030004 | Pulmonary emphysema co-occurrent with fibrosis of lung |
| SNOMED CT | 708038006 | Acute exacerbation of asthma |
| SNOMED CT | 708090002 | Acute severe exacerbation of asthma |
| SNOMED CT | 708093000 | Acute exacerbation of allergic asthma |
| SNOMED CT | 708094006 | Acute exacerbation of intrinsic asthma |
| SNOMED CT | 708095007 | Acute severe exacerbation of immunoglobin E-mediated allergic asthma |
| SNOMED CT | 708096008 | Acute severe exacerbation of intrinsic asthma |
| SNOMED CT | 713525001 | Recurrent bacterial pneumonia |
| SNOMED CT | 713526000 | Recurrent bacterial pneumonia co-occurrent with human immunodeficiency virus infection |
| SNOMED CT | 720575002 | Braddock syndrome |
| SNOMED CT | 723829000 | Trimorphic syndrome |
| SNOMED CT | 724500003 | Idiopathic chronic eosinophilic pneumonia |
| SNOMED CT | 733171006 | Chronic pulmonary aspergillosis |
| SNOMED CT | 733858005 | Acute severe refractory exacerbation of asthma |
| SNOMED CT | 734904007 | Life threatening acute exacerbation of asthma |
| SNOMED CT | 734905008 | Moderate acute exacerbation of asthma |
| SNOMED CT | 735465007 | Protracted bronchitis caused by bacterium |
| SNOMED CT | 735588005 | Uncomplicated allergic asthma |
| SNOMED CT | 735589002 | Uncomplicated non-allergic asthma |
| SNOMED CT | 737180005 | Chronic bronchiolitis |
| SNOMED CT | 762521001 | Acute allergic asthma |
| SNOMED CT | 762618008 | Bronchiolitis obliterans syndrome due to and after lung transplantation |
| SNOMED CT | 771306007 | Hereditary fibrosing poikiloderma, tendon contractures, myopathy, pulmonary fibrosis syndrome |
| SNOMED CT | 776416004 | Hyperuricemia, pulmonary hypertension, renal failure, alkalosis syndrome |
| SNOMED CT | 782513000 | Acute severe exacerbation of allergic asthma |
| SNOMED CT | 782520007 | Exacerbation of allergic asthma due to infection |
| SNOMED CT | 783182004 | Chronic respiratory distress with surfactant metabolism deficiency |
| SNOMED CT | 785736001 | Chronic bronchitis co-occurrent with wheeze |
| SNOMED CT | 786836003 | Near fatal asthma |
| SNOMED CT | 789574002 | Acute exacerbation of idiopathic pulmonary fibrosis |
| SNOMED CT | 829976001 | Thunderstorm asthma |
| SNOMED CT | 836477007 | Chronic emphysema due to vapor |

Continued on next page

**Table 5 – continued from previous page**

| CodeSystem | ConceptCode | ConceptName |
|---|---|---|
| SNOMED CT | 840306007 | Mild pulmonary hypertension |
| SNOMED CT | 840307003 | Moderate pulmonary hypertension |
| SNOMED CT | 840308008 | Severe pulmonary hypertension |
| SNOMED CT | 840350008 | Chronic obliterative bronchiolitis due to chemical fumes |
| SNOMED CT | 840351007 | Chronic obliterative bronchiolitis due to vapor |
| SNOMED CT | 846637007 | Chronic pulmonary fibrosis caused by chemical vapors |
| SNOMED CT | 401000119107 | Asthma with irreversible airway obstruction |
| SNOMED CT | 901000119100 | Persistent asthma, well controlled |
| SNOMED CT | 1741000119102 | Intermittent asthma uncontrolled |
| SNOMED CT | 1751000119100 | Acute exacerbation of chronic obstructive airways disease with asthma |
| SNOMED CT | 5281000124103 | Persistent asthma |
| SNOMED CT | 28791000119105 | Chronic coccidioidomycotic pneumonia |
| SNOMED CT | 72301000119103 | Asthma in pregnancy |
| SNOMED CT | 99031000119107 | Acute exacerbation of asthma co-occurrent with allergic rhinitis |
| SNOMED CT | 102361000119104 | Chronic pneumonia |
| SNOMED CT | 103781000119103 | Allergic bronchopulmonary mycosis |
| SNOMED CT | 106001000119101 | Chronic obstructive lung disease co-occurrent with acute bronchitis |
| SNOMED CT | 124991000119109 | Severe persistent asthma co-occurrent with allergic rhinitis |
| SNOMED CT | 125001000119103 | Moderate persistent asthma co-occurrent with allergic rhinitis |
| SNOMED CT | 125011000119100 | Mild persistent asthma co-occurrent with allergic rhinitis |
| SNOMED CT | 125021000119107 | Intermittent asthma co-occurrent with allergic rhinitis |
| SNOMED CT | 135171000119106 | Acute exacerbation of moderate persistent asthma |
| SNOMED CT | 135181000119109 | Acute exacerbation of mild persistent asthma |
| SNOMED CT | 293241000119100 | Acute exacerbation of chronic obstructive bronchitis |
| SNOMED CT | 434301000124102 | Chronic interstitial lung disease |
| SNOMED CT | 451981000124108 | Eosinophilic bronchitis |
| SNOMED CT | 460561000124109 | PF-ILD-progressive fibrosing interstitial lung disease |
| SNOMED CT | 2360001000004109 | Steroid dependent asthma |
| SNOMED CT | 10674711000119105 | Acute severe exacerbation of asthma co-occurrent with allergic rhinitis |
| SNOMED CT | 10674991000119104 | Intermittent allergic asthma |
| SNOMED CT | 10675391000119101 | Severe controlled persistent asthma |
| SNOMED CT | 10675431000119106 | Severe persistent allergic asthma |
| SNOMED CT | 10675471000119109 | Acute severe exacerbation of severe persistent allergic asthma |
| SNOMED CT | 10675551000119104 | Acute severe exacerbation of severe persistent asthma co-occurrent with allergic rhinitis |
| SNOMED CT | 10675751000119107 | Severe uncontrolled persistent asthma |
| SNOMED CT | 10675871000119106 | Mild persistent allergic asthma |

Continued on next page

**Table 5 – continued from previous page**

| CodeSystem | ConceptCode | ConceptName |
|---|---|---|
| SNOMED CT | 10675911000119109 | Acute severe exacerbation of mild persistent allergic asthma |
| SNOMED CT | 10675991000119100 | Acute severe exacerbation of mild persistent allergic asthma co-occurrent with allergic rhinitis |
| SNOMED CT | 10676391000119108 | Moderate persistent allergic asthma |
| SNOMED CT | 10676431000119103 | Acute severe exacerbation of moderate persistent allergic asthma |
| SNOMED CT | 10676511000119109 | Acute severe exacerbation of moderate persistent asthma co-occurrent with allergic rhinitis |
| SNOMED CT | 10692681000119108 | Aspirin exacerbated respiratory disease |
| SNOMED CT | 10692721000119102 | Chronic obstructive asthma co-occurrent with acute exacerbation of asthma |
| SNOMED CT | 10692761000119107 | Asthma-chronic obstructive pulmonary disease overlap syndrome |
| SNOMED CT | 10742121000119104 | Asthma in mother complicating childbirth |
| SNOMED CT | 16055311000119107 | Inhaled steroid-dependent asthma |
| SNOMED CT | 16584951000119101 | Oral steroid-dependent asthma |
| SNOMED CT | 233674008 | Pulmonary emphysema in alpha-1 primary immunodeficiency deficiency |
| SNOMED CT | 233948000 | Pulmonary hypertension in vasculitis |
| SNOMED CT | 423889005 | Non-immunoglobulin E mediated allergic asthma |
| SNOMED CT | 697899000 | Heritable pulmonary arterial hypertension due to bone morphogenetic protein receptor type II mutation |
| SNOMED CT | 697900005 | Heritable pulmonary arterial hypertension due to activin A receptor type II-like kinase 1 or endoglin mutation |
| SNOMED CT | 703953004 | Asthma caused by house dust mite, Dermatophagoides pteronyssinus |
| SNOMED CT | 703954005 | Asthma caused by house dust mite, Dermatophagoides farinae |
| SNOMED CT | 1010333003 | Emphysema of left lung |
| SNOMED CT | 1010334009 | Emphysema of right lung |
| SNOMED CT | 1010625007 | Pulmonary hypertension due to pulmonary disease with mixed restrictive and obstructive patterns |
| SNOMED CT | 1010627004 | Pulmonary hypertension due to developmental abnormality |
| SNOMED CT | 1010670004 | Chronic endogenous lipoid pneumonia |
| SNOMED CT | 879814007 | Pulmonary hypertension due to pulmonary capillary hemangiomatosis |
| SNOMED CT | 879815008 | Pulmonary hypertension due to pulmonary veno-occlusive disease |
| SNOMED CT | 879816009 | Pulmonary hypertension due to familial pulmonary capillary hemangiomatosis |
| SNOMED CT | 1163150009 | Chronic cavitary pulmonary aspergillosis |
| SNOMED CT | 1177120001 | Bronchiolitis obliterans syndrome due to and following allogeneic stem cell transplant |

Continued on next page

**Table 5 – continued from previous page**

| CodeSystem | ConceptCode | ConceptName |
|---|---|---|
| SNOMED CT | 1228876007 | Severe early-onset pulmonary alveolar proteinosis due to MARS deficiency |
| SNOMED CT | 1163147006 | Chronic fibrosing pulmonary aspergillosis |
| SNOMED CT | 1197476009 | Infantile-onset pulmonary alveolar proteinosis, hypogammaglobulinemia |

#### 38 3.3 Diabetes

Table 6: Concept codes used to define diabetes.

| CodeSystem | ConceptCode | ConceptName |
|---|---|---|
| ICD10CM | O24.83 | Other pre-existing diabetes mellitus in the puerperium |
| ICD9CM | 250.02 | Diabetes mellitus without mention of complication, type II or unspecified type, uncontrolled |
| ICD10CM | O24.33 | Unspecified pre-existing diabetes mellitus in the puerperium |
| ICD9CM | 250.01 | Diabetes mellitus without mention of complication, type I [juvenile type], not stated as uncontrolled |
| ICD10CM | O24.82 | Other pre-existing diabetes mellitus in childbirth |
| ICD9CM | 250.00 | Diabetes mellitus without mention of complication, type II or unspecified type, not stated as uncontrolled |
| ICD10CM | O24.32 | Unspecified pre-existing diabetes mellitus in childbirth |
| SNOMED CT | 237604008 | Maturity onset diabetes of the young, type 2 |
| ICD10CM | O24.819 | Other pre-existing diabetes mellitus in pregnancy, unspecified trimester |
| SNOMED CT | 237599002 | Insulin treated type 2 diabetes mellitus |
| ICD10CM | O24.319 | Unspecified pre-existing diabetes mellitus in pregnancy, unspecified trimester |
| SNOMED CT | 23045005 | Insulin dependent diabetes mellitus type 1A |
| ICD10CM | O24.813 | Other pre-existing diabetes mellitus in pregnancy, third trimester |
| SNOMED CT | 199230006 | Pre-existing type 2 diabetes mellitus |
| ICD10CM | O24.313 | Unspecified pre-existing diabetes mellitus in pregnancy, third trimester |
| SNOMED CT | 199229001 | Pre-existing type 1 diabetes mellitus |
| ICD10CM | O24.812 | Other pre-existing diabetes mellitus in pregnancy, second trimester |
| SNOMED CT | 190389009 | Type 2 diabetes mellitus with ulcer |
| ICD10CM | O24.312 | Unspecified pre-existing diabetes mellitus in pregnancy, second trimester |
| SNOMED CT | 190372001 | Type 1 diabetes mellitus maturity onset |
| ICD10CM | O24.811 | Other pre-existing diabetes mellitus in pregnancy, first trimester |
| SNOMED CT | 190368000 | Type 1 diabetes mellitus with ulcer |
| ICD10CM | O24.311 | Unspecified pre-existing diabetes mellitus in pregnancy, first trimester |
| SNOMED CT | 190331003 | Hyperosmolar coma due to type 2 diabetes mellitus |
| ICD10CM | E10.3539 | Type 1 diabetes mellitus with proliferative diabetic retinopathy with traction retinal detachment not involving the macula, unspecified eye |
| SNOMED CT | 190330002 | Hyperosmolar coma due to type 1 diabetes mellitus |
| ICD10CM | O24.13 | Pre-existing type 2 diabetes mellitus, in the puerperium |

Continued on next page

**Table 6 – continued from previous page**

| CodeSystem | ConceptCode | ConceptName |
|---|---|---|
| SNOMED CT | 1551000119108 | Nonproliferative retinopathy due to type 2 diabetes mellitus |
| ICD10CM | E10.3411 | Type 1 diabetes mellitus with severe nonproliferative diabetic retinopathy with macular edema, right eye |
| SNOMED CT | 1511000119107 | Peripheral neuropathy due to type 2 diabetes mellitus |
| ICD10CM | O24.12 | Pre-existing type 2 diabetes mellitus, in childbirth |
| SNOMED CT | 1501000119109 | Proliferative retinopathy due to type 2 diabetes mellitus |
| ICD10CM | E10.341 | Type 1 diabetes mellitus with severe nonproliferative diabetic retinopathy with macular edema |
| SNOMED CT | 1481000119100 | Diabetes mellitus type 2 without retinopathy |
| ICD10CM | O24.119 | Pre-existing type 2 diabetes mellitus, in pregnancy, unspecified trimester |
| SNOMED CT | 138941000119105 | Severe nonproliferative retinopathy due to diabetes mellitus type 2 |
| ICD10CM | E10.3533 | Type 1 diabetes mellitus with proliferative diabetic retinopathy with traction retinal detachment not involving the macula, bilateral |
| SNOMED CT | 138921000119104 | Moderate nonproliferative retinopathy due to type 2 diabetes mellitus |
| ICD10CM | O24.113 | Pre-existing type 2 diabetes mellitus, in pregnancy, third trimester |
| SNOMED CT | 138911000119106 | Mild nonproliferative retinopathy due to type 2 diabetes mellitus |
| ICD10CM | E10.3532 | Type 1 diabetes mellitus with proliferative diabetic retinopathy with traction retinal detachment not involving the macula, left eye |
| SNOMED CT | 138901000119108 | Severe nonproliferative retinopathy due to diabetes mellitus type 1 |
| ICD10CM | O24.112 | Pre-existing type 2 diabetes mellitus, in pregnancy, second trimester |
| SNOMED CT | 138891000119109 | Moderate nonproliferative retinopathy due to type 1 diabetes mellitus |
| ICD10CM | E10.3531 | Type 1 diabetes mellitus with proliferative diabetic retinopathy with traction retinal detachment not involving the macula, right eye |
| SNOMED CT | 138881000119106 | Mild nonproliferative retinopathy due to type 1 diabetes mellitus |
| ICD10CM | O24.111 | Pre-existing type 2 diabetes mellitus, in pregnancy, first trimester |
| SNOMED CT | 110181000119105 | Peripheral sensory neuropathy due to type 2 diabetes mellitus |
| ICD10CM | E10.3529 | Type 1 diabetes mellitus with proliferative diabetic retinopathy with traction retinal detachment involving the macula, unspecified eye |

Continued on next page

**Table 6 – continued from previous page**

| CodeSystem | ConceptCode | ConceptName |
|---|---|---|
| SNOMED CT | 109171000119104 | Retinal edema due to type 1 diabetes mellitus |
| ICD10CM | O24.03 | Pre-existing type 1 diabetes mellitus, in the puerperium |
| SNOMED CT | 104961000119108 | Ischemia of retina due to type 2 diabetes mellitus |
| ICD10CM | E10.3523 | Type 1 diabetes mellitus with proliferative diabetic retinopathy with traction retinal detachment involving the macula, bilateral |
| SNOMED CT | 104941000119109 | Ischemia of retina due to type 1 diabetes mellitus |
| ICD10CM | O24.02 | Pre-existing type 1 diabetes mellitus, in childbirth |
| SNOMED CT | 102781000119107 | Sensory neuropathy due to type 1 diabetes mellitus |
| ICD10CM | E10.3522 | Type 1 diabetes mellitus with proliferative diabetic retinopathy with traction retinal detachment involving the macula, left eye |
| ICD10CM | O24.019 | Pre-existing type 1 diabetes mellitus, in pregnancy, unspecified trimester |
| ICD10CM | E10.3521 | Type 1 diabetes mellitus with proliferative diabetic retinopathy with traction retinal detachment involving the macula, right eye |
| ICD10CM | O24.013 | Pre-existing type 1 diabetes mellitus, in pregnancy, third trimester |
| ICD10CM | E10.3519 | Type 1 diabetes mellitus with proliferative diabetic retinopathy with macular edema, unspecified eye |
| ICD10CM | O24.012 | Pre-existing type 1 diabetes mellitus, in pregnancy, second trimester |
| ICD10CM | E10.3513 | Type 1 diabetes mellitus with proliferative diabetic retinopathy with macular edema, bilateral |
| ICD10CM | O24.011 | Pre-existing type 1 diabetes mellitus, in pregnancy, first trimester |
| ICD10CM | E10.3512 | Type 1 diabetes mellitus with proliferative diabetic retinopathy with macular edema, left eye |
| ICD10CM | E13.9 | Other specified diabetes mellitus without complications |
| ICD10CM | E10.3511 | Type 1 diabetes mellitus with proliferative diabetic retinopathy with macular edema, right eye |
| ICD10CM | E13.8 | Other specified diabetes mellitus with unspecified complications |
| ICD10CM | E10.351 | Type 1 diabetes mellitus with proliferative diabetic retinopathy with macular edema |
| ICD10CM | E13.69 | Other specified diabetes mellitus with other specified complication |
| ICD10CM | E10.3499 | Type 1 diabetes mellitus with severe nonproliferative diabetic retinopathy without macular edema, unspecified eye |
| ICD10CM | E13.65 | Other specified diabetes mellitus with hyperglycemia |
| ICD10CM | E10.3493 | Type 1 diabetes mellitus with severe nonproliferative diabetic retinopathy without macular edema, bilateral |

Continued on next page

**Table 6 – continued from previous page**

| CodeSystem | ConceptCode | ConceptName |
|---|---|---|
| ICD10CM | E13.649 | Other specified diabetes mellitus with hypoglycemia without coma |
| ICD10CM | E10.3492 | Type 1 diabetes mellitus with severe nonproliferative diabetic retinopathy without macular edema, left eye |
| ICD10CM | E13.641 | Other specified diabetes mellitus with hypoglycemia with coma |
| ICD10CM | E10.3491 | Type 1 diabetes mellitus with severe nonproliferative diabetic retinopathy without macular edema, right eye |
| ICD10CM | E13.638 | Other specified diabetes mellitus with other oral complications |
| ICD10CM | E10.349 | Type 1 diabetes mellitus with severe nonproliferative diabetic retinopathy without macular edema |
| ICD10CM | E13.630 | Other specified diabetes mellitus with periodontal disease |
| ICD10CM | E10.3419 | Type 1 diabetes mellitus with severe nonproliferative diabetic retinopathy with macular edema, unspecified eye |
| ICD10CM | E13.628 | Other specified diabetes mellitus with other skin complications |
| ICD10CM | E10.3413 | Type 1 diabetes mellitus with severe nonproliferative diabetic retinopathy with macular edema, bilateral |
| ICD10CM | E13.622 | Other specified diabetes mellitus with other skin ulcer |
| ICD10CM | E10.3412 | Type 1 diabetes mellitus with severe nonproliferative diabetic retinopathy with macular edema, left eye |
| ICD10CM | E11.37X2 | Type 2 diabetes mellitus with diabetic macular edema, resolved following treatment, left eye |
| ICD10CM | E10.3399 | Type 1 diabetes mellitus with moderate nonproliferative diabetic retinopathy without macular edema, unspecified eye |
| ICD10CM | E11.37X1 | Type 2 diabetes mellitus with diabetic macular edema, resolved following treatment, right eye |
| ICD10CM | E10.3393 | Type 1 diabetes mellitus with moderate nonproliferative diabetic retinopathy without macular edema, bilateral |
| ICD10CM | E11.36 | Type 2 diabetes mellitus with diabetic cataract |
| ICD10CM | E10.3392 | Type 1 diabetes mellitus with moderate nonproliferative diabetic retinopathy without macular edema, left eye |
| ICD10CM | E11.3599 | Type 2 diabetes mellitus with proliferative diabetic retinopathy without macular edema, unspecified eye |
| ICD10CM | E10.3391 | Type 1 diabetes mellitus with moderate nonproliferative diabetic retinopathy without macular edema, right eye |
| ICD10CM | E11.3593 | Type 2 diabetes mellitus with proliferative diabetic retinopathy without macular edema, bilateral |
| ICD10CM | E10.339 | Type 1 diabetes mellitus with moderate nonproliferative diabetic retinopathy without macular edema |
| ICD10CM | E11.3592 | Type 2 diabetes mellitus with proliferative diabetic retinopathy without macular edema, left eye |

Continued on next page

**Table 6 – continued from previous page**

| CodeSystem | ConceptCode | ConceptName |
|---|---|---|
| ICD10CM | E10.3319 | Type 1 diabetes mellitus with moderate nonproliferative diabetic retinopathy with macular edema, unspecified eye |
| ICD10CM | E11.3591 | Type 2 diabetes mellitus with proliferative diabetic retinopathy without macular edema, right eye |
| ICD10CM | E10.3313 | Type 1 diabetes mellitus with moderate nonproliferative diabetic retinopathy with macular edema, bilateral |
| ICD10CM | E11.359 | Type 2 diabetes mellitus with proliferative diabetic retinopathy without macular edema |
| ICD10CM | E10.3312 | Type 1 diabetes mellitus with moderate nonproliferative diabetic retinopathy with macular edema, left eye |
| ICD10CM | E11.3559 | Type 2 diabetes mellitus with stable proliferative diabetic retinopathy, unspecified eye |
| ICD10CM | E10.3311 | Type 1 diabetes mellitus with moderate nonproliferative diabetic retinopathy with macular edema, right eye |
| ICD10CM | E11.3553 | Type 2 diabetes mellitus with stable proliferative diabetic retinopathy, bilateral |
| ICD10CM | E10.331 | Type 1 diabetes mellitus with moderate nonproliferative diabetic retinopathy with macular edema |
| ICD10CM | E11.3552 | Type 2 diabetes mellitus with stable proliferative diabetic retinopathy, left eye |
| ICD10CM | E10.3299 | Type 1 diabetes mellitus with mild nonproliferative diabetic retinopathy without macular edema, unspecified eye |
| ICD10CM | E11.3551 | Type 2 diabetes mellitus with stable proliferative diabetic retinopathy, right eye |
| ICD10CM | E10.3293 | Type 1 diabetes mellitus with mild nonproliferative diabetic retinopathy without macular edema, bilateral |
| ICD10CM | E11.3549 | Type 2 diabetes mellitus with proliferative diabetic retinopathy with combined traction retinal detachment and rhegmatogenous retinal detachment, unspecified eye |
| ICD10CM | E10.3292 | Type 1 diabetes mellitus with mild nonproliferative diabetic retinopathy without macular edema, left eye |
| ICD10CM | E11.3543 | Type 2 diabetes mellitus with proliferative diabetic retinopathy with combined traction retinal detachment and rhegmatogenous retinal detachment, bilateral |
| ICD10CM | E10.3291 | Type 1 diabetes mellitus with mild nonproliferative diabetic retinopathy without macular edema, right eye |
| ICD10CM | E11.3542 | Type 2 diabetes mellitus with proliferative diabetic retinopathy with combined traction retinal detachment and rhegmatogenous retinal detachment, left eye |
| ICD10CM | E10.329 | Type 1 diabetes mellitus with mild nonproliferative diabetic retinopathy without macular edema |

Continued on next page

**Table 6 – continued from previous page**

| CodeSystem | ConceptCode | ConceptName |
|---|---|---|
| ICD10CM | E11.3541 | Type 2 diabetes mellitus with proliferative diabetic retinopathy with combined traction retinal detachment and rhegmatogenous retinal detachment, right eye |
| ICD10CM | E10.3219 | Type 1 diabetes mellitus with mild nonproliferative diabetic retinopathy with macular edema, unspecified eye |
| ICD10CM | E11.3539 | Type 2 diabetes mellitus with proliferative diabetic retinopathy with traction retinal detachment not involving the macula, unspecified eye |
| ICD10CM | E10.3213 | Type 1 diabetes mellitus with mild nonproliferative diabetic retinopathy with macular edema, bilateral |
| ICD10CM | E11.3533 | Type 2 diabetes mellitus with proliferative diabetic retinopathy with traction retinal detachment not involving the macula, bilateral |
| ICD10CM | E10.3212 | Type 1 diabetes mellitus with mild nonproliferative diabetic retinopathy with macular edema, left eye |
| ICD10CM | E11.3532 | Type 2 diabetes mellitus with proliferative diabetic retinopathy with traction retinal detachment not involving the macula, left eye |
| ICD10CM | E10.3211 | Type 1 diabetes mellitus with mild nonproliferative diabetic retinopathy with macular edema, right eye |
| ICD10CM | E11.3531 | Type 2 diabetes mellitus with proliferative diabetic retinopathy with traction retinal detachment not involving the macula, right eye |
| ICD10CM | E10.321 | Type 1 diabetes mellitus with mild nonproliferative diabetic retinopathy with macular edema |
| ICD10CM | E11.3529 | Type 2 diabetes mellitus with proliferative diabetic retinopathy with traction retinal detachment involving the macula, unspecified eye |
| ICD10CM | E10.319 | Type 1 diabetes mellitus with unspecified diabetic retinopathy without macular edema |
| ICD10CM | E11.3523 | Type 2 diabetes mellitus with proliferative diabetic retinopathy with traction retinal detachment involving the macula, bilateral |
| ICD10CM | E10.311 | Type 1 diabetes mellitus with unspecified diabetic retinopathy with macular edema |
| ICD10CM | E11.3522 | Type 2 diabetes mellitus with proliferative diabetic retinopathy with traction retinal detachment involving the macula, left eye |
| ICD10CM | E10.29 | Type 1 diabetes mellitus with other diabetic kidney complication |
| ICD10CM | E11.3521 | Type 2 diabetes mellitus with proliferative diabetic retinopathy with traction retinal detachment involving the macula, right eye |

Continued on next page

**Table 6 – continued from previous page**

| CodeSystem | ConceptCode | ConceptName |
|---|---|---|
| ICD10CM | E10.22 | Type 1 diabetes mellitus with diabetic chronic kidney disease |
| ICD10CM | E11.3519 | Type 2 diabetes mellitus with proliferative diabetic retinopathy with macular edema, unspecified eye |
| ICD10CM | E10.21 | Type 1 diabetes mellitus with diabetic nephropathy |
| ICD10CM | E11.3513 | Type 2 diabetes mellitus with proliferative diabetic retinopathy with macular edema, bilateral |
| ICD10CM | E10.11 | Type 1 diabetes mellitus with ketoacidosis with coma |
| ICD10CM | E11.3512 | Type 2 diabetes mellitus with proliferative diabetic retinopathy with macular edema, left eye |
| ICD10CM | E10.10 | Type 1 diabetes mellitus with ketoacidosis without coma |
| ICD10CM | E11.3511 | Type 2 diabetes mellitus with proliferative diabetic retinopathy with macular edema, right eye |
| SNOMED CT | 9859006 | Acanthosis nigricans due to type 2 diabetes mellitus |
| ICD10CM | E11.351 | Type 2 diabetes mellitus with proliferative diabetic retinopathy with macular edema |
| SNOMED CT | 97331000119101 | Macular edema and retinopathy due to type 2 diabetes mellitus |
| ICD10CM | E11.3499 | Type 2 diabetes mellitus with severe nonproliferative diabetic retinopathy without macular edema, unspecified eye |
| SNOMED CT | 87921000119104 | Cranial nerve palsy with type II diabetes mellitus |
| ICD10CM | E11.3493 | Type 2 diabetes mellitus with severe nonproliferative diabetic retinopathy without macular edema, bilateral |
| SNOMED CT | 82581000119105 | Rubeosis iridis due to type 1 diabetes mellitus |
| ICD10CM | E11.3492 | Type 2 diabetes mellitus with severe nonproliferative diabetic retinopathy without macular edema, left eye |
| SNOMED CT | 82571000119107 | Traction detachment of retina due to type 1 diabetes mellitus |
| ICD10CM | E11.3491 | Type 2 diabetes mellitus with severe nonproliferative diabetic retinopathy without macular edema, right eye |
| ICD10CM | E11.3213 | Type 2 diabetes mellitus with mild nonproliferative diabetic retinopathy with macular edema, bilateral |
| ICD10CM | E11.349 | Type 2 diabetes mellitus with severe nonproliferative diabetic retinopathy without macular edema |
| ICD10CM | E11.3212 | Type 2 diabetes mellitus with mild nonproliferative diabetic retinopathy with macular edema, left eye |
| ICD10CM | E11.3419 | Type 2 diabetes mellitus with severe nonproliferative diabetic retinopathy with macular edema, unspecified eye |
| ICD10CM | E11.3211 | Type 2 diabetes mellitus with mild nonproliferative diabetic retinopathy with macular edema, right eye |
| ICD10CM | E11.3413 | Type 2 diabetes mellitus with severe nonproliferative diabetic retinopathy with macular edema, bilateral |

Continued on next page

**Table 6 – continued from previous page**

| CodeSystem | ConceptCode | ConceptName |
|---|---|---|
| ICD10CM | E11.321 | Type 2 diabetes mellitus with mild nonproliferative diabetic retinopathy with macular edema |
| ICD10CM | E11.3412 | Type 2 diabetes mellitus with severe nonproliferative diabetic retinopathy with macular edema, left eye |
| ICD10CM | E11.319 | Type 2 diabetes mellitus with unspecified diabetic retinopathy without macular edema |
| ICD10CM | E11.3411 | Type 2 diabetes mellitus with severe nonproliferative diabetic retinopathy with macular edema, right eye |
| ICD10CM | E11.311 | Type 2 diabetes mellitus with unspecified diabetic retinopathy with macular edema |
| ICD10CM | E11.341 | Type 2 diabetes mellitus with severe nonproliferative diabetic retinopathy with macular edema |
| ICD10CM | E11.29 | Type 2 diabetes mellitus with other diabetic kidney complication |
| ICD10CM | E11.3399 | Type 2 diabetes mellitus with moderate nonproliferative diabetic retinopathy without macular edema, unspecified eye |
| ICD10CM | E11.22 | Type 2 diabetes mellitus with diabetic chronic kidney disease |
| ICD10CM | E11.3393 | Type 2 diabetes mellitus with moderate nonproliferative diabetic retinopathy without macular edema, bilateral |
| ICD10CM | E11.21 | Type 2 diabetes mellitus with diabetic nephropathy |
| ICD10CM | E11.3392 | Type 2 diabetes mellitus with moderate nonproliferative diabetic retinopathy without macular edema, left eye |
| ICD10CM | E11.11 | Type 2 diabetes mellitus with ketoacidosis with coma |
| ICD10CM | E11.3391 | Type 2 diabetes mellitus with moderate nonproliferative diabetic retinopathy without macular edema, right eye |
| ICD10CM | E11.10 | Type 2 diabetes mellitus with ketoacidosis without coma |
| ICD10CM | E11.339 | Type 2 diabetes mellitus with moderate nonproliferative diabetic retinopathy without macular edema |
| ICD10CM | E11.01 | Type 2 diabetes mellitus with hyperosmolarity with coma |
| ICD10CM | E11.3319 | Type 2 diabetes mellitus with moderate nonproliferative diabetic retinopathy with macular edema, unspecified eye |
| ICD10CM | E11.00 | Type 2 diabetes mellitus with hyperosmolarity without nonketotic hyperglycemic-hyperosmolar coma (NKHHC) |
| ICD10CM | E11.3313 | Type 2 diabetes mellitus with moderate nonproliferative diabetic retinopathy with macular edema, bilateral |
| ICD10CM | E10.9 | Type 1 diabetes mellitus without complications |
| ICD10CM | E11.3312 | Type 2 diabetes mellitus with moderate nonproliferative diabetic retinopathy with macular edema, left eye |
| ICD10CM | E10.8 | Type 1 diabetes mellitus with unspecified complications |
| ICD10CM | E11.3311 | Type 2 diabetes mellitus with moderate nonproliferative diabetic retinopathy with macular edema, right eye |
| ICD10CM | E10.69 | Type 1 diabetes mellitus with other specified complication |

Continued on next page

**Table 6 – continued from previous page**

| CodeSystem | ConceptCode | ConceptName |
|---|---|---|
| ICD10CM | E11.331 | Type 2 diabetes mellitus with moderate nonproliferative diabetic retinopathy with macular edema |
| ICD10CM | E10.65 | Type 1 diabetes mellitus with hyperglycemia |
| ICD10CM | E11.3299 | Type 2 diabetes mellitus with mild nonproliferative diabetic retinopathy without macular edema, unspecified eye |
| ICD10CM | E10.649 | Type 1 diabetes mellitus with hypoglycemia without coma |
| ICD10CM | E11.3293 | Type 2 diabetes mellitus with mild nonproliferative diabetic retinopathy without macular edema, bilateral |
| ICD10CM | E10.641 | Type 1 diabetes mellitus with hypoglycemia with coma |
| ICD10CM | E11.3292 | Type 2 diabetes mellitus with mild nonproliferative diabetic retinopathy without macular edema, left eye |
| ICD10CM | E10.638 | Type 1 diabetes mellitus with other oral complications |
| ICD10CM | E11.3291 | Type 2 diabetes mellitus with mild nonproliferative diabetic retinopathy without macular edema, right eye |
| ICD10CM | E10.630 | Type 1 diabetes mellitus with periodontal disease |
| ICD10CM | E11.329 | Type 2 diabetes mellitus with mild nonproliferative diabetic retinopathy without macular edema |
| ICD10CM | E10.628 | Type 1 diabetes mellitus with other skin complications |
| ICD10CM | E11.3219 | Type 2 diabetes mellitus with mild nonproliferative diabetic retinopathy with macular edema, unspecified eye |
| ICD10CM | E10.622 | Type 1 diabetes mellitus with other skin ulcer |
| ICD10CM | E13.3399 | Other specified diabetes mellitus with moderate nonproliferative diabetic retinopathy without macular edema, unspecified eye |
| ICD10CM | E10.621 | Type 1 diabetes mellitus with foot ulcer |
| ICD10CM | E13.3393 | Other specified diabetes mellitus with moderate nonproliferative diabetic retinopathy without macular edema, bilateral |
| ICD10CM | E10.620 | Type 1 diabetes mellitus with diabetic dermatitis |
| ICD10CM | E13.3392 | Other specified diabetes mellitus with moderate nonproliferative diabetic retinopathy without macular edema, left eye |
| ICD10CM | E10.618 | Type 1 diabetes mellitus with other diabetic arthropathy |
| ICD10CM | E13.3391 | Other specified diabetes mellitus with moderate nonproliferative diabetic retinopathy without macular edema, right eye |
| ICD10CM | E10.610 | Type 1 diabetes mellitus with diabetic neuropathic arthropathy |
| ICD10CM | E13.339 | Other specified diabetes mellitus with moderate nonproliferative diabetic retinopathy without macular edema |
| ICD10CM | E10.59 | Type 1 diabetes mellitus with other circulatory complications |

Continued on next page

**Table 6 – continued from previous page**

| CodeSystem | ConceptCode | ConceptName |
|---|---|---|
| ICD10CM | E13.3319 | Other specified diabetes mellitus with moderate nonproliferative diabetic retinopathy with macular edema, unspecified eye |
| ICD10CM | E10.52 | Type 1 diabetes mellitus with diabetic peripheral angiopathy with gangrene |
| ICD10CM | E13.3313 | Other specified diabetes mellitus with moderate nonproliferative diabetic retinopathy with macular edema, bilateral |
| ICD10CM | E10.51 | Type 1 diabetes mellitus with diabetic peripheral angiopathy without gangrene |
| ICD10CM | E13.3312 | Other specified diabetes mellitus with moderate nonproliferative diabetic retinopathy with macular edema, left eye |
| ICD10CM | E10.49 | Type 1 diabetes mellitus with other diabetic neurological complication |
| ICD10CM | E13.3311 | Other specified diabetes mellitus with moderate nonproliferative diabetic retinopathy with macular edema, right eye |
| ICD10CM | E10.44 | Type 1 diabetes mellitus with diabetic amyotrophy |
| ICD10CM | E13.331 | Other specified diabetes mellitus with moderate nonproliferative diabetic retinopathy with macular edema |
| ICD10CM | E10.43 | Type 1 diabetes mellitus with diabetic autonomic (poly)neuropathy |
| ICD10CM | E13.3299 | Other specified diabetes mellitus with mild nonproliferative diabetic retinopathy without macular edema, unspecified eye |
| ICD10CM | E10.42 | Type 1 diabetes mellitus with diabetic polyneuropathy |
| ICD10CM | E13.3293 | Other specified diabetes mellitus with mild nonproliferative diabetic retinopathy without macular edema, bilateral |
| ICD10CM | E10.41 | Type 1 diabetes mellitus with diabetic mononeuropathy |
| ICD10CM | E13.3292 | Other specified diabetes mellitus with mild nonproliferative diabetic retinopathy without macular edema, left eye |
| ICD10CM | E10.40 | Type 1 diabetes mellitus with diabetic neuropathy, unspecified |
| ICD10CM | E13.3291 | Other specified diabetes mellitus with mild nonproliferative diabetic retinopathy without macular edema, right eye |
| ICD10CM | E10.39 | Type 1 diabetes mellitus with other diabetic ophthalmic complication |
| ICD10CM | E13.329 | Other specified diabetes mellitus with mild nonproliferative diabetic retinopathy without macular edema |
| ICD10CM | E10.37X9 | Type 1 diabetes mellitus with diabetic macular edema, resolved following treatment, unspecified eye |
| ICD10CM | E13.3219 | Other specified diabetes mellitus with mild nonproliferative diabetic retinopathy with macular edema, unspecified eye |
| ICD10CM | E10.37X3 | Type 1 diabetes mellitus with diabetic macular edema, resolved following treatment, bilateral |

Continued on next page

**Table 6 – continued from previous page**

| CodeSystem | ConceptCode | ConceptName |
|---|---|---|
| ICD10CM | E13.3213 | Other specified diabetes mellitus with mild nonproliferative diabetic retinopathy with macular edema, bilateral |
| ICD10CM | E10.37X2 | Type 1 diabetes mellitus with diabetic macular edema, resolved following treatment, left eye |
| ICD10CM | E13.3212 | Other specified diabetes mellitus with mild nonproliferative diabetic retinopathy with macular edema, left eye |
| ICD10CM | E10.37X1 | Type 1 diabetes mellitus with diabetic macular edema, resolved following treatment, right eye |
| ICD10CM | E13.3211 | Other specified diabetes mellitus with mild nonproliferative diabetic retinopathy with macular edema, right eye |
| ICD10CM | E10.36 | Type 1 diabetes mellitus with diabetic cataract |
| ICD10CM | E13.321 | Other specified diabetes mellitus with mild nonproliferative diabetic retinopathy with macular edema |
| ICD10CM | E10.3599 | Type 1 diabetes mellitus with proliferative diabetic retinopathy without macular edema, unspecified eye |
| ICD10CM | E13.319 | Other specified diabetes mellitus with unspecified diabetic retinopathy without macular edema |
| ICD10CM | E10.3593 | Type 1 diabetes mellitus with proliferative diabetic retinopathy without macular edema, bilateral |
| ICD10CM | E13.311 | Other specified diabetes mellitus with unspecified diabetic retinopathy with macular edema |
| ICD10CM | E10.3592 | Type 1 diabetes mellitus with proliferative diabetic retinopathy without macular edema, left eye |
| ICD10CM | E13.29 | Other specified diabetes mellitus with other diabetic kidney complication |
| ICD10CM | E10.3591 | Type 1 diabetes mellitus with proliferative diabetic retinopathy without macular edema, right eye |
| ICD10CM | E13.22 | Other specified diabetes mellitus with diabetic chronic kidney disease |
| ICD10CM | E10.359 | Type 1 diabetes mellitus with proliferative diabetic retinopathy without macular edema |
| ICD10CM | E13.21 | Other specified diabetes mellitus with diabetic nephropathy |
| ICD10CM | E10.3559 | Type 1 diabetes mellitus with stable proliferative diabetic retinopathy, unspecified eye |
| ICD10CM | E13.11 | Other specified diabetes mellitus with ketoacidosis with coma |
| ICD10CM | E10.3553 | Type 1 diabetes mellitus with stable proliferative diabetic retinopathy, bilateral |
| ICD10CM | E13.10 | Other specified diabetes mellitus with ketoacidosis without coma |
| ICD10CM | E10.3552 | Type 1 diabetes mellitus with stable proliferative diabetic retinopathy, left eye |

Continued on next page

**Table 6 – continued from previous page**

| CodeSystem | ConceptCode | ConceptName |
|---|---|---|
| ICD10CM | E13.01 | Other specified diabetes mellitus with hyperosmolarity with coma |
| ICD10CM | E10.3551 | Type 1 diabetes mellitus with stable proliferative diabetic retinopathy, right eye |
| ICD10CM | E13.00 | Other specified diabetes mellitus with hyperosmolarity without nonketotic hyperglycemic-hyperosmolar coma (NKHHC) |
| ICD10CM | E10.3549 | Type 1 diabetes mellitus with proliferative diabetic retinopathy with combined traction retinal detachment and rhegmatogenous retinal detachment, unspecified eye |
| ICD10CM | E11.9 | Type 2 diabetes mellitus without complications |
| ICD10CM | E10.3543 | Type 1 diabetes mellitus with proliferative diabetic retinopathy with combined traction retinal detachment and rhegmatogenous retinal detachment, bilateral |
| ICD10CM | E11.8 | Type 2 diabetes mellitus with unspecified complications |
| ICD10CM | E10.3542 | Type 1 diabetes mellitus with proliferative diabetic retinopathy with combined traction retinal detachment and rhegmatogenous retinal detachment, left eye |
| ICD10CM | E11.69 | Type 2 diabetes mellitus with other specified complication |
| ICD10CM | E10.3541 | Type 1 diabetes mellitus with proliferative diabetic retinopathy with combined traction retinal detachment and rhegmatogenous retinal detachment, right eye |
| ICD10CM | E11.65 | Type 2 diabetes mellitus with hyperglycemia |
| ICD9CM | 362.07 | Diabetic macular edema |
| ICD10CM | E11.649 | Type 2 diabetes mellitus with hypoglycemia without coma |
| ICD9CM | 362.06 | Severe nonproliferative diabetic retinopathy |
| ICD10CM | E11.641 | Type 2 diabetes mellitus with hypoglycemia with coma |
| ICD9CM | 362.05 | Moderate nonproliferative diabetic retinopathy |
| ICD10CM | E11.638 | Type 2 diabetes mellitus with other oral complications |
| ICD9CM | 362.04 | Mild nonproliferative diabetic retinopathy |
| ICD10CM | E11.630 | Type 2 diabetes mellitus with periodontal disease |
| ICD9CM | 362.03 | Nonproliferative diabetic retinopathy NOS |
| ICD10CM | E11.628 | Type 2 diabetes mellitus with other skin complications |
| ICD9CM | 362.02 | Proliferative diabetic retinopathy |
| ICD10CM | E11.622 | Type 2 diabetes mellitus with other skin ulcer |
| ICD9CM | 362.01 | Background diabetic retinopathy |
| ICD10CM | E11.621 | Type 2 diabetes mellitus with foot ulcer |
| SNOMED CT | 359642000 | Type 2 diabetes mellitus in nonobese |
| ICD10CM | E11.620 | Type 2 diabetes mellitus with diabetic dermatitis |
| ICD9CM | 357.2 | Polyneuropathy in diabetes |
| ICD10CM | E11.618 | Type 2 diabetes mellitus with other diabetic arthropathy |
| SNOMED CT | 314903002 | Arthropathy due to type 2 diabetes mellitus |

Continued on next page

**Table 6 – continued from previous page**

| CodeSystem | ConceptCode | ConceptName |
|---|---|---|
| ICD10CM | E11.610 | Type 2 diabetes mellitus with diabetic neuropathic arthropathy |
| SNOMED CT | 314902007 | Type 2 diabetes mellitus with peripheral angiopathy |
| ICD10CM | E11.59 | Type 2 diabetes mellitus with other circulatory complications |
| SNOMED CT | 314893005 | Type 1 diabetes mellitus with arthropathy |
| ICD10CM | E11.52 | Type 2 diabetes mellitus with diabetic peripheral angiopathy with gangrene |
| SNOMED CT | 313436004 | Type 2 diabetes mellitus without complication |
| ICD10CM | E11.51 | Type 2 diabetes mellitus with diabetic peripheral angiopathy without gangrene |
| SNOMED CT | 313435000 | Type 1 diabetes mellitus without complication |
| ICD10CM | E11.49 | Type 2 diabetes mellitus with other diabetic neurological complication |
| SNOMED CT | 31321000119102 | Diabetes mellitus type 1 without retinopathy |
| ICD10CM | E11.44 | Type 2 diabetes mellitus with diabetic amyotrophy |
| SNOMED CT | 31211000119101 | Peripheral angiopathy due to type 1 diabetes mellitus |
| ICD10CM | E11.43 | Type 2 diabetes mellitus with diabetic autonomic (poly)neuropathy |
| SNOMED CT | 28331000119107 | Retinal edema due to type 2 diabetes mellitus |
| ICD10CM | E11.42 | Type 2 diabetes mellitus with diabetic polyneuropathy |
| SNOMED CT | 28032008 | Insulin dependent diabetes mellitus type 1B |
| ICD10CM | E11.41 | Type 2 diabetes mellitus with diabetic mononeuropathy |
| ICD9CM | 250.93 | Diabetes with unspecified complication, type I [juvenile type], uncontrolled |
| ICD10CM | E11.40 | Type 2 diabetes mellitus with diabetic neuropathy, unspecified |
| ICD9CM | 250.92 | Diabetes with unspecified complication, type II or unspecified type, uncontrolled |
| ICD10CM | E11.39 | Type 2 diabetes mellitus with other diabetic ophthalmic complication |
| ICD9CM | 250.91 | Diabetes with unspecified complication, type I [juvenile type], not stated as uncontrolled |
| ICD10CM | E11.37X9 | Type 2 diabetes mellitus with diabetic macular edema, resolved following treatment, unspecified eye |
| ICD9CM | 250.90 | Diabetes with unspecified complication, type II or unspecified type, not stated as uncontrolled |
| ICD10CM | E11.37X3 | Type 2 diabetes mellitus with diabetic macular edema, resolved following treatment, bilateral |
| ICD9CM | 250.83 | Diabetes with other specified manifestations, type I [juvenile type], uncontrolled |
| ICD9CM | 250.82 | Diabetes with other specified manifestations, type II or unspecified type, uncontrolled |

Continued on next page

**Table 6 – continued from previous page**

| CodeSystem | ConceptCode | ConceptName |
|---|---|---|
| ICD9CM | 250.81 | Diabetes with other specified manifestations, type I [juvenile type], not stated as uncontrolled |
| ICD9CM | 250.80 | Diabetes with other specified manifestations, type II or unspecified type, not stated as uncontrolled |
| ICD9CM | 250.73 | Diabetes with peripheral circulatory disorders, type I [juvenile type], uncontrolled |
| ICD9CM | 250.72 | Diabetes with peripheral circulatory disorders, type II or unspecified type, uncontrolled |
| ICD9CM | 250.71 | Diabetes with peripheral circulatory disorders, type I [juvenile type], not stated as uncontrolled |
| ICD9CM | 250.70 | Diabetes with peripheral circulatory disorders, type II or unspecified type, not stated as uncontrolled |
| ICD9CM | 250.63 | Diabetes with neurological manifestations, type I [juvenile type], uncontrolled |
| ICD9CM | 250.62 | Diabetes with neurological manifestations, type II or unspecified type, uncontrolled |
| ICD9CM | 250.61 | Diabetes with neurological manifestations, type I [juvenile type], not stated as uncontrolled |
| ICD9CM | 250.60 | Diabetes with neurological manifestations, type II or unspecified type, not stated as uncontrolled |
| ICD9CM | 250.53 | Diabetes with ophthalmic manifestations, type I [juvenile type], uncontrolled |
| ICD9CM | 250.52 | Diabetes with ophthalmic manifestations, type II or unspecified type, uncontrolled |
| ICD9CM | 250.51 | Diabetes with ophthalmic manifestations, type I [juvenile type], not stated as uncontrolled |
| ICD9CM | 250.50 | Diabetes with ophthalmic manifestations, type II or unspecified type, not stated as uncontrolled |
| ICD9CM | 250.43 | Diabetes with renal manifestations, type I [juvenile type], uncontrolled |
| ICD9CM | 250.42 | Diabetes with renal manifestations, type II or unspecified type, uncontrolled |
| ICD9CM | 250.41 | Diabetes with renal manifestations, type I [juvenile type], not stated as uncontrolled |
| ICD9CM | 250.40 | Diabetes with renal manifestations, type II or unspecified type, not stated as uncontrolled |
| ICD9CM | 250.33 | Diabetes with other coma, type I [juvenile type], uncontrolled |
| ICD9CM | 250.32 | Diabetes with other coma, type II or unspecified type, uncontrolled |
| ICD9CM | 250.31 | Diabetes with other coma, type I [juvenile type], not stated as uncontrolled |

Continued on next page

**Table 6 – continued from previous page**

| CodeSystem | ConceptCode | ConceptName |
|---|---|---|
| ICD9CM | 250.30 | Diabetes with other coma, type II or unspecified type, not stated as uncontrolled |
| ICD9CM | 250.23 | Diabetes with hyperosmolarity, type I [juvenile type], uncontrolled |
| ICD9CM | 250.22 | Diabetes with hyperosmolarity, type II or unspecified type, uncontrolled |
| ICD9CM | 250.21 | Diabetes with hyperosmolarity, type I [juvenile type], not stated as uncontrolled |
| ICD9CM | 250.20 | Diabetes with hyperosmolarity, type II or unspecified type, not stated as uncontrolled |
| ICD9CM | 250.13 | Diabetes with ketoacidosis, type I [juvenile type], uncontrolled |
| ICD9CM | 250.12 | Diabetes with ketoacidosis, type II or unspecified type, uncontrolled |
| ICD9CM | 250.11 | Diabetes with ketoacidosis, type I [juvenile type], not stated as uncontrolled |
| ICD9CM | 250.10 | Diabetes with ketoacidosis, type II or unspecified type, not stated as uncontrolled |
| ICD9CM | 250.03 | Diabetes mellitus without mention of complication, type I [juvenile type], uncontrolled |
| SNOMED CT | 82551000119103 | Rubeosis iridis due to type 2 diabetes mellitus |
| SNOMED CT | 82541000119100 | Traction detachment of retina due to type 2 diabetes mellitus |
| SNOMED CT | 81531005 | Type 2 diabetes mellitus in obese |
| SNOMED CT | 770098001 | Cranial nerve palsy with type I diabetes mellitus |
| SNOMED CT | 739681000 | Disorder of eye due to type 1 diabetes mellitus |
| SNOMED CT | 719216001 | Hypoglycemic coma due to type 2 diabetes mellitus |
| SNOMED CT | 71791000119104 | Peripheral neuropathy co-occurrent and due to type 1 diabetes mellitus |
| SNOMED CT | 71721000119101 | Nephrotic syndrome due to type 1 diabetes mellitus |
| SNOMED CT | 71441000119104 | Nephrotic syndrome due to type 2 diabetes mellitus |
| SNOMED CT | 713706002 | Polyneuropathy due to type 2 diabetes mellitus |
| SNOMED CT | 713705003 | Polyneuropathy due to type 1 diabetes mellitus |
| SNOMED CT | 713703005 | Gastroparesis due to type 2 diabetes mellitus |
| SNOMED CT | 713702000 | Gastroparesis due to type 1 diabetes mellitus |
| SNOMED CT | 712883005 | Autonomic neuropathy due to type 2 diabetes mellitus |
| SNOMED CT | 712882000 | Autonomic neuropathy due to type 1 diabetes mellitus |
| SNOMED CT | 691000119103 | Erectile dysfunction due to type 1 diabetes mellitus |
| ICD9CM | 648.04 | Diabetes mellitus of mother, complicating pregnancy, childbirth, or the puerperium, postpartum condition or complication |

Continued on next page

**Table 6 – continued from previous page**

| CodeSystem | ConceptCode | ConceptName |
|---|---|---|
| ICD9CM | 648.03 | Diabetes mellitus of mother, complicating pregnancy, childbirth, or the puerperium, antepartum condition or complication |
| ICD9CM | 648.02 | Diabetes mellitus of mother, complicating pregnancy, childbirth, or the puerperium, delivered, with mention of postpartum complication |
| ICD9CM | 648.01 | Diabetes mellitus of mother, complicating pregnancy, childbirth, or the puerperium, delivered, with or without mention of antepartum condition |
| ICD9CM | 648.00 | Diabetes mellitus of mother, complicating pregnancy, childbirth, or the puerperium, unspecified as to episode of care or not applicable |
| SNOMED CT | 60991000119100 | Blindness due to type 1 diabetes mellitus |
| SNOMED CT | 60971000119101 | Proliferative retinopathy due to type 1 diabetes mellitus |
| SNOMED CT | 60961000119107 | Nonproliferative diabetic retinopathy due to type 1 diabetes mellitus |
| SNOMED CT | 609567009 | Pre-existing type 2 diabetes mellitus in pregnancy |
| SNOMED CT | 609566000 | Pregnancy and type 1 diabetes mellitus |
| SNOMED CT | 609564002 | Pre-existing type 1 diabetes mellitus in pregnancy |
| SNOMED CT | 609562003 | Maturity onset diabetes of the young, type 1 |
| SNOMED CT | 60951000119105 | Blindness due to type 2 diabetes mellitus |
| SNOMED CT | 46635009 | T1DM - type 1 diabetes mellitus |
| SNOMED CT | 44054006 | Type 2 diabetes mellitus |
| SNOMED CT | 428007007 | Erectile dysfunction due to type 2 diabetes mellitus |
| SNOMED CT | 427571000 | Lumbosacral radiculoplexus neuropathy due to type 1 diabetes mellitus |
| SNOMED CT | 427027005 | Lumbosacral radiculoplexus neuropathy due to type 2 diabetes mellitus |
| SNOMED CT | 422166005 | Peripheral circulatory disorder due to type 2 diabetes mellitus |
| SNOMED CT | 422099009 | Disorder of eye due to type 2 diabetes mellitus |
| SNOMED CT | 422034002 | Retinopathy due to type 2 diabetes mellitus |
| SNOMED CT | 421893009 | Renal disorder due to type 1 diabetes mellitus |
| SNOMED CT | 421847006 | Ketoacidotic coma due to type 2 diabetes mellitus |
| SNOMED CT | 421779007 | Exudative maculopathy due to type 2 diabetes mellitus |
| SNOMED CT | 421468001 | Disorder of nervous system due to type 1 diabetes mellitus |
| SNOMED CT | 421437000 | Hypoglycemic coma due to type 1 diabetes mellitus |
| SNOMED CT | 421365002 | Peripheral circulatory disorder due to type 1 diabetes mellitus |
| SNOMED CT | 421326000 | Disorder of nervous system due to type 2 diabetes mellitus |
| SNOMED CT | 421075007 | Ketoacidotic coma due to type 1 diabetes mellitus |
| SNOMED CT | 420918009 | Mononeuropathy due to type 1 diabetes mellitus |
| SNOMED CT | 420789003 | Retinopathy due to type 1 diabetes mellitus |

Continued on next page

**Table 6 – continued from previous page**

| CodeSystem | ConceptCode | ConceptName |
|---|---|---|
| SNOMED CT | 420486006 | Exudative maculopathy due to type 1 diabetes mellitus |
| SNOMED CT | 420436000 | Mononeuropathy due to type 2 diabetes mellitus |
| SNOMED CT | 420279001 | Renal disorder due to type 2 diabetes mellitus |
| SNOMED CT | 41911000119107 | Glaucoma due to type 2 diabetes mellitus |
| SNOMED CT | 368581000119106 | Neuropathy due to type 2 diabetes mellitus |
| SNOMED CT | 368521000119107 | Disorder of nerve co-occurrent and due to type 1 diabetes mellitus |
| SNOMED CT | 368101000119109 | Diabetes mellitus type 2 with periodontal disease |
| ICD9CM | 366.41 | Diabetic cataract |
| ICD10CM | E13.621 | Other specified diabetes mellitus with foot ulcer |
| ICD10CM | E13.620 | Other specified diabetes mellitus with diabetic dermatitis |
| ICD10CM | E13.618 | Other specified diabetes mellitus with other diabetic arthropathy |
| ICD10CM | E13.610 | Other specified diabetes mellitus with diabetic neuropathic arthropathy |
| ICD10CM | E13.59 | Other specified diabetes mellitus with other circulatory complications |
| ICD10CM | E13.52 | Other specified diabetes mellitus with diabetic peripheral angiopathy with gangrene |
| ICD10CM | E13.51 | Other specified diabetes mellitus with diabetic peripheral angiopathy without gangrene |
| ICD10CM | E13.49 | Other specified diabetes mellitus with other diabetic neurological complication |
| ICD10CM | E13.44 | Other specified diabetes mellitus with diabetic amyotrophy |
| ICD10CM | E13.43 | Other specified diabetes mellitus with diabetic autonomic (poly)neuropathy |
| ICD10CM | E13.42 | Other specified diabetes mellitus with diabetic polyneuropathy |
| ICD10CM | E13.40 | Other specified diabetes mellitus with diabetic neuropathy, unspecified |
| ICD10CM | E13.39 | Other specified diabetes mellitus with other diabetic ophthalmic complication |
| ICD10CM | E13.37X9 | Other specified diabetes mellitus with diabetic macular edema, resolved following treatment, unspecified eye |
| ICD10CM | E13.37X3 | Other specified diabetes mellitus with diabetic macular edema, resolved following treatment, bilateral |
| ICD10CM | E13.37X2 | Other specified diabetes mellitus with diabetic macular edema, resolved following treatment, left eye |
| ICD10CM | E13.37X1 | Other specified diabetes mellitus with diabetic macular edema, resolved following treatment, right eye |
| ICD10CM | E13.36 | Other specified diabetes mellitus with diabetic cataract |
| ICD10CM | E13.3599 | Other specified diabetes mellitus with proliferative diabetic retinopathy without macular edema, unspecified eye |

Continued on next page

**Table 6 – continued from previous page**

| CodeSystem | ConceptCode | ConceptName |
|---|---|---|
| ICD10CM | E13.3593 | Other specified diabetes mellitus with proliferative diabetic retinopathy without macular edema, bilateral |
| ICD10CM | E13.3592 | Other specified diabetes mellitus with proliferative diabetic retinopathy without macular edema, left eye |
| ICD10CM | E13.3591 | Other specified diabetes mellitus with proliferative diabetic retinopathy without macular edema, right eye |
| ICD10CM | E13.359 | Other specified diabetes mellitus with proliferative diabetic retinopathy without macular edema |
| ICD10CM | E13.3559 | Other specified diabetes mellitus with stable proliferative diabetic retinopathy, unspecified eye |
| ICD10CM | E13.3553 | Other specified diabetes mellitus with stable proliferative diabetic retinopathy, bilateral |
| ICD10CM | E13.3552 | Other specified diabetes mellitus with stable proliferative diabetic retinopathy, left eye |
| ICD10CM | E13.3551 | Other specified diabetes mellitus with stable proliferative diabetic retinopathy, right eye |
| ICD10CM | E13.3549 | Other specified diabetes mellitus with proliferative diabetic retinopathy with combined traction retinal detachment and rhegmatogenous retinal detachment, unspecified eye |
| ICD10CM | E13.3543 | Other specified diabetes mellitus with proliferative diabetic retinopathy with combined traction retinal detachment and rhegmatogenous retinal detachment, bilateral |
| ICD10CM | E13.3542 | Other specified diabetes mellitus with proliferative diabetic retinopathy with combined traction retinal detachment and rhegmatogenous retinal detachment, left eye |
| ICD10CM | E13.3541 | Other specified diabetes mellitus with proliferative diabetic retinopathy with combined traction retinal detachment and rhegmatogenous retinal detachment, right eye |
| ICD10CM | E13.3539 | Other specified diabetes mellitus with proliferative diabetic retinopathy with traction retinal detachment not involving the macula, unspecified eye |
| ICD10CM | E13.3533 | Other specified diabetes mellitus with proliferative diabetic retinopathy with traction retinal detachment not involving the macula, bilateral |
| ICD10CM | E13.3532 | Other specified diabetes mellitus with proliferative diabetic retinopathy with traction retinal detachment not involving the macula, left eye |
| ICD10CM | E13.3531 | Other specified diabetes mellitus with proliferative diabetic retinopathy with traction retinal detachment not involving the macula, right eye |
| ICD10CM | E13.3529 | Other specified diabetes mellitus with proliferative diabetic retinopathy with traction retinal detachment involving the macula, unspecified eye |

Continued on next page

**Table 6 – continued from previous page**

| CodeSystem | ConceptCode | ConceptName |
|---|---|---|
| ICD10CM | E13.3523 | Other specified diabetes mellitus with proliferative diabetic retinopathy with traction retinal detachment involving the macula, bilateral |
| ICD10CM | E13.3522 | Other specified diabetes mellitus with proliferative diabetic retinopathy with traction retinal detachment involving the macula, left eye |
| ICD10CM | E13.3521 | Other specified diabetes mellitus with proliferative diabetic retinopathy with traction retinal detachment involving the macula, right eye |
| ICD10CM | E13.3519 | Other specified diabetes mellitus with proliferative diabetic retinopathy with macular edema, unspecified eye |
| ICD10CM | E13.3513 | Other specified diabetes mellitus with proliferative diabetic retinopathy with macular edema, bilateral |
| ICD10CM | E13.3512 | Other specified diabetes mellitus with proliferative diabetic retinopathy with macular edema, left eye |
| ICD10CM | E13.3511 | Other specified diabetes mellitus with proliferative diabetic retinopathy with macular edema, right eye |
| ICD10CM | E13.351 | Other specified diabetes mellitus with proliferative diabetic retinopathy with macular edema |
| ICD10CM | E13.3499 | Other specified diabetes mellitus with severe nonproliferative diabetic retinopathy without macular edema, unspecified eye |
| ICD10CM | E13.3493 | Other specified diabetes mellitus with severe nonproliferative diabetic retinopathy without macular edema, bilateral |
| ICD10CM | E13.3492 | Other specified diabetes mellitus with severe nonproliferative diabetic retinopathy without macular edema, left eye |
| ICD10CM | E13.3491 | Other specified diabetes mellitus with severe nonproliferative diabetic retinopathy without macular edema, right eye |
| ICD10CM | E13.349 | Other specified diabetes mellitus with severe nonproliferative diabetic retinopathy without macular edema |
| ICD10CM | E13.3419 | Other specified diabetes mellitus with severe nonproliferative diabetic retinopathy with macular edema, unspecified eye |
| ICD10CM | E13.41 | Other specified diabetes mellitus with diabetic mononeuropathy |
| ICD10CM | E13.3413 | Other specified diabetes mellitus with severe nonproliferative diabetic retinopathy with macular edema, bilateral |
| ICD10CM | E13.3412 | Other specified diabetes mellitus with severe nonproliferative diabetic retinopathy with macular edema, left eye |
| ICD10CM | E13.3411 | Other specified diabetes mellitus with severe nonproliferative diabetic retinopathy with macular edema, right eye |
| ICD10CM | E13.341 | Other specified diabetes mellitus with severe nonproliferative diabetic retinopathy with macular edema |

#### 3.4 Immunocompromised

Table 7: Concept codes used to define patients with an immunocompromising condition.

| CodeSystem | ConceptCode | ConceptName |
|---|---|---|
| ICD10CM | C01 | Malignant neoplasm of base of tongue |
| ICD10CM | C02 | Malignant neoplasm of other and unspecified parts of tongue |
| ICD10CM | C03 | Malignant neoplasm of gum |
| ICD10CM | C04 | Malignant neoplasm of floor of mouth |
| ICD10CM | C05 | Malignant neoplasm of palate |
| ICD10CM | C06 | Malignant neoplasm of other and unspecified parts of mouth |
| ICD10CM | C07 | Malignant neoplasm of parotid gland |
| ICD10CM | C08 | Malignant neoplasm of other and unspecified major salivary glands |
| ICD10CM | C09 | Malignant neoplasm of tonsil |
| ICD10CM | C10 | Malignant neoplasm of oropharynx |
| ICD10CM | C11 | Malignant neoplasm of nasopharynx |
| ICD10CM | C12 | Malignant neoplasm of pyriform sinus |
| ICD10CM | C13 | Malignant neoplasm of hypopharynx |
| ICD10CM | C14 | Malignant neoplasm of other and ill-defined sites in the lip, oral cavity and pharynx |
| ICD10CM | C15 | Malignant neoplasm of esophagus |
| ICD10CM | C16 | Malignant neoplasm of stomach |
| ICD10CM | C17 | Malignant neoplasm of small intestine |
| ICD10CM | C18 | Malignant neoplasm of colon |
| ICD10CM | C19 | Malignant neoplasm of rectosigmoid junction |
| ICD10CM | C20 | Malignant neoplasm of rectum |
| ICD10CM | C21 | Malignant neoplasm of anus and anal canal |
| ICD10CM | C22 | Malignant neoplasm of liver and intrahepatic bile ducts |
| ICD10CM | C23 | Malignant neoplasm of gallbladder |
| ICD10CM | C24 | Malignant neoplasm of other and unspecified parts of biliary tract |
| ICD10CM | C25 | Malignant neoplasm of pancreas |
| ICD10CM | C26 | Malignant neoplasm of other and ill-defined digestive organs |
| ICD10CM | C30 | Malignant neoplasm of nasal cavity and middle ear |
| ICD10CM | C31 | Malignant neoplasm of accessory sinuses |
| ICD10CM | C32 | Malignant neoplasm of larynx |
| ICD10CM | C33 | Malignant neoplasm of trachea |
| ICD10CM | C34 | Malignant neoplasm of bronchus and lung |
| ICD10CM | C37 | Malignant neoplasm of thymus |
| ICD10CM | C38 | Malignant neoplasm of heart, mediastinum and pleura |
| ICD10CM | C39 | Malignant neoplasm of other and ill-defined sites in the respiratory system and intrathoracic organs |
| ICD10CM | C40 | Malignant neoplasm of bone and articular cartilage of limbs |

Continued on next page

**Table 7 – continued from previous page**

| CodeSystem | ConceptCode | ConceptName |
|---|---|---|
| ICD10CM | C41 | Malignant neoplasm of bone and articular cartilage of other and unspecified sites |
| ICD10CM | C43 | Malignant melanoma of skin |
| ICD10CM | C44 | Other and unspecified malignant neoplasm of skin |
| ICD10CM | C45 | Mesothelioma |
| ICD10CM | C46 | Kaposi's sarcoma |
| ICD10CM | C47 | Malignant neoplasm of peripheral nerves and autonomic nervous system |
| ICD10CM | C48 | Malignant neoplasm of retroperitoneum and peritoneum |
| ICD10CM | C49 | Malignant neoplasm of other connective and soft tissue |
| ICD10CM | C50 | Malignant neoplasm of breast |
| ICD10CM | C51 | Malignant neoplasm of vulva |
| ICD10CM | C52 | Malignant neoplasm of vagina |
| ICD10CM | C53 | Malignant neoplasm of cervix uteri |
| ICD10CM | C54 | Malignant neoplasm of corpus uteri |
| ICD10CM | C55 | Malignant neoplasm of uterus, part unspecified |
| ICD10CM | C56 | Malignant neoplasm of ovary |
| ICD10CM | C57 | Malignant neoplasm of other and unspecified female genital organs |
| ICD10CM | C58 | Malignant neoplasm of placenta |
| ICD10CM | C60 | Malignant neoplasm of penis |
| ICD10CM | C61 | Malignant neoplasm of prostate |
| ICD10CM | C62 | Malignant neoplasm of testis |
| ICD10CM | C63 | Malignant neoplasm of other and unspecified male genital organs |
| ICD10CM | C64 | Malignant neoplasm of kidney, except renal pelvis |
| ICD10CM | C65 | Malignant neoplasm of renal pelvis |
| ICD10CM | C66 | Malignant neoplasm of ureter |
| ICD10CM | C67 | Malignant neoplasm of bladder |
| ICD10CM | C68 | Malignant neoplasm of other and unspecified urinary organs |
| ICD10CM | C69 | Malignant neoplasm of eye and adnexa |
| ICD10CM | C70 | Malignant neoplasm of meninges |
| ICD10CM | C71 | Malignant neoplasm of brain |
| ICD10CM | C72 | Malignant neoplasm of spinal cord, cranial nerves and other parts of central nervous system |
| ICD10CM | C73 | Malignant neoplasm of thyroid gland |
| ICD10CM | C74 | Malignant neoplasm of adrenal gland |
| ICD10CM | C75 | Malignant neoplasm of other endocrine glands and related structures |
| ICD10CM | C76 | Malignant neoplasm of other and ill-defined sites |
| ICD10CM | C77 | Secondary and unspecified malignant neoplasm of lymph nodes |

Continued on next page

**Table 7 – continued from previous page**

| CodeSystem | ConceptCode | ConceptName |
|---|---|---|
| ICD10CM | C78 | Secondary malignant neoplasm of respiratory and digestive organs |
| ICD10CM | C79 | Secondary malignant neoplasm of other and unspecified sites |
| ICD10CM | C7A | Malignant neuroendocrine tumors |
| ICD10CM | C7B | Secondary neuroendocrine tumors |
| ICD10CM | C80 | Malignant neoplasm without specification of site |
| ICD10CM | C81 | Hodgkin lymphoma |
| ICD10CM | C82 | Follicular lymphoma |
| ICD10CM | C83 | Non-follicular lymphoma |
| ICD10CM | C84 | Mature T/NK-cell lymphomas |
| ICD10CM | C85 | Other specified and unspecified types of non-Hodgkin lymphoma |
| ICD10CM | C86 | Other specified types of T/NK-cell lymphoma |
| ICD10CM | C88 | Malignant immunoproliferative diseases and certain other B-cell lymphomas |
| ICD10CM | C90 | Multiple myeloma and malignant plasma cell neoplasms |
| ICD10CM | C91 | Lymphoid leukemia |
| ICD10CM | C92 | Myeloid leukemia |
| ICD10CM | C93 | Monocytic leukemia |
| ICD10CM | C94 | Other leukemias of specified cell type |
| ICD10CM | C95 | Leukemia of unspecified cell type |
| ICD10CM | C96 | Other and unspecified malignant neoplasms of lymphoid, hematopoietic and related tissue |
| ICD10CM | D27.9 | Benign neoplasm of unspecified ovary |
| ICD10CM | D3A | Benign neuroendocrine tumors |
| ICD10CM | D46 | Myelodysplastic syndromes |
| ICD10CM | D47.Z1 | Post-transplant lymphoproliferative disorder (PTLD) |
| ICD10CM | D61.0 | Constitutional aplastic anemia |
| ICD10CM | D61.09 | Other constitutional aplastic anemia |
| ICD10CM | D61.2 | Aplastic anemia due to other external agents |
| ICD10CM | D61.9 | Aplastic anemia, unspecified |
| ICD10CM | D70.0 | Congenital agranulocytosis |
| ICD10CM | D71 | Functional disorders of polymorphonuclear neutrophils |
| ICD10CM | D72.89 | Other specified disorders of white blood cells |
| ICD10CM | D80 | Immunodeficiency with predominantly antibody defects |
| ICD10CM | D81 | Combined immunodeficiencies |
| ICD10CM | D82 | Immunodeficiency associated with other major defects |
| ICD10CM | D83 | Common variable immunodeficiency |
| ICD10CM | D84 | Other immunodeficiencies |
| ICD10CM | D89 | Other disorders involving the immune mechanism, not elsewhere classified |
| ICD10CM | G35 | Multiple sclerosis |

Continued on next page

**Table 7 – continued from previous page**

| CodeSystem | ConceptCode | ConceptName |
|---|---|---|
| ICD10CM | J67.9 | Hypersensitivity pneumonitis due to unspecified organic dust |
| ICD10CM | K70.3 | Alcoholic cirrhosis of liver |
| ICD10CM | K70.4 | Alcoholic hepatic failure |
| ICD10CM | K72 | Hepatic failure, not elsewhere classified |
| ICD10CM | K74.3 | Primary biliary cirrhosis |
| ICD10CM | K74.4 | Secondary biliary cirrhosis |
| ICD10CM | K74.5 | Biliary cirrhosis, unspecified |
| ICD10CM | K74.6 | Other and unspecified cirrhosis of liver |
| ICD10CM | L40.54 | Psoriatic juvenile arthropathy |
| ICD10CM | L40.59 | Other psoriatic arthropathy |
| ICD10CM | L93.0 | Discoid lupus erythematosus |
| ICD10CM | L93.2 | Other local lupus erythematosus |
| ICD10CM | L94 | Other localized connective tissue disorders |
| ICD10CM | M05 | Rheumatoid arthritis with rheumatoid factor |
| ICD10CM | M06 | Other rheumatoid arthritis |
| ICD10CM | M07 | Enteropathic arthropathies |
| ICD10CM | M08 | Juvenile arthritis |
| ICD10CM | M30 | Polyarteritis nodosa and related conditions |
| ICD10CM | M31.3 | Wegener's granulomatosis |
| ICD10CM | M31.5 | Giant cell arteritis with polymyalgia rheumatica |
| ICD10CM | M32 | Systemic lupus erythematosus (SLE) |
| ICD10CM | M33 | Dermatopolymyositis |
| ICD10CM | M34 | Systemic sclerosis [scleroderma] |
| ICD10CM | M35.3 | Polymyalgia rheumatica |
| ICD10CM | M35.8 | Other specified systemic involvement of connective tissue |
| ICD10CM | M35.9 | Systemic involvement of connective tissue, unspecified |
| ICD10CM | M46 | Other inflammatory spondylopathies |
| ICD10CM | T78.40 | Allergy, unspecified |
| ICD10CM | T86 | Complications of transplanted organs and tissue |
| ICD10CM | Z48.2 | Encounter for aftercare following organ transplant |
| ICD10CM | Z51.0 | Encounter for antineoplastic radiation therapy |
| ICD10CM | Z51.1 | Encounter for antineoplastic chemotherapy and immunotherapy |
| ICD10CM | Z94 | Transplanted organ and tissue status |
| ICD10CM | Z98.85 | Transplanted organ removal status |
| ICD10CM | C02.0 | Malignant neoplasm of dorsal surface of tongue |
| ICD10CM | C02.1 | Malignant neoplasm of border of tongue |
| ICD10CM | C02.2 | Malignant neoplasm of ventral surface of tongue |
| ICD10CM | C02.3 | Malignant neoplasm of anterior two-thirds of tongue, part unspecified |
| ICD10CM | C02.4 | Malignant neoplasm of lingual tonsil |
| ICD10CM | C02.8 | Malignant neoplasm of overlapping sites of tongue |

Continued on next page

**Table 7 – continued from previous page**

| CodeSystem | ConceptCode | ConceptName |
|---|---|---|
| ICD10CM | C02.9 | Malignant neoplasm of tongue, unspecified |
| ICD10CM | C03.0 | Malignant neoplasm of upper gum |
| ICD10CM | C03.1 | Malignant neoplasm of lower gum |
| ICD10CM | C03.9 | Malignant neoplasm of gum, unspecified |
| ICD10CM | C04.0 | Malignant neoplasm of anterior floor of mouth |
| ICD10CM | C04.1 | Malignant neoplasm of lateral floor of mouth |
| ICD10CM | C04.8 | Malignant neoplasm of overlapping sites of floor of mouth |
| ICD10CM | C04.9 | Malignant neoplasm of floor of mouth, unspecified |
| ICD10CM | C05.0 | Malignant neoplasm of hard palate |
| ICD10CM | C05.1 | Malignant neoplasm of soft palate |
| ICD10CM | C05.2 | Malignant neoplasm of uvula |
| ICD10CM | C05.8 | Malignant neoplasm of overlapping sites of palate |
| ICD10CM | C05.9 | Malignant neoplasm of palate, unspecified |
| ICD10CM | C06.0 | Malignant neoplasm of cheek mucosa |
| ICD10CM | C06.1 | Malignant neoplasm of vestibule of mouth |
| ICD10CM | C06.2 | Malignant neoplasm of retromolar area |
| ICD10CM | C06.8 | Malignant neoplasm of overlapping sites of other and unspecified parts of mouth |
| ICD10CM | C06.80 | Malignant neoplasm of overlapping sites of unspecified parts of mouth |
| ICD10CM | C06.89 | Malignant neoplasm of overlapping sites of other parts of mouth |
| ICD10CM | C06.9 | Malignant neoplasm of mouth, unspecified |
| ICD10CM | C08.0 | Malignant neoplasm of submandibular gland |
| ICD10CM | C08.1 | Malignant neoplasm of sublingual gland |
| ICD10CM | C08.9 | Malignant neoplasm of major salivary gland, unspecified |
| ICD10CM | C09.0 | Malignant neoplasm of tonsillar fossa |
| ICD10CM | C09.1 | Malignant neoplasm of tonsillar pillar (anterior) (posterior) |
| ICD10CM | C09.8 | Malignant neoplasm of overlapping sites of tonsil |
| ICD10CM | C09.9 | Malignant neoplasm of tonsil, unspecified |
| ICD10CM | C10.0 | Malignant neoplasm of vallecula |
| ICD10CM | C10.1 | Malignant neoplasm of anterior surface of epiglottis |
| ICD10CM | C10.2 | Malignant neoplasm of lateral wall of oropharynx |
| ICD10CM | C10.3 | Malignant neoplasm of posterior wall of oropharynx |
| ICD10CM | C10.4 | Malignant neoplasm of branchial cleft |
| ICD10CM | C10.8 | Malignant neoplasm of overlapping sites of oropharynx |
| ICD10CM | C10.9 | Malignant neoplasm of oropharynx, unspecified |
| ICD10CM | C11.0 | Malignant neoplasm of superior wall of nasopharynx |
| ICD10CM | C11.1 | Malignant neoplasm of posterior wall of nasopharynx |
| ICD10CM | C11.2 | Malignant neoplasm of lateral wall of nasopharynx |
| ICD10CM | C11.3 | Malignant neoplasm of anterior wall of nasopharynx |
| ICD10CM | C11.8 | Malignant neoplasm of overlapping sites of nasopharynx |
| ICD10CM | C11.9 | Malignant neoplasm of nasopharynx, unspecified |

Continued on next page

**Table 7 – continued from previous page**

| CodeSystem | ConceptCode | ConceptName |
|---|---|---|
| ICD10CM | C13.0 | Malignant neoplasm of postcricoid region |
| ICD10CM | C13.1 | Malignant neoplasm of aryepiglottic fold, hypopharyngeal aspect |
| ICD10CM | C13.2 | Malignant neoplasm of posterior wall of hypopharynx |
| ICD10CM | C13.8 | Malignant neoplasm of overlapping sites of hypopharynx |
| ICD10CM | C13.9 | Malignant neoplasm of hypopharynx, unspecified |
| ICD10CM | C14.0 | Malignant neoplasm of pharynx, unspecified |
| ICD10CM | C14.2 | Malignant neoplasm of Waldeyer's ring |
| ICD10CM | C14.8 | Malignant neoplasm of overlapping sites of lip, oral cavity and pharynx |
| ICD10CM | C15.3 | Malignant neoplasm of upper third of esophagus |
| ICD10CM | C15.4 | Malignant neoplasm of middle third of esophagus |
| ICD10CM | C15.5 | Malignant neoplasm of lower third of esophagus |
| ICD10CM | C15.8 | Malignant neoplasm of overlapping sites of esophagus |
| ICD10CM | C15.9 | Malignant neoplasm of esophagus, unspecified |
| ICD10CM | C16.0 | Malignant neoplasm of cardia |
| ICD10CM | C16.1 | Malignant neoplasm of fundus of stomach |
| ICD10CM | C16.2 | Malignant neoplasm of body of stomach |
| ICD10CM | C16.3 | Malignant neoplasm of pyloric antrum |
| ICD10CM | C16.4 | Malignant neoplasm of pylorus |
| ICD10CM | C16.5 | Malignant neoplasm of lesser curvature of stomach, unspecified |
| ICD10CM | C16.6 | Malignant neoplasm of greater curvature of stomach, unspecified |
| ICD10CM | C16.8 | Malignant neoplasm of overlapping sites of stomach |
| ICD10CM | C16.9 | Malignant neoplasm of stomach, unspecified |
| ICD10CM | C17.0 | Malignant neoplasm of duodenum |
| ICD10CM | C17.1 | Malignant neoplasm of jejunum |
| ICD10CM | C17.2 | Malignant neoplasm of ileum |
| ICD10CM | C17.3 | Meckel's diverticulum, malignant |
| ICD10CM | C17.8 | Malignant neoplasm of overlapping sites of small intestine |
| ICD10CM | C17.9 | Malignant neoplasm of small intestine, unspecified |
| ICD10CM | C18.0 | Malignant neoplasm of cecum |
| ICD10CM | C18.1 | Malignant neoplasm of appendix |
| ICD10CM | C18.2 | Malignant neoplasm of ascending colon |
| ICD10CM | C18.3 | Malignant neoplasm of hepatic flexure |
| ICD10CM | C18.4 | Malignant neoplasm of transverse colon |
| ICD10CM | C18.5 | Malignant neoplasm of splenic flexure |
| ICD10CM | C18.6 | Malignant neoplasm of descending colon |
| ICD10CM | C18.7 | Malignant neoplasm of sigmoid colon |
| ICD10CM | C18.8 | Malignant neoplasm of overlapping sites of colon |
| ICD10CM | C18.9 | Malignant neoplasm of colon, unspecified |
| ICD10CM | C21.0 | Malignant neoplasm of anus, unspecified |

Continued on next page

**Table 7 – continued from previous page**

| CodeSystem | ConceptCode | ConceptName |
|---|---|---|
| ICD10CM | C21.1 | Malignant neoplasm of anal canal |
| ICD10CM | C21.2 | Malignant neoplasm of cloacogenic zone |
| ICD10CM | C21.8 | Malignant neoplasm of overlapping sites of rectum, anus and anal canal |
| ICD10CM | C22.0 | Liver cell carcinoma |
| ICD10CM | C22.1 | Intrahepatic bile duct carcinoma |
| ICD10CM | C22.2 | Hepatoblastoma |
| ICD10CM | C22.3 | Angiosarcoma of liver |
| ICD10CM | C22.4 | Other sarcomas of liver |
| ICD10CM | C22.7 | Other specified carcinomas of liver |
| ICD10CM | C22.8 | Malignant neoplasm of liver, primary, unspecified as to type |
| ICD10CM | C22.9 | Malignant neoplasm of liver, not specified as primary or secondary |
| ICD10CM | C24.0 | Malignant neoplasm of extrahepatic bile duct |
| ICD10CM | C24.1 | Malignant neoplasm of ampulla of Vater |
| ICD10CM | C24.8 | Malignant neoplasm of overlapping sites of biliary tract |
| ICD10CM | C24.9 | Malignant neoplasm of biliary tract, unspecified |
| ICD10CM | C25.0 | Malignant neoplasm of head of pancreas |
| ICD10CM | C25.1 | Malignant neoplasm of body of pancreas |
| ICD10CM | C25.2 | Malignant neoplasm of tail of pancreas |
| ICD10CM | C25.3 | Malignant neoplasm of pancreatic duct |
| ICD10CM | C25.4 | Malignant neoplasm of endocrine pancreas |
| ICD10CM | C25.7 | Malignant neoplasm of other parts of pancreas |
| ICD10CM | C25.8 | Malignant neoplasm of overlapping sites of pancreas |
| ICD10CM | C25.9 | Malignant neoplasm of pancreas, unspecified |
| ICD10CM | C26.0 | Malignant neoplasm of intestinal tract, part unspecified |
| ICD10CM | C26.1 | Malignant neoplasm of spleen |
| ICD10CM | C26.9 | Malignant neoplasm of ill-defined sites within the digestive system |
| ICD10CM | C30.0 | Malignant neoplasm of nasal cavity |
| ICD10CM | C30.1 | Malignant neoplasm of middle ear |
| ICD10CM | C31.0 | Malignant neoplasm of maxillary sinus |
| ICD10CM | C31.1 | Malignant neoplasm of ethmoidal sinus |
| ICD10CM | C31.2 | Malignant neoplasm of frontal sinus |
| ICD10CM | C31.3 | Malignant neoplasm of sphenoid sinus |
| ICD10CM | C31.8 | Malignant neoplasm of overlapping sites of accessory sinuses |
| ICD10CM | C31.9 | Malignant neoplasm of accessory sinus, unspecified |
| ICD10CM | C32.0 | Malignant neoplasm of glottis |
| ICD10CM | C32.1 | Malignant neoplasm of supraglottis |
| ICD10CM | C32.2 | Malignant neoplasm of subglottis |
| ICD10CM | C32.3 | Malignant neoplasm of laryngeal cartilage |
| ICD10CM | C32.8 | Malignant neoplasm of overlapping sites of larynx |
| ICD10CM | C32.9 | Malignant neoplasm of larynx, unspecified |

Continued on next page

**Table 7 – continued from previous page**

| CodeSystem | ConceptCode | ConceptName |
|---|---|---|
| ICD10CM | C34.0 | Malignant neoplasm of main bronchus |
| ICD10CM | C34.00 | Malignant neoplasm of unspecified main bronchus |
| ICD10CM | C34.01 | Malignant neoplasm of right main bronchus |
| ICD10CM | C34.02 | Malignant neoplasm of left main bronchus |
| ICD10CM | C34.1 | Malignant neoplasm of upper lobe, bronchus or lung |
| ICD10CM | C34.10 | Malignant neoplasm of upper lobe, unspecified bronchus or lung |
| ICD10CM | C34.11 | Malignant neoplasm of upper lobe, right bronchus or lung |
| ICD10CM | C34.12 | Malignant neoplasm of upper lobe, left bronchus or lung |
| ICD10CM | C34.2 | Malignant neoplasm of middle lobe, bronchus or lung |
| ICD10CM | C34.3 | Malignant neoplasm of lower lobe, bronchus or lung |
| ICD10CM | C34.30 | Malignant neoplasm of lower lobe, unspecified bronchus or lung |
| ICD10CM | C34.31 | Malignant neoplasm of lower lobe, right bronchus or lung |
| ICD10CM | C34.32 | Malignant neoplasm of lower lobe, left bronchus or lung |
| ICD10CM | C34.8 | Malignant neoplasm of overlapping sites of bronchus and lung |
| ICD10CM | C34.80 | Malignant neoplasm of overlapping sites of unspecified bronchus and lung |
| ICD10CM | C34.81 | Malignant neoplasm of overlapping sites of right bronchus and lung |
| ICD10CM | C34.82 | Malignant neoplasm of overlapping sites of left bronchus and lung |
| ICD10CM | C34.9 | Malignant neoplasm of unspecified part of bronchus or lung |
| ICD10CM | C34.90 | Malignant neoplasm of unspecified part of unspecified bronchus or lung |
| ICD10CM | C34.91 | Malignant neoplasm of unspecified part of right bronchus or lung |
| ICD10CM | C34.92 | Malignant neoplasm of unspecified part of left bronchus or lung |
| ICD10CM | C38.0 | Malignant neoplasm of heart |
| ICD10CM | C38.1 | Malignant neoplasm of anterior mediastinum |
| ICD10CM | C38.2 | Malignant neoplasm of posterior mediastinum |
| ICD10CM | C38.3 | Malignant neoplasm of mediastinum, part unspecified |
| ICD10CM | C38.4 | Malignant neoplasm of pleura |
| ICD10CM | C38.8 | Malignant neoplasm of overlapping sites of heart, mediastinum and pleura |
| ICD10CM | C39.0 | Malignant neoplasm of upper respiratory tract, part unspecified |
| ICD10CM | C39.9 | Malignant neoplasm of lower respiratory tract, part unspecified |
| ICD10CM | C40.0 | Malignant neoplasm of scapula and long bones of upper limb |

Continued on next page

**Table 7 – continued from previous page**

| CodeSystem | ConceptCode | ConceptName |
|---|---|---|
| ICD10CM | C40.00 | Malignant neoplasm of scapula and long bones of unspecified upper limb |
| ICD10CM | C40.01 | Malignant neoplasm of scapula and long bones of right upper limb |
| ICD10CM | C40.02 | Malignant neoplasm of scapula and long bones of left upper limb |
| ICD10CM | C40.1 | Malignant neoplasm of short bones of upper limb |
| ICD10CM | C40.10 | Malignant neoplasm of short bones of unspecified upper limb |
| ICD10CM | C40.11 | Malignant neoplasm of short bones of right upper limb |
| ICD10CM | C40.12 | Malignant neoplasm of short bones of left upper limb |
| ICD10CM | C40.2 | Malignant neoplasm of long bones of lower limb |
| ICD10CM | C40.20 | Malignant neoplasm of long bones of unspecified lower limb |
| ICD10CM | C40.21 | Malignant neoplasm of long bones of right lower limb |
| ICD10CM | C40.22 | Malignant neoplasm of long bones of left lower limb |
| ICD10CM | C40.3 | Malignant neoplasm of short bones of lower limb |
| ICD10CM | C40.30 | Malignant neoplasm of short bones of unspecified lower limb |
| ICD10CM | C40.31 | Malignant neoplasm of short bones of right lower limb |
| ICD10CM | C40.32 | Malignant neoplasm of short bones of left lower limb |
| ICD10CM | C40.8 | Malignant neoplasm of overlapping sites of bone and articular cartilage of limb |
| ICD10CM | C40.80 | Malignant neoplasm of overlapping sites of bone and articular cartilage of unspecified limb |
| ICD10CM | C40.81 | Malignant neoplasm of overlapping sites of bone and articular cartilage of right limb |
| ICD10CM | C40.82 | Malignant neoplasm of overlapping sites of bone and articular cartilage of left limb |
| ICD10CM | C40.9 | Malignant neoplasm of unspecified bones and articular cartilage of limb |
| ICD10CM | C40.90 | Malignant neoplasm of unspecified bones and articular cartilage of unspecified limb |
| ICD10CM | C40.91 | Malignant neoplasm of unspecified bones and articular cartilage of right limb |
| ICD10CM | C40.92 | Malignant neoplasm of unspecified bones and articular cartilage of left limb |
| ICD10CM | C41.0 | Malignant neoplasm of bones of skull and face |
| ICD10CM | C41.1 | Malignant neoplasm of mandible |
| ICD10CM | C41.2 | Malignant neoplasm of vertebral column |
| ICD10CM | C41.3 | Malignant neoplasm of ribs, sternum and clavicle |
| ICD10CM | C41.4 | Malignant neoplasm of pelvic bones, sacrum and coccyx |
| ICD10CM | C41.9 | Malignant neoplasm of bone and articular cartilage, unspecified |
| ICD10CM | C43.0 | Malignant melanoma of lip |

Continued on next page

**Table 7 – continued from previous page**

| CodeSystem | ConceptCode | ConceptName |
|---|---|---|
| ICD10CM | C43.1 | Malignant melanoma of eyelid, including canthus |
| ICD10CM | C43.10 | Malignant melanoma of unspecified eyelid, including canthus |
| ICD10CM | C43.11 | Malignant melanoma of right eyelid, including canthus |
| ICD10CM | C43.111 | Malignant melanoma of right upper eyelid, including canthus |
| ICD10CM | C43.112 | Malignant melanoma of right lower eyelid, including canthus |
| ICD10CM | C43.12 | Malignant melanoma of left eyelid, including canthus |
| ICD10CM | C43.121 | Malignant melanoma of left upper eyelid, including canthus |
| ICD10CM | C43.122 | Malignant melanoma of left lower eyelid, including canthus |
| ICD10CM | C43.2 | Malignant melanoma of ear and external auricular canal |
| ICD10CM | C43.20 | Malignant melanoma of unspecified ear and external auricular canal |
| ICD10CM | C43.21 | Malignant melanoma of right ear and external auricular canal |
| ICD10CM | C43.22 | Malignant melanoma of left ear and external auricular canal |
| ICD10CM | C43.3 | Malignant melanoma of other and unspecified parts of face |
| ICD10CM | C43.30 | Malignant melanoma of unspecified part of face |
| ICD10CM | C43.31 | Malignant melanoma of nose |
| ICD10CM | C43.39 | Malignant melanoma of other parts of face |
| ICD10CM | C43.4 | Malignant melanoma of scalp and neck |
| ICD10CM | C43.5 | Malignant melanoma of trunk |
| ICD10CM | C43.51 | Malignant melanoma of anal skin |
| ICD10CM | C43.52 | Malignant melanoma of skin of breast |
| ICD10CM | C43.59 | Malignant melanoma of other part of trunk |
| ICD10CM | C43.6 | Malignant melanoma of upper limb, including shoulder |
| ICD10CM | C43.60 | Malignant melanoma of unspecified upper limb, including shoulder |
| ICD10CM | C43.61 | Malignant melanoma of right upper limb, including shoulder |
| ICD10CM | C43.62 | Malignant melanoma of left upper limb, including shoulder |
| ICD10CM | C43.7 | Malignant melanoma of lower limb, including hip |
| ICD10CM | C43.70 | Malignant melanoma of unspecified lower limb, including hip |
| ICD10CM | C43.71 | Malignant melanoma of right lower limb, including hip |
| ICD10CM | C43.72 | Malignant melanoma of left lower limb, including hip |
| ICD10CM | C43.8 | Malignant melanoma of overlapping sites of skin |
| ICD10CM | C43.9 | Malignant melanoma of skin, unspecified |
| ICD10CM | C44.0 | Other and unspecified malignant neoplasm of skin of lip |
| ICD10CM | C44.00 | Unspecified malignant neoplasm of skin of lip |
| ICD10CM | C44.01 | Basal cell carcinoma of skin of lip |
| ICD10CM | C44.02 | Squamous cell carcinoma of skin of lip |
| ICD10CM | C44.09 | Other specified malignant neoplasm of skin of lip |

Continued on next page

**Table 7 – continued from previous page**

| CodeSystem | ConceptCode | ConceptName |
|---|---|---|
| ICD10CM | C44.1 | Other and unspecified malignant neoplasm of skin of eyelid, including canthus |
| ICD10CM | C44.10 | Unspecified malignant neoplasm of skin of eyelid, including canthus |
| ICD10CM | C44.101 | Unspecified malignant neoplasm of skin of unspecified eyelid, including canthus |
| ICD10CM | C44.102 | Unspecified malignant neoplasm of skin of right eyelid, including canthus |
| ICD10CM | C44.1021 | Unspecified malignant neoplasm of skin of right upper eyelid, including canthus |
| ICD10CM | C44.1022 | Unspecified malignant neoplasm of skin of right lower eyelid, including canthus |
| ICD10CM | C44.109 | Unspecified malignant neoplasm of skin of left eyelid, including canthus |
| ICD10CM | C44.1091 | Unspecified malignant neoplasm of skin of left upper eyelid, including canthus |
| ICD10CM | C44.1092 | Unspecified malignant neoplasm of skin of left lower eyelid, including canthus |
| ICD10CM | C44.11 | Basal cell carcinoma of skin of eyelid, including canthus |
| ICD10CM | C44.111 | Basal cell carcinoma of skin of unspecified eyelid, including canthus |
| ICD10CM | C44.112 | Basal cell carcinoma of skin of right eyelid, including canthus |
| ICD10CM | C44.1121 | Basal cell carcinoma of skin of right upper eyelid, including canthus |
| ICD10CM | C44.1122 | Basal cell carcinoma of skin of right lower eyelid, including canthus |
| ICD10CM | C44.119 | Basal cell carcinoma of skin of left eyelid, including canthus |
| ICD10CM | C44.1191 | Basal cell carcinoma of skin of left upper eyelid, including canthus |
| ICD10CM | C44.1192 | Basal cell carcinoma of skin of left lower eyelid, including canthus |
| ICD10CM | C44.12 | Squamous cell carcinoma of skin of eyelid, including canthus |
| ICD10CM | C44.121 | Squamous cell carcinoma of skin of unspecified eyelid, including canthus |
| ICD10CM | C44.122 | Squamous cell carcinoma of skin of right eyelid, including canthus |
| ICD10CM | C44.1221 | Squamous cell carcinoma of skin of right upper eyelid, including canthus |
| ICD10CM | C44.1222 | Squamous cell carcinoma of skin of right lower eyelid, including canthus |
| ICD10CM | C44.129 | Squamous cell carcinoma of skin of left eyelid, including canthus |

Continued on next page

**Table 7 – continued from previous page**

| CodeSystem | ConceptCode | ConceptName |
|---|---|---|
| ICD10CM | C44.1291 | Squamous cell carcinoma of skin of left upper eyelid, including canthus |
| ICD10CM | C44.1292 | Squamous cell carcinoma of skin of left lower eyelid, including canthus |
| ICD10CM | C44.13 | Sebaceous cell carcinoma of skin of eyelid, including canthus |
| ICD10CM | C44.131 | Sebaceous cell carcinoma of skin of unspecified eyelid, including canthus |
| ICD10CM | C44.132 | Sebaceous cell carcinoma of skin of right eyelid, including canthus |
| ICD10CM | C44.1321 | Sebaceous cell carcinoma of skin of right upper eyelid, including canthus |
| ICD10CM | C44.1322 | Sebaceous cell carcinoma of skin of right lower eyelid, including canthus |
| ICD10CM | C44.139 | Sebaceous cell carcinoma of skin of left eyelid, including canthus |
| ICD10CM | C44.1391 | Sebaceous cell carcinoma of skin of left upper eyelid, including canthus |
| ICD10CM | C44.1392 | Sebaceous cell carcinoma of skin of left lower eyelid, including canthus |
| ICD10CM | C44.19 | Other specified malignant neoplasm of skin of eyelid, including canthus |
| ICD10CM | C44.191 | Other specified malignant neoplasm of skin of unspecified eyelid, including canthus |
| ICD10CM | C44.192 | Other specified malignant neoplasm of skin of right eyelid, including canthus |
| ICD10CM | C44.1921 | Other specified malignant neoplasm of skin of right upper eyelid, including canthus |
| ICD10CM | C44.1922 | Other specified malignant neoplasm of skin of right lower eyelid, including canthus |
| ICD10CM | C44.199 | Other specified malignant neoplasm of skin of left eyelid, including canthus |
| ICD10CM | C44.1991 | Other specified malignant neoplasm of skin of left upper eyelid, including canthus |
| ICD10CM | C44.1992 | Other specified malignant neoplasm of skin of left lower eyelid, including canthus |
| ICD10CM | C44.2 | Other and unspecified malignant neoplasm of skin of ear and external auricular canal |
| ICD10CM | C44.20 | Unspecified malignant neoplasm of skin of ear and external auricular canal |
| ICD10CM | C44.201 | Unspecified malignant neoplasm of skin of unspecified ear and external auricular canal |
| ICD10CM | C44.202 | Unspecified malignant neoplasm of skin of right ear and external auricular canal |

Continued on next page

**Table 7 – continued from previous page**

| CodeSystem | ConceptCode | ConceptName |
|---|---|---|
| ICD10CM | C44.209 | Unspecified malignant neoplasm of skin of left ear and external auricular canal |
| ICD10CM | C44.21 | Basal cell carcinoma of skin of ear and external auricular canal |
| ICD10CM | C44.211 | Basal cell carcinoma of skin of unspecified ear and external auricular canal |
| ICD10CM | C44.212 | Basal cell carcinoma of skin of right ear and external auricular canal |
| ICD10CM | C44.219 | Basal cell carcinoma of skin of left ear and external auricular canal |
| ICD10CM | C44.22 | Squamous cell carcinoma of skin of ear and external auricular canal |
| ICD10CM | C44.221 | Squamous cell carcinoma of skin of unspecified ear and external auricular canal |
| ICD10CM | C44.222 | Squamous cell carcinoma of skin of right ear and external auricular canal |
| ICD10CM | C44.229 | Squamous cell carcinoma of skin of left ear and external auricular canal |
| ICD10CM | C44.29 | Other specified malignant neoplasm of skin of ear and external auricular canal |
| ICD10CM | C44.291 | Other specified malignant neoplasm of skin of unspecified ear and external auricular canal |
| ICD10CM | C44.292 | Other specified malignant neoplasm of skin of right ear and external auricular canal |
| ICD10CM | C44.299 | Other specified malignant neoplasm of skin of left ear and external auricular canal |
| ICD10CM | C44.3 | Other and unspecified malignant neoplasm of skin of other and unspecified parts of face |
| ICD10CM | C44.30 | Unspecified malignant neoplasm of skin of other and unspecified parts of face |
| ICD10CM | C44.300 | Unspecified malignant neoplasm of skin of unspecified part of face |
| ICD10CM | C44.301 | Unspecified malignant neoplasm of skin of nose |
| ICD10CM | C44.309 | Unspecified malignant neoplasm of skin of other parts of face |
| ICD10CM | C44.31 | Basal cell carcinoma of skin of other and unspecified parts of face |
| ICD10CM | C44.310 | Basal cell carcinoma of skin of unspecified parts of face |
| ICD10CM | C44.311 | Basal cell carcinoma of skin of nose |
| ICD10CM | C44.319 | Basal cell carcinoma of skin of other parts of face |
| ICD10CM | C44.32 | Squamous cell carcinoma of skin of other and unspecified parts of face |
| ICD10CM | C44.320 | Squamous cell carcinoma of skin of unspecified parts of face |

Continued on next page

**Table 7 – continued from previous page**

| CodeSystem | ConceptCode | ConceptName |
|---|---|---|
| ICD10CM | C44.321 | Squamous cell carcinoma of skin of nose |
| ICD10CM | C44.329 | Squamous cell carcinoma of skin of other parts of face |
| ICD10CM | C44.39 | Other specified malignant neoplasm of skin of other and unspecified parts of face |
| ICD10CM | C44.390 | Other specified malignant neoplasm of skin of unspecified parts of face |
| ICD10CM | C44.391 | Other specified malignant neoplasm of skin of nose |
| ICD10CM | C44.399 | Other specified malignant neoplasm of skin of other parts of face |
| ICD10CM | C44.4 | Other and unspecified malignant neoplasm of skin of scalp and neck |
| ICD10CM | C44.40 | Unspecified malignant neoplasm of skin of scalp and neck |
| ICD10CM | C44.41 | Basal cell carcinoma of skin of scalp and neck |
| ICD10CM | C44.42 | Squamous cell carcinoma of skin of scalp and neck |
| ICD10CM | C44.49 | Other specified malignant neoplasm of skin of scalp and neck |
| ICD10CM | C44.5 | Other and unspecified malignant neoplasm of skin of trunk |
| ICD10CM | C44.50 | Unspecified malignant neoplasm of skin of trunk |
| ICD10CM | C44.500 | Unspecified malignant neoplasm of anal skin |
| ICD10CM | C44.501 | Unspecified malignant neoplasm of skin of breast |
| ICD10CM | C44.509 | Unspecified malignant neoplasm of skin of other part of trunk |
| ICD10CM | C44.51 | Basal cell carcinoma of skin of trunk |
| ICD10CM | C44.510 | Basal cell carcinoma of anal skin |
| ICD10CM | C44.511 | Basal cell carcinoma of skin of breast |
| ICD10CM | C44.519 | Basal cell carcinoma of skin of other part of trunk |
| ICD10CM | C44.52 | Squamous cell carcinoma of skin of trunk |
| ICD10CM | C44.520 | Squamous cell carcinoma of anal skin |
| ICD10CM | C44.521 | Squamous cell carcinoma of skin of breast |
| ICD10CM | C44.529 | Squamous cell carcinoma of skin of other part of trunk |
| ICD10CM | C44.59 | Other specified malignant neoplasm of skin of trunk |
| ICD10CM | C44.590 | Other specified malignant neoplasm of anal skin |
| ICD10CM | C44.591 | Other specified malignant neoplasm of skin of breast |
| ICD10CM | C44.599 | Other specified malignant neoplasm of skin of other part of trunk |
| ICD10CM | C44.6 | Other and unspecified malignant neoplasm of skin of upper limb, including shoulder |
| ICD10CM | C44.60 | Unspecified malignant neoplasm of skin of upper limb, including shoulder |
| ICD10CM | C44.601 | Unspecified malignant neoplasm of skin of unspecified upper limb, including shoulder |
| ICD10CM | C44.602 | Unspecified malignant neoplasm of skin of right upper limb, including shoulder |

Continued on next page

**Table 7 – continued from previous page**

| CodeSystem | ConceptCode | ConceptName |
|---|---|---|
| ICD10CM | C44.609 | Unspecified malignant neoplasm of skin of left upper limb, including shoulder |
| ICD10CM | C44.61 | Basal cell carcinoma of skin of upper limb, including shoulder |
| ICD10CM | C44.611 | Basal cell carcinoma of skin of unspecified upper limb, including shoulder |
| ICD10CM | C44.612 | Basal cell carcinoma of skin of right upper limb, including shoulder |
| ICD10CM | C44.619 | Basal cell carcinoma of skin of left upper limb, including shoulder |
| ICD10CM | C44.62 | Squamous cell carcinoma of skin of upper limb, including shoulder |
| ICD10CM | C44.621 | Squamous cell carcinoma of skin of unspecified upper limb, including shoulder |
| ICD10CM | C44.622 | Squamous cell carcinoma of skin of right upper limb, including shoulder |
| ICD10CM | C44.629 | Squamous cell carcinoma of skin of left upper limb, including shoulder |
| ICD10CM | C44.69 | Other specified malignant neoplasm of skin of upper limb, including shoulder |
| ICD10CM | C44.691 | Other specified malignant neoplasm of skin of unspecified upper limb, including shoulder |
| ICD10CM | C44.692 | Other specified malignant neoplasm of skin of right upper limb, including shoulder |
| ICD10CM | C44.699 | Other specified malignant neoplasm of skin of left upper limb, including shoulder |
| ICD10CM | C44.7 | Other and unspecified malignant neoplasm of skin of lower limb, including hip |
| ICD10CM | C44.70 | Unspecified malignant neoplasm of skin of lower limb, including hip |
| ICD10CM | C44.701 | Unspecified malignant neoplasm of skin of unspecified lower limb, including hip |
| ICD10CM | C44.702 | Unspecified malignant neoplasm of skin of right lower limb, including hip |
| ICD10CM | C44.709 | Unspecified malignant neoplasm of skin of left lower limb, including hip |
| ICD10CM | C44.71 | Basal cell carcinoma of skin of lower limb, including hip |
| ICD10CM | C44.711 | Basal cell carcinoma of skin of unspecified lower limb, including hip |
| ICD10CM | C44.712 | Basal cell carcinoma of skin of right lower limb, including hip |
| ICD10CM | C44.719 | Basal cell carcinoma of skin of left lower limb, including hip |

Continued on next page

**Table 7 – continued from previous page**

| CodeSystem | ConceptCode | ConceptName |
|---|---|---|
| ICD10CM | C44.72 | Squamous cell carcinoma of skin of lower limb, including hip |
| ICD10CM | C44.721 | Squamous cell carcinoma of skin of unspecified lower limb, including hip |
| ICD10CM | C44.722 | Squamous cell carcinoma of skin of right lower limb, including hip |
| ICD10CM | C44.729 | Squamous cell carcinoma of skin of left lower limb, including hip |
| ICD10CM | C44.79 | Other specified malignant neoplasm of skin of lower limb, including hip |
| ICD10CM | C44.791 | Other specified malignant neoplasm of skin of unspecified lower limb, including hip |
| ICD10CM | C44.792 | Other specified malignant neoplasm of skin of right lower limb, including hip |
| ICD10CM | C44.799 | Other specified malignant neoplasm of skin of left lower limb, including hip |
| ICD10CM | C44.8 | Other and unspecified malignant neoplasm of overlapping sites of skin |
| ICD10CM | C44.80 | Unspecified malignant neoplasm of overlapping sites of skin |
| ICD10CM | C44.81 | Basal cell carcinoma of overlapping sites of skin |
| ICD10CM | C44.82 | Squamous cell carcinoma of overlapping sites of skin |
| ICD10CM | C44.89 | Other specified malignant neoplasm of overlapping sites of skin |
| ICD10CM | C44.9 | Other and unspecified malignant neoplasm of skin, unspecified |
| ICD10CM | C44.90 | Unspecified malignant neoplasm of skin, unspecified |
| ICD10CM | C44.91 | Basal cell carcinoma of skin, unspecified |
| ICD10CM | C44.92 | Squamous cell carcinoma of skin, unspecified |
| ICD10CM | C44.99 | Other specified malignant neoplasm of skin, unspecified |
| ICD10CM | C45.0 | Mesothelioma of pleura |
| ICD10CM | C45.1 | Mesothelioma of peritoneum |
| ICD10CM | C45.2 | Mesothelioma of pericardium |
| ICD10CM | C45.7 | Mesothelioma of other sites |
| ICD10CM | C45.9 | Mesothelioma, unspecified |
| ICD10CM | C46.0 | Kaposi's sarcoma of skin |
| ICD10CM | C46.1 | Kaposi's sarcoma of soft tissue |
| ICD10CM | C46.2 | Kaposi's sarcoma of palate |
| ICD10CM | C46.3 | Kaposi's sarcoma of lymph nodes |
| ICD10CM | C46.4 | Kaposi's sarcoma of gastrointestinal sites |
| ICD10CM | C46.5 | Kaposi's sarcoma of lung |
| ICD10CM | C46.50 | Kaposi's sarcoma of unspecified lung |
| ICD10CM | C46.51 | Kaposi's sarcoma of right lung |
| ICD10CM | C46.52 | Kaposi's sarcoma of left lung |

Continued on next page

**Table 7 – continued from previous page**

| CodeSystem | ConceptCode | ConceptName |
|---|---|---|
| ICD10CM | C46.7 | Kaposi's sarcoma of other sites |
| ICD10CM | C46.9 | Kaposi's sarcoma, unspecified |
| ICD10CM | C47.0 | Malignant neoplasm of peripheral nerves of head, face and neck |
| ICD10CM | C47.1 | Malignant neoplasm of peripheral nerves of upper limb, including shoulder |
| ICD10CM | C47.10 | Malignant neoplasm of peripheral nerves of unspecified upper limb, including shoulder |
| ICD10CM | C47.11 | Malignant neoplasm of peripheral nerves of right upper limb, including shoulder |
| ICD10CM | C47.12 | Malignant neoplasm of peripheral nerves of left upper limb, including shoulder |
| ICD10CM | C47.2 | Malignant neoplasm of peripheral nerves of lower limb, including hip |
| ICD10CM | C47.20 | Malignant neoplasm of peripheral nerves of unspecified lower limb, including hip |
| ICD10CM | C47.21 | Malignant neoplasm of peripheral nerves of right lower limb, including hip |
| ICD10CM | C47.22 | Malignant neoplasm of peripheral nerves of left lower limb, including hip |
| ICD10CM | C47.3 | Malignant neoplasm of peripheral nerves of thorax |
| ICD10CM | C47.4 | Malignant neoplasm of peripheral nerves of abdomen |
| ICD10CM | C47.5 | Malignant neoplasm of peripheral nerves of pelvis |
| ICD10CM | C47.6 | Malignant neoplasm of peripheral nerves of trunk, unspecified |
| ICD10CM | C47.8 | Malignant neoplasm of overlapping sites of peripheral nerves and autonomic nervous system |
| ICD10CM | C47.9 | Malignant neoplasm of peripheral nerves and autonomic nervous system, unspecified |
| ICD10CM | C48.0 | Malignant neoplasm of retroperitoneum |
| ICD10CM | C48.1 | Malignant neoplasm of specified parts of peritoneum |
| ICD10CM | C48.2 | Malignant neoplasm of peritoneum, unspecified |
| ICD10CM | C48.8 | Malignant neoplasm of overlapping sites of retroperitoneum and peritoneum |
| ICD10CM | C49.0 | Malignant neoplasm of connective and soft tissue of head, face and neck |
| ICD10CM | C49.1 | Malignant neoplasm of connective and soft tissue of upper limb, including shoulder |
| ICD10CM | C49.10 | Malignant neoplasm of connective and soft tissue of unspecified upper limb, including shoulder |
| ICD10CM | C49.11 | Malignant neoplasm of connective and soft tissue of right upper limb, including shoulder |

Continued on next page

**Table 7 – continued from previous page**

| CodeSystem | ConceptCode | ConceptName |
|---|---|---|
| ICD10CM | C49.12 | Malignant neoplasm of connective and soft tissue of left upper limb, including shoulder |
| ICD10CM | C49.2 | Malignant neoplasm of connective and soft tissue of lower limb, including hip |
| ICD10CM | C49.20 | Malignant neoplasm of connective and soft tissue of unspecified lower limb, including hip |
| ICD10CM | C49.21 | Malignant neoplasm of connective and soft tissue of right lower limb, including hip |
| ICD10CM | C49.22 | Malignant neoplasm of connective and soft tissue of left lower limb, including hip |
| ICD10CM | C49.3 | Malignant neoplasm of connective and soft tissue of thorax |
| ICD10CM | C49.4 | Malignant neoplasm of connective and soft tissue of abdomen |
| ICD10CM | C49.5 | Malignant neoplasm of connective and soft tissue of pelvis |
| ICD10CM | C49.6 | Malignant neoplasm of connective and soft tissue of trunk, unspecified |
| ICD10CM | C49.8 | Malignant neoplasm of overlapping sites of connective and soft tissue |
| ICD10CM | C49.9 | Malignant neoplasm of connective and soft tissue, unspecified |
| ICD10CM | C49.A | Gastrointestinal stromal tumor |
| ICD10CM | C49.A0 | Gastrointestinal stromal tumor, unspecified site |
| ICD10CM | C49.A1 | Gastrointestinal stromal tumor of esophagus |
| ICD10CM | C49.A2 | Gastrointestinal stromal tumor of stomach |
| ICD10CM | C49.A3 | Gastrointestinal stromal tumor of small intestine |
| ICD10CM | C49.A4 | Gastrointestinal stromal tumor of large intestine |
| ICD10CM | C49.A5 | Gastrointestinal stromal tumor of rectum |
| ICD10CM | C49.A9 | Gastrointestinal stromal tumor of other sites |
| ICD10CM | C50.0 | Malignant neoplasm of nipple and areola |
| ICD10CM | C50.01 | Malignant neoplasm of nipple and areola, female |
| ICD10CM | C50.011 | Malignant neoplasm of nipple and areola, right female breast |
| ICD10CM | C50.012 | Malignant neoplasm of nipple and areola, left female breast |
| ICD10CM | C50.019 | Malignant neoplasm of nipple and areola, unspecified female breast |
| ICD10CM | C50.02 | Malignant neoplasm of nipple and areola, male |
| ICD10CM | C50.021 | Malignant neoplasm of nipple and areola, right male breast |
| ICD10CM | C50.022 | Malignant neoplasm of nipple and areola, left male breast |
| ICD10CM | C50.029 | Malignant neoplasm of nipple and areola, unspecified male breast |
| ICD10CM | C50.1 | Malignant neoplasm of central portion of breast |
| ICD10CM | C50.11 | Malignant neoplasm of central portion of breast, female |
| ICD10CM | C50.111 | Malignant neoplasm of central portion of right female breast |

Continued on next page

**Table 7 – continued from previous page**

| CodeSystem | ConceptCode | ConceptName |
|---|---|---|
| ICD10CM | C50.112 | Malignant neoplasm of central portion of left female breast |
| ICD10CM | C50.119 | Malignant neoplasm of central portion of unspecified female breast |
| ICD10CM | C50.12 | Malignant neoplasm of central portion of breast, male |
| ICD10CM | C50.121 | Malignant neoplasm of central portion of right male breast |
| ICD10CM | C50.122 | Malignant neoplasm of central portion of left male breast |
| ICD10CM | C50.129 | Malignant neoplasm of central portion of unspecified male breast |
| ICD10CM | C50.2 | Malignant neoplasm of upper-inner quadrant of breast |
| ICD10CM | C50.21 | Malignant neoplasm of upper-inner quadrant of breast, female |
| ICD10CM | C50.211 | Malignant neoplasm of upper-inner quadrant of right female breast |
| ICD10CM | C50.212 | Malignant neoplasm of upper-inner quadrant of left female breast |
| ICD10CM | C50.219 | Malignant neoplasm of upper-inner quadrant of unspecified female breast |
| ICD10CM | C50.22 | Malignant neoplasm of upper-inner quadrant of breast, male |
| ICD10CM | C50.221 | Malignant neoplasm of upper-inner quadrant of right male breast |
| ICD10CM | C50.222 | Malignant neoplasm of upper-inner quadrant of left male breast |
| ICD10CM | C50.229 | Malignant neoplasm of upper-inner quadrant of unspecified male breast |
| ICD10CM | C50.3 | Malignant neoplasm of lower-inner quadrant of breast |
| ICD10CM | C50.31 | Malignant neoplasm of lower-inner quadrant of breast, female |
| ICD10CM | C50.311 | Malignant neoplasm of lower-inner quadrant of right female breast |
| ICD10CM | C50.312 | Malignant neoplasm of lower-inner quadrant of left female breast |
| ICD10CM | C50.319 | Malignant neoplasm of lower-inner quadrant of unspecified female breast |
| ICD10CM | C50.32 | Malignant neoplasm of lower-inner quadrant of breast, male |
| ICD10CM | C50.321 | Malignant neoplasm of lower-inner quadrant of right male breast |
| ICD10CM | C50.322 | Malignant neoplasm of lower-inner quadrant of left male breast |
| ICD10CM | C50.329 | Malignant neoplasm of lower-inner quadrant of unspecified male breast |
| ICD10CM | C50.4 | Malignant neoplasm of upper-outer quadrant of breast |
| ICD10CM | C50.41 | Malignant neoplasm of upper-outer quadrant of breast, female |

Continued on next page

**Table 7 – continued from previous page**

| CodeSystem | ConceptCode | ConceptName |
|---|---|---|
| ICD10CM | C50.411 | Malignant neoplasm of upper-outer quadrant of right female breast |
| ICD10CM | C50.412 | Malignant neoplasm of upper-outer quadrant of left female breast |
| ICD10CM | C50.419 | Malignant neoplasm of upper-outer quadrant of unspecified female breast |
| ICD10CM | C50.42 | Malignant neoplasm of upper-outer quadrant of breast, male |
| ICD10CM | C50.421 | Malignant neoplasm of upper-outer quadrant of right male breast |
| ICD10CM | C50.422 | Malignant neoplasm of upper-outer quadrant of left male breast |
| ICD10CM | C50.429 | Malignant neoplasm of upper-outer quadrant of unspecified male breast |
| ICD10CM | C50.5 | Malignant neoplasm of lower-outer quadrant of breast |
| ICD10CM | C50.51 | Malignant neoplasm of lower-outer quadrant of breast, female |
| ICD10CM | C50.511 | Malignant neoplasm of lower-outer quadrant of right female breast |
| ICD10CM | C50.512 | Malignant neoplasm of lower-outer quadrant of left female breast |
| ICD10CM | C50.519 | Malignant neoplasm of lower-outer quadrant of unspecified female breast |
| ICD10CM | C50.52 | Malignant neoplasm of lower-outer quadrant of breast, male |
| ICD10CM | C50.521 | Malignant neoplasm of lower-outer quadrant of right male breast |
| ICD10CM | C50.522 | Malignant neoplasm of lower-outer quadrant of left male breast |
| ICD10CM | C50.529 | Malignant neoplasm of lower-outer quadrant of unspecified male breast |
| ICD10CM | C50.6 | Malignant neoplasm of axillary tail of breast |
| ICD10CM | C50.61 | Malignant neoplasm of axillary tail of breast, female |
| ICD10CM | C50.611 | Malignant neoplasm of axillary tail of right female breast |
| ICD10CM | C50.612 | Malignant neoplasm of axillary tail of left female breast |
| ICD10CM | C50.619 | Malignant neoplasm of axillary tail of unspecified female breast |
| ICD10CM | C50.62 | Malignant neoplasm of axillary tail of breast, male |
| ICD10CM | C50.621 | Malignant neoplasm of axillary tail of right male breast |
| ICD10CM | C50.622 | Malignant neoplasm of axillary tail of left male breast |
| ICD10CM | C50.629 | Malignant neoplasm of axillary tail of unspecified male breast |
| ICD10CM | C50.8 | Malignant neoplasm of overlapping sites of breast |
| ICD10CM | C50.81 | Malignant neoplasm of overlapping sites of breast, female |

Continued on next page

**Table 7 – continued from previous page**

| CodeSystem | ConceptCode | ConceptName |
|---|---|---|
| ICD10CM | C50.811 | Malignant neoplasm of overlapping sites of right female breast |
| ICD10CM | C50.812 | Malignant neoplasm of overlapping sites of left female breast |
| ICD10CM | C50.819 | Malignant neoplasm of overlapping sites of unspecified female breast |
| ICD10CM | C50.82 | Malignant neoplasm of overlapping sites of breast, male |
| ICD10CM | C50.821 | Malignant neoplasm of overlapping sites of right male breast |
| ICD10CM | C50.822 | Malignant neoplasm of overlapping sites of left male breast |
| ICD10CM | C50.829 | Malignant neoplasm of overlapping sites of unspecified male breast |
| ICD10CM | C50.9 | Malignant neoplasm of breast of unspecified site |
| ICD10CM | C50.91 | Malignant neoplasm of breast of unspecified site, female |
| ICD10CM | C50.911 | Malignant neoplasm of unspecified site of right female breast |
| ICD10CM | C50.912 | Malignant neoplasm of unspecified site of left female breast |
| ICD10CM | C50.919 | Malignant neoplasm of unspecified site of unspecified female breast |
| ICD10CM | C50.92 | Malignant neoplasm of breast of unspecified site, male |
| ICD10CM | C50.921 | Malignant neoplasm of unspecified site of right male breast |
| ICD10CM | C50.922 | Malignant neoplasm of unspecified site of left male breast |
| ICD10CM | C50.929 | Malignant neoplasm of unspecified site of unspecified male breast |
| ICD10CM | C51.0 | Malignant neoplasm of labium majus |
| ICD10CM | C51.1 | Malignant neoplasm of labium minus |
| ICD10CM | C51.2 | Malignant neoplasm of clitoris |
| ICD10CM | C51.8 | Malignant neoplasm of overlapping sites of vulva |
| ICD10CM | C51.9 | Malignant neoplasm of vulva, unspecified |
| ICD10CM | C53.0 | Malignant neoplasm of endocervix |
| ICD10CM | C53.1 | Malignant neoplasm of exocervix |
| ICD10CM | C53.8 | Malignant neoplasm of overlapping sites of cervix uteri |
| ICD10CM | C53.9 | Malignant neoplasm of cervix uteri, unspecified |
| ICD10CM | C54.0 | Malignant neoplasm of isthmus uteri |
| ICD10CM | C54.1 | Malignant neoplasm of endometrium |
| ICD10CM | C54.2 | Malignant neoplasm of myometrium |
| ICD10CM | C54.3 | Malignant neoplasm of fundus uteri |
| ICD10CM | C54.8 | Malignant neoplasm of overlapping sites of corpus uteri |
| ICD10CM | C54.9 | Malignant neoplasm of corpus uteri, unspecified |
| ICD10CM | C56.1 | Malignant neoplasm of right ovary |
| ICD10CM | C56.2 | Malignant neoplasm of left ovary |
| ICD10CM | C56.9 | Malignant neoplasm of unspecified ovary |
| ICD10CM | C57.0 | Malignant neoplasm of fallopian tube |
| ICD10CM | C57.00 | Malignant neoplasm of unspecified fallopian tube |
| ICD10CM | C57.01 | Malignant neoplasm of right fallopian tube |
| ICD10CM | C57.02 | Malignant neoplasm of left fallopian tube |

Continued on next page

**Table 7 – continued from previous page**

| CodeSystem | ConceptCode | ConceptName |
|---|---|---|
| ICD10CM | C57.1 | Malignant neoplasm of broad ligament |
| ICD10CM | C57.10 | Malignant neoplasm of unspecified broad ligament |
| ICD10CM | C57.11 | Malignant neoplasm of right broad ligament |
| ICD10CM | C57.12 | Malignant neoplasm of left broad ligament |
| ICD10CM | C57.2 | Malignant neoplasm of round ligament |
| ICD10CM | C57.20 | Malignant neoplasm of unspecified round ligament |
| ICD10CM | C57.21 | Malignant neoplasm of right round ligament |
| ICD10CM | C57.22 | Malignant neoplasm of left round ligament |
| ICD10CM | C57.3 | Malignant neoplasm of parametrium |
| ICD10CM | C57.4 | Malignant neoplasm of uterine adnexa, unspecified |
| ICD10CM | C57.7 | Malignant neoplasm of other specified female genital organs |
| ICD10CM | C57.8 | Malignant neoplasm of overlapping sites of female genital organs |
| ICD10CM | C57.9 | Malignant neoplasm of female genital organ, unspecified |
| ICD10CM | C60.0 | Malignant neoplasm of prepuce |
| ICD10CM | C60.1 | Malignant neoplasm of glans penis |
| ICD10CM | C60.2 | Malignant neoplasm of body of penis |
| ICD10CM | C60.8 | Malignant neoplasm of overlapping sites of penis |
| ICD10CM | C60.9 | Malignant neoplasm of penis, unspecified |
| ICD10CM | C62.0 | Malignant neoplasm of undescended testis |
| ICD10CM | C62.00 | Malignant neoplasm of unspecified undescended testis |
| ICD10CM | C62.01 | Malignant neoplasm of undescended right testis |
| ICD10CM | C62.02 | Malignant neoplasm of undescended left testis |
| ICD10CM | C62.1 | Malignant neoplasm of descended testis |
| ICD10CM | C62.10 | Malignant neoplasm of unspecified descended testis |
| ICD10CM | C62.11 | Malignant neoplasm of descended right testis |
| ICD10CM | C62.12 | Malignant neoplasm of descended left testis |
| ICD10CM | C62.9 | Malignant neoplasm of testis, unspecified whether descended or undescended |
| ICD10CM | C62.90 | Malignant neoplasm of unspecified testis, unspecified whether descended or undescended |
| ICD10CM | C62.91 | Malignant neoplasm of right testis, unspecified whether descended or undescended |
| ICD10CM | C62.92 | Malignant neoplasm of left testis, unspecified whether descended or undescended |
| ICD10CM | C63.0 | Malignant neoplasm of epididymis |
| ICD10CM | C63.00 | Malignant neoplasm of unspecified epididymis |
| ICD10CM | C63.01 | Malignant neoplasm of right epididymis |
| ICD10CM | C63.02 | Malignant neoplasm of left epididymis |
| ICD10CM | C63.1 | Malignant neoplasm of spermatic cord |
| ICD10CM | C63.10 | Malignant neoplasm of unspecified spermatic cord |
| ICD10CM | C63.11 | Malignant neoplasm of right spermatic cord |
| ICD10CM | C63.12 | Malignant neoplasm of left spermatic cord |

Continued on next page

**Table 7 – continued from previous page**

| CodeSystem | ConceptCode | ConceptName |
|---|---|---|
| ICD10CM | C63.2 | Malignant neoplasm of scrotum |
| ICD10CM | C63.7 | Malignant neoplasm of other specified male genital organs |
| ICD10CM | C63.8 | Malignant neoplasm of overlapping sites of male genital organs |
| ICD10CM | C63.9 | Malignant neoplasm of male genital organ, unspecified |
| ICD10CM | C64.1 | Malignant neoplasm of right kidney, except renal pelvis |
| ICD10CM | C64.2 | Malignant neoplasm of left kidney, except renal pelvis |
| ICD10CM | C64.9 | Malignant neoplasm of unspecified kidney, except renal pelvis |
| ICD10CM | C65.1 | Malignant neoplasm of right renal pelvis |
| ICD10CM | C65.2 | Malignant neoplasm of left renal pelvis |
| ICD10CM | C65.9 | Malignant neoplasm of unspecified renal pelvis |
| ICD10CM | C66.1 | Malignant neoplasm of right ureter |
| ICD10CM | C66.2 | Malignant neoplasm of left ureter |
| ICD10CM | C66.9 | Malignant neoplasm of unspecified ureter |
| ICD10CM | C67.0 | Malignant neoplasm of trigone of bladder |
| ICD10CM | C67.1 | Malignant neoplasm of dome of bladder |
| ICD10CM | C67.2 | Malignant neoplasm of lateral wall of bladder |
| ICD10CM | C67.3 | Malignant neoplasm of anterior wall of bladder |
| ICD10CM | C67.4 | Malignant neoplasm of posterior wall of bladder |
| ICD10CM | C67.5 | Malignant neoplasm of bladder neck |
| ICD10CM | C67.6 | Malignant neoplasm of ureteric orifice |
| ICD10CM | C67.7 | Malignant neoplasm of urachus |
| ICD10CM | C67.8 | Malignant neoplasm of overlapping sites of bladder |
| ICD10CM | C67.9 | Malignant neoplasm of bladder, unspecified |
| ICD10CM | C68.0 | Malignant neoplasm of urethra |
| ICD10CM | C68.1 | Malignant neoplasm of paraurethral glands |
| ICD10CM | C68.8 | Malignant neoplasm of overlapping sites of urinary organs |
| ICD10CM | C68.9 | Malignant neoplasm of urinary organ, unspecified |
| ICD10CM | C69.0 | Malignant neoplasm of conjunctiva |
| ICD10CM | C69.00 | Malignant neoplasm of unspecified conjunctiva |
| ICD10CM | C69.01 | Malignant neoplasm of right conjunctiva |
| ICD10CM | C69.02 | Malignant neoplasm of left conjunctiva |
| ICD10CM | C69.1 | Malignant neoplasm of cornea |
| ICD10CM | C69.10 | Malignant neoplasm of unspecified cornea |
| ICD10CM | C69.11 | Malignant neoplasm of right cornea |
| ICD10CM | C69.12 | Malignant neoplasm of left cornea |
| ICD10CM | C69.2 | Malignant neoplasm of retina |
| ICD10CM | C69.20 | Malignant neoplasm of unspecified retina |
| ICD10CM | C69.21 | Malignant neoplasm of right retina |
| ICD10CM | C69.22 | Malignant neoplasm of left retina |
| ICD10CM | C69.3 | Malignant neoplasm of choroid |
| ICD10CM | C69.30 | Malignant neoplasm of unspecified choroid |

Continued on next page

**Table 7 – continued from previous page**

| CodeSystem | ConceptCode | ConceptName |
|---|---|---|
| ICD10CM | C69.31 | Malignant neoplasm of right choroid |
| ICD10CM | C69.32 | Malignant neoplasm of left choroid |
| ICD10CM | C69.4 | Malignant neoplasm of ciliary body |
| ICD10CM | C69.40 | Malignant neoplasm of unspecified ciliary body |
| ICD10CM | C69.41 | Malignant neoplasm of right ciliary body |
| ICD10CM | C69.42 | Malignant neoplasm of left ciliary body |
| ICD10CM | C69.5 | Malignant neoplasm of lacrimal gland and duct |
| ICD10CM | C69.50 | Malignant neoplasm of unspecified lacrimal gland and duct |
| ICD10CM | C69.51 | Malignant neoplasm of right lacrimal gland and duct |
| ICD10CM | C69.52 | Malignant neoplasm of left lacrimal gland and duct |
| ICD10CM | C69.6 | Malignant neoplasm of orbit |
| ICD10CM | C69.60 | Malignant neoplasm of unspecified orbit |
| ICD10CM | C69.61 | Malignant neoplasm of right orbit |
| ICD10CM | C69.62 | Malignant neoplasm of left orbit |
| ICD10CM | C69.8 | Malignant neoplasm of overlapping sites of eye and adnexa |
| ICD10CM | C69.80 | Malignant neoplasm of overlapping sites of unspecified eye and adnexa |
| ICD10CM | C69.81 | Malignant neoplasm of overlapping sites of right eye and adnexa |
| ICD10CM | C69.82 | Malignant neoplasm of overlapping sites of left eye and adnexa |
| ICD10CM | C69.9 | Malignant neoplasm of unspecified site of eye |
| ICD10CM | C69.90 | Malignant neoplasm of unspecified site of unspecified eye |
| ICD10CM | C69.91 | Malignant neoplasm of unspecified site of right eye |
| ICD10CM | C69.92 | Malignant neoplasm of unspecified site of left eye |
| ICD10CM | C70.0 | Malignant neoplasm of cerebral meninges |
| ICD10CM | C70.1 | Malignant neoplasm of spinal meninges |
| ICD10CM | C70.9 | Malignant neoplasm of meninges, unspecified |
| ICD10CM | C71.0 | Malignant neoplasm of cerebrum, except lobes and ventricles |
| ICD10CM | C71.1 | Malignant neoplasm of frontal lobe |
| ICD10CM | C71.2 | Malignant neoplasm of temporal lobe |
| ICD10CM | C71.3 | Malignant neoplasm of parietal lobe |
| ICD10CM | C71.4 | Malignant neoplasm of occipital lobe |
| ICD10CM | C71.5 | Malignant neoplasm of cerebral ventricle |
| ICD10CM | C71.6 | Malignant neoplasm of cerebellum |
| ICD10CM | C71.7 | Malignant neoplasm of brain stem |
| ICD10CM | C71.8 | Malignant neoplasm of overlapping sites of brain |
| ICD10CM | C71.9 | Malignant neoplasm of brain, unspecified |
| ICD10CM | C72.0 | Malignant neoplasm of spinal cord |
| ICD10CM | C72.1 | Malignant neoplasm of cauda equina |
| ICD10CM | C72.2 | Malignant neoplasm of olfactory nerve |
| ICD10CM | C72.20 | Malignant neoplasm of unspecified olfactory nerve |

Continued on next page

**Table 7 – continued from previous page**

| CodeSystem | ConceptCode | ConceptName |
|---|---|---|
| ICD10CM | C72.21 | Malignant neoplasm of right olfactory nerve |
| ICD10CM | C72.22 | Malignant neoplasm of left olfactory nerve |
| ICD10CM | C72.3 | Malignant neoplasm of optic nerve |
| ICD10CM | C72.30 | Malignant neoplasm of unspecified optic nerve |
| ICD10CM | C72.31 | Malignant neoplasm of right optic nerve |
| ICD10CM | C72.32 | Malignant neoplasm of left optic nerve |
| ICD10CM | C72.4 | Malignant neoplasm of acoustic nerve |
| ICD10CM | C72.40 | Malignant neoplasm of unspecified acoustic nerve |
| ICD10CM | C72.41 | Malignant neoplasm of right acoustic nerve |
| ICD10CM | C72.42 | Malignant neoplasm of left acoustic nerve |
| ICD10CM | C72.5 | Malignant neoplasm of other and unspecified cranial nerves |
| ICD10CM | C72.50 | Malignant neoplasm of unspecified cranial nerve |
| ICD10CM | C72.59 | Malignant neoplasm of other cranial nerves |
| ICD10CM | C72.9 | Malignant neoplasm of central nervous system, unspecified |
| ICD10CM | C74.0 | Malignant neoplasm of cortex of adrenal gland |
| ICD10CM | C74.00 | Malignant neoplasm of cortex of unspecified adrenal gland |
| ICD10CM | C74.01 | Malignant neoplasm of cortex of right adrenal gland |
| ICD10CM | C74.02 | Malignant neoplasm of cortex of left adrenal gland |
| ICD10CM | C74.1 | Malignant neoplasm of medulla of adrenal gland |
| ICD10CM | C74.10 | Malignant neoplasm of medulla of unspecified adrenal gland |
| ICD10CM | C74.11 | Malignant neoplasm of medulla of right adrenal gland |
| ICD10CM | C74.12 | Malignant neoplasm of medulla of left adrenal gland |
| ICD10CM | C74.9 | Malignant neoplasm of unspecified part of adrenal gland |
| ICD10CM | C74.90 | Malignant neoplasm of unspecified part of unspecified adrenal gland |
| ICD10CM | C74.91 | Malignant neoplasm of unspecified part of right adrenal gland |
| ICD10CM | C74.92 | Malignant neoplasm of unspecified part of left adrenal gland |
| ICD10CM | C75.0 | Malignant neoplasm of parathyroid gland |
| ICD10CM | C75.1 | Malignant neoplasm of pituitary gland |
| ICD10CM | C75.2 | Malignant neoplasm of craniopharyngeal duct |
| ICD10CM | C75.3 | Malignant neoplasm of pineal gland |
| ICD10CM | C75.4 | Malignant neoplasm of carotid body |
| ICD10CM | C75.5 | Malignant neoplasm of aortic body and other paraganglia |
| ICD10CM | C75.8 | Malignant neoplasm with pluriglandular involvement, unspecified |
| ICD10CM | C75.9 | Malignant neoplasm of endocrine gland, unspecified |
| ICD10CM | C76.0 | Malignant neoplasm of head, face and neck |
| ICD10CM | C76.1 | Malignant neoplasm of thorax |
| ICD10CM | C76.2 | Malignant neoplasm of abdomen |
| ICD10CM | C76.3 | Malignant neoplasm of pelvis |
| ICD10CM | C76.4 | Malignant neoplasm of upper limb |
| ICD10CM | C76.40 | Malignant neoplasm of unspecified upper limb |

Continued on next page

**Table 7 – continued from previous page**

| CodeSystem | ConceptCode | ConceptName |
|---|---|---|
| ICD10CM | C76.41 | Malignant neoplasm of right upper limb |
| ICD10CM | C76.42 | Malignant neoplasm of left upper limb |
| ICD10CM | C76.5 | Malignant neoplasm of lower limb |
| ICD10CM | C76.50 | Malignant neoplasm of unspecified lower limb |
| ICD10CM | C76.51 | Malignant neoplasm of right lower limb |
| ICD10CM | C76.52 | Malignant neoplasm of left lower limb |
| ICD10CM | C76.8 | Malignant neoplasm of other specified ill-defined sites |
| ICD10CM | C77.0 | Secondary and unspecified malignant neoplasm of lymph nodes of head, face and neck |
| ICD10CM | C77.1 | Secondary and unspecified malignant neoplasm of intrathoracic lymph nodes |
| ICD10CM | C77.2 | Secondary and unspecified malignant neoplasm of intra-abdominal lymph nodes |
| ICD10CM | C77.3 | Secondary and unspecified malignant neoplasm of axilla and upper limb lymph nodes |
| ICD10CM | C77.4 | Secondary and unspecified malignant neoplasm of inguinal and lower limb lymph nodes |
| ICD10CM | C77.5 | Secondary and unspecified malignant neoplasm of intrapelvic lymph nodes |
| ICD10CM | C77.8 | Secondary and unspecified malignant neoplasm of lymph nodes of multiple regions |
| ICD10CM | C77.9 | Secondary and unspecified malignant neoplasm of lymph node, unspecified |
| ICD10CM | C78.0 | Secondary malignant neoplasm of lung |
| ICD10CM | C78.00 | Secondary malignant neoplasm of unspecified lung |
| ICD10CM | C78.01 | Secondary malignant neoplasm of right lung |
| ICD10CM | C78.02 | Secondary malignant neoplasm of left lung |
| ICD10CM | C78.1 | Secondary malignant neoplasm of mediastinum |
| ICD10CM | C78.2 | Secondary malignant neoplasm of pleura |
| ICD10CM | C78.3 | Secondary malignant neoplasm of other and unspecified respiratory organs |
| ICD10CM | C78.30 | Secondary malignant neoplasm of unspecified respiratory organ |
| ICD10CM | C78.39 | Secondary malignant neoplasm of other respiratory organs |
| ICD10CM | C78.4 | Secondary malignant neoplasm of small intestine |
| ICD10CM | C78.5 | Secondary malignant neoplasm of large intestine and rectum |
| ICD10CM | C78.6 | Secondary malignant neoplasm of retroperitoneum and peritoneum |
| ICD10CM | C78.7 | Secondary malignant neoplasm of liver and intrahepatic bile duct |
| ICD10CM | C78.8 | Secondary malignant neoplasm of other and unspecified digestive organs |

Continued on next page

**Table 7 – continued from previous page**

| CodeSystem | ConceptCode | ConceptName |
|---|---|---|
| ICD10CM | C78.80 | Secondary malignant neoplasm of unspecified digestive organ |
| ICD10CM | C78.89 | Secondary malignant neoplasm of other digestive organs |
| ICD10CM | C79.0 | Secondary malignant neoplasm of kidney and renal pelvis |
| ICD10CM | C79.00 | Secondary malignant neoplasm of unspecified kidney and renal pelvis |
| ICD10CM | C79.01 | Secondary malignant neoplasm of right kidney and renal pelvis |
| ICD10CM | C79.02 | Secondary malignant neoplasm of left kidney and renal pelvis |
| ICD10CM | C79.1 | Secondary malignant neoplasm of bladder and other and unspecified urinary organs |
| ICD10CM | C79.10 | Secondary malignant neoplasm of unspecified urinary organs |
| ICD10CM | C79.11 | Secondary malignant neoplasm of bladder |
| ICD10CM | C79.19 | Secondary malignant neoplasm of other urinary organs |
| ICD10CM | C79.2 | Secondary malignant neoplasm of skin |
| ICD10CM | C79.3 | Secondary malignant neoplasm of brain and cerebral meninges |
| ICD10CM | C79.31 | Secondary malignant neoplasm of brain |
| ICD10CM | C79.32 | Secondary malignant neoplasm of cerebral meninges |
| ICD10CM | C79.4 | Secondary malignant neoplasm of other and unspecified parts of nervous system |
| ICD10CM | C79.40 | Secondary malignant neoplasm of unspecified part of nervous system |
| ICD10CM | C79.49 | Secondary malignant neoplasm of other parts of nervous system |
| ICD10CM | C79.5 | Secondary malignant neoplasm of bone and bone marrow |
| ICD10CM | C79.51 | Secondary malignant neoplasm of bone |
| ICD10CM | C79.52 | Secondary malignant neoplasm of bone marrow |
| ICD10CM | C79.6 | Secondary malignant neoplasm of ovary |
| ICD10CM | C79.60 | Secondary malignant neoplasm of unspecified ovary |
| ICD10CM | C79.61 | Secondary malignant neoplasm of right ovary |
| ICD10CM | C79.62 | Secondary malignant neoplasm of left ovary |
| ICD10CM | C79.7 | Secondary malignant neoplasm of adrenal gland |
| ICD10CM | C79.70 | Secondary malignant neoplasm of unspecified adrenal gland |
| ICD10CM | C79.71 | Secondary malignant neoplasm of right adrenal gland |
| ICD10CM | C79.72 | Secondary malignant neoplasm of left adrenal gland |
| ICD10CM | C79.8 | Secondary malignant neoplasm of other specified sites |
| ICD10CM | C79.81 | Secondary malignant neoplasm of breast |
| ICD10CM | C79.82 | Secondary malignant neoplasm of genital organs |
| ICD10CM | C79.89 | Secondary malignant neoplasm of other specified sites |
| ICD10CM | C79.9 | Secondary malignant neoplasm of unspecified site |

Continued on next page

**Table 7 – continued from previous page**

| CodeSystem | ConceptCode | ConceptName |
|---|---|---|
| ICD10CM | C7A.0 | Malignant carcinoid tumors |
| ICD10CM | C7A.00 | Malignant carcinoid tumor of unspecified site |
| ICD10CM | C7A.01 | Malignant carcinoid tumors of the small intestine |
| ICD10CM | C7A.010 | Malignant carcinoid tumor of the duodenum |
| ICD10CM | C7A.011 | Malignant carcinoid tumor of the jejunum |
| ICD10CM | C7A.012 | Malignant carcinoid tumor of the ileum |
| ICD10CM | C7A.019 | Malignant carcinoid tumor of the small intestine, unspecified portion |
| ICD10CM | C7A.02 | Malignant carcinoid tumors of the appendix, large intestine, and rectum |
| ICD10CM | C7A.020 | Malignant carcinoid tumor of the appendix |
| ICD10CM | C7A.021 | Malignant carcinoid tumor of the cecum |
| ICD10CM | C7A.022 | Malignant carcinoid tumor of the ascending colon |
| ICD10CM | C7A.023 | Malignant carcinoid tumor of the transverse colon |
| ICD10CM | C7A.024 | Malignant carcinoid tumor of the descending colon |
| ICD10CM | C7A.025 | Malignant carcinoid tumor of the sigmoid colon |
| ICD10CM | C7A.026 | Malignant carcinoid tumor of the rectum |
| ICD10CM | C7A.029 | Malignant carcinoid tumor of the large intestine, unspecified portion |
| ICD10CM | C7A.09 | Malignant carcinoid tumors of other sites |
| ICD10CM | C7A.090 | Malignant carcinoid tumor of the bronchus and lung |
| ICD10CM | C7A.091 | Malignant carcinoid tumor of the thymus |
| ICD10CM | C7A.092 | Malignant carcinoid tumor of the stomach |
| ICD10CM | C7A.093 | Malignant carcinoid tumor of the kidney |
| ICD10CM | C7A.094 | Malignant carcinoid tumor of the foregut, unspecified |
| ICD10CM | C7A.095 | Malignant carcinoid tumor of the midgut, unspecified |
| ICD10CM | C7A.096 | Malignant carcinoid tumor of the hindgut, unspecified |
| ICD10CM | C7A.098 | Malignant carcinoid tumors of other sites |
| ICD10CM | C7A.1 | Malignant poorly differentiated neuroendocrine tumors |
| ICD10CM | C7A.8 | Other malignant neuroendocrine tumors |
| ICD10CM | C7B.0 | Secondary carcinoid tumors |
| ICD10CM | C7B.00 | Secondary carcinoid tumors, unspecified site |
| ICD10CM | C7B.01 | Secondary carcinoid tumors of distant lymph nodes |
| ICD10CM | C7B.02 | Secondary carcinoid tumors of liver |
| ICD10CM | C7B.03 | Secondary carcinoid tumors of bone |
| ICD10CM | C7B.04 | Secondary carcinoid tumors of peritoneum |
| ICD10CM | C7B.09 | Secondary carcinoid tumors of other sites |
| ICD10CM | C7B.1 | Secondary Merkel cell carcinoma |
| ICD10CM | C7B.8 | Other secondary neuroendocrine tumors |
| ICD10CM | C80.0 | Disseminated malignant neoplasm, unspecified |
| ICD10CM | C80.1 | Malignant (primary) neoplasm, unspecified |
| ICD10CM | C80.2 | Malignant neoplasm associated with transplanted organ |
| ICD10CM | C81.0 | Nodular lymphocyte predominant Hodgkin lymphoma |

Continued on next page

**Table 7 – continued from previous page**

| CodeSystem | ConceptCode | ConceptName |
|---|---|---|
| ICD10CM | C81.00 | Nodular lymphocyte predominant Hodgkin lymphoma, unspecified site |
| ICD10CM | C81.01 | Nodular lymphocyte predominant Hodgkin lymphoma, lymph nodes of head, face, and neck |
| ICD10CM | C81.02 | Nodular lymphocyte predominant Hodgkin lymphoma, intrathoracic lymph nodes |
| ICD10CM | C81.03 | Nodular lymphocyte predominant Hodgkin lymphoma, intra-abdominal lymph nodes |
| ICD10CM | C81.04 | Nodular lymphocyte predominant Hodgkin lymphoma, lymph nodes of axilla and upper limb |
| ICD10CM | C81.05 | Nodular lymphocyte predominant Hodgkin lymphoma, lymph nodes of inguinal region and lower limb |
| ICD10CM | C81.06 | Nodular lymphocyte predominant Hodgkin lymphoma, intrapelvic lymph nodes |
| ICD10CM | C81.07 | Nodular lymphocyte predominant Hodgkin lymphoma, spleen |
| ICD10CM | C81.08 | Nodular lymphocyte predominant Hodgkin lymphoma, lymph nodes of multiple sites |
| ICD10CM | C81.09 | Nodular lymphocyte predominant Hodgkin lymphoma, extranodal and solid organ sites |
| ICD10CM | C81.1 | Nodular sclerosis Hodgkin lymphoma |
| ICD10CM | C81.10 | Nodular sclerosis Hodgkin lymphoma, unspecified site |
| ICD10CM | C81.11 | Nodular sclerosis Hodgkin lymphoma, lymph nodes of head, face, and neck |
| ICD10CM | C81.12 | Nodular sclerosis Hodgkin lymphoma, intrathoracic lymph nodes |
| ICD10CM | C81.13 | Nodular sclerosis Hodgkin lymphoma, intra-abdominal lymph nodes |
| ICD10CM | C81.14 | Nodular sclerosis Hodgkin lymphoma, lymph nodes of axilla and upper limb |
| ICD10CM | C81.15 | Nodular sclerosis Hodgkin lymphoma, lymph nodes of inguinal region and lower limb |
| ICD10CM | C81.16 | Nodular sclerosis Hodgkin lymphoma, intrapelvic lymph nodes |
| ICD10CM | C81.17 | Nodular sclerosis Hodgkin lymphoma, spleen |
| ICD10CM | C81.18 | Nodular sclerosis Hodgkin lymphoma, lymph nodes of multiple sites |
| ICD10CM | C81.19 | Nodular sclerosis Hodgkin lymphoma, extranodal and solid organ sites |
| ICD10CM | C81.2 | Mixed cellularity Hodgkin lymphoma |
| ICD10CM | C81.20 | Mixed cellularity Hodgkin lymphoma, unspecified site |
| ICD10CM | C81.21 | Mixed cellularity Hodgkin lymphoma, lymph nodes of head, face, and neck |

Continued on next page

**Table 7 – continued from previous page**

| CodeSystem | ConceptCode | ConceptName |
|---|---|---|
| ICD10CM | C81.22 | Mixed cellularity Hodgkin lymphoma, intrathoracic lymph nodes |
| ICD10CM | C81.23 | Mixed cellularity Hodgkin lymphoma, intra-abdominal lymph nodes |
| ICD10CM | C81.24 | Mixed cellularity Hodgkin lymphoma, lymph nodes of axilla and upper limb |
| ICD10CM | C81.25 | Mixed cellularity Hodgkin lymphoma, lymph nodes of inguinal region and lower limb |
| ICD10CM | C81.26 | Mixed cellularity Hodgkin lymphoma, intrapelvic lymph nodes |
| ICD10CM | C81.27 | Mixed cellularity Hodgkin lymphoma, spleen |
| ICD10CM | C81.28 | Mixed cellularity Hodgkin lymphoma, lymph nodes of multiple sites |
| ICD10CM | C81.29 | Mixed cellularity Hodgkin lymphoma, extranodal and solid organ sites |
| ICD10CM | C81.3 | Lymphocyte depleted Hodgkin lymphoma |
| ICD10CM | C81.30 | Lymphocyte depleted Hodgkin lymphoma, unspecified site |
| ICD10CM | C81.31 | Lymphocyte depleted Hodgkin lymphoma, lymph nodes of head, face, and neck |
| ICD10CM | C81.32 | Lymphocyte depleted Hodgkin lymphoma, intrathoracic lymph nodes |
| ICD10CM | C81.33 | Lymphocyte depleted Hodgkin lymphoma, intra-abdominal lymph nodes |
| ICD10CM | C81.34 | Lymphocyte depleted Hodgkin lymphoma, lymph nodes of axilla and upper limb |
| ICD10CM | C81.35 | Lymphocyte depleted Hodgkin lymphoma, lymph nodes of inguinal region and lower limb |
| ICD10CM | C81.36 | Lymphocyte depleted Hodgkin lymphoma, intrapelvic lymph nodes |
| ICD10CM | C81.37 | Lymphocyte depleted Hodgkin lymphoma, spleen |
| ICD10CM | C81.38 | Lymphocyte depleted Hodgkin lymphoma, lymph nodes of multiple sites |
| ICD10CM | C81.39 | Lymphocyte depleted Hodgkin lymphoma, extranodal and solid organ sites |
| ICD10CM | C81.4 | Lymphocyte-rich Hodgkin lymphoma |
| ICD10CM | C81.40 | Lymphocyte-rich Hodgkin lymphoma, unspecified site |
| ICD10CM | C81.41 | Lymphocyte-rich Hodgkin lymphoma, lymph nodes of head, face, and neck |
| ICD10CM | C81.42 | Lymphocyte-rich Hodgkin lymphoma, intrathoracic lymph nodes |
| ICD10CM | C81.43 | Lymphocyte-rich Hodgkin lymphoma, intra-abdominal lymph nodes |

Continued on next page

**Table 7 – continued from previous page**

| CodeSystem | ConceptCode | ConceptName |
|---|---|---|
| ICD10CM | C81.44 | Lymphocyte-rich Hodgkin lymphoma, lymph nodes of axilla and upper limb |
| ICD10CM | C81.45 | Lymphocyte-rich Hodgkin lymphoma, lymph nodes of inguinal region and lower limb |
| ICD10CM | C81.46 | Lymphocyte-rich Hodgkin lymphoma, intrapelvic lymph nodes |
| ICD10CM | C81.47 | Lymphocyte-rich Hodgkin lymphoma, spleen |
| ICD10CM | C81.48 | Lymphocyte-rich Hodgkin lymphoma, lymph nodes of multiple sites |
| ICD10CM | C81.49 | Lymphocyte-rich Hodgkin lymphoma, extranodal and solid organ sites |
| ICD10CM | C81.7 | Other Hodgkin lymphoma |
| ICD10CM | C81.70 | Other Hodgkin lymphoma, unspecified site |
| ICD10CM | C81.71 | Other Hodgkin lymphoma, lymph nodes of head, face, and neck |
| ICD10CM | C81.72 | Other Hodgkin lymphoma, intrathoracic lymph nodes |
| ICD10CM | C81.73 | Other Hodgkin lymphoma, intra-abdominal lymph nodes |
| ICD10CM | C81.74 | Other Hodgkin lymphoma, lymph nodes of axilla and upper limb |
| ICD10CM | C81.75 | Other Hodgkin lymphoma, lymph nodes of inguinal region and lower limb |
| ICD10CM | C81.76 | Other Hodgkin lymphoma, intrapelvic lymph nodes |
| ICD10CM | C81.77 | Other Hodgkin lymphoma, spleen |
| ICD10CM | C81.78 | Other Hodgkin lymphoma, lymph nodes of multiple sites |
| ICD10CM | C81.79 | Other Hodgkin lymphoma, extranodal and solid organ sites |
| ICD10CM | C81.9 | Hodgkin lymphoma, unspecified |
| ICD10CM | C81.90 | Hodgkin lymphoma, unspecified, unspecified site |
| ICD10CM | C81.91 | Hodgkin lymphoma, unspecified, lymph nodes of head, face, and neck |
| ICD10CM | C81.92 | Hodgkin lymphoma, unspecified, intrathoracic lymph nodes |
| ICD10CM | C81.93 | Hodgkin lymphoma, unspecified, intra-abdominal lymph nodes |
| ICD10CM | C81.94 | Hodgkin lymphoma, unspecified, lymph nodes of axilla and upper limb |
| ICD10CM | C81.95 | Hodgkin lymphoma, unspecified, lymph nodes of inguinal region and lower limb |
| ICD10CM | C81.96 | Hodgkin lymphoma, unspecified, intrapelvic lymph nodes |
| ICD10CM | C81.97 | Hodgkin lymphoma, unspecified, spleen |
| ICD10CM | C81.98 | Hodgkin lymphoma, unspecified, lymph nodes of multiple sites |
| ICD10CM | C81.99 | Hodgkin lymphoma, unspecified, extranodal and solid organ sites |
| ICD10CM | C82.0 | Follicular lymphoma grade I |

Continued on next page

**Table 7 – continued from previous page**

| CodeSystem | ConceptCode | ConceptName |
|---|---|---|
| ICD10CM | C82.00 | Follicular lymphoma grade I, unspecified site |
| ICD10CM | C82.01 | Follicular lymphoma grade I, lymph nodes of head, face, and neck |
| ICD10CM | C82.02 | Follicular lymphoma grade I, intrathoracic lymph nodes |
| ICD10CM | C82.03 | Follicular lymphoma grade I, intra-abdominal lymph nodes |
| ICD10CM | C82.04 | Follicular lymphoma grade I, lymph nodes of axilla and upper limb |
| ICD10CM | C82.05 | Follicular lymphoma grade I, lymph nodes of inguinal region and lower limb |
| ICD10CM | C82.06 | Follicular lymphoma grade I, intrapelvic lymph nodes |
| ICD10CM | C82.07 | Follicular lymphoma grade I, spleen |
| ICD10CM | C82.08 | Follicular lymphoma grade I, lymph nodes of multiple sites |
| ICD10CM | C82.09 | Follicular lymphoma grade I, extranodal and solid organ sites |
| ICD10CM | C82.1 | Follicular lymphoma grade II |
| ICD10CM | C82.10 | Follicular lymphoma grade II, unspecified site |
| ICD10CM | C82.11 | Follicular lymphoma grade II, lymph nodes of head, face, and neck |
| ICD10CM | C82.12 | Follicular lymphoma grade II, intrathoracic lymph nodes |
| ICD10CM | C82.13 | Follicular lymphoma grade II, intra-abdominal lymph nodes |
| ICD10CM | C82.14 | Follicular lymphoma grade II, lymph nodes of axilla and upper limb |
| ICD10CM | C82.15 | Follicular lymphoma grade II, lymph nodes of inguinal region and lower limb |
| ICD10CM | C82.16 | Follicular lymphoma grade II, intrapelvic lymph nodes |
| ICD10CM | C82.17 | Follicular lymphoma grade II, spleen |
| ICD10CM | C82.18 | Follicular lymphoma grade II, lymph nodes of multiple sites |
| ICD10CM | C82.19 | Follicular lymphoma grade II, extranodal and solid organ sites |
| ICD10CM | C82.2 | Follicular lymphoma grade III, unspecified |
| ICD10CM | C82.20 | Follicular lymphoma grade III, unspecified, unspecified site |
| ICD10CM | C82.21 | Follicular lymphoma grade III, unspecified, lymph nodes of head, face, and neck |
| ICD10CM | C82.22 | Follicular lymphoma grade III, unspecified, intrathoracic lymph nodes |
| ICD10CM | C82.23 | Follicular lymphoma grade III, unspecified, intra-abdominal lymph nodes |
| ICD10CM | C82.24 | Follicular lymphoma grade III, unspecified, lymph nodes of axilla and upper limb |
| ICD10CM | C82.25 | Follicular lymphoma grade III, unspecified, lymph nodes of inguinal region and lower limb |
| ICD10CM | C82.26 | Follicular lymphoma grade III, unspecified, intrapelvic lymph nodes |

Continued on next page

**Table 7 – continued from previous page**

| CodeSystem | ConceptCode | ConceptName |
|---|---|---|
| ICD10CM | C82.27 | Follicular lymphoma grade III, unspecified, spleen |
| ICD10CM | C82.28 | Follicular lymphoma grade III, unspecified, lymph nodes of multiple sites |
| ICD10CM | C82.29 | Follicular lymphoma grade III, unspecified, extranodal and solid organ sites |
| ICD10CM | C82.3 | Follicular lymphoma grade IIIa |
| ICD10CM | C82.30 | Follicular lymphoma grade IIIa, unspecified site |
| ICD10CM | C82.31 | Follicular lymphoma grade IIIa, lymph nodes of head, face, and neck |
| ICD10CM | C82.32 | Follicular lymphoma grade IIIa, intrathoracic lymph nodes |
| ICD10CM | C82.33 | Follicular lymphoma grade IIIa, intra-abdominal lymph nodes |
| ICD10CM | C82.34 | Follicular lymphoma grade IIIa, lymph nodes of axilla and upper limb |
| ICD10CM | C82.35 | Follicular lymphoma grade IIIa, lymph nodes of inguinal region and lower limb |
| ICD10CM | C82.36 | Follicular lymphoma grade IIIa, intrapelvic lymph nodes |
| ICD10CM | C82.37 | Follicular lymphoma grade IIIa, spleen |
| ICD10CM | C82.38 | Follicular lymphoma grade IIIa, lymph nodes of multiple sites |
| ICD10CM | C82.39 | Follicular lymphoma grade IIIa, extranodal and solid organ sites |
| ICD10CM | C82.4 | Follicular lymphoma grade IIIb |
| ICD10CM | C82.40 | Follicular lymphoma grade IIIb, unspecified site |
| ICD10CM | C82.41 | Follicular lymphoma grade IIIb, lymph nodes of head, face, and neck |
| ICD10CM | C82.42 | Follicular lymphoma grade IIIb, intrathoracic lymph nodes |
| ICD10CM | C82.43 | Follicular lymphoma grade IIIb, intra-abdominal lymph nodes |
| ICD10CM | C82.44 | Follicular lymphoma grade IIIb, lymph nodes of axilla and upper limb |
| ICD10CM | C82.45 | Follicular lymphoma grade IIIb, lymph nodes of inguinal region and lower limb |
| ICD10CM | C82.46 | Follicular lymphoma grade IIIb, intrapelvic lymph nodes |
| ICD10CM | C82.47 | Follicular lymphoma grade IIIb, spleen |
| ICD10CM | C82.48 | Follicular lymphoma grade IIIb, lymph nodes of multiple sites |
| ICD10CM | C82.49 | Follicular lymphoma grade IIIb, extranodal and solid organ sites |
| ICD10CM | C82.5 | Diffuse follicle center lymphoma |
| ICD10CM | C82.50 | Diffuse follicle center lymphoma, unspecified site |
| ICD10CM | C82.51 | Diffuse follicle center lymphoma, lymph nodes of head, face, and neck |

Continued on next page

**Table 7 – continued from previous page**

| CodeSystem | ConceptCode | ConceptName |
|---|---|---|
| ICD10CM | C82.52 | Diffuse follicle center lymphoma, intrathoracic lymph nodes |
| ICD10CM | C82.53 | Diffuse follicle center lymphoma, intra-abdominal lymph nodes |
| ICD10CM | C82.54 | Diffuse follicle center lymphoma, lymph nodes of axilla and upper limb |
| ICD10CM | C82.55 | Diffuse follicle center lymphoma, lymph nodes of inguinal region and lower limb |
| ICD10CM | C82.56 | Diffuse follicle center lymphoma, intrapelvic lymph nodes |
| ICD10CM | C82.57 | Diffuse follicle center lymphoma, spleen |
| ICD10CM | C82.58 | Diffuse follicle center lymphoma, lymph nodes of multiple sites |
| ICD10CM | C82.59 | Diffuse follicle center lymphoma, extranodal and solid organ sites |
| ICD10CM | C82.6 | Cutaneous follicle center lymphoma |
| ICD10CM | C82.60 | Cutaneous follicle center lymphoma, unspecified site |
| ICD10CM | C82.61 | Cutaneous follicle center lymphoma, lymph nodes of head, face, and neck |
| ICD10CM | C82.62 | Cutaneous follicle center lymphoma, intrathoracic lymph nodes |
| ICD10CM | C82.63 | Cutaneous follicle center lymphoma, intra-abdominal lymph nodes |
| ICD10CM | C82.64 | Cutaneous follicle center lymphoma, lymph nodes of axilla and upper limb |
| ICD10CM | C82.65 | Cutaneous follicle center lymphoma, lymph nodes of inguinal region and lower limb |
| ICD10CM | C82.66 | Cutaneous follicle center lymphoma, intrapelvic lymph nodes |
| ICD10CM | C82.67 | Cutaneous follicle center lymphoma, spleen |
| ICD10CM | C82.68 | Cutaneous follicle center lymphoma, lymph nodes of multiple sites |
| ICD10CM | C82.69 | Cutaneous follicle center lymphoma, extranodal and solid organ sites |
| ICD10CM | C82.8 | Other types of follicular lymphoma |
| ICD10CM | C82.80 | Other types of follicular lymphoma, unspecified site |
| ICD10CM | C82.81 | Other types of follicular lymphoma, lymph nodes of head, face, and neck |
| ICD10CM | C82.82 | Other types of follicular lymphoma, intrathoracic lymph nodes |
| ICD10CM | C82.83 | Other types of follicular lymphoma, intra-abdominal lymph nodes |
| ICD10CM | C82.84 | Other types of follicular lymphoma, lymph nodes of axilla and upper limb |

Continued on next page

**Table 7 – continued from previous page**

| CodeSystem | ConceptCode | ConceptName |
|---|---|---|
| ICD10CM | C82.85 | Other types of follicular lymphoma, lymph nodes of inguinal region and lower limb |
| ICD10CM | C82.86 | Other types of follicular lymphoma, intrapelvic lymph nodes |
| ICD10CM | C82.87 | Other types of follicular lymphoma, spleen |
| ICD10CM | C82.88 | Other types of follicular lymphoma, lymph nodes of multiple sites |
| ICD10CM | C82.89 | Other types of follicular lymphoma, extranodal and solid organ sites |
| ICD10CM | C82.9 | Follicular lymphoma, unspecified |
| ICD10CM | C82.90 | Follicular lymphoma, unspecified, unspecified site |
| ICD10CM | C82.91 | Follicular lymphoma, unspecified, lymph nodes of head, face, and neck |
| ICD10CM | C82.92 | Follicular lymphoma, unspecified, intrathoracic lymph nodes |
| ICD10CM | C82.93 | Follicular lymphoma, unspecified, intra-abdominal lymph nodes |
| ICD10CM | C82.94 | Follicular lymphoma, unspecified, lymph nodes of axilla and upper limb |
| ICD10CM | C82.95 | Follicular lymphoma, unspecified, lymph nodes of inguinal region and lower limb |
| ICD10CM | C82.96 | Follicular lymphoma, unspecified, intrapelvic lymph nodes |
| ICD10CM | C82.97 | Follicular lymphoma, unspecified, spleen |
| ICD10CM | C82.98 | Follicular lymphoma, unspecified, lymph nodes of multiple sites |
| ICD10CM | C82.99 | Follicular lymphoma, unspecified, extranodal and solid organ sites |
| ICD10CM | C83.0 | Small cell B-cell lymphoma |
| ICD10CM | C83.00 | Small cell B-cell lymphoma, unspecified site |
| ICD10CM | C83.01 | Small cell B-cell lymphoma, lymph nodes of head, face, and neck |
| ICD10CM | C83.02 | Small cell B-cell lymphoma, intrathoracic lymph nodes |
| ICD10CM | C83.03 | Small cell B-cell lymphoma, intra-abdominal lymph nodes |
| ICD10CM | C83.04 | Small cell B-cell lymphoma, lymph nodes of axilla and upper limb |
| ICD10CM | C83.05 | Small cell B-cell lymphoma, lymph nodes of inguinal region and lower limb |
| ICD10CM | C83.06 | Small cell B-cell lymphoma, intrapelvic lymph nodes |
| ICD10CM | C83.07 | Small cell B-cell lymphoma, spleen |
| ICD10CM | C83.08 | Small cell B-cell lymphoma, lymph nodes of multiple sites |
| ICD10CM | C83.09 | Small cell B-cell lymphoma, extranodal and solid organ sites |
| ICD10CM | C83.1 | Mantle cell lymphoma |
| ICD10CM | C83.10 | Mantle cell lymphoma, unspecified site |
| ICD10CM | C83.11 | Mantle cell lymphoma, lymph nodes of head, face, and neck |

Continued on next page

**Table 7 – continued from previous page**

| CodeSystem | ConceptCode | ConceptName |
|---|---|---|
| ICD10CM | C83.12 | Mantle cell lymphoma, intrathoracic lymph nodes |
| ICD10CM | C83.13 | Mantle cell lymphoma, intra-abdominal lymph nodes |
| ICD10CM | C83.14 | Mantle cell lymphoma, lymph nodes of axilla and upper limb |
| ICD10CM | C83.15 | Mantle cell lymphoma, lymph nodes of inguinal region and lower limb |
| ICD10CM | C83.16 | Mantle cell lymphoma, intrapelvic lymph nodes |
| ICD10CM | C83.17 | Mantle cell lymphoma, spleen |
| ICD10CM | C83.18 | Mantle cell lymphoma, lymph nodes of multiple sites |
| ICD10CM | C83.19 | Mantle cell lymphoma, extranodal and solid organ sites |
| ICD10CM | C83.3 | Diffuse large B-cell lymphoma |
| ICD10CM | C83.30 | Diffuse large B-cell lymphoma, unspecified site |
| ICD10CM | C83.31 | Diffuse large B-cell lymphoma, lymph nodes of head, face, and neck |
| ICD10CM | C83.32 | Diffuse large B-cell lymphoma, intrathoracic lymph nodes |
| ICD10CM | C83.33 | Diffuse large B-cell lymphoma, intra-abdominal lymph nodes |
| ICD10CM | C83.34 | Diffuse large B-cell lymphoma, lymph nodes of axilla and upper limb |
| ICD10CM | C83.35 | Diffuse large B-cell lymphoma, lymph nodes of inguinal region and lower limb |
| ICD10CM | C83.36 | Diffuse large B-cell lymphoma, intrapelvic lymph nodes |
| ICD10CM | C83.37 | Diffuse large B-cell lymphoma, spleen |
| ICD10CM | C83.38 | Diffuse large B-cell lymphoma, lymph nodes of multiple sites |
| ICD10CM | C83.39 | Diffuse large B-cell lymphoma, extranodal and solid organ sites |
| ICD10CM | C83.5 | Lymphoblastic (diffuse) lymphoma |
| ICD10CM | C83.50 | Lymphoblastic (diffuse) lymphoma, unspecified site |
| ICD10CM | C83.51 | Lymphoblastic (diffuse) lymphoma, lymph nodes of head, face, and neck |
| ICD10CM | C83.52 | Lymphoblastic (diffuse) lymphoma, intrathoracic lymph nodes |
| ICD10CM | C83.53 | Lymphoblastic (diffuse) lymphoma, intra-abdominal lymph nodes |
| ICD10CM | C83.54 | Lymphoblastic (diffuse) lymphoma, lymph nodes of axilla and upper limb |
| ICD10CM | C83.55 | Lymphoblastic (diffuse) lymphoma, lymph nodes of inguinal region and lower limb |
| ICD10CM | C83.56 | Lymphoblastic (diffuse) lymphoma, intrapelvic lymph nodes |
| ICD10CM | C83.57 | Lymphoblastic (diffuse) lymphoma, spleen |
| ICD10CM | C83.58 | Lymphoblastic (diffuse) lymphoma, lymph nodes of multiple sites |

Continued on next page

**Table 7 – continued from previous page**

| CodeSystem | ConceptCode | ConceptName |
|---|---|---|
| ICD10CM | C83.59 | Lymphoblastic (diffuse) lymphoma, extranodal and solid organ sites |
| ICD10CM | C83.7 | Burkitt lymphoma |
| ICD10CM | C83.70 | Burkitt lymphoma, unspecified site |
| ICD10CM | C83.71 | Burkitt lymphoma, lymph nodes of head, face, and neck |
| ICD10CM | C83.72 | Burkitt lymphoma, intrathoracic lymph nodes |
| ICD10CM | C83.73 | Burkitt lymphoma, intra-abdominal lymph nodes |
| ICD10CM | C83.74 | Burkitt lymphoma, lymph nodes of axilla and upper limb |
| ICD10CM | C83.75 | Burkitt lymphoma, lymph nodes of inguinal region and lower limb |
| ICD10CM | C83.76 | Burkitt lymphoma, intrapelvic lymph nodes |
| ICD10CM | C83.77 | Burkitt lymphoma, spleen |
| ICD10CM | C83.78 | Burkitt lymphoma, lymph nodes of multiple sites |
| ICD10CM | C83.79 | Burkitt lymphoma, extranodal and solid organ sites |
| ICD10CM | C83.8 | Other non-follicular lymphoma |
| ICD10CM | C83.80 | Other non-follicular lymphoma, unspecified site |
| ICD10CM | C83.81 | Other non-follicular lymphoma, lymph nodes of head, face, and neck |
| ICD10CM | C83.82 | Other non-follicular lymphoma, intrathoracic lymph nodes |
| ICD10CM | C83.83 | Other non-follicular lymphoma, intra-abdominal lymph nodes |
| ICD10CM | C83.84 | Other non-follicular lymphoma, lymph nodes of axilla and upper limb |
| ICD10CM | C83.85 | Other non-follicular lymphoma, lymph nodes of inguinal region and lower limb |
| ICD10CM | C83.86 | Other non-follicular lymphoma, intrapelvic lymph nodes |
| ICD10CM | C83.87 | Other non-follicular lymphoma, spleen |
| ICD10CM | C83.88 | Other non-follicular lymphoma, lymph nodes of multiple sites |
| ICD10CM | C83.89 | Other non-follicular lymphoma, extranodal and solid organ sites |
| ICD10CM | C83.9 | Non-follicular (diffuse) lymphoma, unspecified |
| ICD10CM | C83.90 | Non-follicular (diffuse) lymphoma, unspecified, unspecified site |
| ICD10CM | C83.91 | Non-follicular (diffuse) lymphoma, unspecified, lymph nodes of head, face, and neck |
| ICD10CM | C83.92 | Non-follicular (diffuse) lymphoma, unspecified, intrathoracic lymph nodes |
| ICD10CM | C83.93 | Non-follicular (diffuse) lymphoma, unspecified, intra-abdominal lymph nodes |
| ICD10CM | C83.94 | Non-follicular (diffuse) lymphoma, unspecified, lymph nodes of axilla and upper limb |

Continued on next page

**Table 7 – continued from previous page**

| CodeSystem | ConceptCode | ConceptName |
|---|---|---|
| ICD10CM | C83.95 | Non-follicular (diffuse) lymphoma, unspecified, lymph nodes of inguinal region and lower limb |
| ICD10CM | C83.96 | Non-follicular (diffuse) lymphoma, unspecified, intrapelvic lymph nodes |
| ICD10CM | C83.97 | Non-follicular (diffuse) lymphoma, unspecified, spleen |
| ICD10CM | C83.98 | Non-follicular (diffuse) lymphoma, unspecified, lymph nodes of multiple sites |
| ICD10CM | C83.99 | Non-follicular (diffuse) lymphoma, unspecified, extranodal and solid organ sites |
| ICD10CM | C84.0 | Mycosis fungoides |
| ICD10CM | C84.00 | Mycosis fungoides, unspecified site |
| ICD10CM | C84.01 | Mycosis fungoides, lymph nodes of head, face, and neck |
| ICD10CM | C84.02 | Mycosis fungoides, intrathoracic lymph nodes |
| ICD10CM | C84.03 | Mycosis fungoides, intra-abdominal lymph nodes |
| ICD10CM | C84.04 | Mycosis fungoides, lymph nodes of axilla and upper limb |
| ICD10CM | C84.05 | Mycosis fungoides, lymph nodes of inguinal region and lower limb |
| ICD10CM | C84.06 | Mycosis fungoides, intrapelvic lymph nodes |
| ICD10CM | C84.07 | Mycosis fungoides, spleen |
| ICD10CM | C84.08 | Mycosis fungoides, lymph nodes of multiple sites |
| ICD10CM | C84.09 | Mycosis fungoides, extranodal and solid organ sites |
| ICD10CM | C84.1 | Sezary disease |
| ICD10CM | C84.10 | Sezary disease, unspecified site |
| ICD10CM | C84.11 | Sezary disease, lymph nodes of head, face, and neck |
| ICD10CM | C84.12 | Sezary disease, intrathoracic lymph nodes |
| ICD10CM | C84.13 | Sezary disease, intra-abdominal lymph nodes |
| ICD10CM | C84.14 | Sezary disease, lymph nodes of axilla and upper limb |
| ICD10CM | C84.15 | Sezary disease, lymph nodes of inguinal region and lower limb |
| ICD10CM | C84.16 | Sezary disease, intrapelvic lymph nodes |
| ICD10CM | C84.17 | Sezary disease, spleen |
| ICD10CM | C84.18 | Sezary disease, lymph nodes of multiple sites |
| ICD10CM | C84.19 | Sezary disease, extranodal and solid organ sites |
| ICD10CM | C84.4 | Peripheral T-cell lymphoma, not classified |
| ICD10CM | C84.40 | Peripheral T-cell lymphoma, not classified, unspecified site |
| ICD10CM | C84.41 | Peripheral T-cell lymphoma, not classified, lymph nodes of head, face, and neck |
| ICD10CM | C84.42 | Peripheral T-cell lymphoma, not classified, intrathoracic lymph nodes |
| ICD10CM | C84.43 | Peripheral T-cell lymphoma, not classified, intra-abdominal lymph nodes |
| ICD10CM | C84.44 | Peripheral T-cell lymphoma, not classified, lymph nodes of axilla and upper limb |

Continued on next page

**Table 7 – continued from previous page**

| CodeSystem | ConceptCode | ConceptName |
|---|---|---|
| ICD10CM | C84.45 | Peripheral T-cell lymphoma, not classified, lymph nodes of inguinal region and lower limb |
| ICD10CM | C84.46 | Peripheral T-cell lymphoma, not classified, intrapelvic lymph nodes |
| ICD10CM | C84.47 | Peripheral T-cell lymphoma, not classified, spleen |
| ICD10CM | C84.48 | Peripheral T-cell lymphoma, not classified, lymph nodes of multiple sites |
| ICD10CM | C84.49 | Peripheral T-cell lymphoma, not classified, extranodal and solid organ sites |
| ICD10CM | C84.6 | Anaplastic large cell lymphoma, ALK-positive |
| ICD10CM | C84.60 | Anaplastic large cell lymphoma, ALK-positive, unspecified site |
| ICD10CM | C84.61 | Anaplastic large cell lymphoma, ALK-positive, lymph nodes of head, face, and neck |
| ICD10CM | C84.62 | Anaplastic large cell lymphoma, ALK-positive, intrathoracic lymph nodes |
| ICD10CM | C84.63 | Anaplastic large cell lymphoma, ALK-positive, intra-abdominal lymph nodes |
| ICD10CM | C84.64 | Anaplastic large cell lymphoma, ALK-positive, lymph nodes of axilla and upper limb |
| ICD10CM | C84.65 | Anaplastic large cell lymphoma, ALK-positive, lymph nodes of inguinal region and lower limb |
| ICD10CM | C84.66 | Anaplastic large cell lymphoma, ALK-positive, intrapelvic lymph nodes |
| ICD10CM | C84.67 | Anaplastic large cell lymphoma, ALK-positive, spleen |
| ICD10CM | C84.68 | Anaplastic large cell lymphoma, ALK-positive, lymph nodes of multiple sites |
| ICD10CM | C84.69 | Anaplastic large cell lymphoma, ALK-positive, extranodal and solid organ sites |
| ICD10CM | C84.7 | Anaplastic large cell lymphoma, ALK-negative |
| ICD10CM | C84.70 | Anaplastic large cell lymphoma, ALK-negative, unspecified site |
| ICD10CM | C84.71 | Anaplastic large cell lymphoma, ALK-negative, lymph nodes of head, face, and neck |
| ICD10CM | C84.72 | Anaplastic large cell lymphoma, ALK-negative, intrathoracic lymph nodes |
| ICD10CM | C84.73 | Anaplastic large cell lymphoma, ALK-negative, intra-abdominal lymph nodes |
| ICD10CM | C84.74 | Anaplastic large cell lymphoma, ALK-negative, lymph nodes of axilla and upper limb |
| ICD10CM | C84.75 | Anaplastic large cell lymphoma, ALK-negative, lymph nodes of inguinal region and lower limb |

Continued on next page

**Table 7 – continued from previous page**

| CodeSystem | ConceptCode | ConceptName |
|---|---|---|
| ICD10CM | C84.76 | Anaplastic large cell lymphoma, ALK-negative, intrapelvic lymph nodes |
| ICD10CM | C84.77 | Anaplastic large cell lymphoma, ALK-negative, spleen |
| ICD10CM | C84.78 | Anaplastic large cell lymphoma, ALK-negative, lymph nodes of multiple sites |
| ICD10CM | C84.79 | Anaplastic large cell lymphoma, ALK-negative, extranodal and solid organ sites |
| ICD10CM | C84.9 | Mature T/NK-cell lymphomas, unspecified |
| ICD10CM | C84.90 | Mature T/NK-cell lymphomas, unspecified, unspecified site |
| ICD10CM | C84.91 | Mature T/NK-cell lymphomas, unspecified, lymph nodes of head, face, and neck |
| ICD10CM | C84.92 | Mature T/NK-cell lymphomas, unspecified, intrathoracic lymph nodes |
| ICD10CM | C84.93 | Mature T/NK-cell lymphomas, unspecified, intra-abdominal lymph nodes |
| ICD10CM | C84.94 | Mature T/NK-cell lymphomas, unspecified, lymph nodes of axilla and upper limb |
| ICD10CM | C84.95 | Mature T/NK-cell lymphomas, unspecified, lymph nodes of inguinal region and lower limb |
| ICD10CM | C84.96 | Mature T/NK-cell lymphomas, unspecified, intrapelvic lymph nodes |
| ICD10CM | C84.97 | Mature T/NK-cell lymphomas, unspecified, spleen |
| ICD10CM | C84.98 | Mature T/NK-cell lymphomas, unspecified, lymph nodes of multiple sites |
| ICD10CM | C84.99 | Mature T/NK-cell lymphomas, unspecified, extranodal and solid organ sites |
| ICD10CM | C84.A | Cutaneous T-cell lymphoma, unspecified |
| ICD10CM | C84.A0 | Cutaneous T-cell lymphoma, unspecified, unspecified site |
| ICD10CM | C84.A1 | Cutaneous T-cell lymphoma, unspecified lymph nodes of head, face, and neck |
| ICD10CM | C84.A2 | Cutaneous T-cell lymphoma, unspecified, intrathoracic lymph nodes |
| ICD10CM | C84.A3 | Cutaneous T-cell lymphoma, unspecified, intra-abdominal lymph nodes |
| ICD10CM | C84.A4 | Cutaneous T-cell lymphoma, unspecified, lymph nodes of axilla and upper limb |
| ICD10CM | C84.A5 | Cutaneous T-cell lymphoma, unspecified, lymph nodes of inguinal region and lower limb |
| ICD10CM | C84.A6 | Cutaneous T-cell lymphoma, unspecified, intrapelvic lymph nodes |
| ICD10CM | C84.A7 | Cutaneous T-cell lymphoma, unspecified, spleen |
| ICD10CM | C84.A8 | Cutaneous T-cell lymphoma, unspecified, lymph nodes of multiple sites |

Continued on next page

**Table 7 – continued from previous page**

| CodeSystem | ConceptCode | ConceptName |
|---|---|---|
| ICD10CM | C84.A9 | Cutaneous T-cell lymphoma, unspecified, extranodal and solid organ sites |
| ICD10CM | C84.Z | Other mature T/NK-cell lymphomas |
| ICD10CM | C84.Z0 | Other mature T/NK-cell lymphomas, unspecified site |
| ICD10CM | C84.Z1 | Other mature T/NK-cell lymphomas, lymph nodes of head, face, and neck |
| ICD10CM | C84.Z2 | Other mature T/NK-cell lymphomas, intrathoracic lymph nodes |
| ICD10CM | C84.Z3 | Other mature T/NK-cell lymphomas, intra-abdominal lymph nodes |
| ICD10CM | C84.Z4 | Other mature T/NK-cell lymphomas, lymph nodes of axilla and upper limb |
| ICD10CM | C84.Z5 | Other mature T/NK-cell lymphomas, lymph nodes of inguinal region and lower limb |
| ICD10CM | C84.Z6 | Other mature T/NK-cell lymphomas, intrapelvic lymph nodes |
| ICD10CM | C84.Z7 | Other mature T/NK-cell lymphomas, spleen |
| ICD10CM | C84.Z8 | Other mature T/NK-cell lymphomas, lymph nodes of multiple sites |
| ICD10CM | C84.Z9 | Other mature T/NK-cell lymphomas, extranodal and solid organ sites |
| ICD10CM | C85.1 | Unspecified B-cell lymphoma |
| ICD10CM | C85.10 | Unspecified B-cell lymphoma, unspecified site |
| ICD10CM | C85.11 | Unspecified B-cell lymphoma, lymph nodes of head, face, and neck |
| ICD10CM | C85.12 | Unspecified B-cell lymphoma, intrathoracic lymph nodes |
| ICD10CM | C85.13 | Unspecified B-cell lymphoma, intra-abdominal lymph nodes |
| ICD10CM | C85.14 | Unspecified B-cell lymphoma, lymph nodes of axilla and upper limb |
| ICD10CM | C85.15 | Unspecified B-cell lymphoma, lymph nodes of inguinal region and lower limb |
| ICD10CM | C85.16 | Unspecified B-cell lymphoma, intrapelvic lymph nodes |
| ICD10CM | C85.17 | Unspecified B-cell lymphoma, spleen |
| ICD10CM | C85.18 | Unspecified B-cell lymphoma, lymph nodes of multiple sites |
| ICD10CM | C85.19 | Unspecified B-cell lymphoma, extranodal and solid organ sites |
| ICD10CM | C85.2 | Mediastinal (thymic) large B-cell lymphoma |
| ICD10CM | C85.20 | Mediastinal (thymic) large B-cell lymphoma, unspecified site |
| ICD10CM | C85.21 | Mediastinal (thymic) large B-cell lymphoma, lymph nodes of head, face, and neck |
| ICD10CM | C85.22 | Mediastinal (thymic) large B-cell lymphoma, intrathoracic lymph nodes |

Continued on next page

**Table 7 – continued from previous page**

| CodeSystem | ConceptCode | ConceptName |
|---|---|---|
| ICD10CM | C85.23 | Mediastinal (thymic) large B-cell lymphoma, intra-abdominal lymph nodes |
| ICD10CM | C85.24 | Mediastinal (thymic) large B-cell lymphoma, lymph nodes of axilla and upper limb |
| ICD10CM | C85.25 | Mediastinal (thymic) large B-cell lymphoma, lymph nodes of inguinal region and lower limb |
| ICD10CM | C85.26 | Mediastinal (thymic) large B-cell lymphoma, intrapelvic lymph nodes |
| ICD10CM | C85.27 | Mediastinal (thymic) large B-cell lymphoma, spleen |
| ICD10CM | C85.28 | Mediastinal (thymic) large B-cell lymphoma, lymph nodes of multiple sites |
| ICD10CM | C85.29 | Mediastinal (thymic) large B-cell lymphoma, extranodal and solid organ sites |
| ICD10CM | C85.8 | Other specified types of non-Hodgkin lymphoma |
| ICD10CM | C85.80 | Other specified types of non-Hodgkin lymphoma, unspecified site |
| ICD10CM | C85.81 | Other specified types of non-Hodgkin lymphoma, lymph nodes of head, face, and neck |
| ICD10CM | C85.82 | Other specified types of non-Hodgkin lymphoma, intrathoracic lymph nodes |
| ICD10CM | C85.83 | Other specified types of non-Hodgkin lymphoma, intra-abdominal lymph nodes |
| ICD10CM | C85.84 | Other specified types of non-Hodgkin lymphoma, lymph nodes of axilla and upper limb |
| ICD10CM | C85.85 | Other specified types of non-Hodgkin lymphoma, lymph nodes of inguinal region and lower limb |
| ICD10CM | C85.86 | Other specified types of non-Hodgkin lymphoma, intrapelvic lymph nodes |
| ICD10CM | C85.87 | Other specified types of non-Hodgkin lymphoma, spleen |
| ICD10CM | C85.88 | Other specified types of non-Hodgkin lymphoma, lymph nodes of multiple sites |
| ICD10CM | C85.89 | Other specified types of non-Hodgkin lymphoma, extranodal and solid organ sites |
| ICD10CM | C85.9 | Non-Hodgkin lymphoma, unspecified |
| ICD10CM | C85.90 | Non-Hodgkin lymphoma, unspecified, unspecified site |
| ICD10CM | C85.91 | Non-Hodgkin lymphoma, unspecified, lymph nodes of head, face, and neck |
| ICD10CM | C85.92 | Non-Hodgkin lymphoma, unspecified, intrathoracic lymph nodes |
| ICD10CM | C85.93 | Non-Hodgkin lymphoma, unspecified, intra-abdominal lymph nodes |
| ICD10CM | C85.94 | Non-Hodgkin lymphoma, unspecified, lymph nodes of axilla and upper limb |

Continued on next page

**Table 7 – continued from previous page**

| CodeSystem | ConceptCode | ConceptName |
|---|---|---|
| ICD10CM | C85.95 | Non-Hodgkin lymphoma, unspecified, lymph nodes of inguinal region and lower limb |
| ICD10CM | C85.96 | Non-Hodgkin lymphoma, unspecified, intrapelvic lymph nodes |
| ICD10CM | C85.97 | Non-Hodgkin lymphoma, unspecified, spleen |
| ICD10CM | C85.98 | Non-Hodgkin lymphoma, unspecified, lymph nodes of multiple sites |
| ICD10CM | C85.99 | Non-Hodgkin lymphoma, unspecified, extranodal and solid organ sites |
| ICD10CM | C86.0 | Extranodal NK/T-cell lymphoma, nasal type |
| ICD10CM | C86.1 | Hepatosplenic T-cell lymphoma |
| ICD10CM | C86.2 | Enteropathy-type (intestinal) T-cell lymphoma |
| ICD10CM | C86.3 | Subcutaneous panniculitis-like T-cell lymphoma |
| ICD10CM | C86.4 | Blastic NK-cell lymphoma |
| ICD10CM | C86.5 | Angioimmunoblastic T-cell lymphoma |
| ICD10CM | C86.6 | Primary cutaneous CD30-positive T-cell proliferations |
| ICD10CM | C88.0 | Waldenstrom macroglobulinemia |
| ICD10CM | C88.2 | Heavy chain disease |
| ICD10CM | C88.3 | Immunoproliferative small intestinal disease |
| ICD10CM | C88.4 | Extranodal marginal zone B-cell lymphoma of mucosa-associated lymphoid tissue [MALT-lymphoma] |
| ICD10CM | C88.8 | Other malignant immunoproliferative diseases |
| ICD10CM | C88.9 | Malignant immunoproliferative disease, unspecified |
| ICD10CM | C90.0 | Multiple myeloma |
| ICD10CM | C90.00 | Multiple myeloma not having achieved remission |
| ICD10CM | C90.01 | Multiple myeloma in remission |
| ICD10CM | C90.02 | Multiple myeloma in relapse |
| ICD10CM | C90.1 | Plasma cell leukemia |
| ICD10CM | C90.10 | Plasma cell leukemia not having achieved remission |
| ICD10CM | C90.11 | Plasma cell leukemia in remission |
| ICD10CM | C90.12 | Plasma cell leukemia in relapse |
| ICD10CM | C90.2 | Extramedullary plasmacytoma |
| ICD10CM | C90.20 | Extramedullary plasmacytoma not having achieved remission |
| ICD10CM | C90.21 | Extramedullary plasmacytoma in remission |
| ICD10CM | C90.22 | Extramedullary plasmacytoma in relapse |
| ICD10CM | C90.3 | Solitary plasmacytoma |
| ICD10CM | C90.30 | Solitary plasmacytoma not having achieved remission |
| ICD10CM | C90.31 | Solitary plasmacytoma in remission |
| ICD10CM | C90.32 | Solitary plasmacytoma in relapse |
| ICD10CM | C91.0 | Acute lymphoblastic leukemia [ALL] |
| ICD10CM | C91.00 | Acute lymphoblastic leukemia not having achieved remission |

Continued on next page

**Table 7 – continued from previous page**

| CodeSystem | ConceptCode | ConceptName |
|---|---|---|
| ICD10CM | C91.01 | Acute lymphoblastic leukemia, in remission |
| ICD10CM | C91.02 | Acute lymphoblastic leukemia, in relapse |
| ICD10CM | C91.1 | Chronic lymphocytic leukemia of B-cell type |
| ICD10CM | C91.10 | Chronic lymphocytic leukemia of B-cell type not having achieved remission |
| ICD10CM | C91.11 | Chronic lymphocytic leukemia of B-cell type in remission |
| ICD10CM | C91.12 | Chronic lymphocytic leukemia of B-cell type in relapse |
| ICD10CM | C91.3 | Prolymphocytic leukemia of B-cell type |
| ICD10CM | C91.30 | Prolymphocytic leukemia of B-cell type not having achieved remission |
| ICD10CM | C91.31 | Prolymphocytic leukemia of B-cell type, in remission |
| ICD10CM | C91.32 | Prolymphocytic leukemia of B-cell type, in relapse |
| ICD10CM | C91.4 | Hairy cell leukemia |
| ICD10CM | C91.40 | Hairy cell leukemia not having achieved remission |
| ICD10CM | C91.41 | Hairy cell leukemia, in remission |
| ICD10CM | C91.42 | Hairy cell leukemia, in relapse |
| ICD10CM | C91.5 | Adult T-cell lymphoma/leukemia (HTLV-1-associated) |
| ICD10CM | C91.50 | Adult T-cell lymphoma/leukemia (HTLV-1-associated) not having achieved remission |
| ICD10CM | C91.51 | Adult T-cell lymphoma/leukemia (HTLV-1-associated), in remission |
| ICD10CM | C91.52 | Adult T-cell lymphoma/leukemia (HTLV-1-associated), in relapse |
| ICD10CM | C91.6 | Prolymphocytic leukemia of T-cell type |
| ICD10CM | C91.60 | Prolymphocytic leukemia of T-cell type not having achieved remission |
| ICD10CM | C91.61 | Prolymphocytic leukemia of T-cell type, in remission |
| ICD10CM | C91.62 | Prolymphocytic leukemia of T-cell type, in relapse |
| ICD10CM | C91.9 | Lymphoid leukemia, unspecified |
| ICD10CM | C91.90 | Lymphoid leukemia, unspecified not having achieved remission |
| ICD10CM | C91.91 | Lymphoid leukemia, unspecified, in remission |
| ICD10CM | C91.92 | Lymphoid leukemia, unspecified, in relapse |
| ICD10CM | C91.A | Mature B-cell leukemia Burkitt-type |
| ICD10CM | C91.A0 | Mature B-cell leukemia Burkitt-type not having achieved remission |
| ICD10CM | C91.A1 | Mature B-cell leukemia Burkitt-type, in remission |
| ICD10CM | C91.A2 | Mature B-cell leukemia Burkitt-type, in relapse |
| ICD10CM | C91.Z | Other lymphoid leukemia |
| ICD10CM | C91.Z0 | Other lymphoid leukemia not having achieved remission |
| ICD10CM | C91.Z1 | Other lymphoid leukemia, in remission |
| ICD10CM | C91.Z2 | Other lymphoid leukemia, in relapse |
| ICD10CM | C92.0 | Acute myeloblastic leukemia |

Continued on next page

**Table 7 – continued from previous page**

| CodeSystem | ConceptCode | ConceptName |
|---|---|---|
| ICD10CM | C92.00 | Acute myeloblastic leukemia, not having achieved remission |
| ICD10CM | C92.01 | Acute myeloblastic leukemia, in remission |
| ICD10CM | C92.02 | Acute myeloblastic leukemia, in relapse |
| ICD10CM | C92.1 | Chronic myeloid leukemia, BCR/ABL-positive |
| ICD10CM | C92.10 | Chronic myeloid leukemia, BCR/ABL-positive, not having achieved remission |
| ICD10CM | C92.11 | Chronic myeloid leukemia, BCR/ABL-positive, in remission |
| ICD10CM | C92.12 | Chronic myeloid leukemia, BCR/ABL-positive, in relapse |
| ICD10CM | C92.2 | Atypical chronic myeloid leukemia, BCR/ABL-negative |
| ICD10CM | C92.20 | Atypical chronic myeloid leukemia, BCR/ABL-negative, not having achieved remission |
| ICD10CM | C92.21 | Atypical chronic myeloid leukemia, BCR/ABL-negative, in remission |
| ICD10CM | C92.22 | Atypical chronic myeloid leukemia, BCR/ABL-negative, in relapse |
| ICD10CM | C92.3 | Myeloid sarcoma |
| ICD10CM | C92.30 | Myeloid sarcoma, not having achieved remission |
| ICD10CM | C92.31 | Myeloid sarcoma, in remission |
| ICD10CM | C92.32 | Myeloid sarcoma, in relapse |
| ICD10CM | C92.4 | Acute promyelocytic leukemia |
| ICD10CM | C92.40 | Acute promyelocytic leukemia, not having achieved remission |
| ICD10CM | C92.41 | Acute promyelocytic leukemia, in remission |
| ICD10CM | C92.42 | Acute promyelocytic leukemia, in relapse |
| ICD10CM | C92.5 | Acute myelomonocytic leukemia |
| ICD10CM | C92.50 | Acute myelomonocytic leukemia, not having achieved remission |
| ICD10CM | C92.51 | Acute myelomonocytic leukemia, in remission |
| ICD10CM | C92.52 | Acute myelomonocytic leukemia, in relapse |
| ICD10CM | C92.6 | Acute myeloid leukemia with 11q23-abnormality |
| ICD10CM | C92.60 | Acute myeloid leukemia with 11q23-abnormality not having achieved remission |
| ICD10CM | C92.61 | Acute myeloid leukemia with 11q23-abnormality in remission |
| ICD10CM | C92.62 | Acute myeloid leukemia with 11q23-abnormality in relapse |
| ICD10CM | C92.9 | Myeloid leukemia, unspecified |
| ICD10CM | C92.90 | Myeloid leukemia, unspecified, not having achieved remission |
| ICD10CM | C92.91 | Myeloid leukemia, unspecified in remission |
| ICD10CM | C92.92 | Myeloid leukemia, unspecified in relapse |
| ICD10CM | C92.A | Acute myeloid leukemia with multilineage dysplasia |
| ICD10CM | C92.A0 | Acute myeloid leukemia with multilineage dysplasia, not having achieved remission |

Continued on next page

**Table 7 – continued from previous page**

| CodeSystem | ConceptCode | ConceptName |
|---|---|---|
| ICD10CM | C92.A1 | Acute myeloid leukemia with multilineage dysplasia, in remission |
| ICD10CM | C92.A2 | Acute myeloid leukemia with multilineage dysplasia, in relapse |
| ICD10CM | C92.Z | Other myeloid leukemia |
| ICD10CM | C92.Z0 | Other myeloid leukemia not having achieved remission |
| ICD10CM | C92.Z1 | Other myeloid leukemia, in remission |
| ICD10CM | C92.Z2 | Other myeloid leukemia, in relapse |
| ICD10CM | C93.0 | Acute monoblastic/monocytic leukemia |
| ICD10CM | C93.00 | Acute monoblastic/monocytic leukemia, not having achieved remission |
| ICD10CM | C93.01 | Acute monoblastic/monocytic leukemia, in remission |
| ICD10CM | C93.02 | Acute monoblastic/monocytic leukemia, in relapse |
| ICD10CM | C93.1 | Chronic myelomonocytic leukemia |
| ICD10CM | C93.10 | Chronic myelomonocytic leukemia not having achieved remission |
| ICD10CM | C93.11 | Chronic myelomonocytic leukemia, in remission |
| ICD10CM | C93.12 | Chronic myelomonocytic leukemia, in relapse |
| ICD10CM | C93.3 | Juvenile myelomonocytic leukemia |
| ICD10CM | C93.30 | Juvenile myelomonocytic leukemia, not having achieved remission |
| ICD10CM | C93.31 | Juvenile myelomonocytic leukemia, in remission |
| ICD10CM | C93.32 | Juvenile myelomonocytic leukemia, in relapse |
| ICD10CM | C93.9 | Monocytic leukemia, unspecified |
| ICD10CM | C93.90 | Monocytic leukemia, unspecified, not having achieved remission |
| ICD10CM | C93.91 | Monocytic leukemia, unspecified in remission |
| ICD10CM | C93.92 | Monocytic leukemia, unspecified in relapse |
| ICD10CM | C93.Z | Other monocytic leukemia |
| ICD10CM | C93.Z0 | Other monocytic leukemia, not having achieved remission |
| ICD10CM | C93.Z1 | Other monocytic leukemia, in remission |
| ICD10CM | C93.Z2 | Other monocytic leukemia, in relapse |
| ICD10CM | C94.0 | Acute erythroid leukemia |
| ICD10CM | C94.00 | Acute erythroid leukemia, not having achieved remission |
| ICD10CM | C94.01 | Acute erythroid leukemia, in remission |
| ICD10CM | C94.02 | Acute erythroid leukemia, in relapse |
| ICD10CM | C94.2 | Acute megakaryoblastic leukemia |
| ICD10CM | C94.20 | Acute megakaryoblastic leukemia not having achieved remission |
| ICD10CM | C94.21 | Acute megakaryoblastic leukemia, in remission |
| ICD10CM | C94.22 | Acute megakaryoblastic leukemia, in relapse |
| ICD10CM | C94.3 | Mast cell leukemia |
| ICD10CM | C94.30 | Mast cell leukemia not having achieved remission |

Continued on next page

**Table 7 – continued from previous page**

| CodeSystem | ConceptCode | ConceptName |
|---|---|---|
| ICD10CM | C94.31 | Mast cell leukemia, in remission |
| ICD10CM | C94.32 | Mast cell leukemia, in relapse |
| ICD10CM | C94.4 | Acute panmyelosis with myelofibrosis |
| ICD10CM | C94.40 | Acute panmyelosis with myelofibrosis not having achieved remission |
| ICD10CM | C94.41 | Acute panmyelosis with myelofibrosis, in remission |
| ICD10CM | C94.42 | Acute panmyelosis with myelofibrosis, in relapse |
| ICD10CM | C94.6 | Myelodysplastic disease, not classified |
| ICD10CM | C94.8 | Other specified leukemias |
| ICD10CM | C94.80 | Other specified leukemias not having achieved remission |
| ICD10CM | C94.81 | Other specified leukemias, in remission |
| ICD10CM | C94.82 | Other specified leukemias, in relapse |
| ICD10CM | C95.0 | Acute leukemia of unspecified cell type |
| ICD10CM | C95.00 | Acute leukemia of unspecified cell type not having achieved remission |
| ICD10CM | C95.01 | Acute leukemia of unspecified cell type, in remission |
| ICD10CM | C95.02 | Acute leukemia of unspecified cell type, in relapse |
| ICD10CM | C95.1 | Chronic leukemia of unspecified cell type |
| ICD10CM | C95.10 | Chronic leukemia of unspecified cell type not having achieved remission |
| ICD10CM | C95.11 | Chronic leukemia of unspecified cell type, in remission |
| ICD10CM | C95.12 | Chronic leukemia of unspecified cell type, in relapse |
| ICD10CM | C95.9 | Leukemia, unspecified |
| ICD10CM | C95.90 | Leukemia, unspecified not having achieved remission |
| ICD10CM | C95.91 | Leukemia, unspecified, in remission |
| ICD10CM | C95.92 | Leukemia, unspecified, in relapse |
| ICD10CM | C96.0 | Multifocal and multisystemic (disseminated) Langerhans-cell histiocytosis |
| ICD10CM | C96.2 | Malignant mast cell neoplasm |
| ICD10CM | C96.20 | Malignant mast cell neoplasm, unspecified |
| ICD10CM | C96.21 | Aggressive systemic mastocytosis |
| ICD10CM | C96.22 | Mast cell sarcoma |
| ICD10CM | C96.29 | Other malignant mast cell neoplasm |
| ICD10CM | C96.4 | Sarcoma of dendritic cells (accessory cells) |
| ICD10CM | C96.5 | Multifocal and unisystemic Langerhans-cell histiocytosis |
| ICD10CM | C96.6 | Unifocal Langerhans-cell histiocytosis |
| ICD10CM | C96.9 | Malignant neoplasm of lymphoid, hematopoietic and related tissue, unspecified |
| ICD10CM | C96.A | Histiocytic sarcoma |
| ICD10CM | C96.Z | Other specified malignant neoplasms of lymphoid, hematopoietic and related tissue |
| ICD10CM | D3A.0 | Benign carcinoid tumors |
| ICD10CM | D3A.00 | Benign carcinoid tumor of unspecified site |

Continued on next page

**Table 7 – continued from previous page**

| CodeSystem | ConceptCode | ConceptName |
|---|---|---|
| ICD10CM | D3A.01 | Benign carcinoid tumors of the small intestine |
| ICD10CM | D3A.010 | Benign carcinoid tumor of the duodenum |
| ICD10CM | D3A.011 | Benign carcinoid tumor of the jejunum |
| ICD10CM | D3A.012 | Benign carcinoid tumor of the ileum |
| ICD10CM | D3A.019 | Benign carcinoid tumor of the small intestine, unspecified portion |
| ICD10CM | D3A.02 | Benign carcinoid tumors of the appendix, large intestine, and rectum |
| ICD10CM | D3A.020 | Benign carcinoid tumor of the appendix |
| ICD10CM | D3A.021 | Benign carcinoid tumor of the cecum |
| ICD10CM | D3A.022 | Benign carcinoid tumor of the ascending colon |
| ICD10CM | D3A.023 | Benign carcinoid tumor of the transverse colon |
| ICD10CM | D3A.024 | Benign carcinoid tumor of the descending colon |
| ICD10CM | D3A.025 | Benign carcinoid tumor of the sigmoid colon |
| ICD10CM | D3A.026 | Benign carcinoid tumor of the rectum |
| ICD10CM | D3A.029 | Benign carcinoid tumor of the large intestine, unspecified portion |
| ICD10CM | D3A.09 | Benign carcinoid tumors of other sites |
| ICD10CM | D3A.090 | Benign carcinoid tumor of the bronchus and lung |
| ICD10CM | D3A.091 | Benign carcinoid tumor of the thymus |
| ICD10CM | D3A.092 | Benign carcinoid tumor of the stomach |
| ICD10CM | D3A.093 | Benign carcinoid tumor of the kidney |
| ICD10CM | D3A.094 | Benign carcinoid tumor of the foregut, unspecified |
| ICD10CM | D3A.095 | Benign carcinoid tumor of the midgut, unspecified |
| ICD10CM | D3A.096 | Benign carcinoid tumor of the hindgut, unspecified |
| ICD10CM | D3A.098 | Benign carcinoid tumors of other sites |
| ICD10CM | D3A.8 | Other benign neuroendocrine tumors |
| ICD10CM | D46.0 | Refractory anemia without ring sideroblasts, so stated |
| ICD10CM | D46.1 | Refractory anemia with ring sideroblasts |
| ICD10CM | D46.2 | Refractory anemia with excess of blasts [RAEB] |
| ICD10CM | D46.20 | Refractory anemia with excess of blasts, unspecified |
| ICD10CM | D46.21 | Refractory anemia with excess of blasts 1 |
| ICD10CM | D46.22 | Refractory anemia with excess of blasts 2 |
| ICD10CM | D46.4 | Refractory anemia, unspecified |
| ICD10CM | D46.9 | Myelodysplastic syndrome, unspecified |
| ICD10CM | D46.A | Refractory cytopenia with multilineage dysplasia |
| ICD10CM | D46.B | Refractory cytopenia with multilineage dysplasia and ring sideroblasts |
| ICD10CM | D46.C | Myelodysplastic syndrome with isolated del(5q) chromosomal abnormality |
| ICD10CM | D46.Z | Other myelodysplastic syndromes |
| ICD10CM | D61.01 | Constitutional (pure) red blood cell aplasia |
| ICD10CM | D80.0 | Hereditary hypogammaglobulinemia |

Continued on next page

**Table 7 – continued from previous page**

| CodeSystem | ConceptCode | ConceptName |
|---|---|---|
| ICD10CM | D80.1 | Nonfamilial hypogammaglobulinemia |
| ICD10CM | D80.2 | Selective deficiency of immunoglobulin A [IgA] |
| ICD10CM | D80.3 | Selective deficiency of immunoglobulin G [IgG] subclasses |
| ICD10CM | D80.4 | Selective deficiency of immunoglobulin M [IgM] |
| ICD10CM | D80.5 | Immunodeficiency with increased immunoglobulin M [IgM] |
| ICD10CM | D80.6 | Antibody deficiency with near-normal immunoglobulins or with hyperimmunoglobulinemia |
| ICD10CM | D80.7 | Transient hypogammaglobulinemia of infancy |
| ICD10CM | D80.8 | Other immunodeficiencies with predominantly antibody defects |
| ICD10CM | D80.9 | Immunodeficiency with predominantly antibody defects, unspecified |
| ICD10CM | D81.0 | Severe combined immunodeficiency [SCID] with reticular dysgenesis |
| ICD10CM | D81.1 | Severe combined immunodeficiency [SCID] with low T- and B-cell numbers |
| ICD10CM | D81.2 | Severe combined immunodeficiency [SCID] with low or normal B-cell numbers |
| ICD10CM | D81.3 | Adenosine deaminase [ADA] deficiency |
| ICD10CM | D81.30 | Adenosine deaminase deficiency, unspecified |
| ICD10CM | D81.31 | Severe combined immunodeficiency due to adenosine deaminase deficiency |
| ICD10CM | D81.32 | Adenosine deaminase 2 deficiency |
| ICD10CM | D81.39 | Other adenosine deaminase deficiency |
| ICD10CM | D81.4 | Nezelof's syndrome |
| ICD10CM | D81.5 | Purine nucleoside phosphorylase [PNP] deficiency |
| ICD10CM | D81.6 | Major histocompatibility complex class I deficiency |
| ICD10CM | D81.7 | Major histocompatibility complex class II deficiency |
| ICD10CM | D81.8 | Other combined immunodeficiencies |
| ICD10CM | D81.81 | Biotin-dependent carboxylase deficiency |
| ICD10CM | D81.810 | Biotinidase deficiency |
| ICD10CM | D81.818 | Other biotin-dependent carboxylase deficiency |
| ICD10CM | D81.819 | Biotin-dependent carboxylase deficiency, unspecified |
| ICD10CM | D81.89 | Other combined immunodeficiencies |
| ICD10CM | D81.9 | Combined immunodeficiency, unspecified |
| ICD10CM | D82.0 | Wiskott-Aldrich syndrome |
| ICD10CM | D82.1 | Di George's syndrome |
| ICD10CM | D82.2 | Immunodeficiency with short-limbed stature |
| ICD10CM | D82.3 | Immunodeficiency following hereditary defective response to Epstein-Barr virus |
| ICD10CM | D82.4 | Hyperimmunoglobulin E [IgE] syndrome |
| ICD10CM | D82.8 | Immunodeficiency associated with other specified major defects |

Continued on next page

**Table 7 – continued from previous page**

| CodeSystem | ConceptCode | ConceptName |
|---|---|---|
| ICD10CM | D82.9 | Immunodeficiency associated with major defect, unspecified |
| ICD10CM | D89.4 | Mast cell activation syndrome and related disorders |
| ICD10CM | D83.0 | Common variable immunodeficiency with predominant abnormalities of B-cell numbers and function |
| ICD10CM | D83.1 | Common variable immunodeficiency with predominant immunoregulatory T-cell disorders |
| ICD10CM | D83.2 | Common variable immunodeficiency with autoantibodies to B- or T-cells |
| ICD10CM | D83.8 | Other common variable immunodeficiencies |
| ICD10CM | D83.9 | Common variable immunodeficiency, unspecified |
| ICD10CM | D84.0 | Lymphocyte function antigen-1 [LFA-1] defect |
| ICD10CM | D84.1 | Defects in the complement system |
| ICD10CM | D84.8 | Other specified immunodeficiencies |
| ICD10CM | D84.9 | Immunodeficiency, unspecified |
| ICD10CM | D89.0 | Polyclonal hypergammaglobulinemia |
| ICD10CM | D89.1 | Cryoglobulinemia |
| ICD10CM | D89.2 | Hypergammaglobulinemia, unspecified |
| ICD10CM | D89.3 | Immune reconstitution syndrome |
| ICD10CM | D89.40 | Mast cell activation, unspecified |
| ICD10CM | D89.41 | Monoclonal mast cell activation syndrome |
| ICD10CM | D89.42 | Idiopathic mast cell activation syndrome |
| ICD10CM | D89.43 | Secondary mast cell activation |
| ICD10CM | D89.49 | Other mast cell activation disorder |
| ICD10CM | D89.8 | Other specified disorders involving the immune mechanism, not elsewhere classified |
| ICD10CM | D89.81 | Graft-versus-host disease |
| ICD10CM | D89.810 | Acute graft-versus-host disease |
| ICD10CM | D89.811 | Chronic graft-versus-host disease |
| ICD10CM | D89.812 | Acute on chronic graft-versus-host disease |
| ICD10CM | D89.813 | Graft-versus-host disease, unspecified |
| ICD10CM | D89.82 | Autoimmune lymphoproliferative syndrome [ALPS] |
| ICD10CM | D89.89 | Other specified disorders involving the immune mechanism, not elsewhere classified |
| ICD10CM | D89.9 | Disorder involving the immune mechanism, unspecified |
| ICD10CM | K70.30 | Alcoholic cirrhosis of liver without ascites |
| ICD10CM | K70.31 | Alcoholic cirrhosis of liver with ascites |
| ICD10CM | K70.40 | Alcoholic hepatic failure without coma |
| ICD10CM | K70.41 | Alcoholic hepatic failure with coma |
| ICD10CM | K72.0 | Acute and subacute hepatic failure |
| ICD10CM | K72.00 | Acute and subacute hepatic failure without coma |
| ICD10CM | K72.01 | Acute and subacute hepatic failure with coma |
| ICD10CM | K72.1 | Chronic hepatic failure |
| ICD10CM | K72.10 | Chronic hepatic failure without coma |

Continued on next page

**Table 7 – continued from previous page**

| CodeSystem | ConceptCode | ConceptName |
|---|---|---|
| ICD10CM | K72.11 | Chronic hepatic failure with coma |
| ICD10CM | K72.9 | Hepatic failure, unspecified |
| ICD10CM | K72.90 | Hepatic failure, unspecified without coma |
| ICD10CM | K72.91 | Hepatic failure, unspecified with coma |
| ICD10CM | K74.60 | Unspecified cirrhosis of liver |
| ICD10CM | K74.69 | Other cirrhosis of liver |
| ICD10CM | L94.0 | Localized scleroderma [morphea] |
| ICD10CM | L94.1 | Linear scleroderma |
| ICD10CM | L94.2 | Calcinosis cutis |
| ICD10CM | L94.3 | Sclerodactyly |
| ICD10CM | L94.4 | Gottron's papules |
| ICD10CM | L94.5 | Poikiloderma vasculare atrophicans |
| ICD10CM | L94.6 | Ainhum |
| ICD10CM | L94.8 | Other specified localized connective tissue disorders |
| ICD10CM | L94.9 | Localized connective tissue disorder, unspecified |
| ICD10CM | M05.0 | Felty's syndrome |
| ICD10CM | M05.00 | Felty's syndrome, unspecified site |
| ICD10CM | M05.01 | Felty's syndrome, shoulder |
| ICD10CM | M05.011 | Felty's syndrome, right shoulder |
| ICD10CM | M05.012 | Felty's syndrome, left shoulder |
| ICD10CM | M05.019 | Felty's syndrome, unspecified shoulder |
| ICD10CM | M05.02 | Felty's syndrome, elbow |
| ICD10CM | M05.021 | Felty's syndrome, right elbow |
| ICD10CM | M05.022 | Felty's syndrome, left elbow |
| ICD10CM | M05.029 | Felty's syndrome, unspecified elbow |
| ICD10CM | M05.03 | Felty's syndrome, wrist |
| ICD10CM | M05.031 | Felty's syndrome, right wrist |
| ICD10CM | M05.032 | Felty's syndrome, left wrist |
| ICD10CM | M05.039 | Felty's syndrome, unspecified wrist |
| ICD10CM | M05.04 | Felty's syndrome, hand |
| ICD10CM | M05.041 | Felty's syndrome, right hand |
| ICD10CM | M05.042 | Felty's syndrome, left hand |
| ICD10CM | M05.049 | Felty's syndrome, unspecified hand |
| ICD10CM | M05.05 | Felty's syndrome, hip |
| ICD10CM | M05.051 | Felty's syndrome, right hip |
| ICD10CM | M05.052 | Felty's syndrome, left hip |
| ICD10CM | M05.059 | Felty's syndrome, unspecified hip |
| ICD10CM | M05.06 | Felty's syndrome, knee |
| ICD10CM | M05.061 | Felty's syndrome, right knee |
| ICD10CM | M05.062 | Felty's syndrome, left knee |
| ICD10CM | M05.069 | Felty's syndrome, unspecified knee |
| ICD10CM | M05.07 | Felty's syndrome, ankle and foot |
| ICD10CM | M05.071 | Felty's syndrome, right ankle and foot |

Continued on next page

**Table 7 – continued from previous page**

| CodeSystem | ConceptCode | ConceptName |
|---|---|---|
| ICD10CM | M05.072 | Felty's syndrome, left ankle and foot |
| ICD10CM | M05.079 | Felty's syndrome, unspecified ankle and foot |
| ICD10CM | M05.09 | Felty's syndrome, multiple sites |
| ICD10CM | M05.1 | Rheumatoid lung disease with rheumatoid arthritis |
| ICD10CM | M05.10 | Rheumatoid lung disease with rheumatoid arthritis of unspecified site |
| ICD10CM | M05.11 | Rheumatoid lung disease with rheumatoid arthritis of shoulder |
| ICD10CM | M05.111 | Rheumatoid lung disease with rheumatoid arthritis of right shoulder |
| ICD10CM | M05.112 | Rheumatoid lung disease with rheumatoid arthritis of left shoulder |
| ICD10CM | M05.119 | Rheumatoid lung disease with rheumatoid arthritis of unspecified shoulder |
| ICD10CM | M05.12 | Rheumatoid lung disease with rheumatoid arthritis of elbow |
| ICD10CM | M05.121 | Rheumatoid lung disease with rheumatoid arthritis of right elbow |
| ICD10CM | M05.122 | Rheumatoid lung disease with rheumatoid arthritis of left elbow |
| ICD10CM | M05.129 | Rheumatoid lung disease with rheumatoid arthritis of unspecified elbow |
| ICD10CM | M05.13 | Rheumatoid lung disease with rheumatoid arthritis of wrist |
| ICD10CM | M05.131 | Rheumatoid lung disease with rheumatoid arthritis of right wrist |
| ICD10CM | M05.132 | Rheumatoid lung disease with rheumatoid arthritis of left wrist |
| ICD10CM | M05.139 | Rheumatoid lung disease with rheumatoid arthritis of unspecified wrist |
| ICD10CM | M05.14 | Rheumatoid lung disease with rheumatoid arthritis of hand |
| ICD10CM | M05.141 | Rheumatoid lung disease with rheumatoid arthritis of right hand |
| ICD10CM | M05.142 | Rheumatoid lung disease with rheumatoid arthritis of left hand |
| ICD10CM | M05.149 | Rheumatoid lung disease with rheumatoid arthritis of unspecified hand |
| ICD10CM | M05.15 | Rheumatoid lung disease with rheumatoid arthritis of hip |
| ICD10CM | M05.151 | Rheumatoid lung disease with rheumatoid arthritis of right hip |
| ICD10CM | M05.152 | Rheumatoid lung disease with rheumatoid arthritis of left hip |
| ICD10CM | M05.159 | Rheumatoid lung disease with rheumatoid arthritis of unspecified hip |
| ICD10CM | M05.16 | Rheumatoid lung disease with rheumatoid arthritis of knee |

Continued on next page

**Table 7 – continued from previous page**

| CodeSystem | ConceptCode | ConceptName |
|---|---|---|
| ICD10CM | M05.161 | Rheumatoid lung disease with rheumatoid arthritis of right knee |
| ICD10CM | M05.162 | Rheumatoid lung disease with rheumatoid arthritis of left knee |
| ICD10CM | M05.169 | Rheumatoid lung disease with rheumatoid arthritis of unspecified knee |
| ICD10CM | M05.17 | Rheumatoid lung disease with rheumatoid arthritis of ankle and foot |
| ICD10CM | M05.171 | Rheumatoid lung disease with rheumatoid arthritis of right ankle and foot |
| ICD10CM | M05.172 | Rheumatoid lung disease with rheumatoid arthritis of left ankle and foot |
| ICD10CM | M05.179 | Rheumatoid lung disease with rheumatoid arthritis of unspecified ankle and foot |
| ICD10CM | M05.19 | Rheumatoid lung disease with rheumatoid arthritis of multiple sites |
| ICD10CM | M05.2 | Rheumatoid vasculitis with rheumatoid arthritis |
| ICD10CM | M05.20 | Rheumatoid vasculitis with rheumatoid arthritis of unspecified site |
| ICD10CM | M05.21 | Rheumatoid vasculitis with rheumatoid arthritis of shoulder |
| ICD10CM | M05.211 | Rheumatoid vasculitis with rheumatoid arthritis of right shoulder |
| ICD10CM | M05.212 | Rheumatoid vasculitis with rheumatoid arthritis of left shoulder |
| ICD10CM | M05.219 | Rheumatoid vasculitis with rheumatoid arthritis of unspecified shoulder |
| ICD10CM | M05.22 | Rheumatoid vasculitis with rheumatoid arthritis of elbow |
| ICD10CM | M05.221 | Rheumatoid vasculitis with rheumatoid arthritis of right elbow |
| ICD10CM | M05.222 | Rheumatoid vasculitis with rheumatoid arthritis of left elbow |
| ICD10CM | M05.229 | Rheumatoid vasculitis with rheumatoid arthritis of unspecified elbow |
| ICD10CM | M05.23 | Rheumatoid vasculitis with rheumatoid arthritis of wrist |
| ICD10CM | M05.231 | Rheumatoid vasculitis with rheumatoid arthritis of right wrist |
| ICD10CM | M05.232 | Rheumatoid vasculitis with rheumatoid arthritis of left wrist |
| ICD10CM | M05.239 | Rheumatoid vasculitis with rheumatoid arthritis of unspecified wrist |
| ICD10CM | M05.24 | Rheumatoid vasculitis with rheumatoid arthritis of hand |
| ICD10CM | M05.241 | Rheumatoid vasculitis with rheumatoid arthritis of right hand |
| ICD10CM | M05.242 | Rheumatoid vasculitis with rheumatoid arthritis of left hand |

Continued on next page

**Table 7 – continued from previous page**

| CodeSystem | ConceptCode | ConceptName |
|---|---|---|
| ICD10CM | M05.249 | Rheumatoid vasculitis with rheumatoid arthritis of unspecified hand |
| ICD10CM | M05.25 | Rheumatoid vasculitis with rheumatoid arthritis of hip |
| ICD10CM | M05.251 | Rheumatoid vasculitis with rheumatoid arthritis of right hip |
| ICD10CM | M05.252 | Rheumatoid vasculitis with rheumatoid arthritis of left hip |
| ICD10CM | M05.259 | Rheumatoid vasculitis with rheumatoid arthritis of unspecified hip |
| ICD10CM | M05.26 | Rheumatoid vasculitis with rheumatoid arthritis of knee |
| ICD10CM | M05.261 | Rheumatoid vasculitis with rheumatoid arthritis of right knee |
| ICD10CM | M05.262 | Rheumatoid vasculitis with rheumatoid arthritis of left knee |
| ICD10CM | M05.269 | Rheumatoid vasculitis with rheumatoid arthritis of unspecified knee |
| ICD10CM | M05.27 | Rheumatoid vasculitis with rheumatoid arthritis of ankle and foot |
| ICD10CM | M05.271 | Rheumatoid vasculitis with rheumatoid arthritis of right ankle and foot |
| ICD10CM | M05.272 | Rheumatoid vasculitis with rheumatoid arthritis of left ankle and foot |
| ICD10CM | M05.279 | Rheumatoid vasculitis with rheumatoid arthritis of unspecified ankle and foot |
| ICD10CM | M05.29 | Rheumatoid vasculitis with rheumatoid arthritis of multiple sites |
| ICD10CM | M05.3 | Rheumatoid heart disease with rheumatoid arthritis |
| ICD10CM | M05.30 | Rheumatoid heart disease with rheumatoid arthritis of unspecified site |
| ICD10CM | M05.31 | Rheumatoid heart disease with rheumatoid arthritis of shoulder |
| ICD10CM | M05.311 | Rheumatoid heart disease with rheumatoid arthritis of right shoulder |
| ICD10CM | M05.312 | Rheumatoid heart disease with rheumatoid arthritis of left shoulder |
| ICD10CM | M05.319 | Rheumatoid heart disease with rheumatoid arthritis of unspecified shoulder |
| ICD10CM | M05.32 | Rheumatoid heart disease with rheumatoid arthritis of elbow |
| ICD10CM | M05.321 | Rheumatoid heart disease with rheumatoid arthritis of right elbow |
| ICD10CM | M05.322 | Rheumatoid heart disease with rheumatoid arthritis of left elbow |
| ICD10CM | M05.329 | Rheumatoid heart disease with rheumatoid arthritis of unspecified elbow |
| ICD10CM | M05.33 | Rheumatoid heart disease with rheumatoid arthritis of wrist |

Continued on next page

**Table 7 – continued from previous page**

| CodeSystem | ConceptCode | ConceptName |
|---|---|---|
| ICD10CM | M05.331 | Rheumatoid heart disease with rheumatoid arthritis of right wrist |
| ICD10CM | M05.332 | Rheumatoid heart disease with rheumatoid arthritis of left wrist |
| ICD10CM | M05.339 | Rheumatoid heart disease with rheumatoid arthritis of unspecified wrist |
| ICD10CM | M05.34 | Rheumatoid heart disease with rheumatoid arthritis of hand |
| ICD10CM | M05.341 | Rheumatoid heart disease with rheumatoid arthritis of right hand |
| ICD10CM | M05.342 | Rheumatoid heart disease with rheumatoid arthritis of left hand |
| ICD10CM | M05.349 | Rheumatoid heart disease with rheumatoid arthritis of unspecified hand |
| ICD10CM | M05.35 | Rheumatoid heart disease with rheumatoid arthritis of hip |
| ICD10CM | M05.351 | Rheumatoid heart disease with rheumatoid arthritis of right hip |
| ICD10CM | M05.352 | Rheumatoid heart disease with rheumatoid arthritis of left hip |
| ICD10CM | M05.359 | Rheumatoid heart disease with rheumatoid arthritis of unspecified hip |
| ICD10CM | M05.36 | Rheumatoid heart disease with rheumatoid arthritis of knee |
| ICD10CM | M05.361 | Rheumatoid heart disease with rheumatoid arthritis of right knee |
| ICD10CM | M05.362 | Rheumatoid heart disease with rheumatoid arthritis of left knee |
| ICD10CM | M05.369 | Rheumatoid heart disease with rheumatoid arthritis of unspecified knee |
| ICD10CM | M05.37 | Rheumatoid heart disease with rheumatoid arthritis of ankle and foot |
| ICD10CM | M05.371 | Rheumatoid heart disease with rheumatoid arthritis of right ankle and foot |
| ICD10CM | M05.372 | Rheumatoid heart disease with rheumatoid arthritis of left ankle and foot |
| ICD10CM | M05.379 | Rheumatoid heart disease with rheumatoid arthritis of unspecified ankle and foot |
| ICD10CM | M05.39 | Rheumatoid heart disease with rheumatoid arthritis of multiple sites |
| ICD10CM | M05.4 | Rheumatoid myopathy with rheumatoid arthritis |
| ICD10CM | M05.40 | Rheumatoid myopathy with rheumatoid arthritis of unspecified site |
| ICD10CM | M05.41 | Rheumatoid myopathy with rheumatoid arthritis of shoulder |

Continued on next page

**Table 7 – continued from previous page**

| CodeSystem | ConceptCode | ConceptName |
|---|---|---|
| ICD10CM | M05.411 | Rheumatoid myopathy with rheumatoid arthritis of right shoulder |
| ICD10CM | M05.412 | Rheumatoid myopathy with rheumatoid arthritis of left shoulder |
| ICD10CM | M05.419 | Rheumatoid myopathy with rheumatoid arthritis of unspecified shoulder |
| ICD10CM | M05.42 | Rheumatoid myopathy with rheumatoid arthritis of elbow |
| ICD10CM | M05.421 | Rheumatoid myopathy with rheumatoid arthritis of right elbow |
| ICD10CM | M05.422 | Rheumatoid myopathy with rheumatoid arthritis of left elbow |
| ICD10CM | M05.429 | Rheumatoid myopathy with rheumatoid arthritis of unspecified elbow |
| ICD10CM | M05.43 | Rheumatoid myopathy with rheumatoid arthritis of wrist |
| ICD10CM | M05.431 | Rheumatoid myopathy with rheumatoid arthritis of right wrist |
| ICD10CM | M05.432 | Rheumatoid myopathy with rheumatoid arthritis of left wrist |
| ICD10CM | M05.439 | Rheumatoid myopathy with rheumatoid arthritis of unspecified wrist |
| ICD10CM | M05.44 | Rheumatoid myopathy with rheumatoid arthritis of hand |
| ICD10CM | M05.441 | Rheumatoid myopathy with rheumatoid arthritis of right hand |
| ICD10CM | M05.442 | Rheumatoid myopathy with rheumatoid arthritis of left hand |
| ICD10CM | M05.449 | Rheumatoid myopathy with rheumatoid arthritis of unspecified hand |
| ICD10CM | M05.45 | Rheumatoid myopathy with rheumatoid arthritis of hip |
| ICD10CM | M05.451 | Rheumatoid myopathy with rheumatoid arthritis of right hip |
| ICD10CM | M05.452 | Rheumatoid myopathy with rheumatoid arthritis of left hip |
| ICD10CM | M05.459 | Rheumatoid myopathy with rheumatoid arthritis of unspecified hip |
| ICD10CM | M05.46 | Rheumatoid myopathy with rheumatoid arthritis of knee |
| ICD10CM | M05.461 | Rheumatoid myopathy with rheumatoid arthritis of right knee |
| ICD10CM | M05.462 | Rheumatoid myopathy with rheumatoid arthritis of left knee |
| ICD10CM | M05.469 | Rheumatoid myopathy with rheumatoid arthritis of unspecified knee |
| ICD10CM | M05.47 | Rheumatoid myopathy with rheumatoid arthritis of ankle and foot |

Continued on next page

**Table 7 – continued from previous page**

| CodeSystem | ConceptCode | ConceptName |
|---|---|---|
| ICD10CM | M05.471 | Rheumatoid myopathy with rheumatoid arthritis of right ankle and foot |
| ICD10CM | M05.472 | Rheumatoid myopathy with rheumatoid arthritis of left ankle and foot |
| ICD10CM | M05.479 | Rheumatoid myopathy with rheumatoid arthritis of unspecified ankle and foot |
| ICD10CM | M05.49 | Rheumatoid myopathy with rheumatoid arthritis of multiple sites |
| ICD10CM | M05.5 | Rheumatoid polyneuropathy with rheumatoid arthritis |
| ICD10CM | M05.50 | Rheumatoid polyneuropathy with rheumatoid arthritis of unspecified site |
| ICD10CM | M05.51 | Rheumatoid polyneuropathy with rheumatoid arthritis of shoulder |
| ICD10CM | M05.511 | Rheumatoid polyneuropathy with rheumatoid arthritis of right shoulder |
| ICD10CM | M05.512 | Rheumatoid polyneuropathy with rheumatoid arthritis of left shoulder |
| ICD10CM | M05.519 | Rheumatoid polyneuropathy with rheumatoid arthritis of unspecified shoulder |
| ICD10CM | M05.52 | Rheumatoid polyneuropathy with rheumatoid arthritis of elbow |
| ICD10CM | M05.521 | Rheumatoid polyneuropathy with rheumatoid arthritis of right elbow |
| ICD10CM | M05.522 | Rheumatoid polyneuropathy with rheumatoid arthritis of left elbow |
| ICD10CM | M05.529 | Rheumatoid polyneuropathy with rheumatoid arthritis of unspecified elbow |
| ICD10CM | M05.53 | Rheumatoid polyneuropathy with rheumatoid arthritis of wrist |
| ICD10CM | M05.531 | Rheumatoid polyneuropathy with rheumatoid arthritis of right wrist |
| ICD10CM | M05.532 | Rheumatoid polyneuropathy with rheumatoid arthritis of left wrist |
| ICD10CM | M05.539 | Rheumatoid polyneuropathy with rheumatoid arthritis of unspecified wrist |
| ICD10CM | M05.54 | Rheumatoid polyneuropathy with rheumatoid arthritis of hand |
| ICD10CM | M05.541 | Rheumatoid polyneuropathy with rheumatoid arthritis of right hand |
| ICD10CM | M05.542 | Rheumatoid polyneuropathy with rheumatoid arthritis of left hand |
| ICD10CM | M05.549 | Rheumatoid polyneuropathy with rheumatoid arthritis of unspecified hand |

Continued on next page

**Table 7 – continued from previous page**

| CodeSystem | ConceptCode | ConceptName |
|---|---|---|
| ICD10CM | M05.55 | Rheumatoid polyneuropathy with rheumatoid arthritis of hip |
| ICD10CM | M05.551 | Rheumatoid polyneuropathy with rheumatoid arthritis of right hip |
| ICD10CM | M05.552 | Rheumatoid polyneuropathy with rheumatoid arthritis of left hip |
| ICD10CM | M05.559 | Rheumatoid polyneuropathy with rheumatoid arthritis of unspecified hip |
| ICD10CM | M05.56 | Rheumatoid polyneuropathy with rheumatoid arthritis of knee |
| ICD10CM | M05.561 | Rheumatoid polyneuropathy with rheumatoid arthritis of right knee |
| ICD10CM | M05.562 | Rheumatoid polyneuropathy with rheumatoid arthritis of left knee |
| ICD10CM | M05.569 | Rheumatoid polyneuropathy with rheumatoid arthritis of unspecified knee |
| ICD10CM | M05.57 | Rheumatoid polyneuropathy with rheumatoid arthritis of ankle and foot |
| ICD10CM | M05.571 | Rheumatoid polyneuropathy with rheumatoid arthritis of right ankle and foot |
| ICD10CM | M05.572 | Rheumatoid polyneuropathy with rheumatoid arthritis of left ankle and foot |
| ICD10CM | M05.579 | Rheumatoid polyneuropathy with rheumatoid arthritis of unspecified ankle and foot |
| ICD10CM | M05.59 | Rheumatoid polyneuropathy with rheumatoid arthritis of multiple sites |
| ICD10CM | M05.6 | Rheumatoid arthritis with involvement of other organs and systems |
| ICD10CM | M05.60 | Rheumatoid arthritis of unspecified site with involvement of other organs and systems |
| ICD10CM | M05.61 | Rheumatoid arthritis of shoulder with involvement of other organs and systems |
| ICD10CM | M05.611 | Rheumatoid arthritis of right shoulder with involvement of other organs and systems |
| ICD10CM | M05.612 | Rheumatoid arthritis of left shoulder with involvement of other organs and systems |
| ICD10CM | M05.619 | Rheumatoid arthritis of unspecified shoulder with involvement of other organs and systems |
| ICD10CM | M05.62 | Rheumatoid arthritis of elbow with involvement of other organs and systems |
| ICD10CM | M05.621 | Rheumatoid arthritis of right elbow with involvement of other organs and systems |

Continued on next page

**Table 7 – continued from previous page**

| CodeSystem | ConceptCode | ConceptName |
|---|---|---|
| ICD10CM | M05.622 | Rheumatoid arthritis of left elbow with involvement of other organs and systems |
| ICD10CM | M05.629 | Rheumatoid arthritis of unspecified elbow with involvement of other organs and systems |
| ICD10CM | M05.63 | Rheumatoid arthritis of wrist with involvement of other organs and systems |
| ICD10CM | M05.631 | Rheumatoid arthritis of right wrist with involvement of other organs and systems |
| ICD10CM | M05.632 | Rheumatoid arthritis of left wrist with involvement of other organs and systems |
| ICD10CM | M05.639 | Rheumatoid arthritis of unspecified wrist with involvement of other organs and systems |
| ICD10CM | M05.64 | Rheumatoid arthritis of hand with involvement of other organs and systems |
| ICD10CM | M05.641 | Rheumatoid arthritis of right hand with involvement of other organs and systems |
| ICD10CM | M05.642 | Rheumatoid arthritis of left hand with involvement of other organs and systems |
| ICD10CM | M05.649 | Rheumatoid arthritis of unspecified hand with involvement of other organs and systems |
| ICD10CM | M05.65 | Rheumatoid arthritis of hip with involvement of other organs and systems |
| ICD10CM | M05.651 | Rheumatoid arthritis of right hip with involvement of other organs and systems |
| ICD10CM | M05.652 | Rheumatoid arthritis of left hip with involvement of other organs and systems |
| ICD10CM | M05.659 | Rheumatoid arthritis of unspecified hip with involvement of other organs and systems |
| ICD10CM | M05.66 | Rheumatoid arthritis of knee with involvement of other organs and systems |
| ICD10CM | M05.661 | Rheumatoid arthritis of right knee with involvement of other organs and systems |
| ICD10CM | M05.662 | Rheumatoid arthritis of left knee with involvement of other organs and systems |
| ICD10CM | M05.669 | Rheumatoid arthritis of unspecified knee with involvement of other organs and systems |
| ICD10CM | M05.67 | Rheumatoid arthritis of ankle and foot with involvement of other organs and systems |
| ICD10CM | M05.671 | Rheumatoid arthritis of right ankle and foot with involvement of other organs and systems |
| ICD10CM | M05.672 | Rheumatoid arthritis of left ankle and foot with involvement of other organs and systems |

Continued on next page

**Table 7 – continued from previous page**

| CodeSystem | ConceptCode | ConceptName |
|---|---|---|
| ICD10CM | M05.679 | Rheumatoid arthritis of unspecified ankle and foot with involvement of other organs and systems |
| ICD10CM | M05.69 | Rheumatoid arthritis of multiple sites with involvement of other organs and systems |
| ICD10CM | M05.7 | Rheumatoid arthritis with rheumatoid factor without organ or systems involvement |
| ICD10CM | M05.70 | Rheumatoid arthritis with rheumatoid factor of unspecified site without organ or systems involvement |
| ICD10CM | M05.71 | Rheumatoid arthritis with rheumatoid factor of shoulder without organ or systems involvement |
| ICD10CM | M05.711 | Rheumatoid arthritis with rheumatoid factor of right shoulder without organ or systems involvement |
| ICD10CM | M05.712 | Rheumatoid arthritis with rheumatoid factor of left shoulder without organ or systems involvement |
| ICD10CM | M05.719 | Rheumatoid arthritis with rheumatoid factor of unspecified shoulder without organ or systems involvement |
| ICD10CM | M05.72 | Rheumatoid arthritis with rheumatoid factor of elbow without organ or systems involvement |
| ICD10CM | M05.721 | Rheumatoid arthritis with rheumatoid factor of right elbow without organ or systems involvement |
| ICD10CM | M05.722 | Rheumatoid arthritis with rheumatoid factor of left elbow without organ or systems involvement |
| ICD10CM | M05.729 | Rheumatoid arthritis with rheumatoid factor of unspecified elbow without organ or systems involvement |
| ICD10CM | M05.73 | Rheumatoid arthritis with rheumatoid factor of wrist without organ or systems involvement |
| ICD10CM | M05.731 | Rheumatoid arthritis with rheumatoid factor of right wrist without organ or systems involvement |
| ICD10CM | M05.732 | Rheumatoid arthritis with rheumatoid factor of left wrist without organ or systems involvement |
| ICD10CM | M05.739 | Rheumatoid arthritis with rheumatoid factor of unspecified wrist without organ or systems involvement |
| ICD10CM | M05.74 | Rheumatoid arthritis with rheumatoid factor of hand without organ or systems involvement |
| ICD10CM | M05.741 | Rheumatoid arthritis with rheumatoid factor of right hand without organ or systems involvement |
| ICD10CM | M05.742 | Rheumatoid arthritis with rheumatoid factor of left hand without organ or systems involvement |
| ICD10CM | M05.749 | Rheumatoid arthritis with rheumatoid factor of unspecified hand without organ or systems involvement |
| ICD10CM | M05.75 | Rheumatoid arthritis with rheumatoid factor of hip without organ or systems involvement |

Continued on next page

**Table 7 – continued from previous page**

| CodeSystem | ConceptCode | ConceptName |
|---|---|---|
| ICD10CM | M05.751 | Rheumatoid arthritis with rheumatoid factor of right hip without organ or systems involvement |
| ICD10CM | M05.752 | Rheumatoid arthritis with rheumatoid factor of left hip without organ or systems involvement |
| ICD10CM | M05.759 | Rheumatoid arthritis with rheumatoid factor of unspecified hip without organ or systems involvement |
| ICD10CM | M05.76 | Rheumatoid arthritis with rheumatoid factor of knee without organ or systems involvement |
| ICD10CM | M05.761 | Rheumatoid arthritis with rheumatoid factor of right knee without organ or systems involvement |
| ICD10CM | M05.762 | Rheumatoid arthritis with rheumatoid factor of left knee without organ or systems involvement |
| ICD10CM | M05.769 | Rheumatoid arthritis with rheumatoid factor of unspecified knee without organ or systems involvement |
| ICD10CM | M05.77 | Rheumatoid arthritis with rheumatoid factor of ankle and foot without organ or systems involvement |
| ICD10CM | M05.771 | Rheumatoid arthritis with rheumatoid factor of right ankle and foot without organ or systems involvement |
| ICD10CM | M05.772 | Rheumatoid arthritis with rheumatoid factor of left ankle and foot without organ or systems involvement |
| ICD10CM | M05.779 | Rheumatoid arthritis with rheumatoid factor of unspecified ankle and foot without organ or systems involvement |
| ICD10CM | M05.852 | Other rheumatoid arthritis with rheumatoid factor of left hip |
| ICD10CM | M05.79 | Rheumatoid arthritis with rheumatoid factor of multiple sites without organ or systems involvement |
| ICD10CM | M05.8 | Other rheumatoid arthritis with rheumatoid factor |
| ICD10CM | M05.80 | Other rheumatoid arthritis with rheumatoid factor of unspecified site |
| ICD10CM | M05.81 | Other rheumatoid arthritis with rheumatoid factor of shoulder |
| ICD10CM | M05.811 | Other rheumatoid arthritis with rheumatoid factor of right shoulder |
| ICD10CM | M05.812 | Other rheumatoid arthritis with rheumatoid factor of left shoulder |
| ICD10CM | M05.819 | Other rheumatoid arthritis with rheumatoid factor of unspecified shoulder |
| ICD10CM | M05.82 | Other rheumatoid arthritis with rheumatoid factor of elbow |
| ICD10CM | M05.821 | Other rheumatoid arthritis with rheumatoid factor of right elbow |
| ICD10CM | M05.822 | Other rheumatoid arthritis with rheumatoid factor of left elbow |

Continued on next page

**Table 7 – continued from previous page**

| CodeSystem | ConceptCode | ConceptName |
|---|---|---|
| ICD10CM | M05.829 | Other rheumatoid arthritis with rheumatoid factor of unspecified elbow |
| ICD10CM | M05.83 | Other rheumatoid arthritis with rheumatoid factor of wrist |
| ICD10CM | M05.831 | Other rheumatoid arthritis with rheumatoid factor of right wrist |
| ICD10CM | M05.832 | Other rheumatoid arthritis with rheumatoid factor of left wrist |
| ICD10CM | M05.839 | Other rheumatoid arthritis with rheumatoid factor of unspecified wrist |
| ICD10CM | M05.84 | Other rheumatoid arthritis with rheumatoid factor of hand |
| ICD10CM | M05.841 | Other rheumatoid arthritis with rheumatoid factor of right hand |
| ICD10CM | M05.842 | Other rheumatoid arthritis with rheumatoid factor of left hand |
| ICD10CM | M05.849 | Other rheumatoid arthritis with rheumatoid factor of unspecified hand |
| ICD10CM | M05.85 | Other rheumatoid arthritis with rheumatoid factor of hip |
| ICD10CM | M05.851 | Other rheumatoid arthritis with rheumatoid factor of right hip |
| ICD10CM | M05.859 | Other rheumatoid arthritis with rheumatoid factor of unspecified hip |
| ICD10CM | M05.86 | Other rheumatoid arthritis with rheumatoid factor of knee |
| ICD10CM | M05.861 | Other rheumatoid arthritis with rheumatoid factor of right knee |
| ICD10CM | M05.862 | Other rheumatoid arthritis with rheumatoid factor of left knee |
| ICD10CM | M05.869 | Other rheumatoid arthritis with rheumatoid factor of unspecified knee |
| ICD10CM | M05.87 | Other rheumatoid arthritis with rheumatoid factor of ankle and foot |
| ICD10CM | M05.871 | Other rheumatoid arthritis with rheumatoid factor of right ankle and foot |
| ICD10CM | M05.872 | Other rheumatoid arthritis with rheumatoid factor of left ankle and foot |
| ICD10CM | M05.879 | Other rheumatoid arthritis with rheumatoid factor of unspecified ankle and foot |
| ICD10CM | M05.89 | Other rheumatoid arthritis with rheumatoid factor of multiple sites |
| ICD10CM | M05.9 | Rheumatoid arthritis with rheumatoid factor, unspecified |
| ICD10CM | M06.0 | Rheumatoid arthritis without rheumatoid factor |
| ICD10CM | M06.00 | Rheumatoid arthritis without rheumatoid factor, unspecified site |
| ICD10CM | M06.01 | Rheumatoid arthritis without rheumatoid factor, shoulder |

Continued on next page

**Table 7 – continued from previous page**

| CodeSystem | ConceptCode | ConceptName |
|---|---|---|
| ICD10CM | M06.011 | Rheumatoid arthritis without rheumatoid factor, right shoulder |
| ICD10CM | M06.012 | Rheumatoid arthritis without rheumatoid factor, left shoulder |
| ICD10CM | M06.019 | Rheumatoid arthritis without rheumatoid factor, unspecified shoulder |
| ICD10CM | M06.02 | Rheumatoid arthritis without rheumatoid factor, elbow |
| ICD10CM | M06.021 | Rheumatoid arthritis without rheumatoid factor, right elbow |
| ICD10CM | M06.022 | Rheumatoid arthritis without rheumatoid factor, left elbow |
| ICD10CM | M06.029 | Rheumatoid arthritis without rheumatoid factor, unspecified elbow |
| ICD10CM | M06.03 | Rheumatoid arthritis without rheumatoid factor, wrist |
| ICD10CM | M06.031 | Rheumatoid arthritis without rheumatoid factor, right wrist |
| ICD10CM | M06.032 | Rheumatoid arthritis without rheumatoid factor, left wrist |
| ICD10CM | M06.039 | Rheumatoid arthritis without rheumatoid factor, unspecified wrist |
| ICD10CM | M06.04 | Rheumatoid arthritis without rheumatoid factor, hand |
| ICD10CM | M06.041 | Rheumatoid arthritis without rheumatoid factor, right hand |
| ICD10CM | M06.042 | Rheumatoid arthritis without rheumatoid factor, left hand |
| ICD10CM | M06.049 | Rheumatoid arthritis without rheumatoid factor, unspecified hand |
| ICD10CM | M06.05 | Rheumatoid arthritis without rheumatoid factor, hip |
| ICD10CM | M06.051 | Rheumatoid arthritis without rheumatoid factor, right hip |
| ICD10CM | M06.052 | Rheumatoid arthritis without rheumatoid factor, left hip |
| ICD10CM | M06.059 | Rheumatoid arthritis without rheumatoid factor, unspecified hip |
| ICD10CM | M06.06 | Rheumatoid arthritis without rheumatoid factor, knee |
| ICD10CM | M06.061 | Rheumatoid arthritis without rheumatoid factor, right knee |
| ICD10CM | M06.062 | Rheumatoid arthritis without rheumatoid factor, left knee |
| ICD10CM | M06.069 | Rheumatoid arthritis without rheumatoid factor, unspecified knee |
| ICD10CM | M06.07 | Rheumatoid arthritis without rheumatoid factor, ankle and foot |
| ICD10CM | M06.071 | Rheumatoid arthritis without rheumatoid factor, right ankle and foot |
| ICD10CM | M06.072 | Rheumatoid arthritis without rheumatoid factor, left ankle and foot |
| ICD10CM | M06.079 | Rheumatoid arthritis without rheumatoid factor, unspecified ankle and foot |
| ICD10CM | M06.08 | Rheumatoid arthritis without rheumatoid factor, vertebrae |
| ICD10CM | M06.09 | Rheumatoid arthritis without rheumatoid factor, multiple sites |

Continued on next page

**Table 7 – continued from previous page**

| CodeSystem | ConceptCode | ConceptName |
|---|---|---|
| ICD10CM | M06.1 | Adult-onset Still's disease |
| ICD10CM | M06.2 | Rheumatoid bursitis |
| ICD10CM | M06.20 | Rheumatoid bursitis, unspecified site |
| ICD10CM | M06.21 | Rheumatoid bursitis, shoulder |
| ICD10CM | M06.211 | Rheumatoid bursitis, right shoulder |
| ICD10CM | M06.212 | Rheumatoid bursitis, left shoulder |
| ICD10CM | M06.219 | Rheumatoid bursitis, unspecified shoulder |
| ICD10CM | M06.22 | Rheumatoid bursitis, elbow |
| ICD10CM | M06.221 | Rheumatoid bursitis, right elbow |
| ICD10CM | M06.222 | Rheumatoid bursitis, left elbow |
| ICD10CM | M06.229 | Rheumatoid bursitis, unspecified elbow |
| ICD10CM | M06.23 | Rheumatoid bursitis, wrist |
| ICD10CM | M06.231 | Rheumatoid bursitis, right wrist |
| ICD10CM | M06.232 | Rheumatoid bursitis, left wrist |
| ICD10CM | M06.239 | Rheumatoid bursitis, unspecified wrist |
| ICD10CM | M06.24 | Rheumatoid bursitis, hand |
| ICD10CM | M06.241 | Rheumatoid bursitis, right hand |
| ICD10CM | M06.242 | Rheumatoid bursitis, left hand |
| ICD10CM | M06.249 | Rheumatoid bursitis, unspecified hand |
| ICD10CM | M06.25 | Rheumatoid bursitis, hip |
| ICD10CM | M06.251 | Rheumatoid bursitis, right hip |
| ICD10CM | M06.252 | Rheumatoid bursitis, left hip |
| ICD10CM | M06.259 | Rheumatoid bursitis, unspecified hip |
| ICD10CM | M06.26 | Rheumatoid bursitis, knee |
| ICD10CM | M06.261 | Rheumatoid bursitis, right knee |
| ICD10CM | M06.262 | Rheumatoid bursitis, left knee |
| ICD10CM | M06.269 | Rheumatoid bursitis, unspecified knee |
| ICD10CM | M06.27 | Rheumatoid bursitis, ankle and foot |
| ICD10CM | M06.271 | Rheumatoid bursitis, right ankle and foot |
| ICD10CM | M06.272 | Rheumatoid bursitis, left ankle and foot |
| ICD10CM | M06.279 | Rheumatoid bursitis, unspecified ankle and foot |
| ICD10CM | M06.28 | Rheumatoid bursitis, vertebrae |
| ICD10CM | M06.29 | Rheumatoid bursitis, multiple sites |
| ICD10CM | M06.3 | Rheumatoid nodule |
| ICD10CM | M06.30 | Rheumatoid nodule, unspecified site |
| ICD10CM | M06.31 | Rheumatoid nodule, shoulder |
| ICD10CM | M06.311 | Rheumatoid nodule, right shoulder |
| ICD10CM | M06.312 | Rheumatoid nodule, left shoulder |
| ICD10CM | M06.319 | Rheumatoid nodule, unspecified shoulder |
| ICD10CM | M06.32 | Rheumatoid nodule, elbow |
| ICD10CM | M06.321 | Rheumatoid nodule, right elbow |
| ICD10CM | M06.322 | Rheumatoid nodule, left elbow |
| ICD10CM | M06.329 | Rheumatoid nodule, unspecified elbow |

Continued on next page

**Table 7 – continued from previous page**

| CodeSystem | ConceptCode | ConceptName |
|---|---|---|
| ICD10CM | M06.33 | Rheumatoid nodule, wrist |
| ICD10CM | M06.331 | Rheumatoid nodule, right wrist |
| ICD10CM | M06.332 | Rheumatoid nodule, left wrist |
| ICD10CM | M06.339 | Rheumatoid nodule, unspecified wrist |
| ICD10CM | M06.34 | Rheumatoid nodule, hand |
| ICD10CM | M06.341 | Rheumatoid nodule, right hand |
| ICD10CM | M06.342 | Rheumatoid nodule, left hand |
| ICD10CM | M06.349 | Rheumatoid nodule, unspecified hand |
| ICD10CM | M06.35 | Rheumatoid nodule, hip |
| ICD10CM | M06.351 | Rheumatoid nodule, right hip |
| ICD10CM | M06.352 | Rheumatoid nodule, left hip |
| ICD10CM | M06.359 | Rheumatoid nodule, unspecified hip |
| ICD10CM | M06.36 | Rheumatoid nodule, knee |
| ICD10CM | M06.361 | Rheumatoid nodule, right knee |
| ICD10CM | M06.362 | Rheumatoid nodule, left knee |
| ICD10CM | M06.369 | Rheumatoid nodule, unspecified knee |
| ICD10CM | M06.37 | Rheumatoid nodule, ankle and foot |
| ICD10CM | M06.371 | Rheumatoid nodule, right ankle and foot |
| ICD10CM | M06.372 | Rheumatoid nodule, left ankle and foot |
| ICD10CM | M06.379 | Rheumatoid nodule, unspecified ankle and foot |
| ICD10CM | M06.38 | Rheumatoid nodule, vertebrae |
| ICD10CM | M06.39 | Rheumatoid nodule, multiple sites |
| ICD10CM | M06.4 | Inflammatory polyarthropathy |
| ICD10CM | M06.8 | Other specified rheumatoid arthritis |
| ICD10CM | M06.80 | Other specified rheumatoid arthritis, unspecified site |
| ICD10CM | M06.81 | Other specified rheumatoid arthritis, shoulder |
| ICD10CM | M06.811 | Other specified rheumatoid arthritis, right shoulder |
| ICD10CM | M06.812 | Other specified rheumatoid arthritis, left shoulder |
| ICD10CM | M06.819 | Other specified rheumatoid arthritis, unspecified shoulder |
| ICD10CM | M06.82 | Other specified rheumatoid arthritis, elbow |
| ICD10CM | M06.821 | Other specified rheumatoid arthritis, right elbow |
| ICD10CM | M06.822 | Other specified rheumatoid arthritis, left elbow |
| ICD10CM | M06.829 | Other specified rheumatoid arthritis, unspecified elbow |
| ICD10CM | M06.83 | Other specified rheumatoid arthritis, wrist |
| ICD10CM | M06.831 | Other specified rheumatoid arthritis, right wrist |
| ICD10CM | M06.832 | Other specified rheumatoid arthritis, left wrist |
| ICD10CM | M06.839 | Other specified rheumatoid arthritis, unspecified wrist |
| ICD10CM | M06.84 | Other specified rheumatoid arthritis, hand |
| ICD10CM | M06.841 | Other specified rheumatoid arthritis, right hand |
| ICD10CM | M06.842 | Other specified rheumatoid arthritis, left hand |
| ICD10CM | M06.849 | Other specified rheumatoid arthritis, unspecified hand |
| ICD10CM | M06.85 | Other specified rheumatoid arthritis, hip |
| ICD10CM | M06.851 | Other specified rheumatoid arthritis, right hip |

Continued on next page

**Table 7 – continued from previous page**

| CodeSystem | ConceptCode | ConceptName |
|---|---|---|
| ICD10CM | M06.852 | Other specified rheumatoid arthritis, left hip |
| ICD10CM | M06.859 | Other specified rheumatoid arthritis, unspecified hip |
| ICD10CM | M06.86 | Other specified rheumatoid arthritis, knee |
| ICD10CM | M06.861 | Other specified rheumatoid arthritis, right knee |
| ICD10CM | M06.862 | Other specified rheumatoid arthritis, left knee |
| ICD10CM | M06.869 | Other specified rheumatoid arthritis, unspecified knee |
| ICD10CM | M06.87 | Other specified rheumatoid arthritis, ankle and foot |
| ICD10CM | M06.871 | Other specified rheumatoid arthritis, right ankle and foot |
| ICD10CM | M06.872 | Other specified rheumatoid arthritis, left ankle and foot |
| ICD10CM | M06.879 | Other specified rheumatoid arthritis, unspecified ankle and foot |
| ICD10CM | M06.88 | Other specified rheumatoid arthritis, vertebrae |
| ICD10CM | M06.89 | Other specified rheumatoid arthritis, multiple sites |
| ICD10CM | M06.9 | Rheumatoid arthritis, unspecified |
| ICD10CM | M07.6 | Enteropathic arthropathies |
| ICD10CM | M07.60 | Enteropathic arthropathies, unspecified site |
| ICD10CM | M07.61 | Enteropathic arthropathies, shoulder |
| ICD10CM | M07.611 | Enteropathic arthropathies, right shoulder |
| ICD10CM | M07.612 | Enteropathic arthropathies, left shoulder |
| ICD10CM | M07.619 | Enteropathic arthropathies, unspecified shoulder |
| ICD10CM | M07.62 | Enteropathic arthropathies, elbow |
| ICD10CM | M07.621 | Enteropathic arthropathies, right elbow |
| ICD10CM | M07.622 | Enteropathic arthropathies, left elbow |
| ICD10CM | M07.629 | Enteropathic arthropathies, unspecified elbow |
| ICD10CM | M07.63 | Enteropathic arthropathies, wrist |
| ICD10CM | M07.631 | Enteropathic arthropathies, right wrist |
| ICD10CM | M07.632 | Enteropathic arthropathies, left wrist |
| ICD10CM | M07.639 | Enteropathic arthropathies, unspecified wrist |
| ICD10CM | M07.64 | Enteropathic arthropathies, hand |
| ICD10CM | M07.641 | Enteropathic arthropathies, right hand |
| ICD10CM | M07.642 | Enteropathic arthropathies, left hand |
| ICD10CM | M07.649 | Enteropathic arthropathies, unspecified hand |
| ICD10CM | M07.65 | Enteropathic arthropathies, hip |
| ICD10CM | M07.651 | Enteropathic arthropathies, right hip |
| ICD10CM | M07.652 | Enteropathic arthropathies, left hip |
| ICD10CM | M07.659 | Enteropathic arthropathies, unspecified hip |
| ICD10CM | M07.66 | Enteropathic arthropathies, knee |
| ICD10CM | M07.661 | Enteropathic arthropathies, right knee |
| ICD10CM | M07.662 | Enteropathic arthropathies, left knee |
| ICD10CM | M07.669 | Enteropathic arthropathies, unspecified knee |
| ICD10CM | M07.67 | Enteropathic arthropathies, ankle and foot |
| ICD10CM | M07.671 | Enteropathic arthropathies, right ankle and foot |
| ICD10CM | M07.672 | Enteropathic arthropathies, left ankle and foot |

Continued on next page

**Table 7 – continued from previous page**

| CodeSystem | ConceptCode | ConceptName |
|---|---|---|
| ICD10CM | M07.679 | Enteropathic arthropathies, unspecified ankle and foot |
| ICD10CM | M07.68 | Enteropathic arthropathies, vertebrae |
| ICD10CM | M07.69 | Enteropathic arthropathies, multiple sites |
| ICD10CM | M08.0 | Unspecified juvenile rheumatoid arthritis |
| ICD10CM | M08.00 | Unspecified juvenile rheumatoid arthritis of unspecified site |
| ICD10CM | M08.01 | Unspecified juvenile rheumatoid arthritis, shoulder |
| ICD10CM | M08.011 | Unspecified juvenile rheumatoid arthritis, right shoulder |
| ICD10CM | M08.012 | Unspecified juvenile rheumatoid arthritis, left shoulder |
| ICD10CM | M08.019 | Unspecified juvenile rheumatoid arthritis, unspecified shoulder |
| ICD10CM | M08.02 | Unspecified juvenile rheumatoid arthritis of elbow |
| ICD10CM | M08.021 | Unspecified juvenile rheumatoid arthritis, right elbow |
| ICD10CM | M08.022 | Unspecified juvenile rheumatoid arthritis, left elbow |
| ICD10CM | M08.029 | Unspecified juvenile rheumatoid arthritis, unspecified elbow |
| ICD10CM | M08.03 | Unspecified juvenile rheumatoid arthritis, wrist |
| ICD10CM | M08.031 | Unspecified juvenile rheumatoid arthritis, right wrist |
| ICD10CM | M08.032 | Unspecified juvenile rheumatoid arthritis, left wrist |
| ICD10CM | M08.039 | Unspecified juvenile rheumatoid arthritis, unspecified wrist |
| ICD10CM | M08.04 | Unspecified juvenile rheumatoid arthritis, hand |
| ICD10CM | M08.041 | Unspecified juvenile rheumatoid arthritis, right hand |
| ICD10CM | M08.042 | Unspecified juvenile rheumatoid arthritis, left hand |
| ICD10CM | M08.049 | Unspecified juvenile rheumatoid arthritis, unspecified hand |
| ICD10CM | M08.05 | Unspecified juvenile rheumatoid arthritis, hip |
| ICD10CM | M08.051 | Unspecified juvenile rheumatoid arthritis, right hip |
| ICD10CM | M08.052 | Unspecified juvenile rheumatoid arthritis, left hip |
| ICD10CM | M08.059 | Unspecified juvenile rheumatoid arthritis, unspecified hip |
| ICD10CM | M08.06 | Unspecified juvenile rheumatoid arthritis, knee |
| ICD10CM | M08.061 | Unspecified juvenile rheumatoid arthritis, right knee |
| ICD10CM | M08.062 | Unspecified juvenile rheumatoid arthritis, left knee |
| ICD10CM | M08.069 | Unspecified juvenile rheumatoid arthritis, unspecified knee |
| ICD10CM | M08.07 | Unspecified juvenile rheumatoid arthritis, ankle and foot |
| ICD10CM | M08.071 | Unspecified juvenile rheumatoid arthritis, right ankle and foot |
| ICD10CM | M08.072 | Unspecified juvenile rheumatoid arthritis, left ankle and foot |
| ICD10CM | M08.079 | Unspecified juvenile rheumatoid arthritis, unspecified ankle and foot |
| ICD10CM | M08.08 | Unspecified juvenile rheumatoid arthritis, vertebrae |
| ICD10CM | M08.09 | Unspecified juvenile rheumatoid arthritis, multiple sites |
| ICD10CM | M08.1 | Juvenile ankylosing spondylitis |
| ICD10CM | M08.2 | Juvenile rheumatoid arthritis with systemic onset |
| ICD10CM | M08.20 | Juvenile rheumatoid arthritis with systemic onset, unspecified site |

Continued on next page

**Table 7 – continued from previous page**

| CodeSystem | ConceptCode | ConceptName |
|---|---|---|
| ICD10CM | M08.21 | Juvenile rheumatoid arthritis with systemic onset, shoulder |
| ICD10CM | M08.211 | Juvenile rheumatoid arthritis with systemic onset, right shoulder |
| ICD10CM | M08.212 | Juvenile rheumatoid arthritis with systemic onset, left shoulder |
| ICD10CM | M08.219 | Juvenile rheumatoid arthritis with systemic onset, unspecified shoulder |
| ICD10CM | M08.22 | Juvenile rheumatoid arthritis with systemic onset, elbow |
| ICD10CM | M08.221 | Juvenile rheumatoid arthritis with systemic onset, right elbow |
| ICD10CM | M08.222 | Juvenile rheumatoid arthritis with systemic onset, left elbow |
| ICD10CM | M08.229 | Juvenile rheumatoid arthritis with systemic onset, unspecified elbow |
| ICD10CM | M08.23 | Juvenile rheumatoid arthritis with systemic onset, wrist |
| ICD10CM | M08.231 | Juvenile rheumatoid arthritis with systemic onset, right wrist |
| ICD10CM | M08.232 | Juvenile rheumatoid arthritis with systemic onset, left wrist |
| ICD10CM | M08.239 | Juvenile rheumatoid arthritis with systemic onset, unspecified wrist |
| ICD10CM | M08.24 | Juvenile rheumatoid arthritis with systemic onset, hand |
| ICD10CM | M08.241 | Juvenile rheumatoid arthritis with systemic onset, right hand |
| ICD10CM | M08.242 | Juvenile rheumatoid arthritis with systemic onset, left hand |
| ICD10CM | M08.249 | Juvenile rheumatoid arthritis with systemic onset, unspecified hand |
| ICD10CM | M08.25 | Juvenile rheumatoid arthritis with systemic onset, hip |
| ICD10CM | M08.251 | Juvenile rheumatoid arthritis with systemic onset, right hip |
| ICD10CM | M08.252 | Juvenile rheumatoid arthritis with systemic onset, left hip |
| ICD10CM | M08.259 | Juvenile rheumatoid arthritis with systemic onset, unspecified hip |
| ICD10CM | M08.26 | Juvenile rheumatoid arthritis with systemic onset, knee |
| ICD10CM | M08.261 | Juvenile rheumatoid arthritis with systemic onset, right knee |
| ICD10CM | M08.262 | Juvenile rheumatoid arthritis with systemic onset, left knee |
| ICD10CM | M08.269 | Juvenile rheumatoid arthritis with systemic onset, unspecified knee |
| ICD10CM | M08.27 | Juvenile rheumatoid arthritis with systemic onset, ankle and foot |
| ICD10CM | M08.271 | Juvenile rheumatoid arthritis with systemic onset, right ankle and foot |
| ICD10CM | M08.272 | Juvenile rheumatoid arthritis with systemic onset, left ankle and foot |

Continued on next page

**Table 7 – continued from previous page**

| CodeSystem | ConceptCode | ConceptName |
|---|---|---|
| ICD10CM | M08.279 | Juvenile rheumatoid arthritis with systemic onset, unspecified ankle and foot |
| ICD10CM | M08.28 | Juvenile rheumatoid arthritis with systemic onset, vertebrae |
| ICD10CM | M08.29 | Juvenile rheumatoid arthritis with systemic onset, multiple sites |
| ICD10CM | M08.3 | Juvenile rheumatoid polyarthritis (seronegative) |
| ICD10CM | M08.4 | Pauciarticular juvenile rheumatoid arthritis |
| ICD10CM | M08.40 | Pauciarticular juvenile rheumatoid arthritis, unspecified site |
| ICD10CM | M08.41 | Pauciarticular juvenile rheumatoid arthritis, shoulder |
| ICD10CM | M08.411 | Pauciarticular juvenile rheumatoid arthritis, right shoulder |
| ICD10CM | M08.412 | Pauciarticular juvenile rheumatoid arthritis, left shoulder |
| ICD10CM | M08.419 | Pauciarticular juvenile rheumatoid arthritis, unspecified shoulder |
| ICD10CM | M08.42 | Pauciarticular juvenile rheumatoid arthritis, elbow |
| ICD10CM | M08.421 | Pauciarticular juvenile rheumatoid arthritis, right elbow |
| ICD10CM | M08.422 | Pauciarticular juvenile rheumatoid arthritis, left elbow |
| ICD10CM | M08.429 | Pauciarticular juvenile rheumatoid arthritis, unspecified elbow |
| ICD10CM | M08.43 | Pauciarticular juvenile rheumatoid arthritis, wrist |
| ICD10CM | M08.431 | Pauciarticular juvenile rheumatoid arthritis, right wrist |
| ICD10CM | M08.432 | Pauciarticular juvenile rheumatoid arthritis, left wrist |
| ICD10CM | M08.439 | Pauciarticular juvenile rheumatoid arthritis, unspecified wrist |
| ICD10CM | M08.44 | Pauciarticular juvenile rheumatoid arthritis, hand |
| ICD10CM | M08.441 | Pauciarticular juvenile rheumatoid arthritis, right hand |
| ICD10CM | M08.442 | Pauciarticular juvenile rheumatoid arthritis, left hand |
| ICD10CM | M08.449 | Pauciarticular juvenile rheumatoid arthritis, unspecified hand |
| ICD10CM | M08.45 | Pauciarticular juvenile rheumatoid arthritis, hip |
| ICD10CM | M08.451 | Pauciarticular juvenile rheumatoid arthritis, right hip |
| ICD10CM | M08.452 | Pauciarticular juvenile rheumatoid arthritis, left hip |
| ICD10CM | M08.459 | Pauciarticular juvenile rheumatoid arthritis, unspecified hip |
| ICD10CM | M08.46 | Pauciarticular juvenile rheumatoid arthritis, knee |
| ICD10CM | M08.461 | Pauciarticular juvenile rheumatoid arthritis, right knee |
| ICD10CM | M08.462 | Pauciarticular juvenile rheumatoid arthritis, left knee |
| ICD10CM | M08.469 | Pauciarticular juvenile rheumatoid arthritis, unspecified knee |
| ICD10CM | M08.47 | Pauciarticular juvenile rheumatoid arthritis, ankle and foot |
| ICD10CM | M08.471 | Pauciarticular juvenile rheumatoid arthritis, right ankle and foot |
| ICD10CM | M08.472 | Pauciarticular juvenile rheumatoid arthritis, left ankle and foot |

Continued on next page

Table 7 – continued from previous page

| CodeSystem | ConceptCode | ConceptName |
|---|---|---|
| ICD10CM | M08.479 | Pauciarticular juvenile rheumatoid arthritis, unspecified ankle and foot |
| ICD10CM | M08.48 | Pauciarticular juvenile rheumatoid arthritis, vertebrae |
| ICD10CM | M08.8 | Other juvenile arthritis |
| ICD10CM | M08.80 | Other juvenile arthritis, unspecified site |
| ICD10CM | M08.81 | Other juvenile arthritis, shoulder |
| ICD10CM | M08.811 | Other juvenile arthritis, right shoulder |
| ICD10CM | M08.812 | Other juvenile arthritis, left shoulder |
| ICD10CM | M08.819 | Other juvenile arthritis, unspecified shoulder |
| ICD10CM | M08.82 | Other juvenile arthritis, elbow |
| ICD10CM | M08.821 | Other juvenile arthritis, right elbow |
| ICD10CM | M08.822 | Other juvenile arthritis, left elbow |
| ICD10CM | M08.829 | Other juvenile arthritis, unspecified elbow |
| ICD10CM | M08.83 | Other juvenile arthritis, wrist |
| ICD10CM | M08.831 | Other juvenile arthritis, right wrist |
| ICD10CM | M08.832 | Other juvenile arthritis, left wrist |
| ICD10CM | M08.839 | Other juvenile arthritis, unspecified wrist |
| ICD10CM | M08.84 | Other juvenile arthritis, hand |
| ICD10CM | M08.841 | Other juvenile arthritis, right hand |
| ICD10CM | M08.842 | Other juvenile arthritis, left hand |
| ICD10CM | M08.849 | Other juvenile arthritis, unspecified hand |
| ICD10CM | M08.85 | Other juvenile arthritis, hip |
| ICD10CM | M08.851 | Other juvenile arthritis, right hip |
| ICD10CM | M08.852 | Other juvenile arthritis, left hip |
| ICD10CM | M08.859 | Other juvenile arthritis, unspecified hip |
| ICD10CM | M08.86 | Other juvenile arthritis, knee |
| ICD10CM | M08.861 | Other juvenile arthritis, right knee |
| ICD10CM | M08.862 | Other juvenile arthritis, left knee |
| ICD10CM | M08.869 | Other juvenile arthritis, unspecified knee |
| ICD10CM | M08.87 | Other juvenile arthritis, ankle and foot |
| ICD10CM | M08.871 | Other juvenile arthritis, right ankle and foot |
| ICD10CM | M08.872 | Other juvenile arthritis, left ankle and foot |
| ICD10CM | M08.879 | Other juvenile arthritis, unspecified ankle and foot |
| ICD10CM | M08.88 | Other juvenile arthritis, other specified site |
| ICD10CM | M08.89 | Other juvenile arthritis, multiple sites |
| ICD10CM | M08.9 | Juvenile arthritis, unspecified |
| ICD10CM | M08.90 | Juvenile arthritis, unspecified, unspecified site |
| ICD10CM | M08.91 | Juvenile arthritis, unspecified, shoulder |
| ICD10CM | M08.911 | Juvenile arthritis, unspecified, right shoulder |
| ICD10CM | M08.912 | Juvenile arthritis, unspecified, left shoulder |
| ICD10CM | M08.919 | Juvenile arthritis, unspecified, unspecified shoulder |
| ICD10CM | M08.92 | Juvenile arthritis, unspecified, elbow |
| ICD10CM | M08.921 | Juvenile arthritis, unspecified, right elbow |

Continued on next page

**Table 7 – continued from previous page**

| CodeSystem | ConceptCode | ConceptName |
|---|---|---|
| ICD10CM | M08.922 | Juvenile arthritis, unspecified, left elbow |
| ICD10CM | M08.929 | Juvenile arthritis, unspecified, unspecified elbow |
| ICD10CM | M08.93 | Juvenile arthritis, unspecified, wrist |
| ICD10CM | M08.931 | Juvenile arthritis, unspecified, right wrist |
| ICD10CM | M08.932 | Juvenile arthritis, unspecified, left wrist |
| ICD10CM | M08.939 | Juvenile arthritis, unspecified, unspecified wrist |
| ICD10CM | M08.94 | Juvenile arthritis, unspecified, hand |
| ICD10CM | M08.941 | Juvenile arthritis, unspecified, right hand |
| ICD10CM | M08.942 | Juvenile arthritis, unspecified, left hand |
| ICD10CM | M08.949 | Juvenile arthritis, unspecified, unspecified hand |
| ICD10CM | M08.95 | Juvenile arthritis, unspecified, hip |
| ICD10CM | M08.951 | Juvenile arthritis, unspecified, right hip |
| ICD10CM | M08.952 | Juvenile arthritis, unspecified, left hip |
| ICD10CM | M08.959 | Juvenile arthritis, unspecified, unspecified hip |
| ICD10CM | M08.96 | Juvenile arthritis, unspecified, knee |
| ICD10CM | M08.961 | Juvenile arthritis, unspecified, right knee |
| ICD10CM | M08.962 | Juvenile arthritis, unspecified, left knee |
| ICD10CM | M08.969 | Juvenile arthritis, unspecified, unspecified knee |
| ICD10CM | M08.97 | Juvenile arthritis, unspecified, ankle and foot |
| ICD10CM | M08.971 | Juvenile arthritis, unspecified, right ankle and foot |
| ICD10CM | M08.972 | Juvenile arthritis, unspecified, left ankle and foot |
| ICD10CM | M08.979 | Juvenile arthritis, unspecified, unspecified ankle and foot |
| ICD10CM | M08.98 | Juvenile arthritis, unspecified, vertebrae |
| ICD10CM | M08.99 | Juvenile arthritis, unspecified, multiple sites |
| ICD10CM | M30.0 | Polyarteritis nodosa |
| ICD10CM | M30.1 | Polyarteritis with lung involvement [Churg-Strauss] |
| ICD10CM | M30.2 | Juvenile polyarteritis |
| ICD10CM | M30.3 | Mucocutaneous lymph node syndrome [Kawasaki] |
| ICD10CM | M30.8 | Other conditions related to polyarteritis nodosa |
| ICD10CM | M31.30 | Wegener's granulomatosis without renal involvement |
| ICD10CM | M31.31 | Wegener's granulomatosis with renal involvement |
| ICD10CM | M32.0 | Drug-induced systemic lupus erythematosus |
| ICD10CM | M32.1 | Systemic lupus erythematosus with organ or system involvement |
| ICD10CM | M32.10 | Systemic lupus erythematosus, organ or system involvement unspecified |
| ICD10CM | M32.11 | Endocarditis in systemic lupus erythematosus |
| ICD10CM | M32.12 | Pericarditis in systemic lupus erythematosus |
| ICD10CM | M32.13 | Lung involvement in systemic lupus erythematosus |
| ICD10CM | M32.14 | Glomerular disease in systemic lupus erythematosus |
| ICD10CM | M32.15 | Tubulo-interstitial nephropathy in systemic lupus erythematosus |

Continued on next page

**Table 7 – continued from previous page**

| CodeSystem | ConceptCode | ConceptName |
|---|---|---|
| ICD10CM | M32.19 | Other organ or system involvement in systemic lupus erythematosus |
| ICD10CM | M32.8 | Other forms of systemic lupus erythematosus |
| ICD10CM | M32.9 | Systemic lupus erythematosus, unspecified |
| ICD10CM | M33.0 | Juvenile dermatomyositis |
| ICD10CM | M33.00 | Juvenile dermatomyositis, organ involvement unspecified |
| ICD10CM | M33.01 | Juvenile dermatomyositis with respiratory involvement |
| ICD10CM | M33.02 | Juvenile dermatomyositis with myopathy |
| ICD10CM | M33.03 | Juvenile dermatomyositis without myopathy |
| ICD10CM | M33.09 | Juvenile dermatomyositis with other organ involvement |
| ICD10CM | M33.1 | Other dermatomyositis |
| ICD10CM | M33.10 | Other dermatomyositis, organ involvement unspecified |
| ICD10CM | M33.11 | Other dermatomyositis with respiratory involvement |
| ICD10CM | M33.12 | Other dermatomyositis with myopathy |
| ICD10CM | M33.13 | Other dermatomyositis without myopathy |
| ICD10CM | M33.19 | Other dermatomyositis with other organ involvement |
| ICD10CM | M33.2 | Polymyositis |
| ICD10CM | M33.20 | Polymyositis, organ involvement unspecified |
| ICD10CM | M33.21 | Polymyositis with respiratory involvement |
| ICD10CM | M33.22 | Polymyositis with myopathy |
| ICD10CM | M33.29 | Polymyositis with other organ involvement |
| ICD10CM | M33.9 | Dermatopolymyositis, unspecified |
| ICD10CM | M33.90 | Dermatopolymyositis, unspecified, organ involvement unspecified |
| ICD10CM | M33.91 | Dermatopolymyositis, unspecified with respiratory involvement |
| ICD10CM | M33.92 | Dermatopolymyositis, unspecified with myopathy |
| ICD10CM | M33.93 | Dermatopolymyositis, unspecified without myopathy |
| ICD10CM | M33.99 | Dermatopolymyositis, unspecified with other organ involvement |
| ICD10CM | M34.0 | Progressive systemic sclerosis |
| ICD10CM | M34.1 | CR(E)ST syndrome |
| ICD10CM | M34.2 | Systemic sclerosis induced by drug and chemical |
| ICD10CM | M34.8 | Other forms of systemic sclerosis |
| ICD10CM | M34.81 | Systemic sclerosis with lung involvement |
| ICD10CM | M34.82 | Systemic sclerosis with myopathy |
| ICD10CM | M34.83 | Systemic sclerosis with polyneuropathy |
| ICD10CM | M34.89 | Other systemic sclerosis |
| ICD10CM | M34.9 | Systemic sclerosis, unspecified |
| ICD10CM | M46.0 | Spinal enthesopathy |
| ICD10CM | M46.00 | Spinal enthesopathy, site unspecified |
| ICD10CM | M46.01 | Spinal enthesopathy, occipito-atlanto-axial region |
| ICD10CM | M46.02 | Spinal enthesopathy, cervical region |

Continued on next page

**Table 7 – continued from previous page**

| CodeSystem | ConceptCode | ConceptName |
|---|---|---|
| ICD10CM | M46.03 | Spinal enthesopathy, cervicothoracic region |
| ICD10CM | M46.04 | Spinal enthesopathy, thoracic region |
| ICD10CM | M46.05 | Spinal enthesopathy, thoracolumbar region |
| ICD10CM | M46.06 | Spinal enthesopathy, lumbar region |
| ICD10CM | M46.07 | Spinal enthesopathy, lumbosacral region |
| ICD10CM | M46.08 | Spinal enthesopathy, sacral and sacrococcygeal region |
| ICD10CM | M46.09 | Spinal enthesopathy, multiple sites in spine |
| ICD10CM | M46.1 | Sacroiliitis, not elsewhere classified |
| ICD10CM | M46.2 | Osteomyelitis of vertebra |
| ICD10CM | M46.20 | Osteomyelitis of vertebra, site unspecified |
| ICD10CM | M46.21 | Osteomyelitis of vertebra, occipito-atlanto-axial region |
| ICD10CM | M46.22 | Osteomyelitis of vertebra, cervical region |
| ICD10CM | M46.23 | Osteomyelitis of vertebra, cervicothoracic region |
| ICD10CM | M46.24 | Osteomyelitis of vertebra, thoracic region |
| ICD10CM | M46.25 | Osteomyelitis of vertebra, thoracolumbar region |
| ICD10CM | M46.26 | Osteomyelitis of vertebra, lumbar region |
| ICD10CM | M46.27 | Osteomyelitis of vertebra, lumbosacral region |
| ICD10CM | M46.28 | Osteomyelitis of vertebra, sacral and sacrococcygeal region |
| ICD10CM | M46.3 | Infection of intervertebral disc (pyogenic) |
| ICD10CM | M46.30 | Infection of intervertebral disc (pyogenic), site unspecified |
| ICD10CM | M46.31 | Infection of intervertebral disc (pyogenic), occipito-atlanto-axial region |
| ICD10CM | M46.32 | Infection of intervertebral disc (pyogenic), cervical region |
| ICD10CM | M46.33 | Infection of intervertebral disc (pyogenic), cervicothoracic region |
| ICD10CM | M46.34 | Infection of intervertebral disc (pyogenic), thoracic region |
| ICD10CM | M46.35 | Infection of intervertebral disc (pyogenic), thoracolumbar region |
| ICD10CM | M46.36 | Infection of intervertebral disc (pyogenic), lumbar region |
| ICD10CM | M46.37 | Infection of intervertebral disc (pyogenic), lumbosacral region |
| ICD10CM | M46.38 | Infection of intervertebral disc (pyogenic), sacral and sacrococcygeal region |
| ICD10CM | M46.39 | Infection of intervertebral disc (pyogenic), multiple sites in spine |
| ICD10CM | M46.4 | Discitis, unspecified |
| ICD10CM | M46.40 | Discitis, unspecified, site unspecified |
| ICD10CM | M46.41 | Discitis, unspecified, occipito-atlanto-axial region |
| ICD10CM | M46.42 | Discitis, unspecified, cervical region |
| ICD10CM | M46.43 | Discitis, unspecified, cervicothoracic region |
| ICD10CM | M46.44 | Discitis, unspecified, thoracic region |
| ICD10CM | M46.45 | Discitis, unspecified, thoracolumbar region |
| ICD10CM | M46.46 | Discitis, unspecified, lumbar region |

Continued on next page

**Table 7 – continued from previous page**

| CodeSystem | ConceptCode | ConceptName |
|---|---|---|
| ICD10CM | M46.47 | Discitis, unspecified, lumbosacral region |
| ICD10CM | M46.48 | Discitis, unspecified, sacral and sacrococcygeal region |
| ICD10CM | M46.49 | Discitis, unspecified, multiple sites in spine |
| ICD10CM | M46.5 | Other infective spondylopathies |
| ICD10CM | M46.50 | Other infective spondylopathies, site unspecified |
| ICD10CM | M46.51 | Other infective spondylopathies, occipito-atlanto-axial region |
| ICD10CM | M46.52 | Other infective spondylopathies, cervical region |
| ICD10CM | M46.53 | Other infective spondylopathies, cervicothoracic region |
| ICD10CM | M46.54 | Other infective spondylopathies, thoracic region |
| ICD10CM | M46.55 | Other infective spondylopathies, thoracolumbar region |
| ICD10CM | M46.56 | Other infective spondylopathies, lumbar region |
| ICD10CM | M46.57 | Other infective spondylopathies, lumbosacral region |
| ICD10CM | M46.58 | Other infective spondylopathies, sacral and sacrococcygeal region |
| ICD10CM | M46.59 | Other infective spondylopathies, multiple sites in spine |
| ICD10CM | M46.8 | Other specified inflammatory spondylopathies |
| ICD10CM | M46.80 | Other specified inflammatory spondylopathies, site unspecified |
| ICD10CM | M46.81 | Other specified inflammatory spondylopathies, occipito-atlanto-axial region |
| ICD10CM | M46.82 | Other specified inflammatory spondylopathies, cervical region |
| ICD10CM | M46.83 | Other specified inflammatory spondylopathies, cervicothoracic region |
| ICD10CM | M46.84 | Other specified inflammatory spondylopathies, thoracic region |
| ICD10CM | M46.85 | Other specified inflammatory spondylopathies, thoracolumbar region |
| ICD10CM | M46.86 | Other specified inflammatory spondylopathies, lumbar region |
| ICD10CM | M46.87 | Other specified inflammatory spondylopathies, lumbosacral region |
| ICD10CM | M46.88 | Other specified inflammatory spondylopathies, sacral and sacrococcygeal region |
| ICD10CM | M46.89 | Other specified inflammatory spondylopathies, multiple sites in spine |
| ICD10CM | M46.9 | Unspecified inflammatory spondylopathy |
| ICD10CM | M46.90 | Unspecified inflammatory spondylopathy, site unspecified |
| ICD10CM | M46.91 | Unspecified inflammatory spondylopathy, occipito-atlanto-axial region |
| ICD10CM | M46.92 | Unspecified inflammatory spondylopathy, cervical region |

Continued on next page

**Table 7 – continued from previous page**

| CodeSystem | ConceptCode | ConceptName |
|---|---|---|
| ICD10CM | M46.93 | Unspecified inflammatory spondylopathy, cervicothoracic region |
| ICD10CM | M46.94 | Unspecified inflammatory spondylopathy, thoracic region |
| ICD10CM | M46.95 | Unspecified inflammatory spondylopathy, thoracolumbar region |
| ICD10CM | M46.96 | Unspecified inflammatory spondylopathy, lumbar region |
| ICD10CM | M46.97 | Unspecified inflammatory spondylopathy, lumbosacral region |
| ICD10CM | M46.98 | Unspecified inflammatory spondylopathy, sacral and sacrococcygeal region |
| ICD10CM | M46.99 | Unspecified inflammatory spondylopathy, multiple sites in spine |
| ICD10CM | T78.40XA | Allergy, unspecified, initial encounter |
| ICD10CM | T78.40XD | Allergy, unspecified, subsequent encounter |
| ICD10CM | T78.40XS | Allergy, unspecified, sequela |
| ICD10CM | T86.30 | Unspecified complication of heart-lung transplant |
| ICD10CM | T86.3 | Complications of heart-lung transplant |
| ICD10CM | T86.0 | Complications of bone marrow transplant |
| ICD10CM | T86.00 | Unspecified complication of bone marrow transplant |
| ICD10CM | T86.01 | Bone marrow transplant rejection |
| ICD10CM | T86.02 | Bone marrow transplant failure |
| ICD10CM | T86.03 | Bone marrow transplant infection |
| ICD10CM | T86.09 | Other complications of bone marrow transplant |
| ICD10CM | T86.1 | Complications of kidney transplant |
| ICD10CM | T86.10 | Unspecified complication of kidney transplant |
| ICD10CM | T86.11 | Kidney transplant rejection |
| ICD10CM | T86.12 | Kidney transplant failure |
| ICD10CM | T86.13 | Kidney transplant infection |
| ICD10CM | T86.19 | Other complication of kidney transplant |
| ICD10CM | T86.2 | Complications of heart transplant |
| ICD10CM | T86.20 | Unspecified complication of heart transplant |
| ICD10CM | T86.21 | Heart transplant rejection |
| ICD10CM | T86.22 | Heart transplant failure |
| ICD10CM | T86.23 | Heart transplant infection |
| ICD10CM | T86.29 | Other complications of heart transplant |
| ICD10CM | T86.290 | Cardiac allograft vasculopathy |
| ICD10CM | T86.298 | Other complications of heart transplant |
| ICD10CM | T86.31 | Heart-lung transplant rejection |
| ICD10CM | T86.32 | Heart-lung transplant failure |
| ICD10CM | T86.33 | Heart-lung transplant infection |
| ICD10CM | T86.39 | Other complications of heart-lung transplant |
| ICD10CM | T86.4 | Complications of liver transplant |
| ICD10CM | T86.40 | Unspecified complication of liver transplant |

Continued on next page

**Table 7 – continued from previous page**

| CodeSystem | ConceptCode | ConceptName |
|---|---|---|
| ICD10CM | T86.41 | Liver transplant rejection |
| ICD10CM | T86.42 | Liver transplant failure |
| ICD10CM | T86.43 | Liver transplant infection |
| ICD10CM | T86.49 | Other complications of liver transplant |
| ICD10CM | T86.5 | Complications of stem cell transplant |
| ICD10CM | T86.8 | Complications of other transplanted organs and tissues |
| ICD10CM | T86.81 | Complications of lung transplant |
| ICD10CM | T86.810 | Lung transplant rejection |
| ICD10CM | T86.811 | Lung transplant failure |
| ICD10CM | T86.812 | Lung transplant infection |
| ICD10CM | T86.818 | Other complications of lung transplant |
| ICD10CM | T86.819 | Unspecified complication of lung transplant |
| ICD10CM | T86.82 | Complications of skin graft (allograft) (autograft) |
| ICD10CM | T86.820 | Skin graft (allograft) rejection |
| ICD10CM | T86.821 | Skin graft (allograft) (autograft) failure |
| ICD10CM | T86.822 | Skin graft (allograft) (autograft) infection |
| ICD10CM | T86.828 | Other complications of skin graft (allograft) (autograft) |
| ICD10CM | T86.829 | Unspecified complication of skin graft (allograft) (autograft) |
| ICD10CM | T86.83 | Complications of bone graft |
| ICD10CM | T86.830 | Bone graft rejection |
| ICD10CM | T86.831 | Bone graft failure |
| ICD10CM | T86.832 | Bone graft infection |
| ICD10CM | T86.838 | Other complications of bone graft |
| ICD10CM | T86.839 | Unspecified complication of bone graft |
| ICD10CM | T86.84 | Complications of corneal transplant |
| ICD10CM | T86.840 | Corneal transplant rejection |
| ICD10CM | T86.841 | Corneal transplant failure |
| ICD10CM | T86.842 | Corneal transplant infection |
| ICD10CM | T86.848 | Other complications of corneal transplant |
| ICD10CM | T86.849 | Unspecified complication of corneal transplant |
| ICD10CM | T86.85 | Complication of intestine transplant |
| ICD10CM | T86.850 | Intestine transplant rejection |
| ICD10CM | T86.851 | Intestine transplant failure |
| ICD10CM | T86.852 | Intestine transplant infection |
| ICD10CM | T86.858 | Other complications of intestine transplant |
| ICD10CM | T86.859 | Unspecified complication of intestine transplant |
| ICD10CM | T86.89 | Complications of other transplanted tissue |
| ICD10CM | T86.890 | Other transplanted tissue rejection |
| ICD10CM | T86.891 | Other transplanted tissue failure |
| ICD10CM | T86.892 | Other transplanted tissue infection |
| ICD10CM | T86.898 | Other complications of other transplanted tissue |
| ICD10CM | T86.899 | Unspecified complication of other transplanted tissue |
| ICD10CM | T86.9 | Complication of unspecified transplanted organ and tissue |

Continued on next page

**Table 7 – continued from previous page**

| CodeSystem | ConceptCode | ConceptName |
|---|---|---|
| ICD10CM | T86.90 | Unspecified complication of unspecified transplanted organ and tissue |
| ICD10CM | T86.91 | Unspecified transplanted organ and tissue rejection |
| ICD10CM | T86.92 | Unspecified transplanted organ and tissue failure |
| ICD10CM | T86.93 | Unspecified transplanted organ and tissue infection |
| ICD10CM | T86.99 | Other complications of unspecified transplanted organ and tissue |
| ICD10CM | Z48.21 | Encounter for aftercare following heart transplant |
| ICD10CM | Z48.22 | Encounter for aftercare following kidney transplant |
| ICD10CM | Z48.23 | Encounter for aftercare following liver transplant |
| ICD10CM | Z48.24 | Encounter for aftercare following lung transplant |
| ICD10CM | Z48.28 | Encounter for aftercare following multiple organ transplant |
| ICD10CM | Z48.280 | Encounter for aftercare following heart-lung transplant |
| ICD10CM | Z48.288 | Encounter for aftercare following multiple organ transplant |
| ICD10CM | Z48.29 | Encounter for aftercare following other organ transplant |
| ICD10CM | Z48.290 | Encounter for aftercare following bone marrow transplant |
| ICD10CM | Z48.298 | Encounter for aftercare following other organ transplant |
| ICD10CM | Z51.11 | Encounter for antineoplastic chemotherapy |
| ICD10CM | Z51.12 | Encounter for antineoplastic immunotherapy |
| ICD10CM | Z94.0 | Kidney transplant status |
| ICD10CM | Z94.1 | Heart transplant status |
| ICD10CM | Z94.2 | Lung transplant status |
| ICD10CM | Z94.3 | Heart and lungs transplant status |
| ICD10CM | Z94.4 | Liver transplant status |
| ICD10CM | Z94.5 | Skin transplant status |
| ICD10CM | Z94.6 | Bone transplant status |
| ICD10CM | Z94.7 | Corneal transplant status |
| ICD10CM | Z94.8 | Other transplanted organ and tissue status |
| ICD10CM | Z94.81 | Bone marrow transplant status |
| ICD10CM | Z94.82 | Intestine transplant status |
| ICD10CM | Z94.83 | Pancreas transplant status |
| ICD10CM | Z94.84 | Stem cells transplant status |
| ICD10CM | Z94.89 | Other transplanted organ and tissue status |
| ICD10CM | Z94.9 | Transplanted organ and tissue status, unspecified |
| ICD10CM | D84.81 | Immunodeficiency due to conditions classified elsewhere |
| ICD10CM | D84.82 | Immunodeficiency due to drugs and external causes |
| ICD10CM | D84.821 | Immunodeficiency due to drugs |
| ICD10CM | D84.822 | Immunodeficiency due to external causes |
| ICD10CM | D84.89 | Other immunodeficiencies |
| ICD10CM | D89.83 | Cytokine release syndrome |
| ICD10CM | D89.831 | Cytokine release syndrome, grade 1 |
| ICD10CM | D89.832 | Cytokine release syndrome, grade 2 |
| ICD10CM | D89.833 | Cytokine release syndrome, grade 3 |

Continued on next page

**Table 7 – continued from previous page**

| CodeSystem | ConceptCode | ConceptName |
|---|---|---|
| ICD10CM | D89.834 | Cytokine release syndrome, grade 4 |
| ICD10CM | D89.835 | Cytokine release syndrome, grade 5 |
| ICD10CM | D89.839 | Cytokine release syndrome, grade unspecified |
| ICD10CM | M05.7A | Rheumatoid arthritis with rheumatoid factor of other specified site without organ or systems involvement |
| ICD10CM | M05.8A | Other rheumatoid arthritis with rheumatoid factor of other specified site |
| ICD10CM | M06.0A | Rheumatoid arthritis without rheumatoid factor, other specified site |
| ICD10CM | M06.8A | Other specified rheumatoid arthritis, other specified site |
| ICD10CM | M08.0A | Unspecified juvenile rheumatoid arthritis, other specified site |
| ICD10CM | M08.2A | Juvenile rheumatoid arthritis with systemic onset, other specified site |
| ICD10CM | M08.4A | Pauciarticular juvenile rheumatoid arthritis, other specified site |
| ICD10CM | M08.9A | Juvenile arthritis, unspecified, other specified site |
| ICD10CM | M35.81 | Multisystem inflammatory syndrome (MIS) |
| ICD10CM | M35.89 | Other specified systemic involvement of connective tissue |
| ICD10CM | T86.8401 | Corneal transplant rejection, right eye |
| ICD10CM | T86.8402 | Corneal transplant rejection, left eye |
| ICD10CM | T86.8403 | Corneal transplant rejection, bilateral |
| ICD10CM | T86.8409 | Corneal transplant rejection, unspecified eye |
| ICD10CM | T86.8411 | Corneal transplant failure, right eye |
| ICD10CM | T86.8412 | Corneal transplant failure, left eye |
| ICD10CM | T86.8413 | Corneal transplant failure, bilateral |
| ICD10CM | T86.8419 | Corneal transplant failure, unspecified eye |
| ICD10CM | T86.8421 | Corneal transplant infection, right eye |
| ICD10CM | T86.8422 | Corneal transplant infection, left eye |
| ICD10CM | T86.8423 | Corneal transplant infection, bilateral |
| ICD10CM | T86.8429 | Corneal transplant infection, unspecified eye |
| ICD10CM | T86.8481 | Other complications of corneal transplant, right eye |
| ICD10CM | T86.8482 | Other complications of corneal transplant, left eye |
| ICD10CM | T86.8483 | Other complications of corneal transplant, bilateral |
| ICD10CM | T86.8489 | Other complications of corneal transplant, unspecified eye |
| ICD10CM | T86.8491 | Unspecified complication of corneal transplant, right eye |
| ICD10CM | T86.8492 | Unspecified complication of corneal transplant, left eye |
| ICD10CM | T86.8493 | Unspecified complication of corneal transplant, bilateral |
| ICD10CM | T86.8499 | Unspecified complication of corneal transplant, unspecified eye |
| ICD10CM | C79.63 | Secondary malignant neoplasm of bilateral ovaries |
| ICD10CM | D89.44 | Hereditary alpha tryptasemia |
| ICD10CM | C84.7A | Anaplastic large cell lymphoma, ALK-negative, breast |

Continued on next page

**Table 7 – continued from previous page**

| CodeSystem | ConceptCode | ConceptName |
|---|---|---|
| ICD10CM | C56.3 | Malignant neoplasm of bilateral ovaries |
| SNOMED CT | 835009 | Angioimmunoblastic T-cell lymphoma |
| SNOMED CT | 3239007 | Lymphocyte disorder |
| SNOMED CT | 3439009 | SCID due to absent peripheral T cell maturation |
| SNOMED CT | 183005 | Autoimmune pancytopenia |
| SNOMED CT | 3902000 | Non dose-related drug-induced neutropenia |
| SNOMED CT | 4434006 | Bloom syndrome |
| SNOMED CT | 4676006 | SLE glomerulonephritis syndrome, WHO class II |
| SNOMED CT | 5876000 | Acquired pancytopenia |
| SNOMED CT | 6398009 | Idiopathic chronic cold agglutinin disease |
| SNOMED CT | 7607008 | Rheumatoid pericarditis |
| SNOMED CT | 7990002 | Immunoglobulinemia with isolated somatotropin deficiency |
| SNOMED CT | 9893005 | Immunodeficiency with thymoma |
| SNOMED CT | 10639003 | Plasmacytoma |
| SNOMED CT | 11013005 | SLE glomerulonephritis syndrome, WHO class VI |
| SNOMED CT | 12814002 | B lymphocyte disorder |
| SNOMED CT | 13048006 | Orbital lymphoma |
| SNOMED CT | 13720003 | Triclonal gammopathy |
| SNOMED CT | 14024008 | Humoral immunologic aplastic anemia |
| SNOMED CT | 15012007 | Granulocyte abnormality due to immune defect |
| SNOMED CT | 16618004 | Polyclonal gammopathy |
| SNOMED CT | 17182001 | Agranulocytosis |
| SNOMED CT | 18827005 | Complement abnormality |
| SNOMED CT | 19340000 | Malignant lymphoma, lymphoplasmacytic |
| SNOMED CT | 19682006 | Lupus hepatitis |
| SNOMED CT | 19944001 | Autologous bone marrow transplant without purging |
| SNOMED CT | 20224008 | Delta heavy chain disease |
| SNOMED CT | 20447006 | Plasma cell dyscrasia with polyneuropathy |
| SNOMED CT | 21043008 | Neutrophil cytomatrix disorder |
| SNOMED CT | 21527007 | Chronic granulomatous disease, type IV |
| SNOMED CT | 22098000 | Chronic idiopathic autoimmune hemolytic anemia |
| SNOMED CT | 22331004 | Acute monocytic leukemia |
| SNOMED CT | 22406001 | SCID due to absent lymphoid stem cells |
| SNOMED CT | 23132008 | AL amyloidosis |
| SNOMED CT | 23238000 | Common variable agammaglobulinemia |
| SNOMED CT | 24072005 | Acute leukemia |
| SNOMED CT | 24181002 | Aplasia of thymus gland with immunodeficiency |
| SNOMED CT | 24419001 | Disorder of complement |
| SNOMED CT | 24743004 | Complement deficiency disease |
| SNOMED CT | 24974008 | Myelokathexis |
| SNOMED CT | 25050002 | Alpha heavy chain disease, respiratory form |
| SNOMED CT | 25109007 | Lymphopenic agammaglobulinemia - short-limbed dwarfism syndrome |

Continued on next page

**Table 7 – continued from previous page**

| CodeSystem | ConceptCode | ConceptName |
|---|---|---|
| SNOMED CT | 25380002 | Pericarditis secondary to systemic lupus erythematosus |
| SNOMED CT | 25502009 | Episodic lymphocytopenia |
| SNOMED CT | 26252007 | Chronic granulomatous disease, type IIA |
| SNOMED CT | 28880005 | Rheumatoid carditis |
| SNOMED CT | 28950004 | Acute promyelocytic leukemia |
| SNOMED CT | 28975000 | Constitutional aplastic anemia |
| SNOMED CT | 29260007 | Immunoglobulin A deficiency |
| SNOMED CT | 29272001 | Chronic granulomatous disease, type I |
| SNOMED CT | 30962008 | Acute myelomonocytic leukemia |
| SNOMED CT | 31323000 | Severe combined immunodeficiency disease |
| SNOMED CT | 31839002 | Myasthenia gravis, adult form |
| SNOMED CT | 32092008 | Toxic neutropenia |
| SNOMED CT | 33286000 | Secondary immune deficiency disorder |
| SNOMED CT | 34916006 | Neutrophil motility disorder |
| SNOMED CT | 35287006 | Myeloid sarcoma |
| SNOMED CT | 36138009 | Congenital immunodeficiency disease |
| SNOMED CT | 36402006 | SLE glomerulonephritis syndrome, WHO class IV |
| SNOMED CT | 36980009 | SCID due to absent adenosine deaminase |
| SNOMED CT | 37465004 | Plasmacytosis |
| SNOMED CT | 37548006 | Hypopigmentation-immunodeficiency disease |
| SNOMED CT | 38970002 | Doan-Wright syndrome |
| SNOMED CT | 39086001 | Hodgkin lymphoma, nodular sclerosis, cellular phase |
| SNOMED CT | 39674000 | Familial C3B inhibitor deficiency syndrome |
| SNOMED CT | 39795003 | Hand-Schüller-Christian disease |
| SNOMED CT | 40145002 | Congenital neutrophil actin dysfunction |
| SNOMED CT | 40197009 | Chronic granulomatous disease, type IA |
| SNOMED CT | 40780007 | Human immunodeficiency virus I infection |
| SNOMED CT | 41529000 | Hodgkin lymphoma, mixed cellularity |
| SNOMED CT | 41814009 | Neutropenia with dysgranulopoiesis |
| SNOMED CT | 43858000 | Secondary aplastic anemia |
| SNOMED CT | 43985008 | Hodgkin lymphoma, nodular sclerosis, grade 2 |
| SNOMED CT | 44306006 | T lymphocyte disorder |
| SNOMED CT | 44371002 | Mixed polyclonal cryoimmunoglobulinemia |
| SNOMED CT | 44940001 | Adenosine deaminase deficiency |
| SNOMED CT | 45390000 | SCID due to absent IL-2 production |
| SNOMED CT | 45572000 | Hodgkin lymphoma, nodular sclerosis, grade 1 |
| SNOMED CT | 45841007 | Kappa light chain disease |
| SNOMED CT | 46280001 | Autologous bone marrow transplant with purging |
| SNOMED CT | 46744002 | Follicular lymphoma, grade 1 |
| SNOMED CT | 46923007 | Hodgkin sarcoma [obs] |
| SNOMED CT | 47318007 | Drug-induced neutropenia |
| SNOMED CT | 48119005 | Dysplasia of thymus gland with immunodeficiency |
| SNOMED CT | 48794007 | HIV infection with infectious mononucleosis-like syndrome |

Continued on next page

**Table 7 – continued from previous page**

| CodeSystem | ConceptCode | ConceptName |
|---|---|---|
| SNOMED CT | 49555001 | SCID due to absent T cell receptor |
| SNOMED CT | 50220002 | Cellular immunologic aplastic anemia |
| SNOMED CT | 51092000 | B-cell chronic lymphocytic leukaemia/small lymphocytic lymphoma |
| SNOMED CT | 51319002 | Light chain disease |
| SNOMED CT | 52042003 | SLE glomerulonephritis syndrome, WHO class V |
| SNOMED CT | 52064007 | Biclonal gammopathy |
| SNOMED CT | 52079000 | Congenital human immunodeficiency virus infection |
| SNOMED CT | 52220008 | Acute megakaryoblastic leukemia |
| SNOMED CT | 52248008 | Hodgkin lymphoma, nodular sclerosis |
| SNOMED CT | 52661003 | Extra-articular rheumatoid process |
| SNOMED CT | 52967002 | Myelofibrosis |
| SNOMED CT | 54072008 | Nonbacterial verrucal endocardiosis |
| SNOMED CT | 54087003 | Hairy cell leukemia variant |
| SNOMED CT | 54097007 | White blood cell disorder |
| SNOMED CT | 55020008 | Follicular lymphoma, grade 2 |
| SNOMED CT | 55051001 | Myasthenia gravis, juvenile form |
| SNOMED CT | 55150002 | Follicular lymphoma |
| SNOMED CT | 55464009 | Systemic lupus erythematosus |
| SNOMED CT | 55907008 | Acquired aplastic anemia |
| SNOMED CT | 55921005 | Multiple myeloma |
| SNOMED CT | 56918001 | Dose-related drug-induced neutropenia |
| SNOMED CT | 57390009 | Mixed cryoimmunoglobulinemia with monoclonal component |
| SNOMED CT | 58034007 | Congenital hypergammaglobulinemia |
| SNOMED CT | 58390007 | Allogeneic bone marrow transplantation |
| SNOMED CT | 58606001 | Primary immune deficiency disorder |
| SNOMED CT | 58648008 | Benign monoclonal gammopathy |
| SNOMED CT | 58776007 | Autologous bone marrow transplant |
| SNOMED CT | 58961005 | Lethal midline granuloma |
| SNOMED CT | 60620005 | Epsilon heavy chain disease |
| SNOMED CT | 60743005 | Purine-nucleoside phosphorylase deficiency |
| SNOMED CT | 61493004 | Mu heavy chain disease |
| SNOMED CT | 62479008 | AIDS |
| SNOMED CT | 63364005 | Chronic myeloid leukemia |
| SNOMED CT | 64575004 | Malignant lymphoma, lymphocytic |
| SNOMED CT | 65623009 | Immune neutropenia |
| SNOMED CT | 66876008 | Lambda light chain disease |
| SNOMED CT | 68504005 | Ataxia-telangiectasia syndrome |
| SNOMED CT | 68712004 | Lymphocytic hypoplasia |
| SNOMED CT | 68815009 | SLE glomerulonephritis syndrome |
| SNOMED CT | 68979007 | Heavy chain disease |
| SNOMED CT | 69077002 | Acute basophilic leukemia |

Continued on next page

**Table 7 – continued from previous page**

| CodeSystem | ConceptCode | ConceptName |
|---|---|---|
| SNOMED CT | 69896004 | RhA - Rheumatoid arthritis |
| SNOMED CT | 70958007 | Plasma cell hyperplasia of bone marrow |
| SNOMED CT | 71436005 | Lazy leukocyte syndrome |
| SNOMED CT | 71904008 | SCID due to absent class II HLA antigens |
| SNOMED CT | 71922006 | Immune defect |
| SNOMED CT | 73286009 | SLE glomerulonephritis syndrome, WHO class I |
| SNOMED CT | 74189002 | Hodgkin granuloma [obs] |
| SNOMED CT | 76243000 | Chronic granulomatous disease, type IVA |
| SNOMED CT | 76521009 | SLE glomerulonephritis syndrome, WHO class III |
| SNOMED CT | 77084001 | Immunologic aplastic anemia |
| SNOMED CT | 77330006 | Chronic granulomatous disease, type II |
| SNOMED CT | 77358003 | Congenital leukocyte adherence deficiency |
| SNOMED CT | 77753005 | Lupus disease of the lung |
| SNOMED CT | 78378009 | Isoimmune neutropenia |
| SNOMED CT | 79019005 | Human immunodeficiency virus II infection |
| SNOMED CT | 79369007 | Complication of transplanted pancreas |
| SNOMED CT | 80570006 | Acute panmyelosis with myelofibrosis |
| SNOMED CT | 80976008 | Myasthenic crisis |
| SNOMED CT | 81166004 | Properdin deficiency disease |
| SNOMED CT | 82178003 | Neonatal myasthenia gravis |
| SNOMED CT | 82317007 | Chronic granulomatous disease, type III |
| SNOMED CT | 82546001 | Reactive immunoproliferative disease |
| SNOMED CT | 83980004 | Impaired phagocytosis |
| SNOMED CT | 86219005 | Uveitis-rheumatoid arthritis syndrome |
| SNOMED CT | 86406008 | Human immunodeficiency virus infection |
| SNOMED CT | 87117006 | HIV infection with acute lymphadenitis |
| SNOMED CT | 89454001 | Shwachman syndrome |
| SNOMED CT | 89655007 | Congenital neutropenia |
| SNOMED CT | 90120004 | Mycosis fungoides |
| SNOMED CT | 90414007 | Chronic acquired pure red cell aplasia |
| SNOMED CT | 91637004 | Myasthenia gravis |
| SNOMED CT | 91854005 | Acute leukemia in remission |
| SNOMED CT | 91855006 | Acute leukemia |
| SNOMED CT | 91856007 | Acute lymphoid leukemia in remission |
| SNOMED CT | 91857003 | Acute lymphoid leukemia |
| SNOMED CT | 91858008 | Acute monocytic leukemia in remission |
| SNOMED CT | 91860005 | Acute myeloid leukemia in remission |
| SNOMED CT | 91861009 | Acute myeloid leukemia, disease |
| SNOMED CT | 91947003 | Asymptomatic human immunodeficiency virus infection |
| SNOMED CT | 91948008 | Asymptomatic human immunodeficiency virus infection in pregnancy |
| SNOMED CT | 92811003 | Chronic leukemia in remission |
| SNOMED CT | 92812005 | Chronic leukemia |

Continued on next page

**Table 7 – continued from previous page**

| CodeSystem | ConceptCode | ConceptName |
|---|---|---|
| SNOMED CT | 92813000 | Chronic lymphoid leukemia in remission |
| SNOMED CT | 92814006 | Chronic lymphoid leukemia, disease |
| SNOMED CT | 92817004 | Chronic myeloid leukemia in remission |
| SNOMED CT | 92818009 | Chronic myeloid leukemia |
| SNOMED CT | 93133006 | Letterer-Siwe disease of intra-abdominal lymph nodes |
| SNOMED CT | 93134000 | Letterer-Siwe disease of intrapelvic lymph nodes |
| SNOMED CT | 93135004 | Letterer-Siwe disease of intrathoracic lymph nodes |
| SNOMED CT | 93136003 | Letterer-Siwe disease of lymph nodes of axilla AND/OR upper limb |
| SNOMED CT | 93137007 | Letterer-Siwe disease of lymph nodes of head, face AND/OR neck |
| SNOMED CT | 93138002 | Letterer-Siwe disease of lymph nodes of inguinal region AND/OR lower limb |
| SNOMED CT | 93139005 | Letterer-Siwe disease of lymph nodes of multiple sites |
| SNOMED CT | 93140007 | Letterer-Siwe disease of spleen |
| SNOMED CT | 93141006 | Letterer-Siwe disease of extranodal AND/OR solid organ site |
| SNOMED CT | 93142004 | Leukemia in remission |
| SNOMED CT | 93143009 | Leukemia |
| SNOMED CT | 93144003 | Leukemic reticuloendotheliosis of intra-abdominal lymph nodes |
| SNOMED CT | 93145002 | Leukemic reticuloendotheliosis of intrapelvic lymph nodes |
| SNOMED CT | 93146001 | Leukemic reticuloendotheliosis of intrathoracic lymph nodes |
| SNOMED CT | 93150008 | Leukemic reticuloendotheliosis of lymph nodes of multiple sites |
| SNOMED CT | 93151007 | Hairy cell leukemia of spleen |
| SNOMED CT | 93152000 | Leukemic reticuloendotheliosis of extranodal AND/OR solid organ site |
| SNOMED CT | 93169003 | Lymphoid leukemia in remission |
| SNOMED CT | 93182006 | Malignant histiocytosis of intra-abdominal lymph nodes |
| SNOMED CT | 93183001 | Malignant histiocytosis of intrapelvic lymph nodes |
| SNOMED CT | 93184007 | Malignant histiocytosis of intrathoracic lymph nodes |
| SNOMED CT | 93185008 | Malignant histiocytosis of lymph nodes of axilla AND/OR upper limb |
| SNOMED CT | 93186009 | Malignant histiocytosis of lymph nodes of head, face AND/OR neck |
| SNOMED CT | 93187000 | Malignant histiocytosis of lymph nodes of inguinal region AND/OR lower limb |
| SNOMED CT | 93188005 | Malignant histiocytosis of lymph nodes of multiple sites |
| SNOMED CT | 93189002 | Malignant histiocytosis of spleen |
| SNOMED CT | 93190006 | Malignant histiocytosis of extranodal AND/OR solid organ site |
| SNOMED CT | 93191005 | Malignant lymphoma of intra-abdominal lymph nodes |

Continued on next page

**Table 7 – continued from previous page**

| CodeSystem | ConceptCode | ConceptName |
|---|---|---|
| SNOMED CT | 93192003 | Malignant lymphoma of intrapelvic lymph nodes |
| SNOMED CT | 93193008 | Malignant lymphoma of intrathoracic lymph nodes |
| SNOMED CT | 93194002 | Malignant lymphoma of lymph nodes of axilla AND/OR upper limb |
| SNOMED CT | 93195001 | Malignant lymphoma of lymph nodes of head, face AND/OR neck |
| SNOMED CT | 93196000 | Malignant lymphoma of lymph nodes of inguinal region AND/OR lower limb |
| SNOMED CT | 93197009 | Malignant lymphoma of lymph nodes of multiple sites |
| SNOMED CT | 93198004 | Malignant lymphoma of spleen |
| SNOMED CT | 93199007 | Malignant lymphoma of extranodal AND/OR solid organ site |
| SNOMED CT | 93200005 | Malignant mast cell tumor of intra-abdominal lymph nodes |
| SNOMED CT | 93201009 | Malignant mast cell tumor of intrapelvic lymph nodes |
| SNOMED CT | 93202002 | Malignant mast cell tumor of intrathoracic lymph nodes |
| SNOMED CT | 93203007 | Malignant mast cell tumor of lymph nodes of axilla AND/OR upper limb |
| SNOMED CT | 93204001 | Malignant mast cell tumor of lymph nodes of head, face AND/OR neck |
| SNOMED CT | 93205000 | Malignant mast cell tumor of lymph nodes of inguinal region AND/OR lower limb |
| SNOMED CT | 94148006 | Megakaryocytic leukemia in remission |
| SNOMED CT | 94686001 | Mixed cell type lymphosarcoma of intra-abdominal lymph nodes |
| SNOMED CT | 94687005 | Mixed cell type lymphosarcoma of intrapelvic lymph nodes |
| SNOMED CT | 94688000 | Mixed cell type lymphosarcoma of intrathoracic lymph nodes |
| SNOMED CT | 94690004 | Mixed cell type lymphosarcoma of lymph nodes of head, face, and neck |
| SNOMED CT | 94704006 | Multiple myeloma in remission |
| SNOMED CT | 94707004 | Mycosis fungoides of intra-abdominal lymph nodes |
| SNOMED CT | 94708009 | Mycosis fungoides of intrapelvic lymph nodes |
| SNOMED CT | 94709001 | Mycosis fungoides of intrathoracic lymph nodes |
| SNOMED CT | 94710006 | Mycosis fungoides of lymph nodes of axilla AND/OR upper limb |
| SNOMED CT | 94711005 | Mycosis fungoides of lymph nodes of head, face AND/OR neck |
| SNOMED CT | 94712003 | Mycosis fungoides of lymph nodes of inguinal region AND/OR lower limb |
| SNOMED CT | 94714002 | Mycosis fungoides of spleen |
| SNOMED CT | 94715001 | Mycosis fungoides of extranodal AND/OR solid organ site |
| SNOMED CT | 94716000 | Myeloid leukemia in remission |
| SNOMED CT | 94718004 | Myeloid sarcoma in remission |

Continued on next page

**Table 7 – continued from previous page**

| CodeSystem | ConceptCode | ConceptName |
|---|---|---|
| SNOMED CT | 95186006 | Nodular lymphoma of intra-abdominal lymph nodes |
| SNOMED CT | 95187002 | Nodular lymphoma of intrapelvic lymph nodes |
| SNOMED CT | 95188007 | Nodular lymphoma of intrathoracic lymph nodes |
| SNOMED CT | 95192000 | Nodular lymphoma of lymph nodes of multiple sites |
| SNOMED CT | 95193005 | Nodular lymphoma of spleen |
| SNOMED CT | 95194004 | Nodular lymphoma of extranodal AND/OR solid organ site |
| SNOMED CT | 95209008 | Plasma cell leukemia in remission |
| SNOMED CT | 95210003 | Plasma cell leukemia |
| SNOMED CT | 95224004 | Reticulosarcoma of intra-abdominal lymph nodes |
| SNOMED CT | 95225003 | Reticulosarcoma of intrapelvic lymph nodes |
| SNOMED CT | 95226002 | Reticulosarcoma of intrathoracic lymph nodes |
| SNOMED CT | 95230004 | Reticulosarcoma of lymph nodes of multiple sites |
| SNOMED CT | 95231000 | Reticulosarcoma of spleen |
| SNOMED CT | 95260009 | Sézary's disease of lymph nodes of head, face AND/OR neck |
| SNOMED CT | 95263006 | Sézary's disease of spleen |
| SNOMED CT | 95264000 | Sézary's disease of extranodal AND/OR solid organ site |
| SNOMED CT | 95644001 | Systemic lupus erythematosus encephalitis |
| SNOMED CT | 95892003 | Persistent generalized lymphadenopathy |
| SNOMED CT | 103075007 | Humoral immune defect |
| SNOMED CT | 103077004 | Immunodeficiency secondary to neoplasm |
| SNOMED CT | 103078009 | Immunodeficiency secondary to trauma |
| SNOMED CT | 103079001 | Immunodeficiency secondary to corticosteroid |
| SNOMED CT | 103080003 | Immunodeficiency secondary to radiation therapy |
| SNOMED CT | 103081004 | Immunodeficiency secondary to chemotherapy |
| SNOMED CT | 103682005 | Subcutaneous panniculitis-like T-cell lymphoma |
| SNOMED CT | 103685007 | Hepatosplenic gamma-delta cell lymphoma |
| SNOMED CT | 103688009 | Acute myeloid leukemia with abnormal marrow eosinophils |
| SNOMED CT | 103689001 | Acute myeloid leukemia, minimal differentiation |
| SNOMED CT | 103690005 | Acute myeloid leukemia without maturation |
| SNOMED CT | 103691009 | Acute myeloid leukemia with maturation |
| SNOMED CT | 105600002 | Disorder of neutrophils |
| SNOMED CT | 105603000 | Non-malignant lymphocyte AND/OR plasma cell disorder |
| SNOMED CT | 109958007 | Hematologic neoplasm of uncertain behavior |
| SNOMED CT | 109975001 | T-zone lymphoma (clinical) |
| SNOMED CT | 109976000 | Lymphoepithelioid lymphoma (clinical) |
| SNOMED CT | 109978004 | T-cell lymphoma (clinical) |
| SNOMED CT | 109980005 | Malignant immunoproliferative disease (clinical) |
| SNOMED CT | 109982002 | Alpha heavy chain disease (clinical) |
| SNOMED CT | 109984001 | Gamma heavy chain disease (clinical) |
| SNOMED CT | 109985000 | Immunoproliferative small intestinal disease (clinical) |
| SNOMED CT | 109988003 | Histiocytic sarcoma (clinical) |
| SNOMED CT | 109989006 | Multiple myeloma |
| SNOMED CT | 109991003 | Acute panmyelosis with myelofibrosis |

Continued on next page

**Table 7 – continued from previous page**

| CodeSystem | ConceptCode | ConceptName |
|---|---|---|
| SNOMED CT | 109993000 | Chronic myeloproliferative disorder (clinical) |
| SNOMED CT | 109994006 | Essential thrombocythemia |
| SNOMED CT | 109995007 | Myelodysplastic syndrome (clinical) |
| SNOMED CT | 109996008 | Refractory anemia (clinical) |
| SNOMED CT | 109998009 | Refractory anemia with ringed sideroblasts associated with marked thrombocytosis |
| SNOMED CT | 110000005 | Refractory anemia with excess blasts in transformation (clinical) |
| SNOMED CT | 110002002 | Mast cell leukemia (clinical) |
| SNOMED CT | 110006004 | Prolymphocytic leukemia (clinical) |
| SNOMED CT | 110459008 | Malignant lymphoma, metastatic |
| SNOMED CT | 111001004 | Gammopathy |
| SNOMED CT | 111584000 | Reticular dysgenesis |
| SNOMED CT | 111585004 | Neutropenia associated with autoimmune disease |
| SNOMED CT | 111587007 | SCID due to absent IL-2 receptor |
| SNOMED CT | 111880001 | Acute HIV infection |
| SNOMED CT | 112687003 | Hodgkin lymphoma, lymphocyte depletion |
| SNOMED CT | 115244002 | Malignant lymphoma - category |
| SNOMED CT | 115245001 | Malignant lymphoma, follicular AND/OR nodular |
| SNOMED CT | 115246000 | Specified cutaneous AND/OR peripheral T cell lymphoma |
| SNOMED CT | 116133005 | Congenital agammaglobulinemia |
| SNOMED CT | 118607005 | Lymphocyte-rich classical Hodgkin lymphoma |
| SNOMED CT | 118611004 | Sézary's disease (clinical) |
| SNOMED CT | 118612006 | Malignant histiocytosis (clinical) |
| SNOMED CT | 118615008 | Malignant mast cell tumor (clinical) |
| SNOMED CT | 118618005 | Mycosis fungoides (clinical) |
| SNOMED CT | 119249001 | Agammaglobulinemia |
| SNOMED CT | 119250001 | Hypogammaglobulinemia |
| SNOMED CT | 123313007 | Alpha heavy chain disease, enteric form |
| SNOMED CT | 123777002 | Autoimmune leukopenia |
| SNOMED CT | 123782009 | Dysgammaglobulinemia |
| SNOMED CT | 124950009 | Disorder of immunoglobulin |
| SNOMED CT | 127034005 | Pancytopenia |
| SNOMED CT | 127069007 | Hemophagocytic syndrome |
| SNOMED CT | 127070008 | Malignant histiocytic disorder |
| SNOMED CT | 127220001 | Malignant lymphoma of lymph nodes |
| SNOMED CT | 127225006 | Chronic myelomonocytic leukemia |
| SNOMED CT | 127388009 | Hypergammaglobulinemia |
| SNOMED CT | 128623006 | Myelodysplastic syndrome |
| SNOMED CT | 128798004 | Composite Hodgkin and non-Hodgkin lymphoma |
| SNOMED CT | 128800006 | Primary effusion lymphoma |
| SNOMED CT | 128801005 | Mediastinal large B-cell lymphoma |
| SNOMED CT | 128802003 | Splenic marginal zone B-cell lymphoma |

Continued on next page

**Table 7 – continued from previous page**

| CodeSystem | ConceptCode | ConceptName |
|---|---|---|
| SNOMED CT | 128803008 | Marginal zone B-cell lymphoma |
| SNOMED CT | 128806000 | Precursor cell lymphoblastic lymphoma |
| SNOMED CT | 128807009 | Precursor B-cell lymphoblastic lymphoma |
| SNOMED CT | 128808004 | Precursor T-cell lymphoblastic lymphoma |
| SNOMED CT | 128813000 | Histiocytic sarcoma |
| SNOMED CT | 128814006 | Langerhans cell sarcoma |
| SNOMED CT | 128815007 | Interdigitating dendritic cell sarcoma |
| SNOMED CT | 128816008 | Follicular dendritic cell sarcoma |
| SNOMED CT | 128818009 | Acute biphenotypic leukemia |
| SNOMED CT | 128820007 | Prolymphocytic leukemia, B-cell type |
| SNOMED CT | 128822004 | Precursor cell lymphoblastic leukemia |
| SNOMED CT | 128823009 | Precursor B-cell lymphoblastic leukemia |
| SNOMED CT | 128824003 | Precursor T-cell lymphoblastic leukemia |
| SNOMED CT | 128825002 | Chronic myelogenous leukemia, BCR/ABL positive |
| SNOMED CT | 128826001 | Atypical chronic myeloid leukemia, BCR/ABL negative |
| SNOMED CT | 128827005 | Acute myeloid leukemia with myelodysplasia-related changes |
| SNOMED CT | 128828000 | Acute myeloid leukemia, t(8;21) (q22;q22) |
| SNOMED CT | 128829008 | Acute myeloid leukemia, 11q23 abnormalities |
| SNOMED CT | 128830003 | Therapy-related acute myeloid leukemia and myelodysplastic syndrome |
| SNOMED CT | 128832006 | Juvenile myelomonocytic leukemia |
| SNOMED CT | 128834007 | Chronic neutrophilic leukemia |
| SNOMED CT | 128836009 | Myelodysplastic syndrome with multilineage dysplasia |
| SNOMED CT | 128837000 | Myelodysplastic syndrome with isolated del (5q) |
| SNOMED CT | 128838005 | Therapy-related myelodysplastic syndrome |
| SNOMED CT | 128843003 | Myelosclerosis with myeloid metaplasia |
| SNOMED CT | 128845005 | Refractory anemia |
| SNOMED CT | 128846006 | Refractory anemia with ring sideroblasts associated with marked thrombocytosis |
| SNOMED CT | 128847002 | Myelodysplastic syndrome with excess blasts |
| SNOMED CT | 128848007 | Refractory anemia with excess blasts in transformation |
| SNOMED CT | 128920006 | Malignant histiocytosis |
| SNOMED CT | 128921005 | Plasmacytoma, extramedullary (not occurring in bone) |
| SNOMED CT | 128922003 | Plasma cell leukemia |
| SNOMED CT | 128923008 | Prolymphocytic leukemia |
| SNOMED CT | 128924002 | Mast cell leukemia |
| SNOMED CT | 128931003 | Leukemia (category) |
| SNOMED CT | 128932005 | Acute leukemia (category) |
| SNOMED CT | 128933000 | Chronic leukemia (category) |
| SNOMED CT | 128934006 | Myeloid leukemia (category) |
| SNOMED CT | 128935007 | Lymphoid leukemia (category) |
| SNOMED CT | 129642004 | Chronic idiopathic immunoneutropenia in adult |

Continued on next page

**Table 7 – continued from previous page**

| CodeSystem | ConceptCode | ConceptName |
|---|---|---|
| SNOMED CT | 129643009 | Chronic hypoplastic neutropenia |
| SNOMED CT | 129644003 | Myeloperoxidase deficiency syndrome |
| SNOMED CT | 129648000 | Hypoalphaglobulinemia |
| SNOMED CT | 129649008 | Hypobetaglobulinemia |
| SNOMED CT | 165816005 | HIV positive |
| SNOMED CT | 186706006 | Human immunodeficiency virus infection constitutional disease |
| SNOMED CT | 186707002 | Human immunodeficiency virus infection with neurological disease |
| SNOMED CT | 186708007 | Human immunodeficiency virus infection with secondary clinical infectious disease |
| SNOMED CT | 188487008 | Lymphosarcoma and reticulosarcoma |
| SNOMED CT | 188489006 | Reticulosarcoma of lymph nodes of head, face and neck |
| SNOMED CT | 188492005 | Reticulosarcoma of lymph nodes of axilla and upper limb |
| SNOMED CT | 188493000 | Reticulosarcoma of lymph nodes of inguinal region and lower limb |
| SNOMED CT | 188498009 | Lymphosarcoma |
| SNOMED CT | 188500005 | Lymphosarcoma of lymph nodes of head, face and neck |
| SNOMED CT | 188501009 | Lymphosarcoma of intrathoracic lymph nodes |
| SNOMED CT | 188502002 | Lymphosarcoma of intra-abdominal lymph nodes |
| SNOMED CT | 188503007 | Lymphosarcoma of lymph nodes of axilla and upper limb |
| SNOMED CT | 188504001 | Lymphosarcoma of lymph nodes of inguinal region and lower limb |
| SNOMED CT | 188505000 | Lymphosarcoma of intrapelvic lymph nodes |
| SNOMED CT | 188506004 | Lymphosarcoma of spleen |
| SNOMED CT | 188507008 | Lymphosarcoma of lymph nodes of multiple sites |
| SNOMED CT | 188609000 | Nodular lymphoma of lymph nodes of head, face and neck |
| SNOMED CT | 188612002 | Nodular lymphoma of lymph nodes of axilla and upper limb |
| SNOMED CT | 188613007 | Nodular lymphoma of lymph nodes of inguinal region and lower limb |
| SNOMED CT | 188627002 | Mycosis fungoides of lymph nodes of multiple sites |
| SNOMED CT | 188630009 | Sézary's disease of lymph nodes of head, face and neck |
| SNOMED CT | 188631008 | Sézary's disease of intrathoracic lymph nodes |
| SNOMED CT | 188633006 | Sézary's disease of lymph nodes of axilla AND/OR upper limb |
| SNOMED CT | 188634000 | Sézary's disease of lymph nodes of inguinal region and lower limb |
| SNOMED CT | 188635004 | Sézary's disease of intrapelvic lymph nodes |
| SNOMED CT | 188637007 | Sézary's disease of lymph nodes of multiple sites |
| SNOMED CT | 188640007 | Malignant histiocytosis of lymph nodes of head, face and neck |
| SNOMED CT | 188641006 | Malignant histiocytosis of lymph nodes of axilla and upper limb |

Continued on next page

**Table 7 – continued from previous page**

| CodeSystem | ConceptCode | ConceptName |
|---|---|---|
| SNOMED CT | 188642004 | Malignant histiocytosis of lymph nodes of inguinal region and lower limb |
| SNOMED CT | 188645002 | Leukemic reticuloendotheliosis of lymph nodes of head, face and neck |
| SNOMED CT | 188648000 | Leukemic reticuloendotheliosis of lymph nodes of axilla and upper limb |
| SNOMED CT | 188649008 | Leukemic reticuloendotheliosis of lymph nodes of inguinal region and lower limb |
| SNOMED CT | 188660004 | Malignant mast cell tumors |
| SNOMED CT | 188662007 | Mast cell malignancy of lymph nodes of head, face and neck |
| SNOMED CT | 188663002 | Mast cell malignancy of intrathoracic lymph nodes |
| SNOMED CT | 188664008 | Mast cell malignancy of intra-abdominal lymph nodes |
| SNOMED CT | 188665009 | Mast cell malignancy of lymph nodes of axilla and upper limb |
| SNOMED CT | 188666005 | Mast cell malignancy of lymph nodes of inguinal region and lower limb |
| SNOMED CT | 188667001 | Mast cell malignancy of intrapelvic lymph nodes |
| SNOMED CT | 188668006 | Mast cell malignancy of spleen |
| SNOMED CT | 188669003 | Mast cell malignancy of lymph nodes of multiple sites |
| SNOMED CT | 188674006 | Diffuse malignant lymphoma - small non-cleaved cell |
| SNOMED CT | 188675007 | Malignant lymphoma - small cleaved cell |
| SNOMED CT | 188676008 | Malignant lymphoma - mixed small and large cell |
| SNOMED CT | 188691005 | Malignant immunoproliferative small intestinal disease |
| SNOMED CT | 188718006 | Extramedullary plasmacytoma |
| SNOMED CT | 188725004 | Lymphoid leukemia |
| SNOMED CT | 188726003 | Subacute lymphoid leukemia |
| SNOMED CT | 188728002 | Aleukemic lymphoid leukemia |
| SNOMED CT | 188729005 | Adult T-cell leukemia |
| SNOMED CT | 188732008 | Myeloid leukemia |
| SNOMED CT | 188733003 | Chronic eosinophilic leukemia |
| SNOMED CT | 188734009 | Chronic neutrophilic leukemia |
| SNOMED CT | 188736006 | Subacute myeloid leukemia |
| SNOMED CT | 188737002 | Chloroma |
| SNOMED CT | 188738007 | Granulocytic sarcoma |
| SNOMED CT | 188741003 | Aleukemic myeloid leukemia |
| SNOMED CT | 188744006 | Monocytic leukemia |
| SNOMED CT | 188745007 | Chronic monocytic leukemia |
| SNOMED CT | 188746008 | Subacute monocytic leukemia |
| SNOMED CT | 188748009 | Aleukemic monocytic leukemia |
| SNOMED CT | 188754005 | Megakaryocytic leukemia |
| SNOMED CT | 188768003 | Myelomonocytic leukemia |
| SNOMED CT | 188770007 | Subacute myelomonocytic leukemia |
| SNOMED CT | 189509003 | Refractory anemia without sideroblasts, so stated |

Continued on next page

**Table 7 – continued from previous page**

| CodeSystem | ConceptCode | ConceptName |
|---|---|---|
| SNOMED CT | 189962004 | Malignant lymphoma, stem cell type |
| SNOMED CT | 189982000 | Reticulosarcoma morphology |
| SNOMED CT | 189984004 | Reticulosarcoma, pleomorphic cell type |
| SNOMED CT | 189985003 | Reticulosarcoma, nodular |
| SNOMED CT | 190030009 | Compound leukemias |
| SNOMED CT | 190055003 | Eosinophilic leukemia |
| SNOMED CT | 190808009 | Polyclonal hypergammaglobulinemia |
| SNOMED CT | 190817009 | Macroglobulinemia |
| SNOMED CT | 190818004 | Waldenström macroglobulinemia |
| SNOMED CT | 190923000 | Sporadic primary amyloidosis |
| SNOMED CT | 190955000 | Histiocytosis X syndrome |
| SNOMED CT | 190959006 | Haemophagocytic lymphohistiocytosis due to infection |
| SNOMED CT | 190979003 | Selective immunoglobulin A deficiency |
| SNOMED CT | 190980000 | Selective immunoglobulin M deficiency |
| SNOMED CT | 190981001 | Selective immunoglobulin G deficiency |
| SNOMED CT | 190986006 | Antibody deficiency with near-normal immunoglobulins or with hyperimmunoglobulinemia |
| SNOMED CT | 190993005 | Autosomal recessive severe combined immunodeficiency |
| SNOMED CT | 190995003 | Thymic aplasia or dysplasia with immunodeficiency |
| SNOMED CT | 190996002 | Severe combined immunodeficiency with reticular dysgenesis |
| SNOMED CT | 190997006 | Severe combined immunodeficiency with low T- and B-cell numbers |
| SNOMED CT | 190998001 | Severe combined immunodeficiency with low or normal B-cell numbers |
| SNOMED CT | 191001007 | Major histocompatibility complex class I deficiency |
| SNOMED CT | 191002000 | Major histocompatibility complex class II deficiency |
| SNOMED CT | 191008001 | Immunodeficiency following hereditary defective response to Epstein-Barr virus |
| SNOMED CT | 191011000 | Common variable immunodeficiency with predominant abnormalities of B-cell numbers and functions |
| SNOMED CT | 191012007 | Common variable immunodeficiency with predominant immunoregulatory T-cell disorders |
| SNOMED CT | 191013002 | Common variable immunodeficiency with autoantibodies to B- or T-cells |
| SNOMED CT | 191018006 | Lymphocyte function antigen-1 defect |
| SNOMED CT | 191244003 | Aplastic anemia due to chronic disease |
| SNOMED CT | 191246001 | Aplastic anemia due to infection |
| SNOMED CT | 191247005 | Aplastic anemia due to radiation |
| SNOMED CT | 191248000 | Aplastic anemia due to toxic cause |
| SNOMED CT | 191255003 | Transient acquired pure red cell aplasia |
| SNOMED CT | 191256002 | Idiopathic aplastic anemia |
| SNOMED CT | 191338000 | Primary splenic neutropenia |

Continued on next page

**Table 7 – continued from previous page**

| CodeSystem | ConceptCode | ConceptName |
|---|---|---|
| SNOMED CT | 191347008 | Cyclical neutropenia |
| SNOMED CT | 193206003 | Persistent neonatal myasthenia gravis |
| SNOMED CT | 193207007 | Juvenile or adult myasthenia gravis |
| SNOMED CT | 196138005 | Lung disease with systemic lupus erythematosus |
| SNOMED CT | 201764007 | Rheumatoid arthritis of cervical spine |
| SNOMED CT | 201766009 | Rheumatoid arthritis of shoulder |
| SNOMED CT | 201767000 | Rheumatoid arthritis of sternoclavicular joint |
| SNOMED CT | 201768005 | Rheumatoid arthritis of acromioclavicular joint |
| SNOMED CT | 201769002 | Rheumatoid arthritis of elbow |
| SNOMED CT | 201770001 | Rheumatoid arthritis of distal radioulnar joint |
| SNOMED CT | 201771002 | Rheumatoid arthritis of wrist |
| SNOMED CT | 201772009 | Rheumatoid arthritis of metacarpophalangeal joint |
| SNOMED CT | 201773004 | Rheumatoid arthritis of proximal interphalangeal joint of finger |
| SNOMED CT | 201774005 | Rheumatoid arthritis of distal interphalangeal joint of finger |
| SNOMED CT | 201775006 | Rheumatoid arthritis of hip |
| SNOMED CT | 201776007 | Rheumatoid arthritis of sacroiliac joint |
| SNOMED CT | 201777003 | Rheumatoid arthritis of knee |
| SNOMED CT | 201778008 | Rheumatoid arthritis of tibiofibular joint |
| SNOMED CT | 201779000 | Rheumatoid arthritis of ankle |
| SNOMED CT | 201780002 | Rheumatoid arthritis of subtalar joint |
| SNOMED CT | 201781003 | Rheumatoid arthritis of talonavicular joint |
| SNOMED CT | 201783000 | Rheumatoid arthritis of first metatarsophalangeal joint |
| SNOMED CT | 201784006 | Rheumatoid arthritis of lesser metatarsophalangeal joint |
| SNOMED CT | 201785007 | Rheumatoid arthritis of interphalangeal joint of toe |
| SNOMED CT | 201791009 | Flare of rheumatoid arthritis |
| SNOMED CT | 209962000 | Myeloma-associated amyloidosis |
| SNOMED CT | 213148006 | Transplanted organ rejection |
| SNOMED CT | 230180003 | Human immunodeficiency virus leucoencephalopathy |
| SNOMED CT | 230202002 | Vacuolar myelopathy |
| SNOMED CT | 230598008 | Neuropathy due to human immunodeficiency virus |
| SNOMED CT | 230669004 | Genetically determined myasthenia |
| SNOMED CT | 230670003 | Familial infantile myasthenia |
| SNOMED CT | 230671004 | Acetylcholine resynthesis deficiency |
| SNOMED CT | 230672006 | Congenital myasthenic syndrome |
| SNOMED CT | 230673001 | Congenital end-plate acetylcholine receptor deficiency |
| SNOMED CT | 230674007 | Pseudomyopathic myasthenia |
| SNOMED CT | 230676009 | Putative defect in acetylcholine synthesis or packaging |
| SNOMED CT | 230677000 | Congenital end-plate acetylcholinesterase deficiency |
| SNOMED CT | 230678005 | Decrease of motor end-plate potential amplitude without acetylcholine receptor deficiency |
| SNOMED CT | 230679002 | Abnormality of synaptic vesicles |
| SNOMED CT | 230683002 | Transient neonatal myasthenia gravis |

Continued on next page

**Table 7 – continued from previous page**

| CodeSystem | ConceptCode | ConceptName |
|---|---|---|
| SNOMED CT | 230684008 | Ocular myasthenia |
| SNOMED CT | 230685009 | Myasthenia gravis associated with thymoma |
| SNOMED CT | 230686005 | Generalized myasthenia |
| SNOMED CT | 230687001 | Myopathy in myasthenia gravis |
| SNOMED CT | 232075002 | Lymphoma of retina |
| SNOMED CT | 234146006 | Hennekam syndrome |
| SNOMED CT | 234331007 | Syngeneic bone marrow transplant |
| SNOMED CT | 234332000 | T-cell depleted allogeneic bone marrow graft |
| SNOMED CT | 234333005 | Imperfect T-cell depleted allogeneic bone marrow graft |
| SNOMED CT | 234334004 | Allogeneic related bone marrow transplant |
| SNOMED CT | 234335003 | Allogeneic unrelated bone marrow transplant |
| SNOMED CT | 234336002 | Hemopoietic stem cell transplant |
| SNOMED CT | 234367000 | Pancytopenia with pancreatitis |
| SNOMED CT | 234370001 | Pure red cell aplasia, acquired |
| SNOMED CT | 234376007 | Acquired red cell aplasia |
| SNOMED CT | 234414004 | Non-malignant white cell disorder |
| SNOMED CT | 234416002 | X-linked hypogammaglobulinemia |
| SNOMED CT | 234424007 | Metabolic neutropenia |
| SNOMED CT | 234425008 | Autoimmune neutropenia |
| SNOMED CT | 234428005 | Lymphocytoid disorder |
| SNOMED CT | 234429002 | Chemotactic disorder |
| SNOMED CT | 234430007 | CR3-receptor deficiency |
| SNOMED CT | 234431006 | Specific granule deficiency |
| SNOMED CT | 234433009 | Myeloperoxidase deficiency |
| SNOMED CT | 234436001 | Hemolytic erythrophagocytic syndrome |
| SNOMED CT | 234437005 | Hemophagocytic lymphohistiocytosis |
| SNOMED CT | 234519006 | Bone marrow transplant rejection |
| SNOMED CT | 234520000 | Bone marrow transplant failure |
| SNOMED CT | 234522008 | Disease relapse in transplant marrow |
| SNOMED CT | 234532001 | Immunodeficiency disorder |
| SNOMED CT | 234533006 | X-linked agammaglobulinemia with growth hormone deficiency |
| SNOMED CT | 234534000 | Autosomal agammaglobulinemia with absent B-cells |
| SNOMED CT | 234535004 | Selective immunoglobulin dysfunction |
| SNOMED CT | 234539005 | Immunoglobulin heavy chain deficiency |
| SNOMED CT | 234540007 | Selective immunoglobulin E deficiency |
| SNOMED CT | 234541006 | Selective immunoglobulin D deficiency |
| SNOMED CT | 234542004 | Selective immunoglobulin M and immunoglobulin A deficiency |
| SNOMED CT | 234543009 | Immunoglobulin light chain deficiency |
| SNOMED CT | 234544003 | Immunoglobulin subclass deficiency |
| SNOMED CT | 234546001 | Immunoglobulin G2 deficiency |
| SNOMED CT | 234547005 | Combined immunoglobulin G2 and G4 deficiency |

Continued on next page

**Table 7 – continued from previous page**

| CodeSystem | ConceptCode | ConceptName |
|---|---|---|
| SNOMED CT | 234548000 | Immunoglobulin G3 deficiency |
| SNOMED CT | 234549008 | Immunoglobulin G4 deficiency |
| SNOMED CT | 234550008 | Immunoglobulin G1 deficiency |
| SNOMED CT | 234551007 | Immunoglobulin A1 deficiency |
| SNOMED CT | 234552000 | Immunoglobulin A2 deficiency |
| SNOMED CT | 234553005 | Immunoglobulin-associated molecule deficiency |
| SNOMED CT | 234554004 | Secretory piece deficiency |
| SNOMED CT | 234555003 | Defective immunoglobulin glycosylation |
| SNOMED CT | 234563002 | Anti-staphylococcal antibody deficiency |
| SNOMED CT | 234564008 | Primary immunoglobulin catabolism abnormality |
| SNOMED CT | 234565009 | Immunoglobulin hypercatabolism |
| SNOMED CT | 234566005 | Familial immunoglobulin hypercatabolism |
| SNOMED CT | 234570002 | Severe combined immunodeficiency with maternofetal engraftment |
| SNOMED CT | 234571003 | Warts, hypogammaglobulinemia, infections, and myelokathexis |
| SNOMED CT | 234572005 | Benign combined immunodeficiency |
| SNOMED CT | 234573000 | Phagocytic cell defect |
| SNOMED CT | 234574006 | Disorder of phagocytic cell number |
| SNOMED CT | 234576008 | Chronic familial neutropenia |
| SNOMED CT | 234577004 | Lipochrome histiocytosis - familial |
| SNOMED CT | 234578009 | Defective phagocytic cell opsonization |
| SNOMED CT | 234580003 | Defective phagocytic cell chemotaxis |
| SNOMED CT | 234581004 | Defective phagocytic cell adhesion |
| SNOMED CT | 234582006 | Leukocyte adhesion deficiency - type 1 |
| SNOMED CT | 234583001 | Leukocyte adhesion deficiency - type 2 |
| SNOMED CT | 234584007 | Tuftsin deficiency |
| SNOMED CT | 234585008 | Defective phagocytic cell killing |
| SNOMED CT | 234587000 | Neutrophil lactoferrin deficiency |
| SNOMED CT | 234588005 | Neutrophil secondary granule deficiency |
| SNOMED CT | 234591005 | Combined phagocytic defect |
| SNOMED CT | 234593008 | Classical complement pathway abnormality |
| SNOMED CT | 234594002 | Complement 1q deficiency |
| SNOMED CT | 234595001 | Complement 1q beta chain deficiency |
| SNOMED CT | 234596000 | Complement 1q dysfunction |
| SNOMED CT | 234597009 | Complement 1r deficiency |
| SNOMED CT | 234598004 | Complement 1s deficiency |
| SNOMED CT | 234599007 | Complement 2 deficiency |
| SNOMED CT | 234600005 | Complement 4 deficiency |
| SNOMED CT | 234601009 | Complement 4A deficiency |
| SNOMED CT | 234602002 | Complement 4B deficiency |
| SNOMED CT | 234603007 | Complement 3 deficiency |
| SNOMED CT | 234604001 | Alternative pathway deficiency |

Continued on next page

**Table 7 – continued from previous page**

| CodeSystem | ConceptCode | ConceptName |
|---|---|---|
| SNOMED CT | 234605000 | Factor B deficiency |
| SNOMED CT | 234607008 | Factor D deficiency |
| SNOMED CT | 234608003 | Terminal component deficiency |
| SNOMED CT | 234609006 | Complement 5 deficiency |
| SNOMED CT | 234611002 | Complement 6 deficiency |
| SNOMED CT | 234612009 | Complement 7 deficiency |
| SNOMED CT | 234613004 | Combined complement 6 and 7 deficiencies |
| SNOMED CT | 234614005 | Complement 8 beta chain deficiency |
| SNOMED CT | 234615006 | Complement 8 beta chain dysfunction |
| SNOMED CT | 234616007 | Complement 8 alpha-gamma deficiency |
| SNOMED CT | 234617003 | Complement 9 deficiency |
| SNOMED CT | 234618008 | Complement regulatory factor defect |
| SNOMED CT | 234621005 | Factor I deficiency |
| SNOMED CT | 234622003 | Factor H deficiency |
| SNOMED CT | 234623008 | Complement 4 binding protein deficiency |
| SNOMED CT | 234624002 | Decay accelerating factor deficiency |
| SNOMED CT | 234625001 | Homologous restriction factor deficiency |
| SNOMED CT | 234626000 | Complement 5a inhibitor deficiency |
| SNOMED CT | 234627009 | Anaphylotoxin inactivator deficiency |
| SNOMED CT | 234628004 | Complement receptor deficiency |
| SNOMED CT | 234629007 | Complement receptor 1 deficiency |
| SNOMED CT | 234630002 | Complement receptor 3 deficiency |
| SNOMED CT | 234631003 | Immunodeficiency with major anomalies |
| SNOMED CT | 234632005 | Immunodeficiency associated with chromosomal abnormality |
| SNOMED CT | 234633000 | Centromeric instability of chromosomes 1,9 and 16 and immunodeficiency |
| SNOMED CT | 234634006 | Chromosome 18 syndromes and antibody deficiency |
| SNOMED CT | 234635007 | Chromosome 22 abnormalities with hypogammaglobulinemia |
| SNOMED CT | 234636008 | Monosomy 22 and absence of immunoglobulin A |
| SNOMED CT | 234637004 | Deletion of X-chromosome and hypogammaglobulinemia |
| SNOMED CT | 234638009 | Microcephaly, normal intelligence and immunodeficiency |
| SNOMED CT | 234639001 | Triple X syndrome, epilepsy, and hypogammaglobulinemia |
| SNOMED CT | 234640004 | Immunodeficiency associated with 18p syndrome |
| SNOMED CT | 234641000 | Immunodeficiency associated with multiple organ system abnormalities |
| SNOMED CT | 234645009 | Drug-induced immunodeficiency |
| SNOMED CT | 234646005 | Graft versus host disease |
| SNOMED CT | 235009000 | Human immunodeficiency virus-associated periodontitis |
| SNOMED CT | 235726002 | Human immunodeficiency virus enteropathy |
| SNOMED CT | 236406007 | Acquired immune deficiency syndrome-related nephropathy |
| SNOMED CT | 236512004 | Leukemic infiltrate of kidney |

Continued on next page

**Table 7 – continued from previous page**

| CodeSystem | ConceptCode | ConceptName |
|---|---|---|
| SNOMED CT | 236513009 | Lymphoma of kidney |
| SNOMED CT | 236585005 | De novo transplant disease |
| SNOMED CT | 239297008 | Lymphomatoid granulomatosis of lung |
| SNOMED CT | 239791005 | Seropositive rheumatoid arthritis |
| SNOMED CT | 239792003 | Seronegative rheumatoid arthritis |
| SNOMED CT | 239795001 | Rheumatoid arthritis with multisystem involvement |
| SNOMED CT | 239886003 | Limited lupus erythematosus |
| SNOMED CT | 239887007 | Systemic lupus erythematosus with organ/system involvement |
| SNOMED CT | 239888002 | Lupus panniculitis |
| SNOMED CT | 239889005 | Bullous systemic lupus erythematosus |
| SNOMED CT | 239890001 | Systemic lupus erythematosus with multisystem involvement |
| SNOMED CT | 239940004 | Lymphomatoid granulomatosis |
| SNOMED CT | 240103002 | Human immunodeficiency virus myopathy |
| SNOMED CT | 240486003 | Parvoviral aplastic crisis |
| SNOMED CT | 247860002 | Familial neutropenia |
| SNOMED CT | 248693006 | Chronic idiopathic neutropenia |
| SNOMED CT | 254067002 | Immuno-osseous dysplasia |
| SNOMED CT | 254290004 | Lymphoproliferative disorder following transplantation |
| SNOMED CT | 254792006 | Proliferating angioendotheliomatosis |
| SNOMED CT | 255048006 | Tumor of lymphoid, hemopoietic and related tissue |
| SNOMED CT | 255102004 | Angioendotheliomatosis |
| SNOMED CT | 255191003 | Localized malignant reticulohistiocytoma |
| SNOMED CT | 263661007 | Complement 5 dysfunction |
| SNOMED CT | 267459007 | Deficiencies of humoral immunity |
| SNOMED CT | 267460002 | Congenital hypogammaglobulinemia |
| SNOMED CT | 267524009 | Constitutional aplastic anemia with malformation |
| SNOMED CT | 267527002 | Aplastic anemia due to drugs |
| SNOMED CT | 267540007 | Neutropenia due to irradiation |
| SNOMED CT | 267543009 | Functional disorders of polymorphonuclear neutrophils |
| SNOMED CT | 269295009 | Transplanted organ failure |
| SNOMED CT | 269475001 | Malignant tumor of lymphoid hemopoietic and related tissue |
| SNOMED CT | 269476000 | Nodular lymphoma |
| SNOMED CT | 274905008 | Malignant lymphoma - lymphocytic, intermediate differentiation |
| SNOMED CT | 275523003 | Pancytopenia-dysmelia |
| SNOMED CT | 275524009 | Immunoproliferative neoplasm |
| SNOMED CT | 276628009 | Chloramphenicol-induced neutropenia |
| SNOMED CT | 276665006 | Congenital acquired immune deficiency syndrome |
| SNOMED CT | 276666007 | Congenital human immunodeficiency virus positive status syndrome |

Continued on next page

**Table 7 – continued from previous page**

| CodeSystem | ConceptCode | ConceptName |
|---|---|---|
| SNOMED CT | 276811008 | Gastric lymphoma |
| SNOMED CT | 276815004 | Lymphoma of intestine |
| SNOMED CT | 276836002 | Primary cerebral lymphoma |
| SNOMED CT | 277473004 | B-cell chronic lymphocytic leukemia |
| SNOMED CT | 277474005 | B-cell chronic lymphocytic leukemia variant |
| SNOMED CT | 277543005 | Malignant white blood cell disorder |
| SNOMED CT | 277545003 | T-cell chronic lymphocytic leukemia |
| SNOMED CT | 277549009 | Chronic lymphocytic prolymphocytic leukemia syndrome |
| SNOMED CT | 277551008 | Splenic lymphoma with villous lymphocytes |
| SNOMED CT | 277567002 | T-cell prolymphocytic leukemia |
| SNOMED CT | 277568007 | Hairy cell leukemia variant |
| SNOMED CT | 277569004 | Large granular lymphocytic leukemia |
| SNOMED CT | 277570003 | Lymphoma with spill |
| SNOMED CT | 277571004 | B-cell acute lymphoblastic leukemia |
| SNOMED CT | 277572006 | Precursor B-cell acute lymphoblastic leukemia |
| SNOMED CT | 277573001 | Common acute lymphoblastic leukemia |
| SNOMED CT | 277574007 | Null cell acute lymphoblastic leukemia |
| SNOMED CT | 277575008 | T-cell acute lymphoblastic leukemia |
| SNOMED CT | 277579002 | Light chain myeloma |
| SNOMED CT | 277580004 | Non-secretory myeloma |
| SNOMED CT | 277587001 | Juvenile chronic myeloid leukemia |
| SNOMED CT | 277589003 | Atypical chronic myeloid leukemia |
| SNOMED CT | 277597005 | Myelodysplastic syndrome with isolated del(5q) |
| SNOMED CT | 277601005 | Acute monoblastic leukemia |
| SNOMED CT | 277602003 | Acute megakaryoblastic leukemia |
| SNOMED CT | 277604002 | Acute eosinophilic leukemia |
| SNOMED CT | 277606000 | Lymphoreticular tumor |
| SNOMED CT | 277613000 | Cutaneous/peripheral T-cell lymphoma |
| SNOMED CT | 277614006 | Prethymic and thymic T-cell lymphoma/leukemia |
| SNOMED CT | 277615007 | Low grade B-cell lymphoma |
| SNOMED CT | 277616008 | Diffuse low grade B-cell lymphoma |
| SNOMED CT | 277617004 | High grade B-cell lymphoma |
| SNOMED CT | 277618009 | Follicular low grade B-cell lymphoma |
| SNOMED CT | 277619001 | B-cell prolymphocytic leukemia |
| SNOMED CT | 277622004 | Mucosa-associated lymphoma |
| SNOMED CT | 277623009 | Monocytoid B-cell lymphoma |
| SNOMED CT | 277624003 | Follicular malignant lymphoma - mixed cell type |
| SNOMED CT | 277625002 | Follicular malignant lymphoma - small cleaved cell |
| SNOMED CT | 277626001 | Diffuse high grade B-cell lymphoma |
| SNOMED CT | 277627005 | Nodular high grade B-cell lymphoma |
| SNOMED CT | 277628000 | Diffuse malignant lymphoma - large cleaved cell |
| SNOMED CT | 277629008 | Diffuse malignant lymphoma - large non-cleaved cell |
| SNOMED CT | 277632006 | Diffuse malignant lymphoma - centroblastic polymorphic |

Continued on next page

**Table 7 – continued from previous page**

| CodeSystem | ConceptCode | ConceptName |
|---|---|---|
| SNOMED CT | 277637000 | Large cell anaplastic lymphoma |
| SNOMED CT | 277641001 | Follicular malignant lymphoma - large cell |
| SNOMED CT | 277642008 | Low grade T-cell lymphoma |
| SNOMED CT | 277643003 | High grade T-cell lymphoma |
| SNOMED CT | 277651000 | Peripheral T-cell lymphoma - pleomorphic small cell |
| SNOMED CT | 277653002 | Peripheral T-cell lymphoma - pleomorphic medium and large cell |
| SNOMED CT | 277664004 | Malignant lymphoma of testis |
| SNOMED CT | 278051002 | Malignant lymphoma of thyroid gland |
| SNOMED CT | 278052009 | Malignant lymphoma of breast |
| SNOMED CT | 278189009 | Hypergranular promyelocytic leukemia |
| SNOMED CT | 278256002 | Cord cell transfusion |
| SNOMED CT | 278453007 | Acute biphenotypic leukemia |
| SNOMED CT | 281388009 | HIV-related sclerosing cholangitis |
| SNOMED CT | 285769009 | Acute promyelocytic leukemia - hypogranular variant |
| SNOMED CT | 285776004 | Intermediate grade B-cell lymphoma |
| SNOMED CT | 285839005 | Acute myelomonocytic leukemia - eosinophilic variant |
| SNOMED CT | 287006005 | Rheumatoid arthritis of multiple joints |
| SNOMED CT | 287007001 | Rheumatoid arthritis - hand joint |
| SNOMED CT | 287008006 | Rheumatoid arthritis - ankle and/or foot |
| SNOMED CT | 288327009 | Granulation anomaly |
| SNOMED CT | 289317009 | Granulocyte granule deficiency |
| SNOMED CT | 302841002 | Malignant lymphoma - small lymphocytic |
| SNOMED CT | 302842009 | Diffuse malignant lymphoma - centroblastic |
| SNOMED CT | 302845006 | Nodular malignant lymphoma, lymphocytic - well differentiated |
| SNOMED CT | 302848008 | Nodular malignant lymphoma, lymphocytic - intermediate differentiation |
| SNOMED CT | 302855005 | Subacute leukemia |
| SNOMED CT | 302856006 | Aleukemic leukemia |
| SNOMED CT | 302874002 | Phagocytic cell dysfunction |
| SNOMED CT | 303017006 | Malignant lymphoma, convoluted cell type |
| SNOMED CT | 303055001 | Malignant lymphoma, follicular center cell |
| SNOMED CT | 303056000 | Malignant lymphoma, follicular center cell, cleaved |
| SNOMED CT | 303057009 | Malignant lymphoma, follicular center cell, non-cleaved |
| SNOMED CT | 304576008 | Metaphyseal chondrodysplasia, McKusick type with associated immunodeficiency |
| SNOMED CT | 306058006 | Aregenerative anemia |
| SNOMED CT | 307340003 | Monosomy 7 syndrome |
| SNOMED CT | 307341004 | Atypical hairy cell leukemia |
| SNOMED CT | 307592006 | Basophilic leukemia |
| SNOMED CT | 307617006 | Neutrophilic leukemia |
| SNOMED CT | 307622006 | Prolymphocytic lymphosarcoma |

Continued on next page

**Table 7 – continued from previous page**

| CodeSystem | ConceptCode | ConceptName |
|---|---|---|
| SNOMED CT | 307623001 | Malignant lymphoma - lymphoplasmacytic |
| SNOMED CT | 307624007 | Diffuse malignant lymphoma - centroblastic-centrocytic |
| SNOMED CT | 307625008 | Malignant lymphoma - centrocytic |
| SNOMED CT | 307636001 | Malignant lymphoma, mixed lymphocytic-histiocytic, nodular |
| SNOMED CT | 307637005 | Malignant lymphoma, centroblastic-centrocytic, follicular |
| SNOMED CT | 307646004 | Malignant lymphoma, lymphocytic, poorly differentiated, nodular |
| SNOMED CT | 307647008 | Malignant lymphoma, centroblastic type, follicular |
| SNOMED CT | 307649006 | Primary central nervous system lymphoma |
| SNOMED CT | 307650006 | Histiocytic medullary reticulosis |
| SNOMED CT | 307651005 | Myelosclerosis with myeloid metaplasia |
| SNOMED CT | 308143008 | Seropositive erosive rheumatoid arthritis |
| SNOMED CT | 309762007 | Systemic lupus erythematosus with pericarditis |
| SNOMED CT | 313427003 | Lambda light chain myeloma |
| SNOMED CT | 314922006 | B-cell lymphoma morphology |
| SNOMED CT | 314923001 | Low grade B-cell lymphoma morphology |
| SNOMED CT | 314924007 | Follicular low grade B-cell lymphoma morphology |
| SNOMED CT | 314925008 | High grade B-cell lymphoma morphology |
| SNOMED CT | 314926009 | T-cell lymphoma morphology |
| SNOMED CT | 314927000 | High grade T-cell lymphoma morphology |
| SNOMED CT | 314929002 | Diffuse low grade B-cell lymphoma morphology |
| SNOMED CT | 314930007 | Low grade T-cell lymphoma morphology |
| SNOMED CT | 314931006 | Nodular high grade B-cell lymphoma morphology |
| SNOMED CT | 314934003 | Diffuse high grade B-cell lymphoma morphology |
| SNOMED CT | 317226009 | Granulocytosis |
| SNOMED CT | 319171004 | Qualitative abnormality of granulocyte |
| SNOMED CT | 351287008 | Reticular dysgenesis with congenital aleukocytosis |
| SNOMED CT | 363009005 | Complement component deficiency |
| SNOMED CT | 363040003 | Congenital immunodeficiency involving the hematopoietic system |
| SNOMED CT | 363138005 | Hereditary disorder of immune system |
| SNOMED CT | 363153002 | Immune system transplantation |
| SNOMED CT | 363265005 | Predominant humoral immune deficiency |
| SNOMED CT | 370388006 | Patient immunocompromised |
| SNOMED CT | 370391006 | Patient immunosuppressed |
| SNOMED CT | 371012000 | Acute lymphoblastic leukemia, transitional pre-B-cell |
| SNOMED CT | 371134001 | Malignant lymphoma, large cell, polymorphous, immunoblastic |
| SNOMED CT | 373168002 | Reticulosarcoma |
| SNOMED CT | 373604002 | Light chain deposition disease |
| SNOMED CT | 387759001 | Chronic granulomatous disease |
| SNOMED CT | 393573009 | Hypereosinophilic syndrome |

Continued on next page

**Table 7 – continued from previous page**

| CodeSystem | ConceptCode | ConceptName |
|---|---|---|
| SNOMED CT | 395692003 | Benign paraproteinemia |
| SNOMED CT | 397007003 | Mast cell disorder |
| SNOMED CT | 397008008 | Aggressive lymphadenopathic mastocytosis with eosinophilia |
| SNOMED CT | 397009000 | Mast cell malignancy |
| SNOMED CT | 397011009 | Mast cell malignancy of lymph nodes |
| SNOMED CT | 397015000 | Systemic mastocytosis with associated clonal hematological non-mast cell lineage disease |
| SNOMED CT | 397016004 | Systemic mast cell disease |
| SNOMED CT | 397338009 | Myelodysplastic syndrome with excess blasts-1 |
| SNOMED CT | 397339001 | Myelodysplastic syndrome with excess blasts-2 |
| SNOMED CT | 397340004 | Acute myeloid leukemia with recurrent genetic abnormality |
| SNOMED CT | 397341000 | Acute myeloid leukemia with multilineage dysplasia following a myelodysplastic syndrome or myelodysplastic syndrome/myeloproliferative disorder |
| SNOMED CT | 397342007 | Acute myeloid leukemia with multilineage dysplasia without antecedent myelodysplastic syndrome |
| SNOMED CT | 397343002 | Therapy-related acute myeloid leukemia and myelodysplastic syndrome, alkylating agent-related type |
| SNOMED CT | 397344008 | Therapy-related acute myeloid leukemia and myelodysplastic syndrome, topoisomerase type II inhibitor-related type |
| SNOMED CT | 397345009 | Acute leukemia of ambiguous lineage |
| SNOMED CT | 397349003 | Nodal marginal zone B-cell lymphoma |
| SNOMED CT | 397350003 | Extranodal marginal zone B-cell lymphoma of mucosa-associated lymphoid tissue |
| SNOMED CT | 397352006 | Primary cutaneous anaplastic large cell lymphoma |
| SNOMED CT | 397355008 | Dendritic cell sarcoma, no International Classification of Diseases for Oncology subtype |
| SNOMED CT | 397400006 | Burkitt lymphoma/leukemia |
| SNOMED CT | 397467006 | Follicular lymphoma, cutaneous follicle center sub-type |
| SNOMED CT | 397468001 | Follicular lymphoma, diffuse follicle center sub-type, grade 1 |
| SNOMED CT | 397469009 | Follicular lymphoma, diffuse follicle center cell sub-type, grade 2 |
| SNOMED CT | 397763006 | Human immunodeficiency virus encephalopathy |
| SNOMED CT | 398055000 | T-lymphocyte deficiency |
| SNOMED CT | 398250003 | Familial hemophagocytic lymphohistiocytosis |
| SNOMED CT | 398271008 | Predominantly T-cell defect |
| SNOMED CT | 398293003 | Cellular immune defect |
| SNOMED CT | 398329009 | Human immunodeficiency virus encephalitis |
| SNOMED CT | 398623004 | Myelodysplastic syndrome with excess blasts |
| SNOMED CT | 398640008 | Rheumatoid arthritis with pneumoconiosis |
| SNOMED CT | 399648005 | Intravascular B-cell lymphoma |

Continued on next page

**Table 7 – continued from previous page**

| CodeSystem | ConceptCode | ConceptName |
|---|---|---|
| SNOMED CT | 399923009 | Rheumatoid arteritis |
| SNOMED CT | 400001003 | Primary cutaneous lymphoma |
| SNOMED CT | 400122007 | Primary cutaneous T-cell lymphoma |
| SNOMED CT | 402355000 | Acute graft-versus-host disease |
| SNOMED CT | 402356004 | Chronic graft-versus-host disease |
| SNOMED CT | 402453006 | Amyloid light chain amyloidosis due to multiple myeloma |
| SNOMED CT | 402454000 | Primary systemic amyloidosis associated with occult plasma cell dyscrasia |
| SNOMED CT | 402483002 | Immunodeficiency with multicarboxylase deficiency |
| SNOMED CT | 402791005 | B-lymphocyte immunodeficiency |
| SNOMED CT | 402792003 | T-lymphocyte immunodeficiency |
| SNOMED CT | 402793008 | Heritable disorder of neutrophil production |
| SNOMED CT | 402794002 | Heritable disorder of neutrophil function |
| SNOMED CT | 402879006 | T-cell leukemic infiltration of skin |
| SNOMED CT | 402880009 | Primary cutaneous large T-cell lymphoma |
| SNOMED CT | 402882001 | Hodgkin's disease affecting skin |
| SNOMED CT | 403486000 | Acute systemic lupus erythematosus |
| SNOMED CT | 403487009 | Fulminating systemic lupus erythematosus |
| SNOMED CT | 403488004 | Systemic lupus erythematosus of childhood |
| SNOMED CT | 403836001 | Autosomal recessive hyperimmunoglobulin M syndrome |
| SNOMED CT | 403837005 | Wiskott-Aldrich autosomal dominant variant syndrome |
| SNOMED CT | 403838000 | Jung syndrome |
| SNOMED CT | 403839008 | Siccardi syndrome |
| SNOMED CT | 404107008 | Patch/plaque stage mycosis fungoides |
| SNOMED CT | 404108003 | Poikilodermatous mycosis fungoides |
| SNOMED CT | 404109006 | Follicular mucinosis type mycosis fungoides |
| SNOMED CT | 404110001 | Hypomelanotic mycosis fungoides |
| SNOMED CT | 404111002 | Lymphomatoid papulosis-associated mycosis fungoides |
| SNOMED CT | 404112009 | Granulomatous mycosis fungoides |
| SNOMED CT | 404113004 | Tumor stage mycosis fungoides |
| SNOMED CT | 404114005 | Erythrodermic mycosis fungoides |
| SNOMED CT | 404115006 | Bullous mycosis fungoides |
| SNOMED CT | 404116007 | Mycosis fungoides with systemic infiltration |
| SNOMED CT | 404117003 | Spongiotic mycosis fungoides |
| SNOMED CT | 404118008 | Syringotropic mycosis fungoides |
| SNOMED CT | 404119000 | Pagetoid reticulosis |
| SNOMED CT | 404120006 | Localized pagetoid reticulosis |
| SNOMED CT | 404121005 | Generalized pagetoid reticulosis |
| SNOMED CT | 404126000 | CD-30 positive pleomorphic large T-cell cutaneous lymphoma |
| SNOMED CT | 404127009 | CD-30 positive T-immunoblastic cutaneous lymphoma |
| SNOMED CT | 404128004 | CD-30 negative cutaneous T-cell lymphoma |
| SNOMED CT | 404129007 | CD-30 negative anaplastic large T-cell cutaneous lymphoma |

Continued on next page

**Table 7 – continued from previous page**

| CodeSystem | ConceptCode | ConceptName |
|---|---|---|
| SNOMED CT | 404130002 | CD-30 negative pleomorphic large T-cell cutaneous lymphoma |
| SNOMED CT | 404131003 | CD-30 negative T-immunoblastic cutaneous lymphoma |
| SNOMED CT | 404133000 | Subcutaneous panniculitis-like T-cell lymphoma |
| SNOMED CT | 404134006 | Anaplastic large T-cell systemic malignant lymphoma |
| SNOMED CT | 404137004 | Precursor B-cell lymphoblastic lymphoma involving skin |
| SNOMED CT | 404138009 | Small lymphocytic B-cell lymphoma involving skin |
| SNOMED CT | 404139001 | Leukemic infiltration of skin in hairy-cell leukemia |
| SNOMED CT | 404140004 | Primary cutaneous marginal zone B-cell lymphoma |
| SNOMED CT | 404141000 | Primary cutaneous immunocytoma |
| SNOMED CT | 404142007 | Primary cutaneous plasmacytoma |
| SNOMED CT | 404144008 | Primary cutaneous diffuse large cell B-cell lymphoma |
| SNOMED CT | 404147001 | Follicular center B-cell lymphoma (nodal/systemic with skin involvement) |
| SNOMED CT | 404148006 | Diffuse large B-cell lymphoma (nodal/systemic with skin involvement) |
| SNOMED CT | 404149003 | Lymphoplasmacytic B-cell lymphoma, nodal/systemic with skin involvement |
| SNOMED CT | 404150003 | Mantle cell B-cell lymphoma (nodal/systemic with skin involvement) |
| SNOMED CT | 404151004 | Leukemic infiltration of skin in myeloid leukemia |
| SNOMED CT | 404152006 | Leukemic infiltration of skin in acute myeloid leukemia |
| SNOMED CT | 404153001 | Leukemic infiltration of skin in chronic myeloid leukemia |
| SNOMED CT | 404154007 | Leukemic infiltration of skin in monocytic leukemia |
| SNOMED CT | 404155008 | Granulocytic sarcoma affecting skin |
| SNOMED CT | 404156009 | Leukemic infiltration of skin |
| SNOMED CT | 404160007 | Langerhans cell histiocytosis - Hashimoto-Pritzker type |
| SNOMED CT | 404169008 | Malignant histiocytosis involving skin |
| SNOMED CT | 404171008 | Mastocytoma |
| SNOMED CT | 404172001 | Mast cell leukemia affecting skin |
| SNOMED CT | 405631006 | Pediatric human immunodeficiency virus infection |
| SNOMED CT | 406565005 | CD4 T lymphocyte deficiency |
| SNOMED CT | 413389003 | Accelerated phase chronic myeloid leukemia |
| SNOMED CT | 413440007 | Acute lymphoblastic leukemia - category |
| SNOMED CT | 413441006 | Acute monocytic leukemia |
| SNOMED CT | 413442004 | Acute monocytic/monoblastic leukemia |
| SNOMED CT | 413443009 | Acute myeloid leukemia |
| SNOMED CT | 413537009 | Angioimmunoblastic T-cell lymphoma |
| SNOMED CT | 413565006 | Aplastic anemia associated with metabolic alteration |
| SNOMED CT | 413566007 | Aplastic anemia associated with pancreatitis |
| SNOMED CT | 413567003 | Aplastic anemia associated with pregnancy |
| SNOMED CT | 413587002 | Smoldering myeloma |
| SNOMED CT | 413656006 | Blastic phase chronic myeloid leukemia |

Continued on next page

**Table 7 – continued from previous page**

| CodeSystem | ConceptCode | ConceptName |
|---|---|---|
| SNOMED CT | 413834006 | Chronic disease of immune function |
| SNOMED CT | 413836008 | Chronic eosinophilic leukemia |
| SNOMED CT | 413840004 | Chronic lymphoid leukemia - category |
| SNOMED CT | 413841000 | Chronic myeloid leukemia - category |
| SNOMED CT | 413842007 | Chronic myeloid leukemia in lymphoid blast crisis |
| SNOMED CT | 413843002 | Chronic myeloid leukemia in myeloid blast crisis |
| SNOMED CT | 413847001 | Chronic phase chronic myeloid leukemia |
| SNOMED CT | 413990004 | Diffuse large B-cell lymphoma - category |
| SNOMED CT | 414029004 | Disorder of immune function |
| SNOMED CT | 414031008 | Immunosecretory disorder |
| SNOMED CT | 414395005 | Hereditary white blood cell disorder |
| SNOMED CT | 414553000 | Kappa light chain myeloma |
| SNOMED CT | 414644002 | Malignant hematopoietic neoplasm |
| SNOMED CT | 414645001 | Malignant histiocytic neoplasm |
| SNOMED CT | 414653009 | Mast cell neoplasm |
| SNOMED CT | 414785000 | Multiple solitary plasmacytomas |
| SNOMED CT | 414825006 | Neoplasm of hematopoietic cell type |
| SNOMED CT | 414927004 | Ocular myasthenia with strabismus |
| SNOMED CT | 415005004 | Panleukopenia |
| SNOMED CT | 415110002 | Plasma cell myeloma/plasmacytoma |
| SNOMED CT | 415111003 | Plasma cell neoplasm |
| SNOMED CT | 415112005 | Plasmacytoma |
| SNOMED CT | 415113000 | Plasmacytoma - category |
| SNOMED CT | 415283002 | Myelodysplastic syndrome with excess blasts-1 |
| SNOMED CT | 415284008 | Myelodysplastic syndrome with excess blasts-2 |
| SNOMED CT | 415285009 | Myelodysplastic syndrome with multilineage dysplasia |
| SNOMED CT | 415286005 | Myelodysplastic syndrome with ring sideroblasts and multilineage dysplasia |
| SNOMED CT | 415287001 | Relapsing chronic myeloid leukemia |
| SNOMED CT | 417152008 | Langerhans cell histiocytosis of lung |
| SNOMED CT | 417167007 | Immunoglobulin deficiency |
| SNOMED CT | 417672002 | Granulocytopenic disorder |
| SNOMED CT | 418265009 | Primary cutaneous B-cell lymphoma - category |
| SNOMED CT | 418628003 | Follicular mycosis fungoides |
| SNOMED CT | 418789003 | Primary cutaneous plasmacytoma |
| SNOMED CT | 419018000 | Primary cutaneous large T-cell lymphoma - category |
| SNOMED CT | 419283005 | Primary cutaneous T-cell lymphoma - category |
| SNOMED CT | 419386004 | Pagetoid reticulosis |
| SNOMED CT | 419392005 | Primary cutaneous lymphoma |
| SNOMED CT | 419586003 | Primary cutaneous T-cell lymphoma, large cell, CD30-negative |
| SNOMED CT | 419662008 | Primary cutaneous follicle center cell lymphoma |
| SNOMED CT | 420028002 | Primary cutaneous marginal zone B-cell lymphoma |

Continued on next page

**Table 7 – continued from previous page**

| CodeSystem | ConceptCode | ConceptName |
|---|---|---|
| SNOMED CT | 420384005 | Hematopoietic system disease with AIDS (acquired immunodeficiency syndrome) |
| SNOMED CT | 420395004 | Acute endocarditis with AIDS (acquired immunodeficiency syndrome) |
| SNOMED CT | 420519005 | Malignant lymphoma of the eye region |
| SNOMED CT | 420524008 | Kaposi's sarcoma with AIDS (acquired immunodeficiency syndrome) |
| SNOMED CT | 420788006 | Primary intraocular non-Hodgkin malignant lymphoma |
| SNOMED CT | 420890002 | Precursor T cell lymphoblastic leukemia/lymphoblastic lymphoma |
| SNOMED CT | 421020000 | Microsporidiosis associated with acquired immunodeficiency syndrome |
| SNOMED CT | 421077004 | Disseminated candidiasis with AIDS (acquired immunodeficiency syndrome) |
| SNOMED CT | 421246008 | Precursor T-cell lymphoblastic lymphoma |
| SNOMED CT | 421696004 | Precursor T-cell neoplasm |
| SNOMED CT | 422127002 | Herpes zoster with AIDS (acquired immunodeficiency syndrome) |
| SNOMED CT | 422172005 | T-cell AND/OR NK-cell neoplasm |
| SNOMED CT | 422853008 | Lymphoma of retroperitoneal space |
| SNOMED CT | 425333006 | Myeloproliferative disorder |
| SNOMED CT | 425563003 | Grafting of cord blood to bone marrow |
| SNOMED CT | 425657001 | Osteosclerotic myeloma |
| SNOMED CT | 425688002 | Philadelphia chromosome-positive acute lymphoblastic leukemia |
| SNOMED CT | 425749006 | Subacute myeloid leukemia in remission |
| SNOMED CT | 425835006 | Disorder of immune reconstitution |
| SNOMED CT | 425843001 | Allogeneic peripheral blood stem cell transplant |
| SNOMED CT | 425869007 | Acute promyelocytic leukemia, FAB M3, in remission |
| SNOMED CT | 425941003 | Precursor B-cell acute lymphoblastic leukemia in remission |
| SNOMED CT | 425983008 | Autologous peripheral blood stem cell transplant |
| SNOMED CT | 426071002 | Hodgkin's disease in remission |
| SNOMED CT | 426124006 | Acute myeloid leukemia with maturation, FAB M2, in remission |
| SNOMED CT | 426202004 | Immune reconstitution syndrome |
| SNOMED CT | 426217000 | Aleukaemic leukaemia in remission |
| SNOMED CT | 426248008 | Aleukemic lymphoid leukemia in remission |
| SNOMED CT | 426336007 | Solitary osseous myeloma |
| SNOMED CT | 426370008 | Subacute lymphoid leukemia in remission |
| SNOMED CT | 426425001 | Allograft of bone marrow from sibling donor |
| SNOMED CT | 426598005 | Amyloid light-chain nephropathy |
| SNOMED CT | 426642002 | Erythroleukemia, FAB M6 in remission |
| SNOMED CT | 426768001 | Engraftment syndrome |

Continued on next page

**Table 7 – continued from previous page**

| CodeSystem | ConceptCode | ConceptName |
|---|---|---|
| SNOMED CT | 426800001 | Febrile granulocytopenia |
| SNOMED CT | 426885008 | Hodgkin's disease, lymphocytic depletion of lymph nodes of head |
| SNOMED CT | 426955004 | Philadelphia chromosome-positive acute lymphoblastic leukemia |
| SNOMED CT | 427056005 | Subacute leukemia in remission |
| SNOMED CT | 427141003 | Malignant lymphoma in remission |
| SNOMED CT | 427374007 | Immunoproliferative neoplasm in remission |
| SNOMED CT | 427423003 | Allograft of bone marrow from matched unrelated donor |
| SNOMED CT | 427615008 | Allograft of cord blood to bone marrow |
| SNOMED CT | 427642009 | T-cell acute lymphoblastic leukemia in remission |
| SNOMED CT | 427658007 | Acute myelomonocytic leukemia, FAB M4, in remission |
| SNOMED CT | 427770001 | Rheumatoid arthritis of temporomandibular joint |
| SNOMED CT | 428103008 | Disorder of transplanted bone marrow |
| SNOMED CT | 429054002 | Disorder related to transplantation |
| SNOMED CT | 429192004 | Rheumatoid arthritis of foot |
| SNOMED CT | 429362002 | Short stature associated with bone marrow transplant |
| SNOMED CT | 429450002 | Disorder related to bone marrow transplantation |
| SNOMED CT | 429490004 | Disorder affecting transplanted structure |
| SNOMED CT | 429514007 | Growth hormone deficiency after bone marrow transplant |
| SNOMED CT | 430338009 | Smoldering chronic lymphocytic leukemia |
| SNOMED CT | 430478003 | Macrophage activation syndrome |
| SNOMED CT | 431456004 | Accelerated rejection of bone marrow transplant |
| SNOMED CT | 431506006 | Rejection of pancreas transplant |
| SNOMED CT | 431862001 | Acute rejection of bone marrow transplant |
| SNOMED CT | 431953002 | Chronic rejection of bone marrow transplant |
| SNOMED CT | 432782005 | Hyperacute rejection of bone marrow transplant |
| SNOMED CT | 433592008 | Acute rejection of pancreas transplant |
| SNOMED CT | 433809002 | Accelerated rejection of pancreas transplant |
| SNOMED CT | 434202008 | Hyperacute rejection of pancreas transplant |
| SNOMED CT | 434271002 | Chronic rejection of pancreas transplant |
| SNOMED CT | 440422002 | Asymptomatic multiple myeloma |
| SNOMED CT | 441313008 | Indolent multiple myeloma |
| SNOMED CT | 441559006 | Mantle cell lymphoma of spleen |
| SNOMED CT | 441962003 | Large cell lymphoma of intrapelvic lymph nodes |
| SNOMED CT | 442134007 | Hepatitis B associated with Human immunodeficiency virus infection |
| SNOMED CT | 442459007 | Combined immunodeficiency disease |
| SNOMED CT | 442537007 | Non-Hodgkin lymphoma associated with Human immunodeficiency virus infection |
| SNOMED CT | 442557006 | Grafting of bone marrow using allograft from unmatched unrelated donor |
| SNOMED CT | 443487006 | Mantle cell lymphoma |

Continued on next page

**Table 7 – continued from previous page**

| CodeSystem | ConceptCode | ConceptName |
|---|---|---|
| SNOMED CT | 444597005 | Mucosa-associated lymphoid tissue (MALT) lymphoma of stomach |
| SNOMED CT | 444910004 | Primary mediastinal (thymic) large B-cell lymphoma |
| SNOMED CT | 444911000 | Acute myeloid leukemia with t(9:11)(p22;q23); MLLT3-MLL |
| SNOMED CT | 445105005 | Blastic plasmacytoid dendritic cell neoplasm |
| SNOMED CT | 445227008 | Juvenile myelomonocytic leukemia |
| SNOMED CT | 445406001 | Hepatosplenic T-cell lymphoma |
| SNOMED CT | 445448008 | Acute myeloid leukemia with myelodysplasia-related changes |
| SNOMED CT | 445738007 | Myelodysplastic/myeloproliferative disease |
| SNOMED CT | 445757003 | Allogeneic bone marrow transplantation without purging |
| SNOMED CT | 445925008 | Lymphomatous infiltration |
| SNOMED CT | 445945000 | Infectious disease with acquired immune deficiency syndrome |
| SNOMED CT | 445964006 | Monomorphic posttransplant lymphoproliferative disorder |
| SNOMED CT | 446253009 | Allogeneic bone marrow transplantation with purging |
| SNOMED CT | 446643000 | Sarcoma of dendritic cells (accessory cells) |
| SNOMED CT | 446688004 | Leukemic infiltration |
| SNOMED CT | 447100004 | Marginal zone lymphoma |
| SNOMED CT | 447596005 | Myelodysplastic/myeloproliferative neoplasm, unclassifiable |
| SNOMED CT | 447656001 | Lymphoma of pylorus of stomach |
| SNOMED CT | 447658000 | Lymphoma of fundus of stomach |
| SNOMED CT | 447766003 | Lymphoma of pyloric antrum of stomach |
| SNOMED CT | 447805007 | Lymphoma of greater curvature of stomach |
| SNOMED CT | 447806008 | Lymphoma of cardia of stomach |
| SNOMED CT | 447989004 | Non-Hodgkin's lymphoma of extranodal site |
| SNOMED CT | 448212009 | Anaplastic large cell lymphoma, ALK negative |
| SNOMED CT | 448213004 | Diffuse non-Hodgkin's lymphoma of prostate |
| SNOMED CT | 448217003 | Follicular non-Hodgkin's lymphoma of prostate |
| SNOMED CT | 448220006 | Non-Hodgkin's lymphoma of bone |
| SNOMED CT | 448231003 | Follicular non-Hodgkin's lymphoma of nose |
| SNOMED CT | 448254007 | Non-Hodgkin's lymphoma of central nervous system |
| SNOMED CT | 448269008 | Lymphoma of lesser curvature of stomach |
| SNOMED CT | 448317000 | Follicular non-Hodgkin's lymphoma of soft tissue |
| SNOMED CT | 448319002 | Diffuse non-Hodgkin's lymphoma of nasopharynx |
| SNOMED CT | 448354009 | Non-Hodgkin's lymphoma of intestine |
| SNOMED CT | 448371005 | Non-Hodgkin's lymphoma of nasopharynx |
| SNOMED CT | 448372003 | Non-Hodgkin's lymphoma of lung |
| SNOMED CT | 448376000 | Non-Hodgkin's lymphoma of ovary |
| SNOMED CT | 448384001 | Non-Hodgkin's lymphoma of nose |
| SNOMED CT | 448386004 | Non-Hodgkin's lymphoma of oral cavity |
| SNOMED CT | 448387008 | Non-Hodgkin's lymphoma of testis |

Continued on next page

**Table 7 – continued from previous page**

| CodeSystem | ConceptCode | ConceptName |
|---|---|---|
| SNOMED CT | 448447004 | Non-Hodgkin's lymphoma of skin |
| SNOMED CT | 448465000 | Diffuse non-Hodgkin's lymphoma of testis |
| SNOMED CT | 448468003 | Diffuse non-Hodgkin's lymphoma of oral cavity |
| SNOMED CT | 448553002 | Lymphoma of pelvis |
| SNOMED CT | 448555009 | Lymphoma of body of stomach |
| SNOMED CT | 448560008 | Diffuse non-Hodgkin's lymphoma of extranodal site |
| SNOMED CT | 448561007 | Follicular non-Hodgkin's lymphoma of extranodal site |
| SNOMED CT | 448607004 | Diffuse non-Hodgkin's lymphoma of uterine cervix |
| SNOMED CT | 448609001 | Diffuse non-Hodgkin's lymphoma of ovary |
| SNOMED CT | 448663003 | Diffuse non-Hodgkin's lymphoma of stomach |
| SNOMED CT | 448666006 | Follicular non-Hodgkin's lymphoma of bone |
| SNOMED CT | 448672006 | Follicular non-Hodgkin's lymphoma of lung |
| SNOMED CT | 448709005 | Non-Hodgkin's lymphoma of stomach |
| SNOMED CT | 448738008 | Non-Hodgkin's lymphoma of soft tissue |
| SNOMED CT | 448774004 | Non-Hodgkin's lymphoma of uterine cervix |
| SNOMED CT | 448865007 | Follicular non-Hodgkin's lymphoma of skin |
| SNOMED CT | 448867004 | Diffuse non-Hodgkin's lymphoma of lung |
| SNOMED CT | 448995000 | Follicular non-Hodgkin's lymphoma of central nervous system |
| SNOMED CT | 449053004 | Lymphoma of lower esophagus |
| SNOMED CT | 449058008 | Follicular non-Hodgkin's lymphoma of tonsil |
| SNOMED CT | 449059000 | Follicular non-Hodgkin's lymphoma of uterine cervix |
| SNOMED CT | 449063007 | Follicular non-Hodgkin's lymphoma of oral cavity |
| SNOMED CT | 449065000 | Diffuse non-Hodgkin's lymphoma of nose |
| SNOMED CT | 449072004 | Gastrointestinal lymphoma |
| SNOMED CT | 449074003 | Lymphoma of small bowel |
| SNOMED CT | 449075002 | Lymphoma of cardioesophageal junction |
| SNOMED CT | 449108003 | Philadelphia chromosome positive chronic myelogenous leukemia |
| SNOMED CT | 449173006 | Diffuse non-Hodgkin's lymphoma of tonsil |
| SNOMED CT | 449176003 | Diffuse non-Hodgkin's lymphoma of intestine |
| SNOMED CT | 449177007 | Diffuse non-Hodgkin's lymphoma of bone |
| SNOMED CT | 449187006 | Kappa light chain deficiency |
| SNOMED CT | 449216004 | Diffuse non-Hodgkin's lymphoma of soft tissue |
| SNOMED CT | 449217008 | Diffuse non-Hodgkin's lymphoma of skin |
| SNOMED CT | 449218003 | Lymphoma of sigmoid colon |
| SNOMED CT | 449219006 | Follicular non-Hodgkin's lymphoma of nasopharynx |
| SNOMED CT | 449220000 | Diffuse follicle center lymphoma |
| SNOMED CT | 449221001 | Diffuse non-Hodgkin's lymphoma of central nervous system |
| SNOMED CT | 449222008 | Follicular non-Hodgkin's lymphoma of stomach |
| SNOMED CT | 449292003 | Non-Hodgkin's lymphoma of tonsil |
| SNOMED CT | 449307001 | Follicular non-Hodgkin's lymphoma of ovary |
| SNOMED CT | 449318001 | Non-Hodgkin's lymphoma of prostate |

Continued on next page

Table 7 – continued from previous page

| CodeSystem | ConceptCode | ConceptName |
|---|---|---|
| SNOMED CT | 449384005 | Lambda light chain deficiency |
| SNOMED CT | 449386007 | Philadelphia chromosome negative chronic myelogenous leukemia |
| SNOMED CT | 449418000 | Follicular non-Hodgkin's lymphoma of testis |
| SNOMED CT | 449419008 | Follicular non-Hodgkin's lymphoma of intestine |
| SNOMED CT | 449784008 | Diffuse infiltrative lymphocytosis syndrome |
| SNOMED CT | 449853003 | Interleukin-12 deficiency |
| SNOMED CT | 450907007 | Hydroa vacciniforme-like lymphoma |
| SNOMED CT | 450908002 | Primary cutaneous gamma-delta T-cell lymphoma |
| SNOMED CT | 450909005 | Plasmablastic lymphoma |
| SNOMED CT | 450910000 | ALK positive large B-cell lymphoma |
| SNOMED CT | 450911001 | Large B-cell lymphoma arising in HHV8 associated multicentric Castleman disease |
| SNOMED CT | 450912008 | Fibroblastic reticular cell tumor |
| SNOMED CT | 450913003 | Mixed phenotype acute leukemia |
| SNOMED CT | 450914009 | Mixed phenotype acute leukemia with t(9;22)(q34;q11.2); BCR-ABL1 |
| SNOMED CT | 450915005 | Mixed phenotype acute leukemia with t(v;11q23); MLL rearranged |
| SNOMED CT | 450916006 | Mixed phenotype acute leukemia B/myeloid |
| SNOMED CT | 450917002 | Mixed phenotype acute leukemia T/myeloid |
| SNOMED CT | 450920005 | B lymphoblastic leukemia / lymphoma - category |
| SNOMED CT | 450928003 | Acute myeloid leukemia with t(6;9)(p23;q34); DEK-NUP214 |
| SNOMED CT | 450929006 | Acute myeloid leukemia with inv(3)(q21q26.2) or t(3;3)(q21;q26.2); RPN1-EVI1 |
| SNOMED CT | 450935006 | Myeloid leukemia associated with Down Syndrome |
| SNOMED CT | 450937003 | Acute myeloid leukemia (megakaryoblastic) with t(1;22)(p13;q13); RBM15-MKL1 |
| SNOMED CT | 450940003 | Myeloid or lymphoid neoplasm with alpha-type platelet-derived growth factor receptor gene rearrangement |
| SNOMED CT | 450945008 | Refractory cytopenia with unilineage dysplasia |
| SNOMED CT | 450946009 | Refractory neutropenia |
| SNOMED CT | 450947000 | Refractory thrombocytopenia |
| SNOMED CT | 450950002 | B lymphoblastic leukemia lymphoma with t(9;22)(q34;q11.2); BCR-ABL1 |
| SNOMED CT | 450951003 | B lymphoblastic leukemia lymphoma with t(v;11q23); MLL rearranged |
| SNOMED CT | 450952005 | B lymphoblastic leukemia lymphoma with t(12;21)(p13;q22); TEL-AML1 (ETV6-RUNX1) |
| SNOMED CT | 450953000 | B lymphoblastic leukemia lymphoma with hyperdiploidy |
| SNOMED CT | 450954006 | B lymphoblastic leukemia lymphoma with hypodiploidy (Hypodiploid ALL) |

Continued on next page

**Table 7 – continued from previous page**

| CodeSystem | ConceptCode | ConceptName |
|---|---|---|
| SNOMED CT | 450955007 | B lymphoblastic leukemia lymphoma with t(5;14)(q31;q32); IL3-IGH |
| SNOMED CT | 450956008 | B lymphoblastic leukemia lymphoma with t(1;19)(q23;p13.3); E2A-PBX1 (TCF3-PBX1) |
| SNOMED CT | 450958009 | Malignant lymphoma, diffuse large B-cell, immunoblastic |
| SNOMED CT | 450959001 | T-cell/histiocyte rich large B-cell lymphoma |
| SNOMED CT | 697922003 | Pulmonary hypertension in Langerhans cell histiocytosis |
| SNOMED CT | 697965002 | Cholangitis with AIDS (acquired immunodeficiency syndrome) |
| SNOMED CT | 698075004 | Syngeneic peripheral blood stem cell transplantation |
| SNOMED CT | 698646006 | Acute monoblastic leukemia in remission |
| SNOMED CT | 698694005 | Systemic lupus erythematosus in remission |
| SNOMED CT | 699293000 | Ischemic myelofibrosis |
| SNOMED CT | 699537002 | Polyostotic sclerosing histiocytosis |
| SNOMED CT | 699657009 | Hepatosplenic gamma-delta cell lymphoma |
| SNOMED CT | 699818003 | T-cell large granular lymphocytosis |
| SNOMED CT | 700053002 | Sepsis with acquired immunodeficiency syndrome |
| SNOMED CT | 702444009 | Autoimmune lymphoproliferative syndrome |
| SNOMED CT | 702446006 | Core binding factor acute myeloid leukemia |
| SNOMED CT | 702476004 | Therapy-related myelodysplastic syndrome |
| SNOMED CT | 702785000 | Large cell (Ki-1 positive) lymphoma |
| SNOMED CT | 702786004 | Follicular non-Hodgkin's lymphoma diffuse follicle center sub-type grade 1 |
| SNOMED CT | 702977001 | Follicular non-Hodgkin's lymphoma diffuse follicle center cell sub-type grade 2 |
| SNOMED CT | 703387000 | Cytogenetically normal acute myeloid leukemia |
| SNOMED CT | 703525006 | Anhidrotic ectodermal dysplasia with immune deficiency |
| SNOMED CT | 703540008 | Majeed syndrome |
| SNOMED CT | 703626001 | Anaplastic large cell lymphoma, T/Null cell, primary systemic type |
| SNOMED CT | 703817002 | Myelodysplastic/myeloproliferative neoplasm with ring sideroblasts and thrombocytosis |
| SNOMED CT | 703818007 | Acute monoblastic and monocytic leukemia |
| SNOMED CT | 703819004 | Acute myeloid leukemia with mutation of CEBPA (CCAAT enhancer binding protein alpha) gene |
| SNOMED CT | 703820005 | Acute myeloid leukemia with mutated NPM1 |
| SNOMED CT | 703821009 | T lymphoblastic leukemia/lymphoma |
| SNOMED CT | 703822002 | Indeterminate dendritic cell tumor |
| SNOMED CT | 705061009 | Childhood myelodysplastic syndrome |
| SNOMED CT | 707152007 | Phagocytic immunodeficiency |
| SNOMED CT | 707431006 | Langerhans cell histiocytosis of diaper area |
| SNOMED CT | 707432004 | Langerhans cell histiocytosis of skin |
| SNOMED CT | 709115004 | Transplantation of autologous hematopoietic stem cell |

Continued on next page

**Table 7 – continued from previous page**

| CodeSystem | ConceptCode | ConceptName |
|---|---|---|
| SNOMED CT | 709465004 | Periodontitis co-occurrent with Chédiak-Higashi syndrome |
| SNOMED CT | 709471005 | Periodontitis co-occurrent with leukemia |
| SNOMED CT | 709535007 | Periodontitis co-occurrent with infantile genetic agranulocytosis |
| SNOMED CT | 709608008 | Periodontitis co-occurrent with acquired neutropenia |
| SNOMED CT | 710735009 | Periodontitis co-occurrent with leukocyte adhesion deficiency |
| SNOMED CT | 710926008 | Periodontitis co-occurrent with familial neutropenia |
| SNOMED CT | 710927004 | Periodontitis co-occurrent with cyclical neutropenia |
| SNOMED CT | 711164003 | STING-associated vasculopathy with onset in infancy |
| SNOMED CT | 711429001 | Transplantation of autologous progenitor cell |
| SNOMED CT | 711480000 | Activated PI3K-delta syndrome |
| SNOMED CT | 713260006 | Subacute adenoviral encephalitis co-occurrent with human immunodeficiency virus infection |
| SNOMED CT | 713275003 | Splenomegaly co-occurrent with human immunodeficiency virus infection |
| SNOMED CT | 713278001 | Neuralgia co-occurrent with human immunodeficiency virus infection |
| SNOMED CT | 713297001 | Candidiasis of esophagus co-occurrent with human immunodeficiency virus infection |
| SNOMED CT | 713298006 | Heart disease co-occurrent with human immunodeficiency virus infection |
| SNOMED CT | 713299003 | Disorder of eye proper co-occurrent with human immunodeficiency virus infection |
| SNOMED CT | 713300006 | Disorder of gastrointestinal tract co-occurrent with human immunodeficiency virus infection |
| SNOMED CT | 713316008 | Eruption of skin co-occurrent with human immunodeficiency virus infection |
| SNOMED CT | 713318009 | Myocarditis co-occurrent with human immunodeficiency virus infection |
| SNOMED CT | 713320007 | Radiculitis co-occurrent with human immunodeficiency virus infection |
| SNOMED CT | 713325002 | Primary cerebral lymphoma co-occurrent with human immunodeficiency virus infection |
| SNOMED CT | 713339002 | Infection caused by Strongyloides co-occurrent with human immunodeficiency virus infection |
| SNOMED CT | 713340000 | Disorder of skin co-occurrent with human immunodeficiency virus infection |
| SNOMED CT | 713341001 | Myelitis co-occurrent with human immunodeficiency virus infection |
| SNOMED CT | 713342008 | Infection caused by Salmonella co-occurrent with human immunodeficiency virus infection |

Continued on next page

**Table 7 – continued from previous page**

| CodeSystem | ConceptCode | ConceptName |
|---|---|---|
| SNOMED CT | 713349004 | Anemia co-occurrent with human immunodeficiency virus infection |
| SNOMED CT | 713444005 | Hemophagocytic syndrome with human immunodeficiency virus infection |
| SNOMED CT | 713445006 | Disseminated infection caused by Strongyloides co-occurrent with human immunodeficiency virus infection |
| SNOMED CT | 713446007 | Chronic infection caused by herpes simplex virus co-occurrent with human immunodeficiency virus infection |
| SNOMED CT | 713483007 | Reticulosarcoma co-occurrent with human immunodeficiency virus infection |
| SNOMED CT | 713484001 | Disorder of respiratory system co-occurrent with human immunodeficiency virus infection |
| SNOMED CT | 713487008 | Progressive multifocal leucoencephalopathy co-occurrent with human immunodeficiency virus infection |
| SNOMED CT | 713488003 | Presenile dementia co-occurrent with human immunodeficiency virus infection |
| SNOMED CT | 713489006 | Polyneuropathy co-occurrent with human immunodeficiency virus infection |
| SNOMED CT | 713490002 | Infection caused by Pneumocystis co-occurrent with human immunodeficiency virus infection |
| SNOMED CT | 713491003 | Organic brain syndrome co-occurrent with human immunodeficiency virus infection |
| SNOMED CT | 713497004 | Candidiasis of mouth co-occurrent with human immunodeficiency virus infection |
| SNOMED CT | 713503007 | Disorder of spinal cord co-occurrent with human immunodeficiency virus infection |
| SNOMED CT | 713504001 | Disorder of kidney co-occurrent with human immunodeficiency virus infection |
| SNOMED CT | 713505000 | Gastrointestinal malabsorption syndrome co-occurrent with human immunodeficiency virus infection |
| SNOMED CT | 713506004 | Neuritis co-occurrent with human immunodeficiency virus infection |
| SNOMED CT | 713507008 | Lymphadenopathy co-occurrent with human immunodeficiency virus infection |
| SNOMED CT | 713508003 | Aplastic anemia co-occurrent with human immunodeficiency virus infection |
| SNOMED CT | 713510001 | Enlargement of liver co-occurrent with human immunodeficiency virus infection |
| SNOMED CT | 713511002 | Acute endocarditis co-occurrent with human immunodeficiency virus infection |
| SNOMED CT | 713516007 | Primary effusion lymphoma |
| SNOMED CT | 713523008 | Cardiomyopathy co-occurrent with human immunodeficiency virus infection |

Continued on next page

**Table 7 – continued from previous page**

| CodeSystem | ConceptCode | ConceptName |
|---|---|---|
| SNOMED CT | 713526000 | Recurrent bacterial pneumonia co-occurrent with human immunodeficiency virus infection |
| SNOMED CT | 713527009 | Disorder of peripheral nervous system co-occurrent with human immunodeficiency virus infection |
| SNOMED CT | 713530002 | Agranulocytosis co-occurrent with human immunodeficiency virus infection |
| SNOMED CT | 713531003 | Visual impairment co-occurrent with human immunodeficiency virus infection |
| SNOMED CT | 713532005 | Infective arthritis co-occurrent with human immunodeficiency virus infection |
| SNOMED CT | 713533000 | Acquired hemolytic anemia co-occurrent with human immunodeficiency virus infection |
| SNOMED CT | 713543002 | Demyelinating disease of central nervous system co-occurrent with human immunodeficiency virus infection |
| SNOMED CT | 713544008 | Bacterial pneumonia co-occurrent with human immunodeficiency virus infection |
| SNOMED CT | 713545009 | Infection caused by Nocardia co-occurrent with human immunodeficiency virus infection |
| SNOMED CT | 713546005 | Isosporiasis co-occurrent with human immunodeficiency virus infection |
| SNOMED CT | 713570009 | Infectious gastroenteritis co-occurrent with human immunodeficiency virus infection |
| SNOMED CT | 713571008 | Disorder of central nervous system co-occurrent with human immunodeficiency virus infection |
| SNOMED CT | 713572001 | Malignant neoplastic disease co-occurrent with human immunodeficiency virus infection |
| SNOMED CT | 713695001 | Nephrotic syndrome co-occurrent with human immunodeficiency virus infection |
| SNOMED CT | 713696000 | Renal failure syndrome co-occurrent with human immunodeficiency virus infection |
| SNOMED CT | 713718006 | Diffuse non-Hodgkin immunoblastic lymphoma co-occurrent with human immunodeficiency virus infection |
| SNOMED CT | 713722001 | Infection caused by Cytomegalovirus co-occurrent with human immunodeficiency virus infection |
| SNOMED CT | 713729005 | Coccidiosis co-occurrent with human immunodeficiency virus infection |
| SNOMED CT | 713730000 | Infection caused by herpes simplex virus co-occurrent with human immunodeficiency virus infection |
| SNOMED CT | 713731001 | Pyrexia of unknown origin co-occurrent with human immunodeficiency virus infection |
| SNOMED CT | 713732008 | Aspergillosis co-occurrent with human immunodeficiency virus infection |

Continued on next page

**Table 7 – continued from previous page**

| CodeSystem | ConceptCode | ConceptName |
|---|---|---|
| SNOMED CT | 713733003 | Infection caused by herpes zoster virus co-occurrent with human immunodeficiency virus infection |
| SNOMED CT | 713734009 | Dermatophytosis co-occurrent with human immunodeficiency virus infection |
| SNOMED CT | 713742005 | Human immunodeficiency virus antibody positive |
| SNOMED CT | 713844000 | Dementia co-occurrent with human immunodeficiency virus infection |
| SNOMED CT | 713845004 | Infection caused by Cryptosporidium co-occurrent with human immunodeficiency virus infection |
| SNOMED CT | 713880000 | Opportunistic mycosis co-occurrent with human immunodeficiency virus infection |
| SNOMED CT | 713881001 | Microsporidiosis co-occurrent with human immunodeficiency virus infection |
| SNOMED CT | 713887002 | Focal segmental glomerulosclerosis co-occurrent with human immunodeficiency virus infection |
| SNOMED CT | 713897006 | Burkitt lymphoma co-occurrent with human immunodeficiency virus infection |
| SNOMED CT | 713910008 | Antibody mediated acquired pure red cell aplasia caused by erythropoiesis stimulating agent |
| SNOMED CT | 713964006 | Multidermatomal infection caused by Herpes zoster co-occurrent with human immunodeficiency virus infection |
| SNOMED CT | 713967004 | Disseminated atypical infection caused by Mycobacterium co-occurrent with human immunodeficiency virus infection |
| SNOMED CT | 714083007 | Recurrent salmonella sepsis co-occurrent with human immunodeficiency virus infection |
| SNOMED CT | 714251006 | Philadelphia chromosome-negative precursor B-cell acute lymphoblastic leukemia |
| SNOMED CT | 714463003 | Primary effusion lymphoma co-occurrent with infection caused by Human herpesvirus 8 |
| SNOMED CT | 714464009 | Immune reconstitution inflammatory syndrome caused by human immunodeficiency virus infection |
| SNOMED CT | 715664005 | Interdigitating dendritic cell sarcoma |
| SNOMED CT | 715950008 | ALK-positive large B-cell lymphoma |
| SNOMED CT | 715982006 | Severe combined immunodeficiency due to DCLRE1C deficiency |
| SNOMED CT | 716192009 | Thong Douglas Ferrante syndrome |
| SNOMED CT | 716378008 | Combined immunodeficiency due to ZAP70 deficiency |
| SNOMED CT | 716655008 | Aggressive systemic mastocytosis |
| SNOMED CT | 716788007 | Epstein-Barr virus positive diffuse large B-cell lymphoma of elderly |
| SNOMED CT | 716789004 | Epstein-Barr virus positive diffuse large B-cell lymphoma of elderly |

Continued on next page

**Table 7 – continued from previous page**

| CodeSystem | ConceptCode | ConceptName |
|---|---|---|
| SNOMED CT | 716869006 | Mendelian susceptibility to mycobacterial disease due to complete IL12RB1 deficiency |
| SNOMED CT | 716871006 | Severe combined immunodeficiency due to deoxyribonucleic acid dependent protein kinase catalytic subunit deficiency |
| SNOMED CT | 717811007 | Combined immunodeficiency due to CRAC (calcium release activated calcium) channel dysfunction |
| SNOMED CT | 718107000 | T-cell negative B-cell positive severe combined immunodeficiency due to JAK3 deficiency |
| SNOMED CT | 718200007 | Primary pulmonary lymphoma |
| SNOMED CT | 718230004 | Mendelian susceptibility to mycobacterial disease due to complete interferon gamma receptor 1 deficiency |
| SNOMED CT | 718232007 | Pyogenic bacterial infection due to deficiency of myeloid differentiation primary response 88 |
| SNOMED CT | 718614004 | TEMPI syndrome |
| SNOMED CT | 718717004 | Primary immunodeficiency syndrome due to p14 deficiency |
| SNOMED CT | 718882006 | X-linked severe congenital neutropenia |
| SNOMED CT | 719019000 | WT limb blood syndrome |
| SNOMED CT | 719156006 | X-linked intellectual disability with hypogammaglobulinemia and progressive neurological deterioration syndrome |
| SNOMED CT | 719522009 | Candidiasis of upper respiratory tract co-occurrent with human immunodeficiency virus infection |
| SNOMED CT | 719685004 | Absent thumb with short stature and immunodeficiency syndrome |
| SNOMED CT | 719789000 | Human immunodeficiency virus 2 antibody positive and Human immunodeficiency virus 1 antibody cross-reactivity |
| SNOMED CT | 719810000 | X-linked intellectual disability with seizure and psoriasis syndrome |
| SNOMED CT | 719814009 | X-linked mendelian susceptibility to mycobacterial disease |
| SNOMED CT | 719824001 | Vici syndrome |
| SNOMED CT | 719827008 | X-linked immunoneurologic disorder |
| SNOMED CT | 720520009 | Attenuated Chédiak-Higashi syndrome |
| SNOMED CT | 720853005 | Severe combined immunodeficiency, microcephaly, growth retardation, sensitivity to ionizing radiation syndrome |
| SNOMED CT | 720986005 | Anhidrotic ectodermal dysplasia, immunodeficiency, osteopetrosis, lymphedema syndrome |
| SNOMED CT | 721166000 | Human immunodeficiency virus complicating pregnancy childbirth and the puerperium |
| SNOMED CT | 721301004 | Myeloid and lymphoid neoplasm with fibroblast growth factor receptor 1 abnormality |
| SNOMED CT | 721302006 | Myelodysplastic/myeloproliferative neoplasm with ring sideroblasts and thrombocytosis |
| SNOMED CT | 721303001 | Refractory neutropenia |
| SNOMED CT | 721304007 | Refractory thrombocytopenia |

Continued on next page

**Table 7 – continued from previous page**

| CodeSystem | ConceptCode | ConceptName |
|---|---|---|
| SNOMED CT | 721305008 | Acute myeloid leukemia due to recurrent genetic abnormality |
| SNOMED CT | 721306009 | Therapy related acute myeloid leukemia and myelodysplastic syndrome |
| SNOMED CT | 721308005 | Acute leukemia of ambiguous lineage |
| SNOMED CT | 721310007 | Aggressive natural killer-cell leukemia |
| SNOMED CT | 721311006 | Systemic Epstein-Barr virus positive T-cell lymphoproliferative disease of childhood |
| SNOMED CT | 721313009 | Indeterminate dendritic cell neoplasm |
| SNOMED CT | 721314003 | Fibroblastic reticular cell neoplasm |
| SNOMED CT | 721555001 | Follicular lymphoma of small intestine |
| SNOMED CT | 721762007 | Adult T-cell leukemia/lymphoma of skin |
| SNOMED CT | 721876004 | Mendelian susceptibility to mycobacterial disease due to complete interferon gamma receptor 2 deficiency |
| SNOMED CT | 721877008 | Mendelian susceptibility to mycobacterial disease due to complete interleukin 12 subunit beta deficiency |
| SNOMED CT | 721903007 | Microcephaly, hypogammaglobulinemia, abnormal immunity syndrome |
| SNOMED CT | 721977007 | Lung fibrosis, immunodeficiency, 46,XX gonadal dysgenesis syndrome |
| SNOMED CT | 722067005 | Severe combined immunodeficiency with hypereosinophilia |
| SNOMED CT | 722281001 | Agammaglobulinemia, microcephaly, craniosynostosis, severe dermatitis syndrome |
| SNOMED CT | 722288007 | Autoimmune enteropathy and endocrinopathy with susceptibility to chronic infection syndrome |
| SNOMED CT | 722290008 | Autoimmune lymphoproliferative syndrome with recurrent viral infection |
| SNOMED CT | 722557007 | Parkinsonism due to human immunodeficiency virus infection |
| SNOMED CT | 722795004 | Meningeal leukemia |
| SNOMED CT | 722953004 | B-cell lymphoma unclassifiable with features intermediate between Burkitt lymphoma and diffuse large B-cell lymphoma |
| SNOMED CT | 722954005 | B-cell lymphoma unclassifiable with features intermediate between classical Hodgkin lymphoma and diffuse large B-cell lymphoma |
| SNOMED CT | 722955006 | Chronic lymphoproliferative disorder of natural killer cells |
| SNOMED CT | 722956007 | Early post-transplant lymphoproliferative disorder |
| SNOMED CT | 722957003 | Reactive plasmacytic hyperplasia post-transplant lymphoproliferative disorder |
| SNOMED CT | 722958008 | Infectious mononucleosis-like post-transplant lymphoproliferative disorder |
| SNOMED CT | 723334006 | FADD-related immunodeficiency |

Continued on next page

**Table 7 – continued from previous page**

| CodeSystem | ConceptCode | ConceptName |
|---|---|---|
| SNOMED CT | 723384004 | Mendelian susceptibility to mycobacterial disease due to complete ISG15 deficiency |
| SNOMED CT | 723385003 | Mendelian susceptibility to mycobacterial disease due to partial IRF8 deficiency |
| SNOMED CT | 723386002 | Mendelian susceptibility to mycobacterial disease due to partial STAT1 deficiency |
| SNOMED CT | 723443003 | Neutrophil immunodeficiency syndrome |
| SNOMED CT | 723508002 | RAS-associated autoimmune leukoproliferative disease |
| SNOMED CT | 723512008 | Revesz syndrome |
| SNOMED CT | 723889003 | B lymphoblastic leukemia lymphoma with t(9:22)(q34;q11.2); BCR-ABL 1 |
| SNOMED CT | 723995003 | Schimke immuno-osseous dysplasia |
| SNOMED CT | 724015007 | Pyogenic arthritis, pyoderma gangrenosum, acne syndrome |
| SNOMED CT | 724177005 | LIG4 syndrome |
| SNOMED CT | 724179008 | Laron syndrome with immunodeficiency |
| SNOMED CT | 724275005 | Primary immunodeficiency with natural killer cell deficiency and adrenal insufficiency |
| SNOMED CT | 724276006 | X-linked immune dysregulation, polyendocrinopathy, enteropathy syndrome |
| SNOMED CT | 724361001 | Hepatic veno-occlusive disease with immunodeficiency syndrome |
| SNOMED CT | 724642009 | Myeloid neoplasm associated with beta-type platelet-derived growth factor receptor gene rearrangement |
| SNOMED CT | 724644005 | Myeloid leukemia co-occurrent with Down syndrome |
| SNOMED CT | 724645006 | T-cell histiocyte rich large B-cell lymphoma |
| SNOMED CT | 724647003 | Diffuse large B-cell lymphoma co-occurrent with chronic inflammation caused by Epstein-Barr virus |
| SNOMED CT | 724648008 | Plasmablastic lymphoma |
| SNOMED CT | 724649000 | Langerhans cell sarcoma |
| SNOMED CT | 724650000 | Primary follicular dendritic cell sarcoma |
| SNOMED CT | 724767000 | Chorea co-occurrent and due to systemic lupus erythematosus |
| SNOMED CT | 724781003 | Demyelination of central nervous system co-occurrent and due to systemic lupus erythematosus |
| SNOMED CT | 725135004 | Combined immunodeficiency due to CD3gamma deficiency |
| SNOMED CT | 725136003 | Immunodeficiency by defective expression of human leukocyte antigen class 1 |
| SNOMED CT | 725137007 | Neutropenia, monocytopenia, deafness syndrome |
| SNOMED CT | 725150008 | Autosomal dominant mendelian susceptibility to mycobacterial disease due to partial interferon gamma receptor 1 deficiency |

Continued on next page

**Table 7 – continued from previous page**

| CodeSystem | ConceptCode | ConceptName |
|---|---|---|
| SNOMED CT | 725151007 | Autosomal dominant mendelian susceptibility to mycobacterial disease due to partial interferon gamma receptor 2 deficiency |
| SNOMED CT | 725290000 | Combined immunodeficiency due to partial RAG1 deficiency |
| SNOMED CT | 725390002 | Acute myeloid leukemia with t(8;16)(p11;p13) translocation |
| SNOMED CT | 725391003 | Acute myeloid leukemia with t(8;16)(p11;p13) translocation |
| SNOMED CT | 725431001 | Autosomal recessive mendelian susceptibility to mycobacterial disease due to partial interferon gamma receptor 1 deficiency |
| SNOMED CT | 725432008 | Autosomal recessive mendelian susceptibility to mycobacterial disease due to partial interferon gamma receptor 2 deficiency |
| SNOMED CT | 725437002 | Chronic lymphocytic leukemia genetic mutation variant |
| SNOMED CT | 726721002 | Nodal marginal zone B-cell lymphoma |
| SNOMED CT | 733028000 | Multiple sclerosis, ichthyosis, factor VIII deficiency syndrome |
| SNOMED CT | 733598001 | Acute myeloid leukemia with t(6;9)(p23;q34) translocation |
| SNOMED CT | 733834006 | Invasive carcinoma of uterine cervix co-occurrent with human immunodeficiency virus infection |
| SNOMED CT | 733835007 | Extrapulmonary tuberculosis co-occurrent with human immunodeficiency virus infection |
| SNOMED CT | 733860007 | Pediatric nodal marginal zone lymphoma |
| SNOMED CT | 733895005 | Primary cutaneous CD8 positive aggressive epidermotropic cytotoxic T-cell lymphoma |
| SNOMED CT | 733917002 | Pediatric follicular lymphoma |
| SNOMED CT | 734044003 | Anaplastic lymphoma kinase positive anaplastic large cell lymphoma |
| SNOMED CT | 734066005 | Diffuse large B-cell lymphoma of central nervous system |
| SNOMED CT | 734067001 | Splenic diffuse red pulp small B-cell lymphoma |
| SNOMED CT | 734076008 | Diffuse large B-cell lymphoma associated with chronic inflammation |
| SNOMED CT | 734141009 | Splenic B-cell lymphoma |
| SNOMED CT | 734142002 | B-cell lymphoma with features intermediate between diffuse large B-cell lymphoma and Burkitt lymphoma |
| SNOMED CT | 734522002 | Acute myeloid leukemia with FMS-like tyrosine kinase-3 mutation |
| SNOMED CT | 734524001 | Acute myeloid leukemia with FMS-like tyrosine kinase-3 mutation |
| SNOMED CT | 735332000 | Primary cutaneous diffuse large cell B-cell lymphoma leg type |
| SNOMED CT | 735436001 | Acquired disorder of neutrophil function |
| SNOMED CT | 735437005 | Disorder of neutrophil adhesion |

Continued on next page

**Table 7 – continued from previous page**

| CodeSystem | ConceptCode | ConceptName |
|---|---|---|
| SNOMED CT | 735438000 | Disorder of neutrophil chemotaxis |
| SNOMED CT | 735443007 | Acquired lymphocytopenia |
| SNOMED CT | 735521001 | Human immunodeficiency virus World Health Organization 2007 stage 1 co-occurrent with tuberculosis |
| SNOMED CT | 735522008 | Human immunodeficiency virus World Health Organization 2007 stage 1 co-occurrent with malaria |
| SNOMED CT | 735523003 | Human immunodeficiency virus World Health Organization 2007 stage 2 co-occurrent with tuberculosis |
| SNOMED CT | 735524009 | Human immunodeficiency virus World Health Organization 2007 stage 2 co-occurrent with malaria |
| SNOMED CT | 735525005 | Human immunodeficiency virus World Health Organization 2007 stage 3 co-occurrent with tuberculosis |
| SNOMED CT | 735526006 | Human immunodeficiency virus World Health Organization 2007 stage 3 co-occurrent with malaria |
| SNOMED CT | 735527002 | Human immunodeficiency virus World Health Organization 2007 stage 4 co-occurrent with tuberculosis |
| SNOMED CT | 735528007 | Human immunodeficiency virus World Health Organization 2007 stage 4 co-occurrent with malaria |
| SNOMED CT | 735536003 | Adult-onset immunodeficiency |
| SNOMED CT | 735599007 | Rheumatoid arthritis with erosion of joint |
| SNOMED CT | 735600005 | Rheumatoid arthritis without erosion |
| SNOMED CT | 736024007 | Pancytopenia caused by medication |
| SNOMED CT | 736322001 | Pediatric follicular lymphoma |
| SNOMED CT | 737223000 | Dendritic cell neoplasm |
| SNOMED CT | 737224006 | Histiocytic neoplasm |
| SNOMED CT | 737307003 | Natural-killer cell deficiency |
| SNOMED CT | 737378009 | World Health Organization 2007 Human immunodeficiency virus infection clinical stage 1 |
| SNOMED CT | 737379001 | World Health Organization 2007 Human immunodeficiency virus infection clinical stage 2 |
| SNOMED CT | 737380003 | World Health Organization 2007 Human immunodeficiency virus infection clinical stage 3 |
| SNOMED CT | 737381004 | World Health Organization 2007 Human immunodeficiency virus infection clinical stage 4 |
| SNOMED CT | 738527001 | Myeloid and/or lymphoid neoplasm associated with platelet derived growth factor receptor alpha rearrangement |
| SNOMED CT | 738770003 | Anaplastic lymphoma kinase positive anaplastic large cell lymphoma |
| SNOMED CT | 739301006 | Osteoporosis co-occurrent and due to multiple myeloma |
| SNOMED CT | 762315004 | Therapy related acute myeloid leukemia due to and following administration of antineoplastic agent |
| SNOMED CT | 762316003 | Polymorphic lymphoproliferative disorder following transplant |

Continued on next page

**Table 7 – continued from previous page**

| CodeSystem | ConceptCode | ConceptName |
|---|---|---|
| SNOMED CT | 762690000 | Classical Hodgkin lymphoma |
| SNOMED CT | 762691001 | Classical Hodgkin lymphoma |
| SNOMED CT | 763309005 | Acute myeloid leukemia with NPM1 somatic mutation |
| SNOMED CT | 763477007 | Primary lymphoma of conjunctiva |
| SNOMED CT | 763623001 | Severe combined immunodeficiency due to CTPS1 deficiency |
| SNOMED CT | 763666008 | Splenic marginal zone B-cell lymphoma |
| SNOMED CT | 763668009 | Lichtenstein syndrome |
| SNOMED CT | 763713000 | Idiopathic CD4 lymphocytopenia |
| SNOMED CT | 763719001 | Hydroa vacciniforme-like lymphoma |
| SNOMED CT | 763796007 | Megakaryoblastic acute myeloid leukemia with t(1;22)(p13;q13) |
| SNOMED CT | 763884007 | Splenic diffuse red pulp small B-cell lymphoma |
| SNOMED CT | 764855007 | Acute myeloid leukemia with CEBPA somatic mutations |
| SNOMED CT | 764858009 | Isolated agammaglobulinemia |
| SNOMED CT | 764940002 | Inherited acute myeloid leukemia |
| SNOMED CT | 764946008 | Constitutional mismatch repair deficiency syndrome |
| SNOMED CT | 765136002 | Primary cutaneous CD8 positive aggressive epidermotropic cytotoxic T-cell lymphoma |
| SNOMED CT | 765145001 | T cell negative B cell positive severe combined immunodeficiency due to gamma chain deficiency |
| SNOMED CT | 765188009 | Severe combined immunodeficiency due to complete RAG1 and/or RAG2 deficiency |
| SNOMED CT | 765327005 | Congenital sideroblastic anemia, B-cell immunodeficiency, periodic fever, developmental delay syndrome |
| SNOMED CT | 765328000 | Classic mycosis fungoides |
| SNOMED CT | 765748009 | Adult pure red cell aplasia |
| SNOMED CT | 766045006 | Acute myeloid leukemia and myelodysplastic syndrome related to alkylating agent |
| SNOMED CT | 766046007 | Acute myeloid leukemia and myelodysplastic syndrome related to topoisomerase type 2 inhibitor |
| SNOMED CT | 766048008 | Acute myeloid leukemia and myelodysplastic syndrome related to radiation |
| SNOMED CT | 766705006 | Immunodeficiency due to ficolin 3 deficiency |
| SNOMED CT | 766879006 | Combined immunodeficiency due to OX40 deficiency |
| SNOMED CT | 766935007 | Primary bone lymphoma |
| SNOMED CT | 766983005 | Susceptibility to respiratory infection associated with CD8alpha chain mutation |
| SNOMED CT | 767263007 | 22q11.2 deletion syndrome |
| SNOMED CT | 767658000 | Neutropenia due to and following chemotherapy |
| SNOMED CT | 768556005 | Ataxia pancytopenia syndrome |
| SNOMED CT | 768560008 | MDA5 deficiency |
| SNOMED CT | 770402000 | Aleukemic mast cell leukemia |

Continued on next page

**Table 7 – continued from previous page**

| CodeSystem | ConceptCode | ConceptName |
|---|---|---|
| SNOMED CT | 770596007 | Rippling muscle disease with myasthenia gravis |
| SNOMED CT | 770625006 | Combined immunodeficiency with faciooculoskeletal anomalies syndrome |
| SNOMED CT | 770687001 | Vasculitis due to adenosine deaminase 2 deficiency |
| SNOMED CT | 770785002 | T-cell immunodeficiency with epidermodysplasia verruciformis |
| SNOMED CT | 770942003 | Kostmann syndrome |
| SNOMED CT | 770947009 | Autosomal dominant severe congenital neutropenia |
| SNOMED CT | 771073006 | Lymphoproliferative disorder caused by methotrexate |
| SNOMED CT | 771078002 | Immunodeficiency due to MASP-2 deficiency |
| SNOMED CT | 771119002 | Infection caused by Coccidia co-occurrent with acquired immunodeficiency syndrome |
| SNOMED CT | 771126002 | Infection caused by Toxoplasma gondii co-occurrent with acquired immunodeficiency syndrome |
| SNOMED CT | 771127006 | Infection caused by Isospora co-occurrent with acquired immunodeficiency syndrome |
| SNOMED CT | 771309000 | Autosomal recessive lymphoproliferative disease |
| SNOMED CT | 771333006 | Immune dysregulation, inflammatory bowel disease, arthritis, recurrent infection syndrome |
| SNOMED CT | 771443008 | Complement component 3 deficiency |
| SNOMED CT | 771479000 | Combined immunodeficiency due to STK4 deficiency |
| SNOMED CT | 771515001 | Facial dysmorphism, immunodeficiency, livedo, short stature syndrome |
| SNOMED CT | 771517009 | Severe combined immunodeficiency due to CARD11 deficiency |
| SNOMED CT | 772126000 | Poikiloderma with neutropenia |
| SNOMED CT | 773333003 | Autosomal systemic lupus erythematosus |
| SNOMED CT | 773404000 | Roifman syndrome |
| SNOMED CT | 773488000 | Combined immunodeficiency due to MALT1 deficiency |
| SNOMED CT | 773537001 | Differentiation syndrome due to and following chemotherapy co-occurrent with acute promyelocytic leukemia |
| SNOMED CT | 773646003 | PLCG2-associated antibody deficiency and immune dysregulation |
| SNOMED CT | 773664005 | Deficiency in anterior pituitary function, variable immunodeficiency syndrome |
| SNOMED CT | 773702002 | Sterile multifocal osteomyelitis with periostitis and pustulosis |
| SNOMED CT | 773730002 | Osteopetrosis hypogammaglobulinemia syndrome |
| SNOMED CT | 773995001 | Primary cutaneous anaplastic large cell lymphoma |
| SNOMED CT | 774071007 | Pancytopenia with developmental delay syndrome |
| SNOMED CT | 774211005 | Severe dermatitis, multiple allergies, metabolic wasting syndrome |

Continued on next page

**Table 7 – continued from previous page**

| CodeSystem | ConceptCode | ConceptName |
|---|---|---|
| SNOMED CT | 775909002 | Congenital neutropenia, myelofibrosis, nephromegaly syndrome |
| SNOMED CT | 778004006 | Autoinflammation-PLCG2-associated antibody deficiency-immune dysregulation |
| SNOMED CT | 778023004 | Syndromic multisystem autoimmune disease due to ITCH deficiency |
| SNOMED CT | 778024005 | Monocytopenia with susceptibility to infections |
| SNOMED CT | 778027003 | Primary CD59 deficiency |
| SNOMED CT | 778028008 | Immunodeficiency due to CD25 deficiency |
| SNOMED CT | 778045003 | Susceptibility to viral and mycobacterial infection |
| SNOMED CT | 780817000 | Undifferentiated myeloproliferative disease |
| SNOMED CT | 780844005 | Acute myeloid leukemia with inv(3)(q21q26.2) or t(3;3)(q21;q26.2); RPN1-EVI1 |
| SNOMED CT | 782750002 | T-cell receptor alpha-beta-positive T-cell deficiency |
| SNOMED CT | 782751003 | Severe combined immunodeficiency due to IKK2 deficiency |
| SNOMED CT | 782759001 | X-linked dyserythropoietic anemia with abnormal platelets and neutropenia |
| SNOMED CT | 782915004 | Acquired hemophagocytic lymphohistiocytosis associated with malignant disease |
| SNOMED CT | 783007005 | Recurrent Neisseria infection due to factor D deficiency |
| SNOMED CT | 783017000 | Acute myeloid leukemia with BCR-ABL1 |
| SNOMED CT | 783058007 | Autosomal recessive severe congenital neutropenia due to G6PC3 deficiency |
| SNOMED CT | 783099001 | RIDDLE syndrome |
| SNOMED CT | 783142006 | Pancytopenia due to IKZF1 mutations |
| SNOMED CT | 783150002 | Iatrogenic immunodeficiency-associated lymphoproliferative disorder |
| SNOMED CT | 783199003 | Autosomal recessive severe congenital neutropenia due to JAGN1 deficiency |
| SNOMED CT | 783200000 | Autosomal recessive severe congenital neutropenia due to CXCR2 deficiency |
| SNOMED CT | 783201001 | Autosomal recessive severe congenital neutropenia due to CSF3R deficiency |
| SNOMED CT | 783220004 | Burkitt-like lymphoma with 11q aberration |
| SNOMED CT | 783245001 | Primary immunodeficiency with post-measles-mumps-rubella vaccine viral infection |
| SNOMED CT | 783248004 | Hyperimmunoglobulin M syndrome with susceptibility to opportunistic infection |
| SNOMED CT | 783249007 | Hyperimmunoglobulin M syndrome without susceptibility to opportunistic infection |
| SNOMED CT | 783255002 | Hereditary isolated aplastic anemia |
| SNOMED CT | 783263001 | Acute myeloid leukemia with mutated RUNX1 |
| SNOMED CT | 783541009 | Breast implant–associated anaplastic large-cell lymphoma |

Continued on next page

**Table 7 – continued from previous page**

| CodeSystem | ConceptCode | ConceptName |
|---|---|---|
| SNOMED CT | 783565007 | Indolent T-cell lymphoproliferative disorder of gastrointestinal tract |
| SNOMED CT | 783615009 | Erythropoietic uroporphyria associated with myeloid malignancy |
| SNOMED CT | 783617001 | Severe combined immunodeficiency due to LCK deficiency |
| SNOMED CT | 783621008 | Immunodeficiency with factor I anomaly |
| SNOMED CT | 783743009 | Combined immunodeficiency with granulomatosis |
| SNOMED CT | 783744003 | B-lymphoblastic leukemia lymphoma BCR-ABL1-like |
| SNOMED CT | 784339002 | Deficiency of interleukin 36 receptor antagonist |
| SNOMED CT | 784340000 | Combined immunodeficiency due to interleukin 21 receptor deficiency |
| SNOMED CT | 784392009 | Adult chronic idiopathic neutropenia |
| SNOMED CT | 784393004 | Adult-onset immunodeficiency with anti-interferon-gamma autoantibodies |
| SNOMED CT | 784551004 | Follicular T-cell lymphoma |
| SNOMED CT | 785825000 | B lymphoblastic leukemia lymphoma with iAMP21 |
| SNOMED CT | 786909001 | High grade B-cell lymphoma with MYC and BCL2 and/or BCL6 rearrangements |
| SNOMED CT | 786960000 | Large B-cell lymphoma with IRF4 rearrangement |
| SNOMED CT | 787036009 | Monomorphic epitheliotropic intestinal T-cell lymphoma |
| SNOMED CT | 787198005 | Primary cutaneous acral CD8 positive T-cell lymphoma |
| SNOMED CT | 787565006 | Diffuse large B-cell lymphoma activated B-cell subtype |
| SNOMED CT | 787594004 | Diffuse large B-cell lymphoma germinal center B-cell subtype |
| SNOMED CT | 787941004 | Myeloid and/or lymphoid neoplasm with PCM1-JAK2 |
| SNOMED CT | 788674000 | Primary cutaneous CD4 positive small/medium T-cell lymphoproliferative disorder |
| SNOMED CT | 788740009 | Acute myeloid leukemia with biallelic mutation of CEBPA (CCAAT enhancer binding protein alpha) gene |
| SNOMED CT | 788874003 | B-cell prolymphocytic leukemia in remission |
| SNOMED CT | 788972003 | Juvenile myelomonocytic leukemia in remission |
| SNOMED CT | 789435006 | Myelodysplastic syndrome with ring sideroblasts and multilineage dysplasia |
| SNOMED CT | 789689004 | Malignant lymphomatoid granulomatosis grade 3 |
| SNOMED CT | 789690008 | Malignant lymphomatoid granulomatosis grade 3 of lung |
| SNOMED CT | 789777007 | Short-limb skeletal dysplasia with severe combined immunodeficiency |
| SNOMED CT | 829973009 | Autoinflammatory syndrome with pyogenic bacterial infection and amylopectinosis |
| SNOMED CT | 830057003 | Relapsing classical Hodgkin lymphoma |
| SNOMED CT | 836276000 | Hodgkin sarcoma |
| SNOMED CT | 836277009 | Hodgkin granuloma |
| SNOMED CT | 836486002 | Lymphomatous infiltrate of kidney |

Continued on next page

**Table 7 – continued from previous page**

| CodeSystem | ConceptCode | ConceptName |
|---|---|---|
| SNOMED CT | 838340006 | B lymphoblastic leukemia lymphoma with t(5;14)(q31;q32); IL3-IGH |
| SNOMED CT | 838341005 | B lymphoblastic leukemia lymphoma with t(v;11q23); MLL rearranged |
| SNOMED CT | 838342003 | B lymphoblastic leukemia lymphoma with t(12;21) (p13;q22); TEL/AML1 (ETV6-RUNX1) |
| SNOMED CT | 838343008 | B lymphoblastic leukemia lymphoma with t(1;19)(Q23;P13.3); E2A-PBX1 (TCF3/PBX1) |
| SNOMED CT | 838344002 | B lymphoblastic leukemia lymphoma with hypodiploidy |
| SNOMED CT | 838346000 | B lymphoblastic leukemia lymphoma with hyperdiploidy |
| SNOMED CT | 838355002 | Acute myeloid leukemia with inv(16)(p13.1q22) or t(16;16)(p13.1;q22) CBFB-MYH11 |
| SNOMED CT | 838377003 | Chronic hepatitis C co-occurrent with human immunodeficiency virus infection |
| SNOMED CT | 840423002 | Diffuse large B-cell lymphoma of small intestine |
| SNOMED CT | 840424008 | Diffuse large B-cell lymphoma of stomach |
| SNOMED CT | 840442003 | Encephalitis caused by human immunodeficiency virus type 2 |
| SNOMED CT | 840472009 | Deficiency of immunoglobulin A with immunoglobulin G subclass deficiency |
| SNOMED CT | 840498003 | Encephalitis caused by human immunodeficiency virus type 1 |
| SNOMED CT | 866098005 | Large B-cell lymphoma arising in HHV8-associated multicentric Castleman disease |
| SNOMED CT | 866109006 | Lymphocytic hypereosinophilic syndrome |
| SNOMED CT | 871540009 | Post-polycythemia vera myelofibrosis |
| SNOMED CT | 871541008 | Post-essential thrombocythemia myelofibrosis |
| SNOMED CT | 81000119104 | Symptomatic human immunodeficiency virus infection |
| SNOMED CT | 12281000132104 | Relapsing acute myeloid leukemia |
| SNOMED CT | 12291000132102 | Refractory acute myeloid leukemia |
| SNOMED CT | 12301000132103 | Acute lymphoid leukemia relapse |
| SNOMED CT | 12311000132101 | Refractory acute lymphoid leukemia |
| SNOMED CT | 12341000132100 | B-cell lymphoma, unclassifiable, with features intermediate between diffuse large B-cell lymphoma and Burkitt lymphoma |
| SNOMED CT | 12351000132102 | B-cell lymphoma, unclassifiable, with features intermediate between diffuse large B-cell lymphoma and Hodgkin lymphoma |
| SNOMED CT | 12631000119106 | Immunoglobulin G deficiency |
| SNOMED CT | 61291000119103 | Disorder of central nervous system co-occurrent and due to acute lymphoid leukemia in remission |
| SNOMED CT | 61301000119102 | Disorder of central nervous system co-occurrent and due to acute lymphoid leukemia |

Continued on next page

**Table 7 – continued from previous page**

| CodeSystem | ConceptCode | ConceptName |
|---|---|---|
| SNOMED CT | 77461000119109 | Myasthenia gravis with exacerbation |
| SNOMED CT | 77471000119103 | Myasthenia gravis without exacerbation |
| SNOMED CT | 80191000119101 | Symptomatic human immunodeficiency virus (HIV) I infection |
| SNOMED CT | 90681000119107 | Asymptomatic human immunodeficiency virus A1 infection |
| SNOMED CT | 90691000119105 | Asymptomatic human immunodeficiency virus A2 infection |
| SNOMED CT | 116691000119101 | Marginal zone lymphoma of spleen |
| SNOMED CT | 116811000119106 | Non-Hodgkin lymphoma of central nervous system metastatic to lymph node of lower limb |
| SNOMED CT | 116821000119104 | Non-Hodgkin lymphoma of central nervous system metastatic to lymph node of upper limb |
| SNOMED CT | 116841000119105 | Marginal zone lymphoma of lymph nodes of multiple sites |
| SNOMED CT | 116871000119103 | Mantle cell lymphoma of lymph nodes of multiple sites |
| SNOMED CT | 117211000119105 | Peripheral T-cell lymphoma of lymph nodes of multiple sites |
| SNOMED CT | 118791000119106 | Aplastic anemia caused by antineoplastic agent |
| SNOMED CT | 121131000119109 | Chronic graft versus host disease after transplantation of bone marrow |
| SNOMED CT | 122491000119108 | Lymphoproliferative disorder after transplantation of bone marrow |
| SNOMED CT | 133751000119102 | Lymphoma of colon |
| SNOMED CT | 295101000119105 | Nephropathy co-occurrent and due to systemic lupus erythematosus |
| SNOMED CT | 295111000119108 | Nephrotic syndrome co-occurrent and due to systemic lupus erythematosus |
| SNOMED CT | 295121000119101 | Nephrosis co-occurrent and due to systemic lupus erythematosus |
| SNOMED CT | 319081000119109 | Seronegative rheumatoid arthritis of multiple joints |
| SNOMED CT | 319841000119107 | Rheumatoid lung disease with rheumatoid arthritis |
| SNOMED CT | 328301000119102 | Pancytopenia due to antineoplastic chemotherapy |
| SNOMED CT | 328611000119105 | Adult pulmonary Langerhans cell histiocytosis |
| SNOMED CT | 350951000119101 | B-cell lymphoma of intra-abdominal lymph nodes |
| SNOMED CT | 351211000119104 | B-cell lymphoma of lymph nodes of multiple sites |
| SNOMED CT | 352251000119109 | Small lymphocytic B-cell lymphoma of lymph nodes of multiple sites |
| SNOMED CT | 352411000119109 | Small lymphocytic B-cell lymphoma of intra-abdominal lymph nodes |
| SNOMED CT | 352791000119108 | Non-Hodgkin's lymphoma of lymph nodes of multiple sites |
| SNOMED CT | 354851000119101 | Follicular non-Hodgkin's lymphoma of lymph nodes of multiple sites |
| SNOMED CT | 1073731000119100 | Rheumatoid arthritis of left knee |
| SNOMED CT | 1073801000119100 | Rheumatoid arthritis of right hip |
| SNOMED CT | 10755671000119100 | Human immunodeficiency virus in mother complicating childbirth |

Continued on next page

**Table 7 – continued from previous page**

| CodeSystem | ConceptCode | ConceptName |
|---|---|---|
| SNOMED CT | 16623961000119100 | Pancytopenia caused by immunosuppressant |
| SNOMED CT | 1929004 | B-cell lymphoma, no ICD-O subtype |
| SNOMED CT | 3172003 | Peripheral T-cell lymphoma, no International Classification of Diseases for Oncology subtype |
| SNOMED CT | 4950009 | Sézary disease |
| SNOMED CT | 14317002 | AML (acute myeloid leukemia), M6 type |
| SNOMED CT | 14537002 | Hodgkin lymphoma, no International Classification of Diseases for Oncology subtype |
| SNOMED CT | 16893006 | [M]Hodgkin's disease, lymphocytic depletion, diffuse fibrosis |
| SNOMED CT | 17788007 | Acute myeloid leukemia, no International Classification of Diseases for Oncology subtype |
| SNOMED CT | 21964009 | Malignant lymphoma, no International Classification of Diseases for Oncology subtype |
| SNOMED CT | 22197008 | Burkitt cell leukaemia |
| SNOMED CT | 23719005 | Transfusion of bone marrow |
| SNOMED CT | 28054005 | Cutaneous lymphoma |
| SNOMED CT | 30575002 | Pancytopenia-dysmelia syndrome |
| SNOMED CT | 32280000 | Subacute lymphoid leukemia [obs] |
| SNOMED CT | 36070007 | Eczema, thrombocytopenia, immunodeficiency syndrome |
| SNOMED CT | 37810007 | Eosinophilic leukemia |
| SNOMED CT | 40411000 | Malignant lymphoma, lymphocytic, well differentiated, nodular [obs] |
| SNOMED CT | 46732000 | Angiotropic lymphoma |
| SNOMED CT | 46760003 | Estren-Dameshek anaemia |
| SNOMED CT | 50102004 | Malignant lymphoma, centroblastic-centrocytic, diffuse [obs] |
| SNOMED CT | 50926003 | Recurring cold staphylococcal abscesses |
| SNOMED CT | 53237008 | Large cell (Ki-1 positive) lymphoma |
| SNOMED CT | 55602000 | Congenital thymic dysplasia syndrome |
| SNOMED CT | 57160007 | Felty syndrome |
| SNOMED CT | 65399007 | Differentiated progressive histiocytosis |
| SNOMED CT | 65880007 | X linked agammaglobulinemia |
| SNOMED CT | 70600005 | Hodgkin paragranuloma, nodular [obs] |
| SNOMED CT | 71109004 | [M]Hodgkin's disease, lymphocytic depletion, reticular type |
| SNOMED CT | 74654000 | Mantle zone lymphoma [obs] |
| SNOMED CT | 77121009 | Familial fatal Epstein-Barr infection |
| SNOMED CT | 77381001 | Burkitt's tumor |
| SNOMED CT | 77430005 | [M]Adult T-cell leukemia/lymphoma |
| SNOMED CT | 79268002 | Polyneuropathy, organomegaly, endocrinopathy, monoclonal gammopathy, and skin changes syndrome |
| SNOMED CT | 82286005 | Immunodeficiency with IgM hypergammaglobulinemia |
| SNOMED CT | 87163000 | Leukemia, no International Classification of Diseases for Oncology subtype |

Continued on next page

**Table 7 – continued from previous page**

| CodeSystem | ConceptCode | ConceptName |
|---|---|---|
| SNOMED CT | 92511007 | Burkitt's tumor of lymph nodes of axilla and upper limb |
| SNOMED CT | 92512000 | Burkitt's tumor of lymph nodes of head, face, and neck |
| SNOMED CT | 92513005 | Burkitt's tumor of lymph nodes of inguinal region and lower limb |
| SNOMED CT | 92516002 | Burkitt's tumor of unspecified, extranodal, or solid organ site |
| SNOMED CT | 93451002 | Erythroleukaemia, FAB M6 |
| SNOMED CT | 93487009 | Hodgkin disease, lymphocytic depletion of lymph nodes of axilla AND/OR upper limb |
| SNOMED CT | 93488004 | Hodgkin disease, lymphocytic depletion of lymph nodes of head, face AND/OR neck |
| SNOMED CT | 93489007 | Hodgkin disease, lymphocytic depletion of lymph nodes of inguinal region AND/OR lower limb |
| SNOMED CT | 93492006 | Hodgkin disease, lymphocytic depletion of extranodal AND/OR solid organ site |
| SNOMED CT | 93493001 | Hodgkin's paragranuloma of intra-abdominal lymph nodes |
| SNOMED CT | 93494007 | Hodgkin disease, lymphocytic-histiocytic predominance of intrapelvic lymph nodes |
| SNOMED CT | 93495008 | Hodgkin disease, lymphocytic-histiocytic predominance of intrathoracic lymph nodes |
| SNOMED CT | 93496009 | Hodgkin disease, lymphocytic-histiocytic predominance of lymph nodes of axilla AND/OR upper limb |
| SNOMED CT | 93497000 | Hodgkin disease, lymphocytic-histiocytic predominance of lymph nodes of head, face AND/OR neck |
| SNOMED CT | 93498005 | Hodgkin disease, lymphocytic-histiocytic predominance of lymph nodes of inguinal region AND/OR lower limb |
| SNOMED CT | 93500006 | Hodgkin paragranuloma of spleen |
| SNOMED CT | 93501005 | Hodgkin disease, lymphocytic-histiocytic predominance of extranodal AND/OR solid organ site |
| SNOMED CT | 93505001 | Hodgkin disease, mixed cellularity of lymph nodes of axilla AND/OR upper limb |
| SNOMED CT | 93506000 | Hodgkin disease, mixed cellularity of lymph nodes of head, face AND/OR neck |
| SNOMED CT | 93507009 | Hodgkin disease, mixed cellularity of lymph nodes of inguinal region AND/OR lower limb |
| SNOMED CT | 93509007 | Hodgkin disease, mixed cellularity of spleen |
| SNOMED CT | 93510002 | Hodgkin disease, mixed cellularity of extranodal AND/OR solid organ site |
| SNOMED CT | 93514006 | Hodgkin disease, nodular sclerosis of lymph nodes of axilla AND/OR upper limb |
| SNOMED CT | 93515007 | Hodgkin disease, nodular sclerosis of lymph nodes of head, face AND/OR neck |

Continued on next page

**Table 7 – continued from previous page**

| CodeSystem | ConceptCode | ConceptName |
|---|---|---|
| SNOMED CT | 93516008 | Hodgkin disease, nodular sclerosis of lymph nodes of inguinal region AND/OR lower limb |
| SNOMED CT | 93518009 | Hodgkin disease, nodular sclerosis of spleen |
| SNOMED CT | 93519001 | Hodgkin disease, nodular sclerosis of extranodal AND/OR solid organ site |
| SNOMED CT | 93520007 | Hodgkin disease of intra-abdominal lymph nodes |
| SNOMED CT | 93521006 | Hodgkin disease of intrapelvic lymph nodes |
| SNOMED CT | 93522004 | Hodgkin disease of intrathoracic lymph nodes |
| SNOMED CT | 93523009 | Hodgkin disease of lymph nodes of axilla AND/OR upper limb |
| SNOMED CT | 93524003 | Hodgkin disease of lymph nodes of head, face AND/OR neck |
| SNOMED CT | 93525002 | Hodgkin disease of lymph nodes of inguinal region AND/OR lower limb |
| SNOMED CT | 93526001 | Hodgkin disease of lymph nodes of multiple sites |
| SNOMED CT | 93527005 | Hodgkin disease of spleen |
| SNOMED CT | 93528000 | Hodgkin disease of extranodal AND/OR solid organ site |
| SNOMED CT | 93530003 | Hodgkin granuloma of intrapelvic lymph nodes |
| SNOMED CT | 93531004 | Hodgkin granuloma of intrathoracic lymph nodes |
| SNOMED CT | 93532006 | Hodgkin granuloma of lymph nodes of axilla AND/OR upper limb |
| SNOMED CT | 93533001 | Hodgkin granuloma of lymph nodes of head, face AND/OR neck |
| SNOMED CT | 93534007 | Hodgkin granuloma of lymph nodes of inguinal region AND/OR lower limb |
| SNOMED CT | 93536009 | Hodgkin granuloma of spleen |
| SNOMED CT | 93537000 | Hodgkin granuloma of extranodal AND/OR solid organ site |
| SNOMED CT | 93541001 | Hodgkin paragranuloma of lymph nodes of axilla AND/OR upper limb |
| SNOMED CT | 93542008 | Hodgkin paragranuloma of lymph nodes of head, face AND/OR neck |
| SNOMED CT | 93543003 | Hodgkin paragranuloma of lymph nodes of inguinal region AND/OR lower limb |
| SNOMED CT | 93546006 | Hodgkin paragranuloma of extranodal AND/OR solid organ site |
| SNOMED CT | 93547002 | Hodgkin sarcoma of intra-abdominal lymph nodes |
| SNOMED CT | 93548007 | Hodgkin sarcoma of intrapelvic lymph nodes |
| SNOMED CT | 93549004 | Hodgkin sarcoma of intrathoracic lymph nodes |
| SNOMED CT | 93550004 | Hodgkin sarcoma of lymph nodes of axilla AND/OR upper limb |
| SNOMED CT | 93551000 | Hodgkin sarcoma of lymph nodes of head, face AND/OR neck |

Continued on next page

**Table 7 – continued from previous page**

| CodeSystem | ConceptCode | ConceptName |
|---|---|---|
| SNOMED CT | 93552007 | Hodgkin sarcoma of lymph nodes of inguinal region AND/OR lower limb |
| SNOMED CT | 93554008 | Hodgkin sarcoma of spleen |
| SNOMED CT | 93555009 | Hodgkin sarcoma of extranodal AND/OR solid organ site |
| SNOMED CT | 94719007 | Granulocytic sarcoma |
| SNOMED CT | 103686008 | Enteropathy associated T-cell lymphoma |
| SNOMED CT | 109391009 | Kaposi sarcoma of lymph nodes |
| SNOMED CT | 109962001 | Nonfollicular lymphoma |
| SNOMED CT | 109964000 | Diffuse non-Hodgkin lymphoma, undifferentiated |
| SNOMED CT | 109965004 | Lymphoblastoma |
| SNOMED CT | 109966003 | Malignant lymphoma - immunoblastic |
| SNOMED CT | 109967007 | Diffuse non-Hodgkin lymphoma, small cleaved cell |
| SNOMED CT | 109968002 | Diffuse non-Hodgkin lymphoma, small cell |
| SNOMED CT | 109969005 | DLBCL - diffuse large B cell lymphoma |
| SNOMED CT | 109970006 | Follicular lymphoma, grade 1 |
| SNOMED CT | 109971005 | Follicular lymphoma grade 2 |
| SNOMED CT | 109972003 | Follicular lymphoma grade 3 |
| SNOMED CT | 109977009 | Peripheral T-cell lymphoma pleomorphic small cell |
| SNOMED CT | 109979007 | B-cell lymphoma |
| SNOMED CT | 109983007 | Monoclonal paraproteinemia |
| SNOMED CT | 110004001 | Acute promyelocytic leukemia (clinical) |
| SNOMED CT | 110005000 | Acute myelomonocytic leukemia (clinical) |
| SNOMED CT | 110007008 | Adult T-cell leukemia |
| SNOMED CT | 111396008 | Hereditary gigantism of cytoplasmic organelles |
| SNOMED CT | 118599009 | Lymphoma, Hodgkins |
| SNOMED CT | 118600007 | Malignant lymphoma (clinical) |
| SNOMED CT | 118601006 | Non-Hodgkin lymphoma |
| SNOMED CT | 118602004 | Hodgkin granuloma |
| SNOMED CT | 118605002 | Hodgkin lymphoma, nodular lymphocyte predominance |
| SNOMED CT | 118606001 | Hodgkin sarcoma |
| SNOMED CT | 118608000 | Hodgkin disease, nodular sclerosis |
| SNOMED CT | 118609008 | Hodgkin mixed cellularity lymphoma |
| SNOMED CT | 118610003 | Hodgkin disease, lymphocytic depletion |
| SNOMED CT | 118613001 | Leukemic reticuloendotheliosis |
| SNOMED CT | 118614007 | Acute infancy reticulosis |
| SNOMED CT | 118617000 | Burkitt lymphoma |
| SNOMED CT | 128799007 | Classical Hodgkin lymphoma, lymphocyte-rich |
| SNOMED CT | 128805001 | T/NK-cell lymphoma, nasal and nasal type |
| SNOMED CT | 128809007 | Langerhans cell granulomatosis |
| SNOMED CT | 128812005 | Langerhans cell granulomatosis |
| SNOMED CT | 128819001 | T-cell large granular lymphocytic leukaemia |
| SNOMED CT | 128821006 | T-cell prolymphocytic leukemia |
| SNOMED CT | 128833001 | Aggressive natural killer-cell leukemia |

Continued on next page

**Table 7 – continued from previous page**

| CodeSystem | ConceptCode | ConceptName |
|---|---|---|
| SNOMED CT | 128875000 | Primary cutaneous CD30 antigen positive large T-cell lymphoma |
| SNOMED CT | 128929007 | Non-Hodgkin lymphoma - category |
| SNOMED CT | 128930002 | Hodgkin lymphoma - category |
| SNOMED CT | 188510001 | Burkitt lymphoma of lymph nodes of head, face and neck |
| SNOMED CT | 188511002 | Burkitt tumor of intrathoracic lymph nodes |
| SNOMED CT | 188512009 | Burkitt tumor of intra-abdominal lymph nodes |
| SNOMED CT | 188513004 | Burkitt lymphoma of lymph nodes of axilla and upper limb |
| SNOMED CT | 188514005 | Burkitt lymphoma of lymph nodes of inguinal region and lower limb |
| SNOMED CT | 188515006 | Burkitt tumor of intrapelvic lymph nodes |
| SNOMED CT | 188516007 | Burkitt tumor of spleen |
| SNOMED CT | 188517003 | Burkitt tumor of lymph nodes of multiple sites |
| SNOMED CT | 188524002 | Hodgkin paragranuloma of intrathoracic lymph nodes |
| SNOMED CT | 188529007 | Hodgkin paragranuloma of intrapelvic lymph nodes |
| SNOMED CT | 188531003 | Hodgkin paragranuloma of lymph nodes of multiple sites |
| SNOMED CT | 188534006 | Hodgkin granuloma of lymph nodes of head, face and neck |
| SNOMED CT | 188536008 | Hodgkin granuloma of intra-abdominal lymph nodes |
| SNOMED CT | 188537004 | Hodgkin granuloma of lymph nodes of axilla and upper limb |
| SNOMED CT | 188538009 | Hodgkin granuloma of lymph nodes of inguinal region and lower limb |
| SNOMED CT | 188541000 | Hodgkin granuloma of lymph nodes of multiple sites |
| SNOMED CT | 188544008 | Hodgkin sarcoma of lymph nodes of head, face and neck |
| SNOMED CT | 188547001 | Hodgkin sarcoma of lymph nodes of axilla and upper limb |
| SNOMED CT | 188548006 | Hodgkin sarcoma of lymph nodes of inguinal region and lower limb |
| SNOMED CT | 188551004 | Hodgkin sarcoma of lymph nodes of multiple sites |
| SNOMED CT | 188554007 | Hodgkin disease, lymphocytic-histiocytic predominance of lymph nodes of head, face and neck |
| SNOMED CT | 188558005 | Hodgkin disease, lymphocytic-histiocytic predominance of lymph nodes of axilla and upper limb |
| SNOMED CT | 188559002 | Hodgkin disease, lymphocytic-histiocytic predominance of lymph nodes of inguinal region and lower limb |
| SNOMED CT | 188562004 | Hodgkin disease, lymphocytic-histiocytic predominance of lymph nodes of multiple sites |
| SNOMED CT | 188565002 | Hodgkin disease, nodular sclerosis of lymph nodes of head, face and neck |
| SNOMED CT | 188566001 | Hodgkin disease, nodular sclerosis of intrathoracic lymph nodes |
| SNOMED CT | 188567005 | Hodgkin disease, nodular sclerosis of intra-abdominal lymph nodes |
| SNOMED CT | 188568000 | Hodgkin disease, nodular sclerosis of lymph nodes of axilla and upper limb |

Continued on next page

**Table 7 – continued from previous page**

| CodeSystem | ConceptCode | ConceptName |
|---|---|---|
| SNOMED CT | 188569008 | Hodgkin disease, nodular sclerosis of lymph nodes of inguinal region and lower limb |
| SNOMED CT | 188570009 | Hodgkin disease, nodular sclerosis of intrapelvic lymph nodes |
| SNOMED CT | 188572001 | Hodgkin disease, nodular sclerosis of lymph nodes of multiple sites |
| SNOMED CT | 188575004 | Hodgkin disease, mixed cellularity of lymph nodes of head, face and neck |
| SNOMED CT | 188576003 | Hodgkin disease, mixed cellularity of intrathoracic lymph nodes |
| SNOMED CT | 188577007 | Hodgkin disease, mixed cellularity of intra-abdominal lymph nodes |
| SNOMED CT | 188578002 | Hodgkin disease, mixed cellularity of lymph nodes of axilla and upper limb |
| SNOMED CT | 188579005 | Hodgkin disease, mixed cellularity of lymph nodes of inguinal region and lower limb |
| SNOMED CT | 188580008 | Hodgkin disease, mixed cellularity of intrapelvic lymph nodes |
| SNOMED CT | 188582000 | Hodgkin disease, mixed cellularity of lymph nodes of multiple sites |
| SNOMED CT | 188585003 | Hodgkin disease, lymphocytic depletion of lymph nodes of head, face and neck |
| SNOMED CT | 188586002 | Hodgkin disease, lymphocytic depletion of intrathoracic lymph nodes |
| SNOMED CT | 188587006 | Hodgkin disease, lymphocytic depletion of intra-abdominal lymph nodes |
| SNOMED CT | 188589009 | Hodgkin disease, lymphocytic depletion of lymph nodes of axilla and upper limb |
| SNOMED CT | 188590000 | Hodgkin disease, lymphocytic depletion of lymph nodes of inguinal region and lower limb |
| SNOMED CT | 188591001 | Hodgkin disease, lymphocytic depletion of intrapelvic lymph nodes |
| SNOMED CT | 188592008 | Hodgkin disease, lymphocytic depletion of spleen |
| SNOMED CT | 188593003 | Hodgkin disease, lymphocytic depletion of lymph nodes of multiple sites |
| SNOMED CT | 188632001 | Lymphoma intra abdominal, Sezarys |
| SNOMED CT | 188672005 | Follicular non-Hodgkin mixed small cleaved and large cell lymphoma |
| SNOMED CT | 188679001 | Diffuse non-Hodgkin lymphoma undifferentiated (diffuse) |
| SNOMED CT | 240531002 | African Burkitt lymphoma |
| SNOMED CT | 277550009 | Richter syndrome |
| SNOMED CT | 277609007 | Hodgkin disease, lymphocytic predominance - diffuse |

Continued on next page

**Table 7 – continued from previous page**

| CodeSystem | ConceptCode | ConceptName |
|---|---|---|
| SNOMED CT | 277610002 | Hodgkin disease, nodular sclerosis - lymphocytic predominance |
| SNOMED CT | 277611003 | Hodgkin disease, nodular sclerosis - mixed cellularity |
| SNOMED CT | 277612005 | Hodgkin disease, nodular sclerosis - lymphocytic depletion |
| SNOMED CT | 277654008 | Enteropathy-type T-cell lymphoma |
| SNOMED CT | 278257006 | Peripheral blood stem cell transplantation |
| SNOMED CT | 285420006 | Immunoglobulin A myeloma |
| SNOMED CT | 285421005 | Immunoglobulin G myeloma |
| SNOMED CT | 285422003 | Immunoglobulin D myeloma |
| SNOMED CT | 307633009 | Hodgkin disease, lymphocytic depletion, diffuse fibrosis |
| SNOMED CT | 307634003 | Hodgkin disease, lymphocytic depletion, reticular type |
| SNOMED CT | 307635002 | Hodgkin disease, nodular sclerosis - cellular phase |
| SNOMED CT | 308121000 | Follicular non-Hodgkin lymphoma |
| SNOMED CT | 315019000 | Human immunodeficiency virus infection with aseptic meningitis |
| SNOMED CT | 350353007 | Immunodeficiency with generalized hematopoietic hypoplasia |
| SNOMED CT | 359631009 | M0 - Acute myeloblastic leukemia - undifferentiated |
| SNOMED CT | 359640008 | Acute myelogenous leukemia without maturation |
| SNOMED CT | 359648001 | M2 - Acute myeloblastic leukemia with maturation |
| SNOMED CT | 362993009 | Autosomal recessive SCID (severe combined immunodeficiency disease) |
| SNOMED CT | 373381004 | Myelodysplastic syndrome, unclassifiable |
| SNOMED CT | 402881008 | Cutaneous B-cell lymphoma |
| SNOMED CT | 403835002 | X-linked hyper-immunoglobulin M syndrome |
| SNOMED CT | 404106004 | Lymphomatoid papulosis with Hodgkin disease |
| SNOMED CT | 404122003 | Leukaemic infiltration of skin (chronic T-cell lymphocytic leukaemia) |
| SNOMED CT | 404123008 | Leukaemic infiltration of skin (T-cell prolymphocytic leukaemia) |
| SNOMED CT | 404124002 | Leukaemic infiltration of skin (T-cell lymphoblastic leukaemia) |
| SNOMED CT | 404135007 | Angiocentric lymphoma involving skin |
| SNOMED CT | 404136008 | Aggressive natural killer-cell leukemia involving skin |
| SNOMED CT | 404143002 | Crosti's lymphoma |
| SNOMED CT | 404157000 | Specific skin infiltration in Hodgkin disease |
| SNOMED CT | 413527004 | Anaplastic large cell lymphoma, T- and null cell primary systemic types |
| SNOMED CT | 414166008 | Extranodal natural killer/T-cell lymphoma, nasal type |
| SNOMED CT | 414780005 | Mucosa-associated lymphoid tissue lymphoma of orbit |
| SNOMED CT | 416729007 | Neutropenia with AIDS (acquired immunodeficiency syndrome) |
| SNOMED CT | 419094004 | Immunodeficiency-associated Burkitt lymphoma |

Continued on next page

**Table 7 – continued from previous page**

| CodeSystem | ConceptCode | ConceptName |
|---|---|---|
| SNOMED CT | 419770008 | Endemic Burkitt lymphoma |
| SNOMED CT | 419879004 | Atypical Burkitt lymphoma |
| SNOMED CT | 420063007 | Sporadic Burkitt lymphoma |
| SNOMED CT | 420244003 | Encephalitis with AIDS (acquired immunodeficiency syndrome) |
| SNOMED CT | 420281004 | Skin rash with AIDS (acquired immunodeficiency syndrome) |
| SNOMED CT | 420302007 | Reticulosarcoma with AIDS (acquired immunodeficiency syndrome) |
| SNOMED CT | 420308006 | Retinal vascular changes with AIDS (acquired immunodeficiency syndrome) |
| SNOMED CT | 420321004 | Intestinal malabsorption with AIDS (acquired immunodeficiency syndrome) |
| SNOMED CT | 420403001 | Pneumocystosis with AIDS (acquired immunodeficiency syndrome) |
| SNOMED CT | 420452002 | Myelopathy with AIDS (acquired immunodeficiency syndrome) |
| SNOMED CT | 420543008 | Anemia associated with acquired immunodeficiency syndrome |
| SNOMED CT | 420544002 | Bacterial pneumonia with AIDS (acquired immunodeficiency syndrome) |
| SNOMED CT | 420549007 | Salivary gland disease with AIDS (acquired immunodeficiency syndrome) |
| SNOMED CT | 420554003 | Progressive multifocal leukoencephalopathy with AIDS (acquired immunodeficiency syndrome) |
| SNOMED CT | 420614009 | Organic dementia with AIDS (acquired immunodeficiency syndrome) |
| SNOMED CT | 420658009 | Radiculitis with AIDS (acquired immunodeficiency syndrome) |
| SNOMED CT | 420687005 | Ill-defined intestinal infection associated with acquired immunodeficiency syndrome |
| SNOMED CT | 420691000 | Nutritional deficiency with AIDS (acquired immunodeficiency syndrome) |
| SNOMED CT | 420718004 | Central nervous system demyelinating disease with AIDS (acquired immunodeficiency syndrome) |
| SNOMED CT | 420721002 | Acquired immunodeficiency syndrome-associated disorder |
| SNOMED CT | 420764009 | Salmonella infection with AIDS (acquired immunodeficiency syndrome) |
| SNOMED CT | 420774007 | Organic brain syndrome with AIDS (acquired immunodeficiency syndrome) |
| SNOMED CT | 420787001 | Pneumococcal pneumonia with AIDS (acquired immunodeficiency syndrome) |
| SNOMED CT | 420801006 | Malaise with AIDS (acquired immunodeficiency syndrome) |

Continued on next page

**Table 7 – continued from previous page**

| CodeSystem | ConceptCode | ConceptName |
|---|---|---|
| SNOMED CT | 420818005 | Mycobacteriosis with AIDS (acquired immunodeficiency syndrome) |
| SNOMED CT | 420877009 | Dermatomycosis with AIDS (acquired immunodeficiency syndrome) |
| SNOMED CT | 420900006 | Fatigue with AIDS (acquired immunodeficiency syndrome) |
| SNOMED CT | 420938005 | Subacute endocarditis with AIDS (acquired immunodeficiency syndrome) |
| SNOMED CT | 420945005 | Histoplasmosis with AIDS (acquired immunodeficiency syndrome) |
| SNOMED CT | 421023003 | Presenile dementia with AIDS (acquired immunodeficiency syndrome) |
| SNOMED CT | 421047005 | Candidiasis of lung with AIDS (acquired immunodeficiency syndrome) |
| SNOMED CT | 421102007 | Aplastic anemia with AIDS (acquired immunodeficiency syndrome) |
| SNOMED CT | 421230000 | Hepatomegaly with AIDS (acquired immunodeficiency syndrome) |
| SNOMED CT | 421272004 | Subacute myocarditis with AIDS (acquired immunodeficiency syndrome) |
| SNOMED CT | 421283008 | Primary lymphoma of brain with AIDS (acquired immunodeficiency syndrome) |
| SNOMED CT | 421312009 | Agranulocytosis with AIDS (acquired immunodeficiency syndrome) |
| SNOMED CT | 421315006 | Myelitis with AIDS (acquired immunodeficiency syndrome) |
| SNOMED CT | 421394009 | Skin disorder with AIDS (acquired immunodeficiency syndrome) |
| SNOMED CT | 421403008 | Cryptococcosis with AIDS (acquired immunodeficiency syndrome) |
| SNOMED CT | 421415007 | Subacute adenoviral encephalitis with AIDS (acquired immunodeficiency syndrome) |
| SNOMED CT | 421418009 | Mature T-cell AND/OR natural killer cell neoplasm |
| SNOMED CT | 421431004 | Nocardiosis with AIDS (acquired immunodeficiency syndrome) |
| SNOMED CT | 421454008 | Infectious gastroenteritis with AIDS (acquired immunodeficiency syndrome) |
| SNOMED CT | 421460008 | Retinopathy with AIDS (acquired immunodeficiency syndrome) |
| SNOMED CT | 421508002 | Viral pneumonia with AIDS (acquired immunodeficiency syndrome) |
| SNOMED CT | 421529006 | Dementia with AIDS (acquired immunodeficiency syndrome) |
| SNOMED CT | 421571007 | Tuberculosis with AIDS (acquired immunodeficiency syndrome) |

Continued on next page

**Table 7 – continued from previous page**

| CodeSystem | ConceptCode | ConceptName |
|---|---|---|
| SNOMED CT | 421597001 | Polyneuropathy with AIDS (acquired immunodeficiency syndrome) |
| SNOMED CT | 421660003 | Failure to thrive in infant with AIDS (acquired immunodeficiency syndrome) |
| SNOMED CT | 421671002 | Pneumonia with AIDS (acquired immunodeficiency syndrome) |
| SNOMED CT | 421695000 | Abnormal weight loss associated with acquired immunodeficiency syndrome |
| SNOMED CT | 421706001 | Blindness with AIDS (acquired immunodeficiency syndrome) |
| SNOMED CT | 421708000 | Infective arthritis with AIDS (acquired immunodeficiency syndrome) |
| SNOMED CT | 421710003 | Candidiasis of mouth with AIDS (acquired immunodeficiency syndrome) |
| SNOMED CT | 421766003 | Thrombocytopenia with AIDS (acquired immunodeficiency syndrome) |
| SNOMED CT | 421827003 | Encephalopathy with AIDS (acquired immunodeficiency syndrome) |
| SNOMED CT | 421851008 | Acquired hemolytic anemia associated with acquired immunodeficiency syndrome |
| SNOMED CT | 421874007 | Respiratory disorder with AIDS (acquired immunodeficiency syndrome) |
| SNOMED CT | 421883002 | Strongyloidiasis with AIDS (acquired immunodeficiency syndrome) |
| SNOMED CT | 421929001 | Myocarditis with AIDS (acquired immunodeficiency syndrome) |
| SNOMED CT | 421983003 | Noninfectious gastroenteritis with AIDS (acquired immunodeficiency syndrome) |
| SNOMED CT | 421998001 | Central nervous disorder with AIDS (acquired immunodeficiency syndrome) |
| SNOMED CT | 422003001 | Slim disease |
| SNOMED CT | 422012004 | Neuritis with AIDS (acquired immunodeficiency syndrome) |
| SNOMED CT | 422089004 | Encephalomyelitis with AIDS (acquired immunodeficiency syndrome) |
| SNOMED CT | 422136003 | Neuralgia with AIDS (acquired immunodeficiency syndrome) |
| SNOMED CT | 422177004 | Dyspnea with AIDS (acquired immunodeficiency syndrome) |
| SNOMED CT | 422189002 | Low vision with AIDS (acquired immunodeficiency syndrome) |
| SNOMED CT | 422194002 | Hyperhidrosis with AIDS (acquired immunodeficiency syndrome) |
| SNOMED CT | 422282000 | Malignant neoplasm with AIDS (acquired immunodeficiency syndrome) |

Continued on next page

**Table 7 – continued from previous page**

| CodeSystem | ConceptCode | ConceptName |
|---|---|---|
| SNOMED CT | 422337001 | Coccidioidomycosis with AIDS (acquired immunodeficiency syndrome) |
| SNOMED CT | 423294001 | Idiopathic hypereosinophilic syndrome (HES) |
| SNOMED CT | 445269007 | Extranodal marginal zone B-cell lymphoma of mucosa-associated lymphoid tissue |
| SNOMED CT | 450949002 | B lymphoblastic leukemia lymphoma, no International Classification of Diseases for Oncology subtype |
| SNOMED CT | 720345008 | T-cell immunodeficiency, congenital alopecia, and nail dystrophy |
| SNOMED CT | 733627006 | Primary cutaneous gamma-delta T-cell lymphoma |
| SNOMED CT | 781206002 | Rheumatoid spondylitis |
| SNOMED CT | 783787000 | Retinal vasculopathy with cerebral leucoencephalopathy and systemic manifestations |
| SNOMED CT | 1003381002 | Onycho-tricho-dysplasia neutropenia syndrome |
| SNOMED CT | 1003473002 | B cell lymphocyte aplasia caused by drug |
| SNOMED CT | 1003679009 | Human immunodeficiency virus Centers for Disease Control and Prevention dual infection with HIV-1 and HIV-2 |
| SNOMED CT | 1003680007 | Human immunodeficiency virus Centers for Disease Control and Prevention undifferentiated type |
| SNOMED CT | 1003682004 | Human immunodeficiency virus Centers for Disease Control and Prevention stage 1 |
| SNOMED CT | 1003683009 | Human immunodeficiency virus Centers for Disease Control and Prevention stage 2 |
| SNOMED CT | 1003684003 | Human immunodeficiency virus Centers for Disease Control and Prevention stage 3 |
| SNOMED CT | 1004006009 | World Health Organization human immunodeficiency virus infection clinical stage |
| SNOMED CT | 143291000119104 | Non-Hodgkin's lymphoma in remission |
| SNOMED CT | 318861000119101 | Seropositive rheumatoid arthritis of left hand |
| SNOMED CT | 318931000119100 | Seropositive rheumatoid arthritis of right hand |
| SNOMED CT | 878857009 | Malignant lymphomatoid granulomatosis grade 3 |
| SNOMED CT | 878858004 | Malignant lymphomatoid granulomatosis grade 3 of lung |
| SNOMED CT | 898197001 | Adult T-cell leukemia/lymphoma in remission |
| SNOMED CT | 898198006 | T-cell prolymphocytic leukemia in remission |
| SNOMED CT | 1229988003 | Myeloid and/or lymphoid neoplasm with PCM1-JAK2 |
| SNOMED CT | 1204344007 | Myelodysplastic syndrome with single lineage dysplasia |
| SNOMED CT | 16415791000119104 | Myasthenia gravis in remission |
| SNOMED CT | 1204460007 | Myocardial dysfunction with sepsis |
| SNOMED CT | 348741000119101 | Corneal infiltrate of left eye following penetrating keratoplasty |
| SNOMED CT | 348291000119100 | Corneal infiltrate of right eye following penetrating keratoplasty |
| SNOMED CT | 1217301006 | Precursor cell lymphoblastic lymphoma |

Continued on next page

**Table 7 – continued from previous page**

| CodeSystem | ConceptCode | ConceptName |
|---|---|---|
| SNOMED CT | 756003 | Chronic rheumatic myopericarditis |
| SNOMED CT | 961007 | Erythema nodosum, acute form |
| SNOMED CT | 1125006 | Sepsis during labor |
| SNOMED CT | 1824008 | Allergic gastritis |
| SNOMED CT | 2557004 | Chronic leukemia |
| SNOMED CT | 2772003 | Acquired epidermolysis bullosa |
| SNOMED CT | 2858002 | Puerperal sepsis |
| SNOMED CT | 2897005 | Immune thrombocytopenia |
| SNOMED CT | 2990008 | Lymphocytic leukaemoid reaction |
| SNOMED CT | 3533007 | Pustular bacterid |
| SNOMED CT | 3978000 | Warm autoimmune hemolytic anemia |
| SNOMED CT | 4089001 | Meningococcemia |
| SNOMED CT | 4448006 | Allergic headache |
| SNOMED CT | 5085001 | Gonococcemia |
| SNOMED CT | 1208892009 | Alopecia areata of eyebrow |
| SNOMED CT | 5567000 | Gas gangrene septicemia |
| SNOMED CT | 5860009 | Ophiasis |
| SNOMED CT | 7021009 | Gastrointestinal eosinophilic granuloma |
| SNOMED CT | 7119001 | Cutaneous lupus erythematosus |
| SNOMED CT | 7548000 | Rheumatic pneumonia |
| SNOMED CT | 8090002 | Eosinophilic granuloma of oral mucosa |
| SNOMED CT | 8214000 | Telangiectasia macularis eruptiva perstans |
| SNOMED CT | 8326008 | Acute allergic mucoid otitis media |
| SNOMED CT | 8554002 | Brazilian purpuric fever |
| SNOMED CT | 9012003 | Septicemic plague |
| SNOMED CT | 9014002 | Psoriasis |
| SNOMED CT | 9768003 | Tuberculosis cutis lichenoides |
| SNOMED CT | 10001005 | Bacterial sepsis |
| SNOMED CT | 10294000 | Reactive mastocytosis |
| SNOMED CT | 10713006 | Diffuse interstitial rheumatoid disease of lung |
| SNOMED CT | 10785007 | Vinyard sprayers' lung |
| SNOMED CT | 10803007 | Jarisch Herxheimer reaction |
| SNOMED CT | 10934005 | Cryofibrinogenemia |
| SNOMED CT | 11244009 | Polyglandular autoimmune syndrome, type 1 |
| SNOMED CT | 11352009 | Primary cryoglobulinemia |
| SNOMED CT | 11354005 | Tuberculosis cutis indurativa |
| SNOMED CT | 11373009 | Illegal termination of pregnancy with sepsis |
| SNOMED CT | 11781007 | Autoimmune hemolytic anemia due to complement |
| SNOMED CT | 11944003 | Feather-pickers' disease |
| SNOMED CT | 12088005 | Budgerigar-fanciers' disease |
| SNOMED CT | 12219003 | Aleukemic lymphoid leukemia |
| SNOMED CT | 12263007 | Type 1 hypersensitivity response |
| SNOMED CT | 12761008 | Acute photoallergic dermatitis |

Continued on next page

**Table 7 – continued from previous page**

| CodeSystem | ConceptCode | ConceptName |
|---|---|---|
| SNOMED CT | 12834001 | Subacute monocytic leukemia |
| SNOMED CT | 13160009 | Complication of transplanted intestines |
| SNOMED CT | 13172003 | Chronic idiopathic thrombocytopenic purpura |
| SNOMED CT | 13394002 | Suberosis |
| SNOMED CT | 13470001 | Chronic ulcerative ileocolitis |
| SNOMED CT | 14311001 | Severe chronic ulcerative colitis |
| SNOMED CT | 14333004 | Alloimmune neonatal neutropenia |
| SNOMED CT | 14589007 | Phacoanaphylaxis |
| SNOMED CT | 14654002 | Localized anaphylaxis |
| SNOMED CT | 15084002 | Lupus erythematosus profundus |
| SNOMED CT | 15342002 | Chronic ulcerative enterocolitis |
| SNOMED CT | 15470004 | Toxic diffuse goiter with acropachy |
| SNOMED CT | 15676002 | Acute rheumatic pericarditis |
| SNOMED CT | 16623004 | Coffee-workers' lung |
| SNOMED CT | 16751003 | Acute tubulointerstitial nephritis associated with systemic infection |
| SNOMED CT | 16935003 | Allergic arthritis |
| SNOMED CT | 17866004 | Acute allergic sanguinous otitis media |
| SNOMED CT | 18192007 | Acute rheumatic endocarditis |
| SNOMED CT | 18323000 | Drug-induced immune hemolytic anemia, immune complex type |
| SNOMED CT | 18690003 | Farmers' lung |
| SNOMED CT | 19076009 | Sick building syndrome |
| SNOMED CT | 19087001 | Tinea kerion |
| SNOMED CT | 19267009 | Lupus anticoagulant disorder |
| SNOMED CT | 19274004 | Grain-handlers' disease |
| SNOMED CT | 19307009 | Drug-induced immune thrombocytopenia |
| SNOMED CT | 19514005 | Arthritis mutilans |
| SNOMED CT | 19636003 | Monocytosis |
| SNOMED CT | 20005002 | Recurrent rheumatic heart disease |
| SNOMED CT | 21983002 | Hashimoto thyroiditis |
| SNOMED CT | 22231002 | Allergic enteritis of intestine |
| SNOMED CT | 22310002 | Chronic idiopathic monocytosis |
| SNOMED CT | 22695001 | Aleukemic monocytic leukemia |
| SNOMED CT | 22775002 | Allergic vulvitis |
| SNOMED CT | 22888007 | Drug-induced lupus erythematosus due to procainamide |
| SNOMED CT | 22972008 | Erythema multiforme, dermal type |
| SNOMED CT | 23315001 | Sequoiosis |
| SNOMED CT | 24159007 | Persistent lymphocytosis |
| SNOMED CT | 24363009 | Rheumatic fever without heart involvement |
| SNOMED CT | 24444009 | Failed attempted termination of pregnancy with sepsis |
| SNOMED CT | 24526004 | Inflammatory bowel disease |
| SNOMED CT | 24700007 | Multiple sclerosis |

Continued on next page

**Table 7 – continued from previous page**

| CodeSystem | ConceptCode | ConceptName |
|---|---|---|
| SNOMED CT | 24829000 | Eosinophilic ulcerative colitis |
| SNOMED CT | 25042006 | Tracheostomy sepsis |
| SNOMED CT | 25044007 | Neuromyelitis optica |
| SNOMED CT | 25847004 | Seborrheic psoriasis |
| SNOMED CT | 25887009 | Allergic sigmoiditis |
| SNOMED CT | 25897000 | Malt-workers' lung |
| SNOMED CT | 26006005 | Allergic gastroenteritis |
| SNOMED CT | 26045000 | Chronic allergic conjunctivitis |
| SNOMED CT | 26151008 | Graves' disease with acropachy AND with thyrotoxic crisis |
| SNOMED CT | 26169004 | Allergic otitis media |
| SNOMED CT | 26843008 | Antiphospholipid syndrome |
| SNOMED CT | 27040004 | Experimental allergic encephalomyelitis |
| SNOMED CT | 28122003 | Pulmonary eosinophilic granuloma |
| SNOMED CT | 28505005 | Acute idiopathic thrombocytopenic purpura |
| SNOMED CT | 28664002 | Herpes iris |
| SNOMED CT | 28807005 | Secondary cryoglobulinemia |
| SNOMED CT | 28840001 | Generalized pustular psoriasis, exanthematous type |
| SNOMED CT | 29120000 | Eosinophilic colitis |
| SNOMED CT | 29789008 | Monocytic leukemoid reaction |
| SNOMED CT | 30304000 | Allergic colitis |
| SNOMED CT | 30911005 | Cryoglobulinemia |
| SNOMED CT | 30981000 | Secondary eosinophilia |
| SNOMED CT | 31712002 | Primary biliary cholangitis |
| SNOMED CT | 32273002 | Idiopathic thrombocytopenic purpura |
| SNOMED CT | 32295003 | Allergic ileitis |
| SNOMED CT | 32861005 | Erythema nodosum |
| SNOMED CT | 33167004 | Complication of transplanted liver |
| SNOMED CT | 33339001 | Psoriasis with arthropathy |
| SNOMED CT | 33560006 | Subacute lymphoid leukemia |
| SNOMED CT | 33719002 | Subcutaneous rheumatoid nodule |
| SNOMED CT | 34015007 | Bakers' asthma |
| SNOMED CT | 34250006 | Benign mucous membrane pemphigoid |
| SNOMED CT | 34739009 | Urticaria pigmentosa, adult form |
| SNOMED CT | 35154004 | Pemphigus foliaceus |
| SNOMED CT | 35703006 | Post-viral paroxysmal cold hemoglobinuria |
| SNOMED CT | 35750002 | Aleukemic myeloid leukemia |
| SNOMED CT | 36715001 | Erythema multiforme |
| SNOMED CT | 36739006 | Pemphigus erythematosus |
| SNOMED CT | 37042000 | Guttate psoriasis |
| SNOMED CT | 37471005 | Extrinsic allergic alveolitis |
| SNOMED CT | 37981002 | Allergic bronchopulmonary aspergillosis |
| SNOMED CT | 38064006 | Reactive monocytosis |
| SNOMED CT | 38675009 | Monoclonal cryoimmunoglobulinemia |

Continued on next page

**Table 7 – continued from previous page**

| CodeSystem | ConceptCode | ConceptName |
|---|---|---|
| SNOMED CT | 38729007 | Wheat weevil disease |
| SNOMED CT | 38877003 | Rheumatoid aortitis |
| SNOMED CT | 38933006 | Recurrent rheumatic fever |
| SNOMED CT | 39246002 | Legal termination of pregnancy with sepsis |
| SNOMED CT | 39579001 | Anaphylaxis |
| SNOMED CT | 40178009 | Allergic urticaria |
| SNOMED CT | 40712008 | Allergic contact dermatitis caused by Arnica |
| SNOMED CT | 41030002 | Coccidioidomycosis with erythema multiforme |
| SNOMED CT | 41156006 | Tuberculosis papulonecrotica |
| SNOMED CT | 41816006 | Secondary cryofibrinogenemia |
| SNOMED CT | 41864002 | Autoimmune polyendocrinopathy |
| SNOMED CT | 42852008 | Infectious lymphocytosis |
| SNOMED CT | 42939002 | Pyemia |
| SNOMED CT | 43355006 | Eosinopenia |
| SNOMED CT | 44165003 | Complication of transplanted lung |
| SNOMED CT | 44206008 | Hapten type low affinity hemolytic anemia |
| SNOMED CT | 45309006 | Allergic contact dermatitis due to secretions |
| SNOMED CT | 46165000 | Chronic rheumatic endocarditis |
| SNOMED CT | 46359005 | Neutropenia associated with infectious disease |
| SNOMED CT | 46459009 | Brazilian pemphigus foliaceus |
| SNOMED CT | 46586006 | Diffuse alopecia areata |
| SNOMED CT | 46826000 | Rheumatic chorea |
| SNOMED CT | 46848006 | Sepsis following molar AND/OR ectopic pregnancy |
| SNOMED CT | 47144000 | Acute neutrophilia |
| SNOMED CT | 47986005 | Genetic anomaly of leukocyte |
| SNOMED CT | 48269007 | Allergic parotitis |
| SNOMED CT | 48516007 | Acute infectious lymphocytosis |
| SNOMED CT | 48813009 | Lymphocytopenia |
| SNOMED CT | 49237006 | Allergic diarrhea |
| SNOMED CT | 49420001 | Pemphigus vulgaris |
| SNOMED CT | 50253007 | Secondary paroxysmal cold hemoglobinuria |
| SNOMED CT | 50776006 | Allergic encephalomyelitis |
| SNOMED CT | 51264003 | Limbal AND/OR corneal involvement in vernal conjunctivitis |
| SNOMED CT | 51831006 | Sepsis following infusion, injection, transfusion AND/OR vaccination |
| SNOMED CT | 52168008 | Monocytic leukemia |
| SNOMED CT | 52333004 | Mushroom workers' lung |
| SNOMED CT | 52506002 | Chronic ulcerative rectosigmoiditis |
| SNOMED CT | 54867000 | Rheumatoid fibrosing alveolitis |
| SNOMED CT | 54912002 | Drug-induced lupus erythematosus due to hydralazine |
| SNOMED CT | 55444004 | Transient neonatal neutropenia |
| SNOMED CT | 55807009 | Exophthalmos due to toxic diffuse goiter |

Continued on next page

**Table 7 – continued from previous page**

| CodeSystem | ConceptCode | ConceptName |
|---|---|---|
| SNOMED CT | 55985003 | Atopic reaction |
| SNOMED CT | 56478004 | Leukemoid reaction |
| SNOMED CT | 56968009 | Asthma caused by wood dust |
| SNOMED CT | 57803009 | Rheumatic chorea with heart involvement |
| SNOMED CT | 58718002 | Rheumatic fever |
| SNOMED CT | 58797008 | Disorder of transplanted kidney |
| SNOMED CT | 59165007 | Rheumatoid scleritis |
| SNOMED CT | 59275002 | Acute allergic serous otitis media |
| SNOMED CT | 59940009 | Hypersensitivity alveolitis in lungworm infection |
| SNOMED CT | 59957008 | Neonatal Graves' disease |
| SNOMED CT | 60216004 | Toxic diffuse goiter with thyrotoxic crisis |
| SNOMED CT | 60268006 | Toxic diffuse goiter with pretibial myxedema |
| SNOMED CT | 60967006 | Allergic vaginitis |
| SNOMED CT | 61012002 | Chronic rheumatic myocarditis |
| SNOMED CT | 61458006 | Drug-induced lupus erythematosus due to diphenylhydantoin |
| SNOMED CT | 61582004 | Allergic rhinitis |
| SNOMED CT | 61802005 | Primary cryofibrinogenemia |
| SNOMED CT | 62853008 | Anti-glomerular basement membrane tubulointerstitial nephritis |
| SNOMED CT | 62871001 | Idiopathic paroxysmal cold hemoglobinuria |
| SNOMED CT | 63088003 | Allergic asthma without status asthmaticus |
| SNOMED CT | 63175003 | Localized extracutaneous mastocytosis |
| SNOMED CT | 63484008 | Drug-induced neutrophilia |
| SNOMED CT | 64072006 | Acute rheumatic pancarditis |
| SNOMED CT | 64249002 | Allergic eosinophilia |
| SNOMED CT | 64766004 | Ulcerative colitis |
| SNOMED CT | 64936001 | Simple pulmonary eosinophilia |
| SNOMED CT | 65172003 | Pemphigus |
| SNOMED CT | 65394002 | Allergic contact dermatitis caused by urushiol from Chinese lacquer tree |
| SNOMED CT | 65539006 | Impetigo herpetiformis |
| SNOMED CT | 66291003 | Tuberculous phlyctenular keratoconjunctivitis |
| SNOMED CT | 66628005 | Toxic diffuse goiter with exophthalmos AND with thyrotoxic storm |
| SNOMED CT | 66944004 | Autoimmune thyroiditis |
| SNOMED CT | 67023009 | Lymphocytosis |
| SNOMED CT | 67242002 | Bagassosis |
| SNOMED CT | 67465009 | Miscarriage with sepsis |
| SNOMED CT | 67678004 | Acute atopic conjunctivitis |
| SNOMED CT | 67824002 | Lymphosarcoma |
| SNOMED CT | 67895005 | Phlyctenular keratoconjunctivitis |
| SNOMED CT | 68225006 | Alopecia areata |

Continued on next page

**Table 7 – continued from previous page**

| CodeSystem | ConceptCode | ConceptName |
|---|---|---|
| SNOMED CT | 68333005 | Furriers' lung |
| SNOMED CT | 69688007 | Chronic erythema nodosum |
| SNOMED CT | 70139002 | Harara |
| SNOMED CT | 70349007 | Pseudoneutrophilia |
| SNOMED CT | 70884002 | Allergic encephalitis |
| SNOMED CT | 70910003 | Indolent systemic mastocytosis |
| SNOMED CT | 71610005 | Neutrophilic leukaemoid reaction |
| SNOMED CT | 71692003 | Leukoerythroblastotic reaction |
| SNOMED CT | 71958004 | Chronic allergic otitis media |
| SNOMED CT | 72272002 | Non malignant mast cell disease |
| SNOMED CT | 72275000 | Relapsing polychondritis |
| SNOMED CT | 72681001 | Allergic jejunitis |
| SNOMED CT | 72986009 | Acute hemorrhagic leukoencephalitis |
| SNOMED CT | 73397007 | Heparin-induced thrombocytopenia |
| SNOMED CT | 73448002 | Fish-meal workers' lung |
| SNOMED CT | 73658009 | Erythema multiforme, epidermal type |
| SNOMED CT | 74326002 | Subacute myeloid leukemia |
| SNOMED CT | 74610006 | Tuberculous erythema nodosum |
| SNOMED CT | 75316000 | Autoimmune hypoparathyroidism |
| SNOMED CT | 75331009 | Evans syndrome |
| SNOMED CT | 76092003 | Benign mucous membrane pemphigoid with ocular involvement |
| SNOMED CT | 76097009 | Erythema nodosum migrans |
| SNOMED CT | 76481008 | Lymphoepithelioid lymphoma |
| SNOMED CT | 76621000 | Urticaria pigmentosa, infantile form |
| SNOMED CT | 76762001 | Eosinophilic myopathy |
| SNOMED CT | 77090002 | Bullous pemphigoid |
| SNOMED CT | 77206006 | Puerperal pelvic sepsis |
| SNOMED CT | 77363004 | Concentric alopecia areata |
| SNOMED CT | 77522006 | Rheumatoid episcleritis |
| SNOMED CT | 78069008 | Chronic rheumatic pericarditis |
| SNOMED CT | 78079005 | Allergic contact dermatitis due to excretions |
| SNOMED CT | 78223000 | Coccidioidomycosis with erythema nodosum |
| SNOMED CT | 78324009 | Mild chronic ulcerative colitis |
| SNOMED CT | 78712000 | Moderate chronic ulcerative colitis |
| SNOMED CT | 78745000 | Urticaria pigmentosa |
| SNOMED CT | 79291003 | Discoid lupus erythematosus of eyelid |
| SNOMED CT | 79336007 | Familial eosinophilia |
| SNOMED CT | 79958002 | Grain fever |
| SNOMED CT | 80255009 | Maternal transfer neutropenia |
| SNOMED CT | 80258006 | Drug-induced lupus erythematosus |
| SNOMED CT | 80369006 | Chronic neutrophilia |
| SNOMED CT | 80988005 | Mixed cryofibrinogenemia |

Continued on next page

**Table 7 – continued from previous page**

| CodeSystem | ConceptCode | ConceptName |
|---|---|---|
| SNOMED CT | 81077008 | Acute rheumatic arthritis |
| SNOMED CT | 81271001 | Localized pustular psoriasis |
| SNOMED CT | 81285006 | Pemphigus vegetans |
| SNOMED CT | 82275008 | Eosinophilic granulomatosis with polyangiitis |
| SNOMED CT | 82939003 | Chronic rheumatoid nodular fibrosis |
| SNOMED CT | 83728000 | Polyglandular autoimmune syndrome, type 2 |
| SNOMED CT | 83839005 | Acrodermatitis continua of Hallopeau |
| SNOMED CT | 83942000 | Acute disseminated encephalomyelitis |
| SNOMED CT | 84479000 | Allergic inhalant dermatitis |
| SNOMED CT | 84730007 | Cryopathy |
| SNOMED CT | 84828003 | Leukopenia |
| SNOMED CT | 84871007 | Acute rheumatic valvulitis |
| SNOMED CT | 85512009 | Contact dermatitis due to jewelry |
| SNOMED CT | 85828009 | Autoimmune disease |
| SNOMED CT | 86081009 | Herpes gestationis |
| SNOMED CT | 86142006 | Pemphigoid |
| SNOMED CT | 86166000 | Alopecia universalis |
| SNOMED CT | 86225009 | Hapten type high affinity hemolytic anemia |
| SNOMED CT | 86563000 | Erythema multiforme, mixed dermal-epidermal type |
| SNOMED CT | 86638007 | Maple-bark strippers' lung |
| SNOMED CT | 88714009 | Transient hypogammaglobulinemia of infancy |
| SNOMED CT | 89099002 | Chronic allergic bronchitis |
| SNOMED CT | 89155008 | Systemic sclerosis |
| SNOMED CT | 89456004 | Chronic rheumatic valvulitis |
| SNOMED CT | 90453003 | Chronic rheumatic mediastinopericarditis |
| SNOMED CT | 91302008 | Sepsis |
| SNOMED CT | 91623008 | Hydroa herpetiformis |
| SNOMED CT | 91924004 | Allergic gingivitis |
| SNOMED CT | 91925003 | Allergic rhinitis due to animals |
| SNOMED CT | 91926002 | Allergic rhinitis due to grass pollen |
| SNOMED CT | 91927006 | Allergic rhinitis due to tree pollen |
| SNOMED CT | 91928001 | Allergic rhinitis due to weed pollen |
| SNOMED CT | 91941002 | Food anaphylaxis |
| SNOMED CT | 92816008 | Chronic monocytic leukemia |
| SNOMED CT | 92831004 | Complication of transplanted embryo |
| SNOMED CT | 95261008 | Sézary's disease of lymph nodes of inguinal region AND/OR lower limb |
| SNOMED CT | 95329006 | Autoimmune skin disease |
| SNOMED CT | 95332009 | Rash of systemic lupus erythematosus |
| SNOMED CT | 95416007 | Eosinophilia myalgia syndrome |
| SNOMED CT | 95609003 | Neonatal lupus erythematosus |
| SNOMED CT | 95624007 | Neonatal lymphocytopenia |
| SNOMED CT | 95643007 | Autoimmune encephalitis |

Continued on next page

**Table 7 – continued from previous page**

| CodeSystem | ConceptCode | ConceptName |
|---|---|---|
| SNOMED CT | 95733001 | Eosinophilic keratitis |
| SNOMED CT | 95742008 | Corneal graft rejection |
| SNOMED CT | 105601003 | Quantitative disorder of neutrophils |
| SNOMED CT | 105602005 | Quantitative abnormality of granulocytes |
| SNOMED CT | 109250009 | Chronic bullous dermatosis of childhood |
| SNOMED CT | 109992005 | Polycythemia vera (clinical) |
| SNOMED CT | 111037009 | Basophilic hyperplasia |
| SNOMED CT | 111196000 | Dermatitis herpetiformis |
| SNOMED CT | 111209006 | Photoallergic dermatitis |
| SNOMED CT | 111583006 | Leukocytosis |
| SNOMED CT | 111588002 | Heparin-induced thrombocytopenia with thrombosis |
| SNOMED CT | 115666004 | Animal handlers' lung |
| SNOMED CT | 123950001 | Rheumatic chorea without heart involvement |
| SNOMED CT | 127049002 | Primary (idiopathic) autoimmune hemolytic anemia |
| SNOMED CT | 127050002 | Secondary autoimmune hemolytic anemia |
| SNOMED CT | 127052005 | Secondary warm autoimmune hemolytic anemia |
| SNOMED CT | 127053000 | Post-infectious cold agglutinin disease |
| SNOMED CT | 127055007 | Chronic cold agglutinin disease |
| SNOMED CT | 127056008 | Chronic cold agglutinin disease associated with B-cell neoplasm |
| SNOMED CT | 127057004 | Paroxysmal cold hemoglobinuria |
| SNOMED CT | 127058009 | Paroxysmal cold hemoglobinuria associated with tertiary syphilis |
| SNOMED CT | 127060006 | Drug-induced immune hemolytic anemia, hapten type |
| SNOMED CT | 127061005 | Autoimmune hemolytic anemia, categorized by antibody class AND/OR complement |
| SNOMED CT | 127066000 | Familial polycythemia vera |
| SNOMED CT | 127067009 | Stress neutrophilia |
| SNOMED CT | 127218004 | Reactive follicular hyperplasia in the elderly |
| SNOMED CT | 128091003 | Primary ITP (immune thrombocytopenia) |
| SNOMED CT | 128092005 | Secondary autoimmune thrombocytopenia |
| SNOMED CT | 128093000 | Alloimmune thrombocytopenia |
| SNOMED CT | 128094006 | Alloimmune platelet transfusion refractoriness |
| SNOMED CT | 128460000 | Systemic sclerosis, diffuse |
| SNOMED CT | 128461001 | Systemic sclerosis sine scleroderma |
| SNOMED CT | 128600008 | Acute ulcerative colitis |
| SNOMED CT | 128831004 | Chronic myelomonocytic leukemia |
| SNOMED CT | 129563009 | Rheumatoid osteoperiostitis |
| SNOMED CT | 129639005 | Hereditary neutrophilia |
| SNOMED CT | 129640007 | Benign granulocytopenia in childhood |
| SNOMED CT | 129641006 | Chronic benign neutropenia of childhood |
| SNOMED CT | 156370009 | Psoriatic arthritis |
| SNOMED CT | 165531008 | Lymphocytosis - absolute |

Continued on next page

**Table 7 – continued from previous page**

| CodeSystem | ConceptCode | ConceptName |
|---|---|---|
| SNOMED CT | 186365005 | Acute meningococcemia |
| SNOMED CT | 190454008 | Autoimmune parathyroiditis |
| SNOMED CT | 191210005 | Primary cold-type hemolytic anemia |
| SNOMED CT | 191211009 | Primary warm-type hemolytic anemia |
| SNOMED CT | 191212002 | Secondary cold-type hemolytic anemia |
| SNOMED CT | 191213007 | Secondary warm-type hemolytic anemia |
| SNOMED CT | 191273000 | Leukemoid reaction of the newborn |
| SNOMED CT | 191306005 | Immunoglobulin A vasculitis |
| SNOMED CT | 191345000 | Acquired neutropenia in newborn |
| SNOMED CT | 191358004 | Hereditary eosinophilia |
| SNOMED CT | 191360002 | Drug-induced eosinophilia |
| SNOMED CT | 192926004 | Multiple sclerosis of the brainstem |
| SNOMED CT | 192927008 | Multiple sclerosis of the spinal cord |
| SNOMED CT | 192929006 | Exacerbation of multiple sclerosis |
| SNOMED CT | 194394004 | Gram positive sepsis |
| SNOMED CT | 194709000 | Acute rheumatic myocarditis |
| SNOMED CT | 195528001 | Acute rheumatic fever |
| SNOMED CT | 195663004 | Allergic pharyngitis |
| SNOMED CT | 195989002 | Pituitary snuff-takers' disease |
| SNOMED CT | 196133001 | Lung disease with systemic sclerosis |
| SNOMED CT | 196578009 | Crohn's disease of oral soft tissues |
| SNOMED CT | 196853004 | Septicemic pasteurellosis |
| SNOMED CT | 196987008 | Ulcerative ileocolitis |
| SNOMED CT | 197024006 | Allergic enterocolitis |
| SNOMED CT | 199127006 | Herpes gestationis - delivered |
| SNOMED CT | 199128001 | Herpes gestationis - delivered with postnatal complication |
| SNOMED CT | 199129009 | Herpes gestationis - not delivered |
| SNOMED CT | 199710008 | Sepsis during labor, delivered |
| SNOMED CT | 199711007 | Sepsis during labor with antenatal problem |
| SNOMED CT | 200196001 | Puerperal sepsis with postnatal complication |
| SNOMED CT | 200842003 | Allergic contact dermatitis due to adhesive |
| SNOMED CT | 200843008 | Allergic contact dermatitis due to cosmetic |
| SNOMED CT | 200844002 | Allergic contact dermatitis due to drug in contact with skin |
| SNOMED CT | 200845001 | Allergic contact dermatitis due to dye |
| SNOMED CT | 200847009 | Allergic contact dermatitis due to food in contact with the skin |
| SNOMED CT | 200907003 | Drug-induced pemphigus |
| SNOMED CT | 200936003 | Lupus erythematosus |
| SNOMED CT | 200937007 | Lupus erythematosus chronicus |
| SNOMED CT | 200938002 | Discoid lupus erythematosus |
| SNOMED CT | 200939005 | Lupus erythematosus migrans |
| SNOMED CT | 200940007 | Lupus erythematosus nodularis |
| SNOMED CT | 200941006 | Lupus erythematosus tumidus |

Continued on next page

**Table 7 – continued from previous page**

| CodeSystem | ConceptCode | ConceptName |
|---|---|---|
| SNOMED CT | 200942004 | Lupus erythematosus unguium mutilans |
| SNOMED CT | 200951007 | Erythema marginatum in acute rheumatic fever |
| SNOMED CT | 200956002 | Psoriatic arthritis with spine involvement |
| SNOMED CT | 200962007 | Psoriasis annularis |
| SNOMED CT | 200963002 | Psoriasis circinata |
| SNOMED CT | 200964008 | Psoriasis diffusa |
| SNOMED CT | 200965009 | Plaque psoriasis |
| SNOMED CT | 200966005 | Psoriasis geographica |
| SNOMED CT | 200967001 | Psoriasis gyrata |
| SNOMED CT | 200968006 | Psoriasis inveterata |
| SNOMED CT | 200969003 | Rupioid psoriasis |
| SNOMED CT | 200970002 | Psoriasis palmaris |
| SNOMED CT | 200971003 | Psoriasis plantaris |
| SNOMED CT | 200972005 | Psoriasis punctata |
| SNOMED CT | 200973000 | Pustular psoriasis |
| SNOMED CT | 200974006 | Psoriasis universalis |
| SNOMED CT | 200975007 | Psoriasis vulgaris |
| SNOMED CT | 200977004 | Erythrodermic psoriasis |
| SNOMED CT | 201436003 | Drug-induced systemic lupus erythematosus |
| SNOMED CT | 201443009 | Systemic sclerosis induced by drugs and chemicals |
| SNOMED CT | 201789001 | Rheumatoid nodule |
| SNOMED CT | 201796004 | Systemic onset juvenile chronic arthritis |
| SNOMED CT | 201805000 | Juvenile arthritis in Crohn disease |
| SNOMED CT | 201807008 | Juvenile arthritis in ulcerative colitis |
| SNOMED CT | 201964002 | Allergic arthritis of the shoulder region |
| SNOMED CT | 201967009 | Allergic arthritis of the hand |
| SNOMED CT | 201968004 | Allergic arthritis of the pelvic region and thigh |
| SNOMED CT | 201970008 | Allergic arthritis of the ankle and/or foot |
| SNOMED CT | 201972000 | Allergic arthritis of multiple sites |
| SNOMED CT | 206376005 | Sepsis of the newborn |
| SNOMED CT | 206380000 | Sepsis of newborn due to anaerobes |
| SNOMED CT | 1208834007 | Alopecia areata of eyelash |
| SNOMED CT | 213085001 | Fracture of bone allograft |
| SNOMED CT | 213086000 | Displacement of bone allograft |
| SNOMED CT | 213088004 | Rejection of bone allograft |
| SNOMED CT | 213127009 | Infection of bone allograft |
| SNOMED CT | 213150003 | Kidney transplant failure and rejection |
| SNOMED CT | 213151004 | Heart transplant failure and rejection |
| SNOMED CT | 213152006 | Heart-lung transplant failure and rejection |
| SNOMED CT | 213153001 | Liver transplant failure and rejection |
| SNOMED CT | 213192008 | Skin graft failure |
| SNOMED CT | 213203004 | Complications of attempted introduction of embryo in embryo transfer |

Continued on next page

**Table 7 – continued from previous page**

| CodeSystem | ConceptCode | ConceptName |
|---|---|---|
| SNOMED CT | 213320003 | Anaphylactic shock due to serum |
| SNOMED CT | 230372003 | Acute relapsing multiple sclerosis |
| SNOMED CT | 230591002 | Motor neuropathy with multiple conduction block |
| SNOMED CT | 231854006 | Atopic conjunctivitis |
| SNOMED CT | 231855007 | Seasonal allergic conjunctivitis |
| SNOMED CT | 231856008 | Perennial allergic conjunctivitis |
| SNOMED CT | 231857004 | Giant papillary conjunctivitis |
| SNOMED CT | 232234000 | Acute allergic otitis externa |
| SNOMED CT | 232240007 | Chronic allergic otitis externa |
| SNOMED CT | 232308006 | Autoimmune disorder of inner ear |
| SNOMED CT | 232353008 | Perennial allergic rhinitis with seasonal variation |
| SNOMED CT | 232430006 | Recurrent allergic croup |
| SNOMED CT | 232439007 | Relapsing polychondritis of larynx |
| SNOMED CT | 233634002 | Post-lung transplantation bronchiectasis |
| SNOMED CT | 233683003 | Hay fever with asthma |
| SNOMED CT | 233687002 | Colophony asthma |
| SNOMED CT | 233691007 | Eosinophilic asthma |
| SNOMED CT | 233692000 | Cryptogenic pulmonary eosinophilia |
| SNOMED CT | 233694004 | Dog house disease |
| SNOMED CT | 233695003 | Dry rot lung |
| SNOMED CT | 233696002 | Lycoperdonosis |
| SNOMED CT | 233697006 | New Guinea lung |
| SNOMED CT | 233698001 | Paprika splitters' lung |
| SNOMED CT | 233699009 | Pyrethrum alveolitis |
| SNOMED CT | 233700005 | Rodent handlers' lung |
| SNOMED CT | 233701009 | Sewage workers' lung |
| SNOMED CT | 233702002 | Summer-type hypersensitivity pneumonitis |
| SNOMED CT | 233774000 | Humidifier fever |
| SNOMED CT | 233844002 | Accelerated coronary artery disease in transplanted heart |
| SNOMED CT | 233932001 | Cardiac transplant disorder |
| SNOMED CT | 233933006 | Cardiac transplant rejection |
| SNOMED CT | 233934000 | Cardiac transplant failure |
| SNOMED CT | 233979001 | Compression of transplanted artery |
| SNOMED CT | 233971003 | Stenosis of transplant artery |
| SNOMED CT | 234001008 | Dissection of transplant artery |
| SNOMED CT | 234004000 | Hemorrhage of transplant artery |
| SNOMED CT | 234008002 | Rupture of transplanted artery |
| SNOMED CT | 234066000 | Stenosis of transplant vein |
| SNOMED CT | 234077006 | Hemorrhage of transplanted vein |
| SNOMED CT | 234079009 | Rupture of transplanted vein |
| SNOMED CT | 234381003 | Maternal autoimmune hemolytic anemia |
| SNOMED CT | 234417006 | T-cell lymphocytosis |
| SNOMED CT | 234418001 | Chronic benign granulocytopenia |

Continued on next page

**Table 7 – continued from previous page**

| CodeSystem | ConceptCode | ConceptName |
|---|---|---|
| SNOMED CT | 234423001 | Chronic benign neutropenia |
| SNOMED CT | 234426009 | Corticosteroid-induced neutrophilia |
| SNOMED CT | 234427000 | Monocytoid disorder |
| SNOMED CT | 234434003 | Alders syndrome |
| SNOMED CT | 234435002 | Hereditary hypersegmentation |
| SNOMED CT | 234513007 | Post-splenectomy leukocytosis |
| SNOMED CT | 234538002 | Hyperimmunoglobulin D with periodic fever |
| SNOMED CT | 234556002 | Specific antibody deficiency |
| SNOMED CT | 234557006 | Anti-polysaccharide antibody deficiency |
| SNOMED CT | 234559009 | Anti-pneumococcal polysaccharide antibody deficiency |
| SNOMED CT | 234560004 | Anti-meningococcal polysaccharide A antibody deficiency |
| SNOMED CT | 234561000 | Anti-meningococcal polysaccharide C antibody deficiency |
| SNOMED CT | 234562007 | Anti-protein antibody deficiency |
| SNOMED CT | 234579001 | Mannan-binding protein deficiency |
| SNOMED CT | 234586009 | Leukocyte glucose-6-phosphate dehydrogenase deficiency |
| SNOMED CT | 234589002 | Glutathione synthetase deficiency |
| SNOMED CT | 234590006 | Gluthathione peroxidase deficiency |
| SNOMED CT | 234642007 | Age-related immunodeficiency |
| SNOMED CT | 234643002 | Transient immunodeficiency of infancy |
| SNOMED CT | 234999001 | Crohn's disease of gingivae |
| SNOMED CT | 235607002 | Crohn's disease of esophagus |
| SNOMED CT | 235657005 | Sepsis-related gastritis |
| SNOMED CT | 235714007 | Ulcerative enterocolitis |
| SNOMED CT | 235728001 | Autoimmune enteropathy |
| SNOMED CT | 235890007 | Autoimmune liver disease |
| SNOMED CT | 235910007 | Liver transplant disorder |
| SNOMED CT | 235911006 | Liver transplant rejection |
| SNOMED CT | 235912004 | Liver transplant failure |
| SNOMED CT | 236436003 | End stage renal failure with renal transplant |
| SNOMED CT | 236502006 | Renal involvement in scleroderma |
| SNOMED CT | 236503001 | Acute scleroderma renal crisis |
| SNOMED CT | 236569000 | Primary non-function of renal transplant |
| SNOMED CT | 236570004 | Renal transplant rejection |
| SNOMED CT | 236571000 | Hyperacute rejection of renal transplant |
| SNOMED CT | 236572007 | Accelerated rejection of renal transplant |
| SNOMED CT | 236573002 | Very mild acute rejection of renal transplant |
| SNOMED CT | 236574008 | Acute rejection of renal transplant |
| SNOMED CT | 236575009 | Acute rejection of renal transplant - grade I |
| SNOMED CT | 236576005 | Acute rejection of renal transplant - grade II |
| SNOMED CT | 236577001 | Acute rejection of renal transplant - grade III |
| SNOMED CT | 236578006 | Chronic rejection of renal transplant |
| SNOMED CT | 236579003 | Chronic rejection of renal transplant - grade I |
| SNOMED CT | 236580000 | Chronic rejection of renal transplant - grade II |

Continued on next page

**Table 7 – continued from previous page**

| CodeSystem | ConceptCode | ConceptName |
|---|---|---|
| SNOMED CT | 236581001 | Chronic rejection of renal transplant - grade III |
| SNOMED CT | 236582008 | Acute-on-chronic rejection of renal transplant |
| SNOMED CT | 236583003 | Failed renal transplant |
| SNOMED CT | 236584009 | Perfusion injury of renal transplant |
| SNOMED CT | 236586006 | De novo glomerulonephritis |
| SNOMED CT | 236587002 | Transplant glomerulopathy |
| SNOMED CT | 236588007 | Transplant glomerulopathy - early form |
| SNOMED CT | 236589004 | Transplant glomerulopathy - late form |
| SNOMED CT | 236614007 | Perirenal and periureteric post-transplant lymphocele |
| SNOMED CT | 237499004 | Thyrotoxicosis due to Graves' disease |
| SNOMED CT | 237500008 | Hashitoxicosis - transient |
| SNOMED CT | 237519003 | Autoimmune hypothyroidism |
| SNOMED CT | 237652003 | Insulin resistance - type B |
| SNOMED CT | 237706000 | Autoimmune hypophysitis |
| SNOMED CT | 237790001 | Autoimmune primary ovarian failure |
| SNOMED CT | 237822008 | Autoimmune endocrine disease |
| SNOMED CT | 237824009 | Juvenile Graves' disease |
| SNOMED CT | 238152004 | Sepsis-related gastrointestinal ulceration |
| SNOMED CT | 238428001 | Papular tuberculide |
| SNOMED CT | 238429009 | Nodular tuberculide |
| SNOMED CT | 238476000 | Eosinophilic granuloma of bone |
| SNOMED CT | 238575004 | Allergic contact dermatitis |
| SNOMED CT | 238576003 | Allergic contact dermatitis caused by nickel from implant |
| SNOMED CT | 238577007 | Ingested allergen contact dermatitis |
| SNOMED CT | 238578002 | Occupational allergic contact dermatitis |
| SNOMED CT | 238600001 | Flexural psoriasis |
| SNOMED CT | 238601002 | Unstable psoriasis |
| SNOMED CT | 238602009 | Psoriasis with eczema |
| SNOMED CT | 238603004 | Eczematized psoriasis |
| SNOMED CT | 238604005 | Psoriasis of nail |
| SNOMED CT | 238605006 | Psoriatic nail dystrophy |
| SNOMED CT | 238606007 | Psoriatic nail pitting |
| SNOMED CT | 238607003 | Psoriatic onycholysis |
| SNOMED CT | 238608008 | Psoriasis of scalp |
| SNOMED CT | 238609000 | Köbner psoriasis |
| SNOMED CT | 238611009 | Lapiere type of psoriasis |
| SNOMED CT | 238613007 | Generalized pustular psoriasis of pregnancy |
| SNOMED CT | 238615000 | Juvenile pustular psoriasis |
| SNOMED CT | 238616004 | Infantile pustular psoriasis |
| SNOMED CT | 238617008 | Circinate and annular pustular psoriasis |
| SNOMED CT | 238672005 | Lichenoid allergic contact dermatitis |
| SNOMED CT | 238675007 | Sarcoidosis-induced erythema nodosum |
| SNOMED CT | 238676008 | Lofgrens syndrome |

Continued on next page

**Table 7 – continued from previous page**

| CodeSystem | ConceptCode | ConceptName |
|---|---|---|
| SNOMED CT | 238687000 | Familial cold urticaria |
| SNOMED CT | 238818000 | Non-bullous erythema multiforme |
| SNOMED CT | 238820002 | Erythema multiforme of pregnancy |
| SNOMED CT | 238850005 | Mouth and genital ulcers with inflamed cartilage syndrome |
| SNOMED CT | 238926009 | Lupus erythematosus and erythema multiforme-like syndrome |
| SNOMED CT | 238927000 | Chronic discoid lupus erythematosus |
| SNOMED CT | 238928005 | Chilblain lupus erythematosus |
| SNOMED CT | 238930007 | Sclerodermiform reaction |
| SNOMED CT | 238931006 | Eosinophilic cellulitis |
| SNOMED CT | 238938000 | Prebullous pemphigoid |
| SNOMED CT | 238939008 | Localized pemphigoid |
| SNOMED CT | 238940005 | Pemphigoid vegetans |
| SNOMED CT | 238942002 | Adult linear immunoglobulin A disease |
| SNOMED CT | 238943007 | Eosinophilic spongiosis |
| SNOMED CT | 239098009 | Acropustulosis of infancy |
| SNOMED CT | 239105004 | Transplacental herpes gestationis |
| SNOMED CT | 239147000 | Mast cell nevus |
| SNOMED CT | 239799007 | Early onset polyarticular juvenile chronic arthritis |
| SNOMED CT | 239800006 | Late onset polyarticular juvenile chronic arthritis |
| SNOMED CT | 239802003 | Juvenile psoriatic arthritis |
| SNOMED CT | 239803008 | Juvenile psoriatic arthritis with psoriasis |
| SNOMED CT | 239804002 | Juvenile psoriatic arthritis without psoriasis |
| SNOMED CT | 239806000 | Juvenile spondyloarthropathy |
| SNOMED CT | 239809007 | Juvenile arthritis of inflammatory bowel disease |
| SNOMED CT | 239812005 | Psoriatic arthritis with distal interphalangeal joint involvement |
| SNOMED CT | 239813000 | Psoriatic dactylitis |
| SNOMED CT | 239891002 | Subacute cutaneous lupus erythematosus |
| SNOMED CT | 239892009 | Primary antiphospholipid syndrome |
| SNOMED CT | 239894005 | Primary antiphospholipid syndrome with multisystem involvement |
| SNOMED CT | 239895006 | Secondary antiphospholipid syndrome |
| SNOMED CT | 239897003 | Secondary antiphospholipid syndrome with multisystem involvement |
| SNOMED CT | 239910001 | Toxic oil syndrome |
| SNOMED CT | 239943002 | Rheumatoid necrotizing vasculitis |
| SNOMED CT | 239946005 | Postinfective immunoglobulin A vasculitis |
| SNOMED CT | 239947001 | Essential mixed cryoglobulinemia |
| SNOMED CT | 240041006 | Rheumatic fever nodule |
| SNOMED CT | 240360007 | Yersinia erythema nodosum |
| SNOMED CT | 240385003 | Septicemic glanders |
| SNOMED CT | 240389009 | Septicemic melioidosis |

Continued on next page

**Table 7 – continued from previous page**

| CodeSystem | ConceptCode | ConceptName |
|---|---|---|
| SNOMED CT | 240426001 | Chronic meningococcemia |
| SNOMED CT | 241929008 | Acute allergic reaction |
| SNOMED CT | 241930003 | Anaphylaxis caused by venom |
| SNOMED CT | 241931004 | Bee sting-induced anaphylaxis |
| SNOMED CT | 241932006 | Wasp sting-induced anaphylaxis |
| SNOMED CT | 241933001 | Peanut-induced anaphylaxis |
| SNOMED CT | 241934007 | Seafood-induced anaphylaxis |
| SNOMED CT | 241935008 | Egg white-induced anaphylaxis |
| SNOMED CT | 241936009 | Cow's milk protein-induced anaphylaxis |
| SNOMED CT | 241937000 | Drug-induced anaphylaxis |
| SNOMED CT | 241938005 | Penicillin-induced anaphylaxis |
| SNOMED CT | 241939002 | Insulin-induced anaphylaxis |
| SNOMED CT | 241941001 | Seminal fluid-induced anaphylaxis |
| SNOMED CT | 241952007 | Exercise anaphylaxis |
| SNOMED CT | 241954008 | Idiopathic anaphylaxis |
| SNOMED CT | 241956005 | Venom-induced angioedema-urticaria |
| SNOMED CT | 241957001 | Food-induced angioedema-urticaria |
| SNOMED CT | 241965003 | Latex-induced angioedema-urticaria |
| SNOMED CT | 254289008 | Post-transplant neoplasia |
| SNOMED CT | 255101006 | Sézary disease of skin |
| SNOMED CT | 259097007 | Delayed photoallergic dermatitis |
| SNOMED CT | 262945004 | Transection of transplanted artery |
| SNOMED CT | 266451002 | Allergic gastroenteritis and colitis |
| SNOMED CT | 267454002 | Acatalasemia |
| SNOMED CT | 267800008 | Benign mucous membrane pemphigoid with no eye involvement |
| SNOMED CT | 272006008 | Gonococcal arthritis dermatitis syndrome |
| SNOMED CT | 274083001 | Tinea kerion of beard |
| SNOMED CT | 275446004 | Gardner-Diamond syndrome |
| SNOMED CT | 276177000 | Thyroid eye disease |
| SNOMED CT | 276229008 | Sepsis-associated gastrointestinal hemorrhage |
| SNOMED CT | 276288002 | Recurrent salmonella septicemia |
| SNOMED CT | 276575001 | Autoimmune neonatal thrombocytopenia |
| SNOMED CT | 276576000 | Neutropenia of the small for gestational age baby |
| SNOMED CT | 276669000 | Bacterial sepsis of newborn |
| SNOMED CT | 276741000 | Sepsis-associated myocardial dysfunction |
| SNOMED CT | 277010001 | Unexplained episode of renal transplant dysfunction |
| SNOMED CT | 277011002 | Pre-existing disease in renal transplant |
| SNOMED CT | 277475006 | Pemphigus neonatorum |
| SNOMED CT | 277577000 | Monoclonal gammopathy of uncertain significance |
| SNOMED CT | 277638005 | Sepsis-associated left ventricular failure |
| SNOMED CT | 277639002 | Sepsis-associated right ventricular failure |
| SNOMED CT | 277785006 | Penicillin-induced angioedema-urticaria |

Continued on next page

**Table 7 – continued from previous page**

| CodeSystem | ConceptCode | ConceptName |
|---|---|---|
| SNOMED CT | 278484009 | Tropical pulmonary eosinophilia |
| SNOMED CT | 282093000 | Allergic reaction to venom |
| SNOMED CT | 282094006 | Allergic reaction to insect venom |
| SNOMED CT | 282095007 | Allergic reaction to bee sting |
| SNOMED CT | 285430002 | Light chain monoclonal gammopathy of uncertain significance |
| SNOMED CT | 286947004 | Chronic rheumatic mitral valvulitis |
| SNOMED CT | 295046003 | Ulcerative proctocolitis |
| SNOMED CT | 297940005 | Drug-induced erythema nodosum |
| SNOMED CT | 297942002 | Drug-induced erythema multiforme |
| SNOMED CT | 298285004 | Systemic sclerosis with limited cutaneous involvement |
| SNOMED CT | 299276009 | Limited systemic sclerosis |
| SNOMED CT | 300909004 | Allergic reaction to wasp sting |
| SNOMED CT | 300985007 | Coliform septicemia |
| SNOMED CT | 300986008 | Proteus septicemia |
| SNOMED CT | 303011007 | Neutropenic disorder |
| SNOMED CT | 307591004 | Malignant mastocytosis |
| SNOMED CT | 309742004 | Drug-induced autoimmune hemolytic anemia |
| SNOMED CT | 310649002 | Vancomycin resistant enterococcal septicemia |
| SNOMED CT | 310669007 | Septicemia due to enterococcus |
| SNOMED CT | 312591002 | Acute rheumatic heart disease |
| SNOMED CT | 312603000 | Lung transplant rejection |
| SNOMED CT | 313437008 | Meningococcal meningitis with acute meningococcal septicemia |
| SNOMED CT | 314002005 | Corneal allograft rejection |
| SNOMED CT | 314130008 | Meningococcal meningitis with meningococcal septicemia |
| SNOMED CT | 314498009 | Contact lens related giant papillary conjunctivitis |
| SNOMED CT | 314551004 | Corneal graft disorder |
| SNOMED CT | 314552006 | Corneal graft astigmatism |
| SNOMED CT | 314553001 | Corneal graft infection |
| SNOMED CT | 314554007 | Corneal graft vascularization |
| SNOMED CT | 314757003 | Ocular cicatricial pemphigoid |
| SNOMED CT | 314760005 | Corneal graft failure |
| SNOMED CT | 317349009 | Vernal keratoconjunctivitis |
| SNOMED CT | 318316003 | Vernal conjunctivitis |
| SNOMED CT | 320150004 | Idiopathic eosinophilia |
| SNOMED CT | 361098001 | Allergic transfusion reaction |
| SNOMED CT | 361106007 | Transfusion reaction due to allergen in blood |
| SNOMED CT | 361125005 | Subacute autoimmune thyroiditis |
| SNOMED CT | 361212009 | Oral pemphigus vulgaris |
| SNOMED CT | 362985009 | Antibody-mediated activation and inactivation |
| SNOMED CT | 362986005 | Antibody-mediated cytolysis |
| SNOMED CT | 362992004 | Autoimmune connective tissue disorder |

Continued on next page

**Table 7 – continued from previous page**

| CodeSystem | ConceptCode | ConceptName |
|---|---|---|
| SNOMED CT | 367498001 | Seasonal allergic rhinitis |
| SNOMED CT | 367542003 | Pulmonary eosinophilia |
| SNOMED CT | 370474006 | Eosinophilic myositis |
| SNOMED CT | 371044001 | Transient respiratory distress with sepsis |
| SNOMED CT | 373674001 | Anaphylactic urticaria |
| SNOMED CT | 389145006 | Allergic asthma |
| SNOMED CT | 397012002 | Cutaneous mastocytosis |
| SNOMED CT | 397013007 | Solitary cutaneous mastocytoma |
| SNOMED CT | 397014001 | Diffuse erythrodermic mastocytosis |
| SNOMED CT | 397173003 | Crohn's disease of intestine |
| SNOMED CT | 397843005 | Sepsis-related gastrointestinal lesions |
| SNOMED CT | 397856003 | Systemic lupus erythematosus-related syndrome |
| SNOMED CT | 398049005 | Mixed collagen vascular disease |
| SNOMED CT | 398726004 | Rheumatoid lung disease |
| SNOMED CT | 398937006 | Cold autoimmune hemolytic anemia |
| SNOMED CT | 399456004 | Plasma cell myeloma/plasmacytoma |
| SNOMED CT | 399946006 | Crohn's disease of skin |
| SNOMED CT | 400054000 | Rheumatoid vasculitis |
| SNOMED CT | 402188000 | Atopic dermatitis aggravated by type 1 immune reaction |
| SNOMED CT | 402244002 | Lichenoid allergic contact reaction due to color developer |
| SNOMED CT | 402245001 | Angry back syndrome |
| SNOMED CT | 402247009 | Lichenoid allergic contact reaction affecting oral mucosa |
| SNOMED CT | 402248004 | Lichenoid allergic contact reaction due to tattoo ink |
| SNOMED CT | 402249007 | Allergic contact dermatitis of eyelid |
| SNOMED CT | 402250007 | Allergic contact cheilitis |
| SNOMED CT | 402251006 | Allergic contact gingivostomatitis |
| SNOMED CT | 402252004 | Allergic contact dermatitis of perianal skin |
| SNOMED CT | 402254003 | Allergic contact dermatitis of hands |
| SNOMED CT | 402255002 | Allergic contact dermatitis of lower leg |
| SNOMED CT | 402256001 | Allergic contact dermatitis of male genitalia |
| SNOMED CT | 402257005 | Allergic contact dermatitis of female genitalia |
| SNOMED CT | 402304007 | Allergic contact urticaria |
| SNOMED CT | 402307000 | Chronic large plaque psoriasis |
| SNOMED CT | 402308005 | Chronic small plaque psoriasis |
| SNOMED CT | 402309002 | Chronic guttate pattern psoriasis |
| SNOMED CT | 402310007 | Chronic stable plaque psoriasis |
| SNOMED CT | 402311006 | Actively extending plaque psoriasis |
| SNOMED CT | 402312004 | Acute guttate psoriasis |
| SNOMED CT | 402313009 | Guttate flare of psoriasis with preexisting plaques |
| SNOMED CT | 402314003 | Psoriasis of vulva |
| SNOMED CT | 402315002 | Psoriasis of penis |
| SNOMED CT | 402316001 | Psoriasis of perianal skin |
| SNOMED CT | 402317005 | Generalized psoriasis |

Continued on next page

**Table 7 – continued from previous page**

| CodeSystem | ConceptCode | ConceptName |
|---|---|---|
| SNOMED CT | 402318000 | Photoaggravated psoriasis |
| SNOMED CT | 402319008 | Psoriasis of scalp margin |
| SNOMED CT | 402320002 | Psoriasis of face |
| SNOMED CT | 402321003 | Non-pustular psoriasis of hands |
| SNOMED CT | 402322005 | Hypertrophic palmar psoriasis |
| SNOMED CT | 402323000 | Hypertrophic palmoplantar psoriasis |
| SNOMED CT | 402324006 | Non-pustular psoriasis of hands and feet |
| SNOMED CT | 402325007 | Drug-exacerbated psoriasis |
| SNOMED CT | 402326008 | Acute generalized pustular psoriasis de novo |
| SNOMED CT | 402327004 | Acute generalized pustular flare of preexisting plaque psoriasis |
| SNOMED CT | 402329001 | Early onset psoriasis type 1 |
| SNOMED CT | 402330006 | Onset of psoriasis in infancy (<1 year) |
| SNOMED CT | 402331005 | Onset of psoriasis in childhood (1-10 years) |
| SNOMED CT | 402332003 | Onset of psoriasis in adolescence (10-20 years) |
| SNOMED CT | 402333008 | Onset of psoriasis in early adulthood (20-40 years) |
| SNOMED CT | 402334002 | Late onset psoriasis type 2 |
| SNOMED CT | 402335001 | Familial psoriasis |
| SNOMED CT | 402336000 | Familial psoriasis with affected first degree relative |
| SNOMED CT | 402337009 | Familial psoriasis without affected first degree relative |
| SNOMED CT | 402357008 | Lichenoid chronic graft-versus-host disease |
| SNOMED CT | 402358003 | Sclerodermoid chronic graft-versus-host disease |
| SNOMED CT | 402360001 | Materno-fetal graft-versus-host disease |
| SNOMED CT | 402375009 | Crohn's disease of vulva |
| SNOMED CT | 402376005 | Crohn's disease of penis |
| SNOMED CT | 402377001 | Crohn's disease of scrotum |
| SNOMED CT | 402384009 | Allergic urticaria due to ingested food |
| SNOMED CT | 402385005 | Allergic urticaria due to bite and/or sting |
| SNOMED CT | 402386006 | Allergic urticaria due to inhaled allergen |
| SNOMED CT | 402387002 | Allergic angioedema |
| SNOMED CT | 402388007 | Allergic angioedema due to ingested food |
| SNOMED CT | 402389004 | Allergic angioedema due to bite and/or sting |
| SNOMED CT | 402390008 | Anaphylaxis due to ingested food |
| SNOMED CT | 402392000 | Allergic urticaria and/or angioedema |
| SNOMED CT | 402397006 | Autoimmune urticaria |
| SNOMED CT | 402398001 | Acute autoimmune urticaria |
| SNOMED CT | 402399009 | Chronic autoimmune urticaria |
| SNOMED CT | 402400002 | Episodic autoimmune urticaria |
| SNOMED CT | 402401003 | Autoimmune angioedema |
| SNOMED CT | 402402005 | Autoimmune urticaria and/or angioedema |
| SNOMED CT | 402404006 | Episodic angioedema with eosinophilia |
| SNOMED CT | 402426007 | Rheumatoid nodulosis |
| SNOMED CT | 402427003 | Accelerated rheumatoid nodulosis |

Continued on next page

**Table 7 – continued from previous page**

| CodeSystem | ConceptCode | ConceptName |
|---|---|---|
| SNOMED CT | 402428008 | Accelerated rheumatoid nodulosis caused by methotrexate |
| SNOMED CT | 402431009 | Cutaneous atrophy due to rheumatoid arthritis |
| SNOMED CT | 402432002 | Rheumatoid neutrophilic dermatitis |
| SNOMED CT | 402433007 | Rheumatoid arthritis with nailfold and finger-pulp infarcts |
| SNOMED CT | 402434001 | Cutaneous complication of rheumatoid disease |
| SNOMED CT | 402435000 | Eosinophilic spongiosis (prebullous pemphigus) |
| SNOMED CT | 402436004 | Eosinophilic spongiosis (prebullous pemphigoid) |
| SNOMED CT | 402437008 | Localized bullous pemphigoid of vulva |
| SNOMED CT | 402438003 | Pemphigoid nodularis |
| SNOMED CT | 402439006 | Childhood bullous pemphigoid |
| SNOMED CT | 402440008 | Oral mucous membrane involvement by bullous pemphigoid |
| SNOMED CT | 402441007 | Oral involvement by mucous membrane pemphigoid |
| SNOMED CT | 402442000 | Brunsting-Perry type cicatricial pemphigoid |
| SNOMED CT | 402444004 | Oral mucosal involvement by dermatitis herpetiformis |
| SNOMED CT | 402447006 | Epidermolysis bullosa acquisita, classical acral type |
| SNOMED CT | 402448001 | Epidermolysis bullosa acquisita, bullous pemphigoid-like |
| SNOMED CT | 402449009 | Epidermolysis bullosa acquisita, cicatricial pemphigoid-like |
| SNOMED CT | 402450009 | Epidermolysis bullosa acquisita, Brunsting-Perry type |
| SNOMED CT | 402451008 | Epidermolysis bullosa acquisita, oral mucosal involvement |
| SNOMED CT | 402576006 | Lichenoid allergic contact reaction |
| SNOMED CT | 402712002 | Occupational scleroderma |
| SNOMED CT | 402717008 | IgA pemphigus |
| SNOMED CT | 402718003 | Pemphigus paraneoplastica |
| SNOMED CT | 402856005 | Acute hemorrhagic edema of childhood |
| SNOMED CT | 402865003 | Systemic lupus erythematosus-associated antiphospholipid syndrome |
| SNOMED CT | 402961006 | Pseudomonas septicemia with skin involvement |
| SNOMED CT | 402968000 | Erythema nodosum caused by Bacteria |
| SNOMED CT | 402969008 | Erythema nodosum caused by Genus Streptococcus |
| SNOMED CT | 402970009 | Erythema nodosum caused by Yersinia enterocolitica |
| SNOMED CT | 402971008 | Erythema multiforme due to mycoplasma infection |
| SNOMED CT | 403211002 | Papular form erythema multiforme |
| SNOMED CT | 403212009 | Vesicobullous form erythema multiforme |
| SNOMED CT | 403213004 | Erythema multiforme due to malignancy |
| SNOMED CT | 403217003 | Subcorneal pustular dermatosis with paraproteinemia |
| SNOMED CT | 403320008 | Circumscribed alopecia areata of scalp |
| SNOMED CT | 403321007 | Circumscribed alopecia areata of eyelashes/eyebrows |
| SNOMED CT | 403322000 | Circumscribed alopecia areata of beard area |
| SNOMED CT | 403323005 | Circumscribed alopecia areata of limbs |
| SNOMED CT | 403324004 | Circumscribed alopecia areata of trunk |
| SNOMED CT | 403434009 | Atopic keratoconjunctivitis |
| SNOMED CT | 403489007 | Subacute cutaneous lupus erythematosus, annular/polycyclic type |

Continued on next page

**Table 7 – continued from previous page**

| CodeSystem | ConceptCode | ConceptName |
|---|---|---|
| SNOMED CT | 403490003 | Subacute cutaneous lupus erythematosus, papulosquamous type |
| SNOMED CT | 403491004 | Erythema multiforme-like lupus erythematosus |
| SNOMED CT | 403492006 | Hypertrophic type discoid lupus erythematosus |
| SNOMED CT | 403493001 | Rosaceous type discoid lupus erythematosus |
| SNOMED CT | 403494007 | Discoid lupus erythematosus of mucous membranes |
| SNOMED CT | 403495008 | Discoid lupus erythematosus of oral mucosa |
| SNOMED CT | 403496009 | Discoid lupus erythematosus of genital mucous membranes |
| SNOMED CT | 403497000 | Discoid lupus erythematosus of scalp |
| SNOMED CT | 403498005 | Discoid lupus erythematosus of face |
| SNOMED CT | 403499002 | Discoid lupus erythematosus of lip |
| SNOMED CT | 403500006 | Discoid lupus erythematosus of hands |
| SNOMED CT | 403501005 | Discoid lupus erythematosus of foot |
| SNOMED CT | 403502003 | Disseminated discoid lupus erythematosus |
| SNOMED CT | 403513000 | Lupus erythematosus-associated nail dystrophy |
| SNOMED CT | 403629000 | Drug-induced bullous pemphigoid |
| SNOMED CT | 403630005 | Drug-induced pemphigus vulgaris |
| SNOMED CT | 403631009 | Drug-induced pemphigus foliaceus |
| SNOMED CT | 403632002 | Drug-induced epidermolysis bullosa acquisita |
| SNOMED CT | 403735006 | Eosinophilia-myalgia syndrome from tryptophan |
| SNOMED CT | 404145009 | Primary cutaneous anaplastic large cell B-cell lymphoma |
| SNOMED CT | 404165002 | Necrobiotic xanthogranuloma with paraproteinemia |
| SNOMED CT | 404170009 | Bullous urticaria pigmentosa |
| SNOMED CT | 404173006 | Familial mastocytosis |
| SNOMED CT | 404804003 | Platinum asthma |
| SNOMED CT | 404805002 | Platinum sensitivity |
| SNOMED CT | 404807005 | Cheese-washers' lung |
| SNOMED CT | 405720007 | Allergic bronchitis |
| SNOMED CT | 405821001 | Allergic otitis externa |
| SNOMED CT | 408335007 | Autoimmune hepatitis |
| SNOMED CT | 408539000 | Insulin autoimmune syndrome |
| SNOMED CT | 409089005 | Febrile neutropenia |
| SNOMED CT | 410482007 | Iritis with psoriatic arthritis |
| SNOMED CT | 410484008 | Iritis with ulcerative colitis |
| SNOMED CT | 410502007 | Juvenile rheumatoid arthritis |
| SNOMED CT | 410796000 | Juvenile seropositive polyarthritis |
| SNOMED CT | 410797009 | Juvenile seronegative polyarthritis |
| SNOMED CT | 410798004 | Juvenile idiopathic arthritis, oligoarthritis |
| SNOMED CT | 410799007 | Juvenile idiopathic arthritis, persistent oligoarthritis |
| SNOMED CT | 410800006 | Juvenile idiopathic arthritis, extended oligoarthritis |
| SNOMED CT | 410801005 | Juvenile idiopathic arthritis, enthesitis related arthritis |
| SNOMED CT | 410802003 | Juvenile idiopathic arthritis, undifferentiated arthritis |
| SNOMED CT | 413603009 | Autoimmune hemolytic anemia |

Continued on next page

**Table 7 – continued from previous page**

| CodeSystem | ConceptCode | ConceptName |
|---|---|---|
| SNOMED CT | 414061000 | Drug-induced cicatricial pemphigoid |
| SNOMED CT | 414156000 | Exacerbation of ulcerative colitis |
| SNOMED CT | 414450004 | Idiopathic hypereosinophilic syndrome |
| SNOMED CT | 414850009 | Neutrophilia disorder |
| SNOMED CT | 416093006 | Allergic reaction to drug |
| SNOMED CT | 417389002 | Disorder of basophils |
| SNOMED CT | 417516000 | Anaphylaxis due to substance |
| SNOMED CT | 417967008 | Disorder of eosinophil |
| SNOMED CT | 418176003 | Allergic reaction to plant, except food |
| SNOMED CT | 418282006 | Allergic reaction to cosmetics |
| SNOMED CT | 418325008 | Allergic reaction to adhesive |
| SNOMED CT | 418364006 | Allergic reaction to pollen |
| SNOMED CT | 418367004 | Allergic reaction to grass pollen |
| SNOMED CT | 418448002 | Allergic reaction to animal |
| SNOMED CT | 418579000 | Allergic reaction to colophony |
| SNOMED CT | 418634005 | Allergic reaction to substance |
| SNOMED CT | 418943003 | Allergic reaction to tree pollen |
| SNOMED CT | 419076005 | Allergic reaction |
| SNOMED CT | 419121001 | Allergic contact dermatitis of pinna |
| SNOMED CT | 419375001 | Allergic reaction to oil |
| SNOMED CT | 419447004 | Allergic reaction to flour dust |
| SNOMED CT | 419452009 | Allergic reaction to food |
| SNOMED CT | 419455006 | Disorder characterized by eosinophilia |
| SNOMED CT | 419515007 | Allergic reaction to detergent |
| SNOMED CT | 419519001 | Allergic reaction to weed pollen |
| SNOMED CT | 419838000 | Allergic reaction to chemical |
| SNOMED CT | 419884005 | Allergic reaction to flea bite |
| SNOMED CT | 419957005 | Allergic contact dermatitis of external auditory canal |
| SNOMED CT | 420198007 | Allergic reaction to platinum |
| SNOMED CT | 420373003 | Allergic disorder of digestive system |
| SNOMED CT | 420505005 | Corneal endothelial allograft rejection |
| SNOMED CT | 420571002 | Corneal subepithelial allograft rejection |
| SNOMED CT | 420881009 | Allergic disorder caused by substance |
| SNOMED CT | 421092003 | Allergic disorder of respiratory system |
| SNOMED CT | 421187003 | Corneal epithelial allograft rejection |
| SNOMED CT | 422290000 | Allergic disorder of skin |
| SNOMED CT | 423486005 | Disseminated eosinophilic collagen disease |
| SNOMED CT | 424643009 | IgE-mediated allergic asthma |
| SNOMED CT | 425011002 | Allergic fungal sinusitis |
| SNOMED CT | 425500002 | Secondary progressive multiple sclerosis |
| SNOMED CT | 426065008 | Hypersensitivity disorder mediated by immune complex |
| SNOMED CT | 426136000 | Delayed renal graft function |
| SNOMED CT | 426364002 | Allergic hepatitis |

Continued on next page

**Table 7 – continued from previous page**

| CodeSystem | ConceptCode | ConceptName |
|---|---|---|
| SNOMED CT | 426373005 | Relapsing remitting multiple sclerosis |
| SNOMED CT | 426549001 | Crohn's disease in remission |
| SNOMED CT | 426760008 | Delayed hypersensitivity disorder |
| SNOMED CT | 426871003 | Cell-mediated cytotoxic disorder |
| SNOMED CT | 427213005 | Autoimmune vasculitis |
| SNOMED CT | 427245000 | Febrile leukopenia |
| SNOMED CT | 427435004 | Punctate keratopathy after penetrating keratoplasty |
| SNOMED CT | 427439005 | Immune hypersensitivity disorder by mechanism |
| SNOMED CT | 427833000 | Anaphylaxis due to mollusk |
| SNOMED CT | 427889009 | Hypertension associated with transplantation |
| SNOMED CT | 427903006 | Anaphylaxis due to fish |
| SNOMED CT | 427927008 | Disorder related to cardiac transplantation |
| SNOMED CT | 427928003 | Disorder related to lung transplantation |
| SNOMED CT | 428218004 | Allergic arthritis of spine |
| SNOMED CT | 428470000 | Autoimmune sensorineural hearing loss |
| SNOMED CT | 428575007 | Hypertension secondary to kidney transplant |
| SNOMED CT | 428700003 | Primary progressive multiple sclerosis |
| SNOMED CT | 428795003 | Anaphylaxis due to vegetable |
| SNOMED CT | 429195002 | Allergic rhinitis due to animal dander |
| SNOMED CT | 429257001 | Disorder of transplanted heart |
| SNOMED CT | 429451003 | Disorder related to renal transplantation |
| SNOMED CT | 429751004 | Anaphylaxis due to fruit |
| SNOMED CT | 430822009 | Basophilic leukaemoid reaction |
| SNOMED CT | 430980000 | Anaphylaxis due to hymenoptera venom |
| SNOMED CT | 431186002 | Accelerated rejection of cardiac transplant |
| SNOMED CT | 431222008 | Acute rejection of liver transplant |
| SNOMED CT | 431223003 | Acute rejection of lung transplant |
| SNOMED CT | 431505005 | Rejection of intestine transplant |
| SNOMED CT | 431507002 | Accelerated rejection of lung transplant |
| SNOMED CT | 431896008 | Chronic rejection of cardiac transplant |
| SNOMED CT | 432265007 | Rejection of corneal graft after penetrating keratoplasty |
| SNOMED CT | 432772009 | Hyperacute rejection of liver transplant |
| SNOMED CT | 432773004 | Hyperacute rejection of cardiac transplant |
| SNOMED CT | 432774005 | Hyperacute rejection of lung transplant |
| SNOMED CT | 432777003 | Accelerated rejection of liver transplant |
| SNOMED CT | 432843002 | Acute rejection of cardiac transplant |
| SNOMED CT | 432908002 | Chronic rejection of liver transplant |
| SNOMED CT | 432958009 | Chronic rejection of lung transplant |
| SNOMED CT | 432989008 | Allergic reaction to tattoo ink |
| SNOMED CT | 433087001 | Primary failure of corneal graft after penetrating keratoplasty |
| SNOMED CT | 433600001 | Hyperacute rejection of intestine transplant |
| SNOMED CT | 433804007 | Accelerated rejection of intestine transplant |

Continued on next page

**Table 7 – continued from previous page**

| CodeSystem | ConceptCode | ConceptName |
|---|---|---|
| SNOMED CT | 434238001 | Acute rejection of intestine transplant |
| SNOMED CT | 434270001 | Chronic rejection of intestine transplant |
| SNOMED CT | 438511000 | Benign multiple sclerosis |
| SNOMED CT | 439567002 | Malignant multiple sclerosis |
| SNOMED CT | 441492003 | Anaphylaxis due to seed |
| SNOMED CT | 441495001 | Anaphylaxis due to tree nut |
| SNOMED CT | 441593005 | Anaphylaxis due to latex |
| SNOMED CT | 441971007 | Chronic left-sided ulcerative colitis |
| SNOMED CT | 441978001 | Allergic rhinitis due to food |
| SNOMED CT | 442052005 | Anaphylaxis due to shellfish |
| SNOMED CT | 442159003 | Chronic ulcerative pancolitis |
| SNOMED CT | 443872005 | Systemic sclerosis due to chemical |
| SNOMED CT | 443899007 | Autoimmune inflammation of skeletal muscle |
| SNOMED CT | 443980004 | Neutropenic sepsis |
| SNOMED CT | 444133002 | Progressive systemic sclerosis |
| SNOMED CT | 444546002 | Chronic ulcerative colitis |
| SNOMED CT | 444547006 | Cutaneous graft-versus-host disease |
| SNOMED CT | 444548001 | Ulcerative pancolitis |
| SNOMED CT | 444855007 | Arteriosclerosis of coronary artery bypass graft of transplanted heart |
| SNOMED CT | 445030005 | Blastic plasmacytoid dendritic cell neoplasm |
| SNOMED CT | 445187004 | Antisynthetase syndrome |
| SNOMED CT | 445243001 | Left sided ulcerative colitis |
| SNOMED CT | 445260006 | Posttransplant diabetes mellitus |
| SNOMED CT | 445261005 | Posttransplant hyperlipidemia |
| SNOMED CT | 446021007 | Limbal vernal conjunctivitis |
| SNOMED CT | 446096008 | Perennial allergic rhinitis |
| SNOMED CT | 446682003 | Pediatric autoimmune neuropsychiatric disorder associated with streptococcal infection |
| SNOMED CT | 447843005 | Severe sepsis with acute organ dysfunction caused by anaerobic bacteria |
| SNOMED CT | 448542008 | Autoimmune pancreatitis |
| SNOMED CT | 448784003 | Sepsis due to herpes simplex |
| SNOMED CT | 449729000 | Allergic rhinitis due to house dust mite |
| SNOMED CT | 449730005 | Autoimmune polyendocrine syndrome type 4 |
| SNOMED CT | 449731009 | Autoimmune polyendocrine syndrome type 3 |
| SNOMED CT | 1197483002 | Functional disorder of polymorphonuclear neutrophil |
| SNOMED CT | 473104002 | Allergic reaction to hair dye |
| SNOMED CT | 473460002 | Allergic conjunctivitis |
| SNOMED CT | 608874000 | Eaton Lambert syndrome with underlying malignancy |
| SNOMED CT | 609329007 | Catastrophic antiphospholipid syndrome |
| SNOMED CT | 609485004 | Induced termination of pregnancy complicated by sepsis |
| SNOMED CT | 609552005 | Allergic reaction to radiocontrast media |

Continued on next page

**Table 7 – continued from previous page**

| CodeSystem | ConceptCode | ConceptName |
|---|---|---|
| SNOMED CT | 697969008 | Ulcerative colitis in remission |
| SNOMED CT | 698819004 | Postoperative sepsis |
| SNOMED CT | 700050004 | Overwhelming infection in asplenic patient |
| SNOMED CT | 700051000 | Sepsis in asplenic subject |
| SNOMED CT | 700052007 | Post-splenectomy sepsis |
| SNOMED CT | 700054008 | Sepsis due to fungus |
| SNOMED CT | 700055009 | Sepsis with cutaneous manifestations |
| SNOMED CT | 700057001 | Emberger syndrome |
| SNOMED CT | 700269001 | Discoid lupus erythematosus of lower eyelid |
| SNOMED CT | 700334001 | Discoid lupus erythematosus of upper eyelid |
| SNOMED CT | 700340008 | Allergic contact dermatitis of upper eyelid |
| SNOMED CT | 700341007 | Allergic contact dermatitis of lower eyelid |
| SNOMED CT | 702575003 | Retinocochleocerebral vasculopathy |
| SNOMED CT | 702617007 | Acute generalized exanthematous pustulosis |
| SNOMED CT | 703048006 | Post-renal transplant vesicoureteric reflux |
| SNOMED CT | 703133002 | Lichenoid allergic contact reaction due to amalgam |
| SNOMED CT | 703538003 | Mannose-binding lectin deficiency |
| SNOMED CT | 703802001 | Acquired angioedema due to C1 inhibitor autoantibody |
| SNOMED CT | 707148007 | Recurrent post-transplant renal disease |
| SNOMED CT | 707271004 | Sepsis due to oral infection |
| SNOMED CT | 707301001 | Lupus erythematosus of oral mucous membrane |
| SNOMED CT | 707314000 | Erythema multiforme co-occurrent with ulcer of oral mucous membrane |
| SNOMED CT | 707441009 | Milk-induced pulmonary disease in infant |
| SNOMED CT | 707443007 | Autoimmune pulmonary alveolar proteinosis |
| SNOMED CT | 707986004 | Mucous membrane pemphigoid with esophageal involvement |
| SNOMED CT | 708093000 | Acute exacerbation of allergic asthma |
| SNOMED CT | 708095007 | Acute severe exacerbation of immunoglobin E-mediated allergic asthma |
| SNOMED CT | 710027002 | Cytokine release syndrome |
| SNOMED CT | 711160007 | Eosinophil peroxidase deficiency |
| SNOMED CT | 711435001 | Chronic migratory panniculitis |
| SNOMED CT | 711526005 | Chronic localized pemphigoid |
| SNOMED CT | 713225000 | Gingival disease due to lupus erythematosus |
| SNOMED CT | 713229006 | Gingival disease due to erythema multiforme |
| SNOMED CT | 713236007 | Allergic contact gingivitis caused by acrylic dental material |
| SNOMED CT | 713654004 | Autoimmune cholangitis |
| SNOMED CT | 713825007 | Transplant renal artery stenosis |
| SNOMED CT | 713854001 | Perinatal sepsis |
| SNOMED CT | 713865006 | Perinatal sepsis caused by Streptococcus agalactiae |
| SNOMED CT | 713866007 | Perinatal sepsis caused by Escherichia coli |
| SNOMED CT | 714148005 | Allergic contact gingivitis caused by nickel |

Continued on next page

**Table 7 – continued from previous page**

| CodeSystem | ConceptCode | ConceptName |
|---|---|---|
| SNOMED CT | 714149002 | Allergic contact gingivitis caused by mercury |
| SNOMED CT | 714522006 | Allergic gingival disease |
| SNOMED CT | 715070000 | Sepsis caused by Peptostreptococcus |
| SNOMED CT | 715071001 | Sepsis caused by anaerobic streptococcus |
| SNOMED CT | 715401008 | Reynolds syndrome |
| SNOMED CT | 715770009 | Acute motor axonal neuropathy |
| SNOMED CT | 715840006 | Pemphigus vulgaris of gingival mucous membrane |
| SNOMED CT | 715863001 | Autoimmune necrotizing myopathy |
| SNOMED CT | 715887004 | Anaphylaxis caused by sulfite salt |
| SNOMED CT | 716374005 | Anaphylaxis caused by sulfur dioxide |
| SNOMED CT | 716722005 | Acute motor sensory axonal neuropathy |
| SNOMED CT | 716723000 | Acute inflammatory demyelinating polyradiculoneuropathy |
| SNOMED CT | 717780007 | Monoclonal B-cell lymphocytosis |
| SNOMED CT | 717872008 | Allergic duodenitis |
| SNOMED CT | 718175009 | Macrophagic myofasciitis |
| SNOMED CT | 718471008 | Oral pemphigoid of gingival mucous membrane |
| SNOMED CT | 718716008 | Autoimmune hemolytic anemia mixed type |
| SNOMED CT | 719040004 | Anaphylaxis caused by lupin flour |
| SNOMED CT | 720497002 | Anti-human leukocyte antigen hyperimmunization |
| SNOMED CT | 720587009 | Donor derived melanoma |
| SNOMED CT | 720588004 | Non-immunoglobulin E-mediated drug sensitivity |
| SNOMED CT | 720980004 | Alopecia, psychomotor epilepsy, periodontal pyorrhea, intellectual disability syndrome |
| SNOMED CT | 721093000 | Dianzani autoimmune lymphoproliferative disease |
| SNOMED CT | 721104000 | Sepsis due to urinary tract infection |
| SNOMED CT | 721196005 | Pemphigus of vulva |
| SNOMED CT | 721278008 | Neonatal sepsis caused by Malassezia |
| SNOMED CT | 721307000 | Transient abnormal myelopoiesis |
| SNOMED CT | 721538000 | Psoriasis of anogenital region |
| SNOMED CT | 721572008 | Sepsis caused by Pseudomonas aeruginosa |
| SNOMED CT | 721686000 | Inflammatory polyps co-occurrent and due to inflammatory bowel disease |
| SNOMED CT | 721702009 | Crohn disease of anal canal |
| SNOMED CT | 721711009 | Autoimmune hepatitis type 1 |
| SNOMED CT | 721712002 | Autoimmune hepatitis type 2 |
| SNOMED CT | 721713007 | Autoimmune hepatitis type 3 |
| SNOMED CT | 722386009 | Celiac disease with epilepsy and cerebral calcification syndrome |
| SNOMED CT | 722401001 | Severe fever with thrombocytopenia syndrome virus |
| SNOMED CT | 722453009 | Hennekam Beemer syndrome |
| SNOMED CT | 722850002 | Crohn's disease of gastrointestinal anastomosis |
| SNOMED CT | 722870008 | IgG4-related sclerosing cholangitis |
| SNOMED CT | 722872000 | Autoimmune pancreatitis type 1 |

Continued on next page

**Table 7 – continued from previous page**

| CodeSystem | ConceptCode | ConceptName |
|---|---|---|
| SNOMED CT | 722925004 | Transient neonatal neutropenia due to congenital viral infection |
| SNOMED CT | 722926003 | Transient neonatal neutropenia due to neonatal bacterial sepsis |
| SNOMED CT | 722991004 | Autoimmune myopathy |
| SNOMED CT | 723004005 | Autoimmune bullous dermatosis |
| SNOMED CT | 723022008 | Autoimmune keratitis |
| SNOMED CT | 723674005 | Simple cryoglobulinemia |
| SNOMED CT | 724603009 | Pediatric onset systemic sclerosis |
| SNOMED CT | 724643004 | Transient abnormal myelopoiesis co-occurrent with Down syndrome |
| SNOMED CT | 724778008 | Progressive relapsing multiple sclerosis |
| SNOMED CT | 724782005 | Demyelination of central nervous system co-occurrent and due to Sjogren disease |
| SNOMED CT | 724809006 | Autoimmune retinopathy |
| SNOMED CT | 724825001 | Allergic contact dermatitis caused by hairdressing product |
| SNOMED CT | 724826000 | Allergic contact blepharoconjunctivitis |
| SNOMED CT | 725415009 | House allergic alveolitis |
| SNOMED CT | 726078000 | Sporadic Blau syndrome |
| SNOMED CT | 726101009 | Extraintestinal Crohn's |
| SNOMED CT | 732960002 | Secondary autoimmune hemolytic anemia co-occurrent and due to systemic lupus erythematosus |
| SNOMED CT | 732962005 | Secondary autoimmune hemolytic anemia co-occurrent and due to chronic inflammatory disease |
| SNOMED CT | 732963000 | Secondary autoimmune hemolytic anemia co-occurrent and due to lymphoproliferative disorder |
| SNOMED CT | 732965007 | Secondary autoimmune hemolytic anemia co-occurrent and due to rheumatic disorder |
| SNOMED CT | 732966008 | Secondary autoimmune hemolytic anemia co-occurrent and due to ulcerative colitis |
| SNOMED CT | 733142005 | Sepsis following obstructed labor |
| SNOMED CT | 733155006 | IgE mediated allergic enteritis of small intestine |
| SNOMED CT | 733157003 | Crohn disease of appendix |
| SNOMED CT | 733207001 | Necrosis of myocutaneous flap |
| SNOMED CT | 735173007 | Shock due to anaphylaxis |
| SNOMED CT | 735434003 | Acquired neutrophilia |
| SNOMED CT | 735435002 | Constitutional neutrophilia |
| SNOMED CT | 735439008 | Constitutional eosinopenia |
| SNOMED CT | 735440005 | Acquired eosinopenia |
| SNOMED CT | 735442002 | Acquired eosinophilia |
| SNOMED CT | 735444001 | Acquired lymphocytosis |
| SNOMED CT | 735445000 | Allergic disorder of respiratory tract |
| SNOMED CT | 735447008 | Anaphylaxis caused by insect venom |

Continued on next page

**Table 7 – continued from previous page**

| CodeSystem | ConceptCode | ConceptName |
|---|---|---|
| SNOMED CT | 735448003 | Anaphylaxis due to mast cell disorder |
| SNOMED CT | 735454002 | Allergic bronchospasm caused by dietary substance |
| SNOMED CT | 735588005 | Uncomplicated allergic asthma |
| SNOMED CT | 735637004 | Sepsis of neonate caused by Streptococcus pyogenes |
| SNOMED CT | 735638009 | Sepsis of fetus caused by Streptococcus pyogenes |
| SNOMED CT | 735975001 | Shock co-occurrent and due to anaphylaxis caused by serum |
| SNOMED CT | 737193000 | Allergic enteritis of small intestine |
| SNOMED CT | 737195007 | Crohn disease of upper gastrointestinal tract |
| SNOMED CT | 737249005 | Photodermatitis co-occurrent and due to autoimmune disease |
| SNOMED CT | 737315000 | Food-protein induced enterocolitis syndrome |
| SNOMED CT | 737316004 | Food protein-induced colitis in infant |
| SNOMED CT | 737317008 | Food protein-induced proctitis in infant |
| SNOMED CT | 762282007 | Immunoglobulin G4 related pachymeningitis |
| SNOMED CT | 762303003 | Pediatric onset Sjögren syndrome |
| SNOMED CT | 762375006 | Erythema multiforme due to viral disease |
| SNOMED CT | 762379000 | Allergic contact dermatitis caused by dental material |
| SNOMED CT | 762380002 | Allergic contact dermatitis caused by food flavor |
| SNOMED CT | 762381003 | Allergic contact dermatitis caused by food additive |
| SNOMED CT | 762382005 | Allergic contact dermatitis caused by agrochemical biocide |
| SNOMED CT | 762383000 | Allergic contact dermatitis caused by cutting oil |
| SNOMED CT | 762384006 | Allergic contact dermatitis caused by disinfectant |
| SNOMED CT | 762386008 | Allergic contact dermatitis caused by rubber compound |
| SNOMED CT | 762387004 | Allergic contact dermatitis caused by systemic medicament |
| SNOMED CT | 762455001 | Anaphylaxis caused by low temperature |
| SNOMED CT | 762521001 | Acute allergic asthma |
| SNOMED CT | 762528007 | Sepsis caused by Klebsiella pneumoniae |
| SNOMED CT | 762529004 | Sepsis caused by Acinetobacter baumannii |
| SNOMED CT | 762545002 | Occupational allergic contact dermatitis caused by footwear |
| SNOMED CT | 762548000 | Occupational allergic contact dermatitis caused by cosmetic |
| SNOMED CT | 762549008 | Occupational allergic contact dermatitis caused by fragrance |
| SNOMED CT | 762552000 | Occupational allergic contact dermatitis caused by dental material |
| SNOMED CT | 762554004 | Occupational allergic contact dermatitis caused by food flavor |
| SNOMED CT | 762555003 | Occupational allergic contact dermatitis caused by food additive |
| SNOMED CT | 762558001 | Occupational allergic contact dermatitis caused by hairdressing product |
| SNOMED CT | 762560004 | Occupational allergic contact dermatitis caused by agrochemical biocide |

Continued on next page

**Table 7 – continued from previous page**

| CodeSystem | ConceptCode | ConceptName |
|---|---|---|
| SNOMED CT | 762561000 | Occupational allergic contact dermatitis caused by cutting oil |
| SNOMED CT | 762562007 | Occupational allergic contact dermatitis caused by disinfectant |
| SNOMED CT | 762566005 | Occupational allergic contact dermatitis caused by metal compound |
| SNOMED CT | 762570002 | Occupational allergic contact dermatitis caused by industrial preservative |
| SNOMED CT | 762571003 | Occupational allergic contact dermatitis caused by biocide |
| SNOMED CT | 762574006 | Occupational allergic contact dermatitis caused by rubber compound |
| SNOMED CT | 762576008 | Occupational allergic contact dermatitis caused by systemic drug or medicament |
| SNOMED CT | 762578009 | Occupational allergic contact dermatitis caused by topical medicament |
| SNOMED CT | 762618008 | Bronchiolitis obliterans syndrome due to and after lung transplantation |
| SNOMED CT | 762726008 | Occupational allergic contact dermatitis caused by clothing |
| SNOMED CT | 762754001 | Allergic contact dermatitis caused by fragrance |
| SNOMED CT | 762755000 | Allergic contact dermatitis caused by footwear |
| SNOMED CT | 762757008 | Allergic contact dermatitis caused by clothing |
| SNOMED CT | 763630007 | Satoyoshi syndrome |
| SNOMED CT | 763793004 | Limbic encephalitis with contactin-associated protein-like 2 antibodies |
| SNOMED CT | 763794005 | Limbic encephalitis with leucine-rich glioma-inactivated 1 antibodies |
| SNOMED CT | 763864008 | Persistent polyclonal B-cell lymphocytosis |
| SNOMED CT | 765106006 | Early-onset neonatal sepsis |
| SNOMED CT | 765107002 | Late-onset neonatal sepsis |
| SNOMED CT | 765751002 | Autoimmune encephalopathy with parasomnia and obstructive sleep apnea |
| SNOMED CT | 766049000 | Acute sensory ataxic neuropathy |
| SNOMED CT | 766246000 | Marburg acute multiple sclerosis |
| SNOMED CT | 766252004 | Juvenile overlap myositis |
| SNOMED CT | 766722008 | Paraparetic variant of Guillain-Barré syndrome |
| SNOMED CT | 770349000 | Sepsis caused by virus |
| SNOMED CT | 770545005 | Allergic contact dermatitis caused by iodine |
| SNOMED CT | 770588002 | Allergic contact dermatitis caused by iodophore |
| SNOMED CT | 770589005 | Allergic contact dermatitis caused by povidone iodine |
| SNOMED CT | 771145006 | Herpetiform pemphigus |
| SNOMED CT | 771271000 | Steroid-responsive encephalopathy associated with autoimmune thyroiditis |
| SNOMED CT | 773275000 | Post-transplant acute limbic encephalitis |

Continued on next page

**Table 7 – continued from previous page**

| CodeSystem | ConceptCode | ConceptName |
|---|---|---|
| SNOMED CT | 773662009 | Neonatal inflammatory skin and bowel disease |
| SNOMED CT | 774080007 | Neonatal scleroderma |
| SNOMED CT | 774082004 | Neonatal dermatomyositis |
| SNOMED CT | 774083009 | Neonatal autoimmune hemolytic anemia |
| SNOMED CT | 774084003 | Neonatal antiphospholipid syndrome |
| SNOMED CT | 778050009 | Idiopathic eosinophilic myositis |
| SNOMED CT | 781474001 | Allergic disorder |
| SNOMED CT | 782309008 | Immune-mediated neuropathy |
| SNOMED CT | 782513000 | Acute severe exacerbation of allergic asthma |
| SNOMED CT | 782520007 | Exacerbation of allergic asthma due to infection |
| SNOMED CT | 782866008 | Eosinophilic peritonitis |
| SNOMED CT | 783010003 | Pharyngeal-cervical-brachial variant of Guillain-Barré syndrome |
| SNOMED CT | 783187005 | Aseptic peritoneal eosinophilia after dialysis |
| SNOMED CT | 783205005 | Alopecia antibody deficiency |
| SNOMED CT | 783211008 | Non-chronic lymphocytic leukemia monoclonal B-cell lymphocytosis |
| SNOMED CT | 783244002 | Acute pure sensory neuropathy |
| SNOMED CT | 783566008 | Mucocutaneous ulcer caused by Epstein-Barr virus |
| SNOMED CT | 784327005 | Pustular psoriasis of sole of foot |
| SNOMED CT | 784328000 | Pustular psoriasis of palm of hand |
| SNOMED CT | 786855003 | Monoclonal B-cell lymphocytosis chronic lymphocytic leukemia-type |
| SNOMED CT | 788613004 | Anti-glomerular basement membrane disease |
| SNOMED CT | 788718000 | Iritis co-occurrent with inflammatory bowel disease |
| SNOMED CT | 788803005 | Allergic reaction to galactose-alpha 1,3 galactose |
| SNOMED CT | 789043007 | Sepsis without septic shock |
| SNOMED CT | 789296007 | Phlyctenular keratoconjunctivitis caused by staphylococcus toxin |
| SNOMED CT | 789697006 | Mast cell gastritis |
| SNOMED CT | 792842004 | Coronary arteriosclerosis in artery of transplanted heart |
| SNOMED CT | 816984002 | Progressive multiple sclerosis |
| SNOMED CT | 818950005 | Blau syndrome |
| SNOMED CT | 829971006 | Non-amyloid monoclonal immunoglobulin deposition disease |
| SNOMED CT | 829976001 | Thunderstorm asthma |
| SNOMED CT | 838324008 | Autoimmune optic neuropathy |
| SNOMED CT | 838330008 | Autoimmune ganglionopathy |
| SNOMED CT | 838359008 | Acute cutaneous lupus erythematosus |
| SNOMED CT | 838366009 | Anti-synthetase syndrome due to dermatomyositis |
| SNOMED CT | 838367000 | Allergic rhinosinusitis caused by Aspergillus |
| SNOMED CT | 838551007 | Autoimmune progesterone dermatitis |

Continued on next page

**Table 7 – continued from previous page**

| CodeSystem | ConceptCode | ConceptName |
|---|---|---|
| SNOMED CT | 840505007 | Down syndrome co-occurrent with leukemoid reaction associated transient neonatal pustulosis |
| SNOMED CT | 846606007 | Graft versus host disease of liver |
| SNOMED CT | 860800001 | Erythema multiforme due to systemic fungal infection |
| SNOMED CT | 860801002 | Erythema nodosum due to systemic fungal infection |
| SNOMED CT | 860824009 | Eosinophilia due to infectious disease |
| SNOMED CT | 860825005 | Hypertrophic pachymeningitis due to disorder of immune function |
| SNOMED CT | 860827002 | Eosinopenia due to infectious disease |
| SNOMED CT | 860869003 | Erythema multiforme due to Herpes simplex infection |
| SNOMED CT | 860870002 | Erythema multiforme due to Mycoplasma pneumoniae infection |
| SNOMED CT | 860924005 | Allergic contact dermatitis caused by alkali-metal soap |
| SNOMED CT | 866046008 | Mucous membrane pemphigoid of penile mucosa |
| SNOMED CT | 866084002 | Lymphopenia due to infection |
| SNOMED CT | 866151004 | Lymphocytopenia due to Severe acute respiratory syndrome coronavirus 2 |
| SNOMED CT | 870588003 | Sepsis due to disease caused by severe acute respiratory syndrome coronavirus 2 |
| SNOMED CT | 870749003 | Allergic contact dermatitis due to usnic acid |
| SNOMED CT | 871920005 | Allergic reaction to insect bite |
| SNOMED CT | 871926004 | Anaphylaxis caused by insect bite and/or insect sting |
| SNOMED CT | 871927008 | Allergic reaction caused by insect bite and/or insect sting |
| SNOMED CT | 871930001 | Anaphylaxis after allergen immunotherapy |
| SNOMED CT | 871931002 | Allergic reaction after allergen immunotherapy |
| SNOMED CT | 2701000124105 | Recurrent acute disseminated encephalomyelitis |
| SNOMED CT | 8161000119106 | Arthritis co-occurrent and due to Crohn's disease |
| SNOMED CT | 22241000175107 | Sepsis due to methicillin-sensitive Staphylococcus aureus |
| SNOMED CT | 32751000119100 | Line sepsis associated with dialysis catheter |
| SNOMED CT | 61311000119104 | Acute myeloid leukemia, minimal differentiation, FAB M0 in remission |
| SNOMED CT | 61321000119106 | Acute myeloid leukemia without maturation, FAB M1 in remission |
| SNOMED CT | 68261000119102 | Diffuse non-Hodgkin's lymphoma Lugano stage I |
| SNOMED CT | 68271000119108 | Diffuse non-Hodgkin's lymphoma Lugano stage II |
| SNOMED CT | 68291000119109 | Diffuse non-Hodgkin's lymphoma Lugano stage IV |
| SNOMED CT | 72161000119100 | Antiphospholipid syndrome in pregnancy |
| SNOMED CT | 72631000119101 | Human immunodeficiency virus (HIV) II infection category B2 |
| SNOMED CT | 76981000119106 | Human immunodeficiency virus (HIV) infection category B1 |
| SNOMED CT | 76991000119109 | Human immunodeficiency virus (HIV) infection category B2 |
| SNOMED CT | 84811000119107 | Follicular dendritic sarcoma of intraabdominal lymph node |
| SNOMED CT | 84831000119102 | Follicular dendritic sarcoma of lymph nodes of multiple sites |

Continued on next page

**Table 7 – continued from previous page**

| CodeSystem | ConceptCode | ConceptName |
|---|---|---|
| SNOMED CT | 89681000119101 | Glomerulonephritis co-occurrent and due to scleroderma |
| SNOMED CT | 99031000119107 | Acute exacerbation of asthma co-occurrent with allergic rhinitis |
| SNOMED CT | 103781000119103 | Allergic bronchopulmonary mycosis |
| SNOMED CT | 116711000119103 | Marginal zone lymphoma of inguinal lymph node |
| SNOMED CT | 116741000119104 | Marginal zone lymphoma of thoracic lymph node |
| SNOMED CT | 117061000119101 | Peripheral T-cell lymphoma of spleen |
| SNOMED CT | 117091000119108 | Peripheral T-cell lymphoma of axillary lymph node |
| SNOMED CT | 117111000119100 | Peripheral T-cell lymphoma of thoracic lymph node |
| SNOMED CT | 119031000119101 | Myelofibrosis due to another disorder |
| SNOMED CT | 121121000119106 | Acute graft-versus-host reaction following bone marrow transplant |
| SNOMED CT | 121881000119102 | Intractable allergic migraine |
| SNOMED CT | 122501000119101 | Lymphoproliferative disorder following lung transplant |
| SNOMED CT | 122511000119103 | Lymphoproliferative disorder following heart transplantation |
| SNOMED CT | 122521000119105 | Lymphoproliferative disorder following liver transplant |
| SNOMED CT | 122531000119108 | Lymphoproliferative disorder following kidney transplant |
| SNOMED CT | 122881000119107 | Chronic monocytic leukemia in relapse |
| SNOMED CT | 122901000119109 | Myeloid leukemia in relapse |
| SNOMED CT | 122951000119108 | Lymphoid leukemia in relapse |
| SNOMED CT | 122961000119105 | Chronic lymphoid leukemia in relapse |
| SNOMED CT | 122981000119101 | Plasma cell leukemia in relapse |
| SNOMED CT | 123571000119104 | Erythema multiforme minor |
| SNOMED CT | 123781000119107 | Chronic leukemia in relapse |
| SNOMED CT | 124991000119109 | Severe persistent asthma co-occurrent with allergic rhinitis |
| SNOMED CT | 125001000119103 | Moderate persistent asthma co-occurrent with allergic rhinitis |
| SNOMED CT | 125011000119100 | Mild persistent asthma co-occurrent with allergic rhinitis |
| SNOMED CT | 125021000119107 | Intermittent asthma co-occurrent with allergic rhinitis |
| SNOMED CT | 126811000119102 | Sepsis without acute organ dysfunction caused by Serratia species |
| SNOMED CT | 127121000119101 | Sepsis without acute organ dysfunction caused by Streptococcus pneumoniae |
| SNOMED CT | 127141000119107 | Septic shock co-occurrent with acute organ dysfunction due to anaerobic bacteria |
| SNOMED CT | 127171000119100 | Septic shock co-occurrent with acute organ dysfunction due to Haemophilus influenzae |
| SNOMED CT | 127241000119104 | Septic shock co-occurrent with acute organ dysfunction due to Staphylococcus |
| SNOMED CT | 127751000119108 | Sepsis caused by Staphylococcus without acute organ dysfunction |
| SNOMED CT | 127961000119108 | Aleukemic myeloid leukemia in relapse |

Continued on next page

**Table 7 – continued from previous page**

| CodeSystem | ConceptCode | ConceptName |
|---|---|---|
| SNOMED CT | 128631000119109 | Chronic graft-versus-host disease following kidney transplant |
| SNOMED CT | 128971000119101 | Cryoglobulinemia due to chronic hepatitis C |
| SNOMED CT | 129571000119102 | Allergic reaction due to allergy skin test |
| SNOMED CT | 129781000119105 | Neoplasia following lung transplant |
| SNOMED CT | 137421000119106 | Graves' disease in remission |
| SNOMED CT | 143441000119108 | Rheumatoid arthritis in remission |
| SNOMED CT | 145611000119107 | Allergic migraine |
| SNOMED CT | 156051000119109 | Allergic rhinitis due to animal hair and dander |
| SNOMED CT | 182961000119101 | Acute disseminated encephalomyelitis following infectious disease |
| SNOMED CT | 292991000119106 | Eaton Lambert syndrome without underlying malignancy |
| SNOMED CT | 300961000119108 | Juvenile idiopathic arthritis of left hand |
| SNOMED CT | 301051000119101 | Juvenile idiopathic arthritis of right hand |
| SNOMED CT | 318871000119107 | Seropositive rheumatoid arthritis of left hip |
| SNOMED CT | 318881000119105 | Seropositive rheumatoid arthritis of left knee |
| SNOMED CT | 318941000119109 | Seropositive rheumatoid arthritis of right hip |
| SNOMED CT | 318951000119106 | Seropositive rheumatoid arthritis of right knee |
| SNOMED CT | 319031000119108 | Seronegative rheumatoid arthritis of left hand |
| SNOMED CT | 319111000119104 | Seronegative rheumatoid arthritis of right hand |
| SNOMED CT | 319861000119106 | Rheumatoid nodule of subcutaneous tissue of left elbow |
| SNOMED CT | 319871000119100 | Rheumatoid nodule of subcutaneous tissue of left hand |
| SNOMED CT | 319941000119103 | Rheumatoid nodule of subcutaneous tissue of right elbow |
| SNOMED CT | 319951000119101 | Rheumatoid nodule of subcutaneous tissue of right hand |
| SNOMED CT | 333131000119102 | Phlyctenular keratoconjunctivitis of right eye |
| SNOMED CT | 335981000119102 | Chronic giant papillary conjunctivitis of right eye |
| SNOMED CT | 336601000119106 | Acute atopic conjunctivitis of right eye |
| SNOMED CT | 338741000119102 | Phlyctenular keratoconjunctivitis of left eye |
| SNOMED CT | 341591000119109 | Chronic giant papillary conjunctivitis of left eye |
| SNOMED CT | 342211000119106 | Acute atopic conjunctivitis of left eye |
| SNOMED CT | 342761000119104 | Allergic contact dermatitis of right upper eyelid |
| SNOMED CT | 342771000119105 | Allergic contact dermatitis of right lower eyelid |
| SNOMED CT | 342781000119108 | Allergic contact dermatitis of right eyelid |
| SNOMED CT | 342801000119107 | Allergic contact dermatitis of left lower eyelid |
| SNOMED CT | 342811000119105 | Allergic contact dermatitis of left eyelid |
| SNOMED CT | 344511000119102 | Bilateral phlyctenular keratoconjunctivitis |
| SNOMED CT | 346811000119109 | Bilateral chronic giant papillary conjunctivitis of eyes |
| SNOMED CT | 347371000119108 | Acute atopic conjunctivitis of bilateral eyes |
| SNOMED CT | 428241000124101 | Anaphylaxis caused by human papillomavirus vaccine |
| SNOMED CT | 428281000124107 | Anaphylactic reaction caused by diphtheria and tetanus vaccine |
| SNOMED CT | 428291000124105 | Anaphylaxis caused by tetanus, diphtheria and acellular pertussis vaccine |

Continued on next page

**Table 7 – continued from previous page**

| CodeSystem | ConceptCode | ConceptName |
|---|---|---|
| SNOMED CT | 428301000124106 | Anaphylaxis caused by meningococcal vaccine |
| SNOMED CT | 428321000124101 | Anaphylaxis due to Hepatitis B vaccine |
| SNOMED CT | 432451000124103 | Multiphasic acute disseminated encephalomyelitis |
| SNOMED CT | 433621000124101 | Anaphylaxis due to Hemophilus influenzae type b vaccine |
| SNOMED CT | 435181000124108 | Sepsis of newborn due to Streptococcus agalactiae |
| SNOMED CT | 436741000124105 | septicaemia associated with vascular access catheter |
| SNOMED CT | 436751000124107 | Sepsis associated with internal vascular access |
| SNOMED CT | 450521000124109 | Marginal zone lymphoma of axillary lymph node |
| SNOMED CT | 451011000124102 | Allergic conjunctivitis of left eye |
| SNOMED CT | 452281000124106 | Autoimmune encephalitis caused by N-methyl D-aspartate receptor antibody |
| SNOMED CT | 452291000124109 | Relapse multiple myeloma |
| SNOMED CT | 456911000124108 | Diffuse non-Hodgkin's lymphoma Lugano stage III |
| SNOMED CT | 459911000124100 | Rheumatoid factor positive rheumatoid arthritis |
| SNOMED CT | 459921000124108 | Rheumatoid factor positive rheumatoid arthritis of the shoulder |
| SNOMED CT | 461311000124107 | Refractory Hodgkin's lymphoma |
| SNOMED CT | 461351000124108 | Breast implant-associated anaplastic large cell lymphoma |
| SNOMED CT | 679351000119102 | Allergic conjunctivitis of bilateral eyes |
| SNOMED CT | 679371000119106 | Allergic conjunctivitis of right eye |
| SNOMED CT | 1073681000119109 | Rheumatoid arthritis of left ankle |
| SNOMED CT | 1073691000119107 | Rheumatoid arthritis of left elbow |
| SNOMED CT | 1073711000119105 | Rheumatoid arthritis of left hand |
| SNOMED CT | 1073721000119103 | Rheumatoid arthritis of left hip |
| SNOMED CT | 1073741000119109 | Rheumatoid arthritis of left shoulder |
| SNOMED CT | 1073751000119106 | Rheumatoid arthritis of left wrist |
| SNOMED CT | 1073761000119108 | Rheumatoid arthritis of right ankle |
| SNOMED CT | 1073771000119102 | Rheumatoid arthritis of right elbow |
| SNOMED CT | 1073791000119101 | Rheumatoid arthritis of right hand |
| SNOMED CT | 1073811000119102 | Rheumatoid arthritis of right knee |
| SNOMED CT | 1073821000119109 | Rheumatoid arthritis of right shoulder |
| SNOMED CT | 1073831000119107 | Rheumatoid arthritis of right wrist |
| SNOMED CT | 1073861000119104 | Deformity of right hand co-occurrent and due to rheumatoid arthritis |
| SNOMED CT | 1073871000119105 | Deformity of left hand co-occurrent and due to rheumatoid arthritis |
| SNOMED CT | 1073881000119108 | Deformity of right foot co-occurrent and due to rheumatoid arthritis |
| SNOMED CT | 1073891000119106 | Deformity of left foot co-occurrent and due to rheumatoid arthritis |
| SNOMED CT | 1083041000119104 | Acute recurrent allergic otitis media of bilateral middle ears |
| SNOMED CT | 1083081000119109 | Acute allergic serous otitis media of bilateral middle ears |
| SNOMED CT | 1083191000119108 | Autoimmune disorder of bilateral inner ears |

Continued on next page

**Table 7 – continued from previous page**

| CodeSystem | ConceptCode | ConceptName |
|---|---|---|
| SNOMED CT | 1085131000119105 | Abscess of intestine co-occurrent and due to chronic ulcerative pancolitis |
| SNOMED CT | 1085231000119100 | Abscess of intestine co-occurrent and due to chronic ulcerative rectosigmoiditis |
| SNOMED CT | 1085751000119100 | Abscess of intestine co-occurrent and due to Crohn's disease of large intestine |
| SNOMED CT | 1085801000119106 | Abscess of intestine co-occurrent and due to Crohn's disease of small and large intestine |
| SNOMED CT | 1092461000119104 | Sepsis without acute organ dysfunction |
| SNOMED CT | 1092471000119105 | Septic shock co-occurrent with acute organ dysfunction due to Chromobacterium |
| SNOMED CT | 1092481000119108 | Septic shock co-occurrent with acute organ dysfunction due to coagulase-negative Staphylococcus |
| SNOMED CT | 1092491000119106 | Septic shock co-occurrent with acute organ dysfunction due to Enterococcus |
| SNOMED CT | 1092501000119104 | Septic shock co-occurrent with acute organ dysfunction due to Gonococcus |
| SNOMED CT | 1092511000119101 | Septic shock co-occurrent with acute organ dysfunction due to Gram-positive coccus |
| SNOMED CT | 1092531000119106 | Septic shock co-occurrent with acute organ dysfunction due to Group A streptococcus |
| SNOMED CT | 1092541000119102 | Septic shock co-occurrent with acute organ dysfunction due to Group B streptococcus |
| SNOMED CT | 1092571000119109 | Sepsis caused by Chromobacterium |
| SNOMED CT | 1092591000119105 | Sepsis caused by Enterococcus |
| SNOMED CT | 1092601000119103 | Sepsis caused by Gonococcus |
| SNOMED CT | 1092841000119100 | Abscess of intestine co-occurrent and due to ulcerative colitis |
| SNOMED CT | 10628671000119100 | Allergic contact dermatitis caused by latex |
| SNOMED CT | 10628711000119101 | Allergic contact dermatitis caused by plant material |
| SNOMED CT | 10628751000119100 | Allergic contact dermatitis caused by pollen |
| SNOMED CT | 10629311000119107 | Psoriatic arthritis mutilans |
| SNOMED CT | 10629391000119103 | Allergic rhinitis caused by feathers |
| SNOMED CT | 10629431000119108 | Allergic rhinitis caused by insects |
| SNOMED CT | 10629471000119106 | Allergic rhinitis caused by mold |
| SNOMED CT | 10674711000119105 | Acute severe exacerbation of asthma co-occurrent with allergic rhinitis |
| SNOMED CT | 10674991000119104 | Intermittent allergic asthma |
| SNOMED CT | 10675431000119106 | Severe persistent allergic asthma |
| SNOMED CT | 10675471000119109 | Acute severe exacerbation of severe persistent allergic asthma |
| SNOMED CT | 10675551000119104 | Acute severe exacerbation of severe persistent asthma co-occurrent with allergic rhinitis |

Continued on next page

**Table 7 – continued from previous page**

| CodeSystem | ConceptCode | ConceptName |
|---|---|---|
| SNOMED CT | 10675631000119109 | Severe persistent asthma controlled co-occurrent with allergic rhinitis |
| SNOMED CT | 10675711000119106 | Severe persistent asthma uncontrolled co-occurrent with allergic rhinitis |
| SNOMED CT | 10675871000119106 | Mild persistent allergic asthma |
| SNOMED CT | 10675911000119109 | Acute severe exacerbation of mild persistent allergic asthma |
| SNOMED CT | 10675991000119100 | Acute severe exacerbation of mild persistent allergic asthma co-occurrent with allergic rhinitis |
| SNOMED CT | 10676111000119102 | Mild persistent asthma controlled co-occurrent with allergic rhinitis |
| SNOMED CT | 10676231000119102 | Mild persistent asthma uncontrolled co-occurrent with allergic rhinitis |
| SNOMED CT | 10676391000119108 | Moderate persistent allergic asthma |
| SNOMED CT | 10676431000119103 | Acute severe exacerbation of moderate persistent allergic asthma |
| SNOMED CT | 10676511000119109 | Acute severe exacerbation of moderate persistent asthma co-occurrent with allergic rhinitis |
| SNOMED CT | 10676591000119100 | Moderate persistent controlled asthma co-occurrent with allergic rhinitis |
| SNOMED CT | 10676671000119102 | Moderate persistent asthma uncontrolled co-occurrent with allergic rhinitis |
| SNOMED CT | 10676871000119103 | Allergic contact dermatitis caused by chemical |
| SNOMED CT | 10743231000119101 | Crohn's disease of small intestine with stenosis |
| SNOMED CT | 10743271000119103 | Immunoglobulin G4 related systemic disease |
| SNOMED CT | 10746341000119109 | Acquired immune deficiency syndrome complicating childbirth |
| SNOMED CT | 10751351000119104 | Sepsis due to ectopic pregnancy |
| SNOMED CT | 10812001000119100 | Sepsis due to incomplete miscarriage |
| SNOMED CT | 10838971000119103 | Immunodeficiency caused by long term therapeutic use of drug |
| SNOMED CT | 11055151000119108 | Seropositive rheumatoid arthritis of multiple joints |
| SNOMED CT | 13008751000119103 | Allergic proctocolitis caused by food protein |
| SNOMED CT | 15673361000119100 | Bilateral rheumatoid nodule of subcutaneous tissue of elbows |
| SNOMED CT | 15673521000119101 | Bilateral rheumatoid nodule of subcutaneous tissue of hands |
| SNOMED CT | 15679961000119104 | Vernal conjunctivitis of right eye |
| SNOMED CT | 15680001000119101 | Vernal conjunctivitis of left eye |
| SNOMED CT | 15680041000119104 | Vernal conjunctivitis of bilateral eyes |
| SNOMED CT | 15685921000119102 | Rheumatoid arthritis of bilateral temporomandibular joints |
| SNOMED CT | 15685961000119107 | Rheumatoid arthritis of right temporomandibular joint |
| SNOMED CT | 15686001000119104 | Rheumatoid arthritis of left temporomandibular joint |
| SNOMED CT | 15686281000119101 | Bilateral rheumatoid arthritis of feet |
| SNOMED CT | 15686321000119106 | Rheumatoid arthritis of bilateral elbows |

Continued on next page

**Table 7 – continued from previous page**

| CodeSystem | ConceptCode | ConceptName |
|---|---|---|
| SNOMED CT | 15687201000119107 | Bilateral rheumatoid arthritis of knees |
| SNOMED CT | 15687321000119109 | Bilateral rheumatoid arthritis of hands |
| SNOMED CT | 15687841000119108 | Rheumatoid arthritis of bilateral hips |
| SNOMED CT | 15691161000119108 | Seronegative rheumatoid arthritis of bilateral hands |
| SNOMED CT | 15691721000119102 | Rheumatoid arthritis of bilateral ankles |
| SNOMED CT | 15691761000119107 | Rheumatoid arthritis of bilateral wrists |
| SNOMED CT | 15691801000119104 | Seropositive rheumatoid arthritis of bilateral hands |
| SNOMED CT | 15911801000119104 | Allergic reaction caused by penicillin |
| SNOMED CT | 15919441000119101 | Allergic reaction caused by hymenoptera venom |
| SNOMED CT | 15919781000119104 | Allergic reaction caused by antibacterial agent |
| SNOMED CT | 15919821000119109 | Allergic reaction caused by sulfonamide |
| SNOMED CT | 15920041000119105 | Allergic reaction caused by nonsteroidal antiinflammatory agent |
| SNOMED CT | 15920121000119103 | Allergic reaction caused by vaccine product |
| SNOMED CT | 15920161000119108 | Allergic reaction caused by egg protein |
| SNOMED CT | 15920201000119103 | Allergic reaction caused by fish |
| SNOMED CT | 15920361000119106 | Allergic reaction caused by peanut |
| SNOMED CT | 15920401000119102 | Allergic reaction caused by seafood |
| SNOMED CT | 15920481000119105 | Allergic reaction caused by shellfish |
| SNOMED CT | 15920521000119105 | Allergic reaction caused by tree nut |
| SNOMED CT | 15920601000119100 | Allergic reaction caused by latex |
| SNOMED CT | 15928141000119107 | Human immunodeficiency virus infection with cognitive impairment |
| SNOMED CT | 15988671000119100 | Perennial allergic conjunctivitis of bilateral eyes |
| SNOMED CT | 16024431000119108 | Acute polyarticular juvenile rheumatoid arthritis |
| SNOMED CT | 16041671000119109 | Juvenile rheumatoid arthritis of bilateral knees |
| SNOMED CT | 16044751000119106 | Polyarticular juvenile idiopathic arthritis |
| SNOMED CT | 16058671000119103 | Transplant rejection of cornea of left eye |
| SNOMED CT | 16058711000119104 | Transplant failure of cornea of right eye |
| SNOMED CT | 16058831000119102 | Transplant rejection of cornea of right eye |
| SNOMED CT | 16058871000119104 | Transplant failure of cornea of left eye |
| SNOMED CT | 16060271000119109 | Neonatal sepsis caused by Staphylococcus |
| SNOMED CT | 16060311000119109 | Neonatal sepsis caused by Streptococcus |
| SNOMED CT | 16098491000119109 | Chronic autoimmune hepatitis |
| SNOMED CT | 16208561000119106 | Juvenile rheumatoid arthritis of left knee |
| SNOMED CT | 16208721000119109 | Juvenile rheumatoid arthritis of right knee |
| SNOMED CT | 16228891000119105 | Allergic contact dermatitis due to metal |
| SNOMED CT | 16318001000119107 | Hypogammaglobulinemia due to multiple myeloma |
| SNOMED CT | 16697871000119100 | Mast cell activation syndrome |
| SNOMED CT | 674814021000119106 | Acute respiratory distress syndrome due to COVID-19 |
| SNOMED CT | 479003 | Graves' disease with pretibial myxoedema AND with thyrotoxic crisis |
| SNOMED CT | 1089000 | Intrauterine foetal sepsis |

Continued on next page

**Table 7 – continued from previous page**

| CodeSystem | ConceptCode | ConceptName |
|---|---|---|
| SNOMED CT | 3815005 | Regional enteritis of rectum |
| SNOMED CT | 5603006 | Autoimmune hemolytic anemia caused by immunoglobulin G |
| SNOMED CT | 7620006 | Regional enteritis of the large bowel |
| SNOMED CT | 18700001 | Septicemia caused by Chromobacterium |
| SNOMED CT | 19719003 | Jessner's lymphocytic infiltrate |
| SNOMED CT | 21719001 | Hayfever |
| SNOMED CT | 27520001 | Pustular psoriasis of the palms AND/OR soles |
| SNOMED CT | 27538003 | Hashitoxicosis |
| SNOMED CT | 31848007 | CREST - Calcinosis, Raynaud's phenomenon, esophageal dysfunction, sclerodactyly, telangiectasia |
| SNOMED CT | 33491002 | Autoimmune hemolytic anemia caused by IgA plus complement |
| SNOMED CT | 34000006 | Crohns disease |
| SNOMED CT | 38106008 | Crohn disease of ileum |
| SNOMED CT | 40175007 | Allergic dermatitis caused by bite of Ctenocephalides felis |
| SNOMED CT | 40956001 | Guillain Barre syndrome |
| SNOMED CT | 41936006 | Septicemia caused by Erysipelothrix insidiosa |
| SNOMED CT | 48347002 | Air-conditioner and humidifier lung |
| SNOMED CT | 50440006 | Regional colitis |
| SNOMED CT | 50581000 | Pulmonary-renal syndrome |
| SNOMED CT | 51696001 | Subcutaneous lipogranulomatosis |
| SNOMED CT | 52983002 | Kerion caused by Microsporum |
| SNOMED CT | 56287005 | Regional enteritis of the duodenum |
| SNOMED CT | 56689002 | Granulomatous enteritis |
| SNOMED CT | 56989000 | Myasthenic syndrome |
| SNOMED CT | 61424003 | Crohn disease of pylorus |
| SNOMED CT | 62382002 | Calcinosis, Raynaud's phenomenon, sclerodactyly, and telangiectasia syndrome |
| SNOMED CT | 62609001 | Autoimmune hemolytic anemia caused by IgG plus complement |
| SNOMED CT | 69339004 | Pigeon fancier's lung |
| SNOMED CT | 70622003 | Crohn disease of pyloric antrum |
| SNOMED CT | 71832003 | Autoimmune hemolytic anemia caused by immunoglobulin M |
| SNOMED CT | 71833008 | Crohn's disease of small and large intestines |
| SNOMED CT | 73860009 | Kerion caused by Trichophyton |
| SNOMED CT | 76715008 | Autoimmune adrenal atrophy |
| SNOMED CT | 78946008 | Keratoconjunctivitis sicca, in Sjögren syndrome |
| SNOMED CT | 79337003 | Blood product-induced anaphylaxis |
| SNOMED CT | 79587009 | Septicemia caused by Bacteroides |
| SNOMED CT | 83901003 | Sicca syndrome |
| SNOMED CT | 85559002 | Pelger Huet anomaly |

Continued on next page

**Table 7 – continued from previous page**

| CodeSystem | ConceptCode | ConceptName |
|---|---|---|
| SNOMED CT | 87442008 | Pericarditis co-occurrent and due to scleroderma |
| SNOMED CT | 91178003 | Allergic dermatitis caused by bite of Ctenocephalides canis |
| SNOMED CT | 91390005 | Crohn disease of jejunum |
| SNOMED CT | 91411007 | Autoimmune hemolytic anemia caused by IgA |
| SNOMED CT | 95330001 | Linear immunoglobulin A dermatosis |
| SNOMED CT | 123583002 | Tuberculids |
| SNOMED CT | 126766000 | Lymphoepithelial sialadenitis of Sjögren syndrome |
| SNOMED CT | 127054006 | Cold agglutinin disease caused by Epstein-Barr virus infection |
| SNOMED CT | 129000002 | Langerhans cell histiocytosis, unifocal |
| SNOMED CT | 129645002 | Adult glucose-6-phosphate dehydrogenase deficiency of leukocytes syndrome |
| SNOMED CT | 193497004 | Vogt-Koyanagi syndrome |
| SNOMED CT | 196137000 | Lung disease with Sjögren disease |
| SNOMED CT | 196977009 | Crohn disease of terminal ileum |
| SNOMED CT | 206378006 | Sepsis of newborn caused by Staphylococcus aureus |
| SNOMED CT | 206379003 | Sepsis of newborn caused by Escherichia coli |
| SNOMED CT | 234558001 | Anti-hemophilus influenzae B polysaccharide antibody deficiency |
| SNOMED CT | 234619000 | Hereditary angioneurotic edema - type 1 |
| SNOMED CT | 234620006 | Hereditary angioneurotic edema - type 2 |
| SNOMED CT | 235664007 | Crohn disease of stomach |
| SNOMED CT | 235796008 | Perianal Crohn disease |
| SNOMED CT | 237520009 | Hypothyroidism due to Hashimoto thyroiditis |
| SNOMED CT | 237521008 | Hypothyroidism due to thyroid stimulating hormone receptor blocking antibody |
| SNOMED CT | 238612002 | Generalized pustular psoriasis of von Zumbusch |
| SNOMED CT | 238937005 | Pemphigus vegetans (Neumann type) |
| SNOMED CT | 239912009 | Primary Sjögren syndrome |
| SNOMED CT | 239915006 | Secondary Sjögren syndrome |
| SNOMED CT | 285423008 | Immunoglobulin A monoclonal gammopathy of uncertain significance |
| SNOMED CT | 285424002 | Immunoglobulin G monoclonal gammopathy of uncertain significance |
| SNOMED CT | 285426000 | Immunoglobulin M monoclonal gammopathy of uncertain significance |
| SNOMED CT | 285428004 | Immunoglobulin D monoclonal gammopathy of uncertain significance |
| SNOMED CT | 353295004 | Exophthalmic goiter |
| SNOMED CT | 397172008 | Gastrointestinal Crohn disease |
| SNOMED CT | 402378006 | Metastatic Crohn disease of skin |
| SNOMED CT | 402446002 | Oral mucosal involvement by linear immunoglobulin A disease |

Continued on next page

**Table 7 – continued from previous page**

| CodeSystem | ConceptCode | ConceptName |
|---|---|---|
| SNOMED CT | 403158001 | Allergic contact dermatitis caused by Bryozoa |
| SNOMED CT | 403633007 | Drug-induced linear immunoglobulin A disease |
| SNOMED CT | 410485009 | Iritis with Crohn disease |
| SNOMED CT | 414153008 | Exacerbation of Crohn disease of large intestine |
| SNOMED CT | 414154002 | Exacerbation of Crohn disease of small intestine |
| SNOMED CT | 421310001 | Non-immunoglobulin E-mediated atopic disorder |
| SNOMED CT | 421871004 | Atopic allergy |
| SNOMED CT | 422076005 | Immediate hypersensitivity disorder |
| SNOMED CT | 422169003 | Nonatopic immunoglobulin E-mediated allergic disorder |
| SNOMED CT | 422339003 | Non-immunoglobulin E-mediated allergic disorder |
| SNOMED CT | 423889005 | Non-immunoglobulin E mediated allergic asthma |
| SNOMED CT | 447684006 | Erysipelothrix sepsis |
| SNOMED CT | 447685007 | Severe sepsis with acute organ dysfunction caused by Haemophilus influenzae |
| SNOMED CT | 447841007 | Sepsis caused by Candida |
| SNOMED CT | 447894003 | Severe sepsis with acute organ dysfunction caused by Staphylococcus |
| SNOMED CT | 447897005 | Sepsis caused by Acinetobacter |
| SNOMED CT | 447898000 | Sepsis caused by Actinomyces |
| SNOMED CT | 447899008 | Sepsis caused by Escherichia coli |
| SNOMED CT | 448417001 | Sepsis caused by Staphylococcus aureus |
| SNOMED CT | 448418006 | Severe sepsis with acute organ dysfunction caused by Streptococcus |
| SNOMED CT | 448419003 | Severe sepsis with acute organ dysfunction due to Group B streptococcus |
| SNOMED CT | 448420009 | Sepsis caused by Streptococcus group D |
| SNOMED CT | 448421008 | Severe sepsis with acute organ dysfunction caused by Pneumococcus |
| SNOMED CT | 448422001 | Sepsis caused by Streptococcus suis |
| SNOMED CT | 448482003 | Sepsis caused by Bacillus anthracis |
| SNOMED CT | 448483008 | Sepsis caused by Enterobacter |
| SNOMED CT | 448812000 | Severe sepsis with acute organ dysfunction caused by methicillin resistant Staphylococcus aureus |
| SNOMED CT | 448813005 | Severe sepsis with acute organ dysfunction caused by Pseudomonas |
| SNOMED CT | 449082003 | Severe sepsis with acute organ dysfunction caused by Gram-negative bacteria |
| SNOMED CT | 449083008 | Severe sepsis with acute organ dysfunction caused by Salmonella |
| SNOMED CT | 449084002 | Severe sepsis with acute organ dysfunction caused by Serratia |
| SNOMED CT | 449335002 | Listerial sepsis |

Continued on next page

**Table 7 – continued from previous page**

| CodeSystem | ConceptCode | ConceptName |
|---|---|---|
| SNOMED CT | 449504009 | Severe sepsis with acute organ dysfunction due to Group A streptococcus |
| SNOMED CT | 449505005 | Severe sepsis with acute organ dysfunction due to coagulase negative Staphylococcus |
| SNOMED CT | 449546000 | Sepsis caused by Haemophilus influenzae type B |
| SNOMED CT | 699861000 | Blau syndrome |
| SNOMED CT | 703903005 | Dermatophagoides farinae allergic rhinitis |
| SNOMED CT | 703937002 | Dermatophagoides pteronyssinus allergic rhinitis |
| SNOMED CT | 703951002 | Allergic conjunctivitis caused by Dermatophagoides farinae |
| SNOMED CT | 703952009 | Allergic conjunctivitis caused by Dermatophagoides pteronyssinus |
| SNOMED CT | 703953004 | Asthma caused by house dust mite, Dermatophagoides pteronyssinus |
| SNOMED CT | 703954005 | Asthma caused by house dust mite, Dermatophagoides farinae |
| SNOMED CT | 762385007 | Allergic contact dermatitis caused by allergenic hapten derived from plants or organic matter |
| SNOMED CT | 72621000119104 | Human immunodeficiency virus |
| SNOMED CT | 127091000119100 | Sepsis of newborn caused by Streptococcus agalactiae |
| SNOMED CT | 127181000119102 | Septic shock co-occurrent with acute organ dysfunction caused by Meningococcus |
| SNOMED CT | 127191000119104 | Septic shock co-occurrent with acute organ dysfunction caused by methicillin susceptible Staphylococcus aureus |
| SNOMED CT | 127201000119101 | Septic shock co-occurrent with acute organ dysfunction caused by Pseudomonas |
| SNOMED CT | 127231000119108 | Septic shock co-occurrent with acute organ dysfunction caused by Serratia |
| SNOMED CT | 127251000119102 | Septic shock co-occurrent with acute organ dysfunction caused by Streptococcus |
| SNOMED CT | 127261000119100 | Septic shock co-occurrent with acute organ dysfunction caused by Pneumococcus |
| SNOMED CT | 127311000119106 | Sepsis caused by Meningococcus |
| SNOMED CT | 127331000119101 | Sepsis caused by methicillin susceptible Staphylococcus aureus |
| SNOMED CT | 128381000119104 | Septic shock co-occurrent with acute organ dysfunction caused by methicillin resistant Staphylococcus aureus |
| SNOMED CT | 342791000119106 | Contact dermatitis, left upper eyelid |
| SNOMED CT | 428331000124103 | Anaphylaxis caused by rotavirus vaccine |
| SNOMED CT | 1073701000119107 | Rheumatoid arthritis of left foot |
| SNOMED CT | 1073781000119104 | Rheumatoid arthritis of right foot |
| SNOMED CT | 1085851000119105 | Abscess of small intestine due to Crohn's disease |
| SNOMED CT | 1085901000119101 | Abscess of intestine due to Crohn's disease |

Continued on next page

**Table 7 – continued from previous page**

| CodeSystem | ConceptCode | ConceptName |
|---|---|---|
| SNOMED CT | 1003756003 | Hevea brasiliensis latex protein-induced angioedema-urticaria |
| SNOMED CT | 1003757007 | Allergic reaction caused by Hevea brasiliensis latex protein |
| SNOMED CT | 1003758002 | Anaphylaxis due to Hevea brasiliensis latex protein |
| SNOMED CT | 1052325005 | Psoriasis exacerbated by human immunodeficiency virus infection |
| SNOMED CT | 1078231000 | Allergic contact dermatitis due to jewelry |
| SNOMED CT | 1082871000119108 | Bilateral acute allergic otitis media of middle ears |
| SNOMED CT | 1083261000119107 | Bilateral chronic allergic otitis media of middle ears |
| SNOMED CT | 1087981000119103 | Acute allergic otitis media of left middle ear |
| SNOMED CT | 1088321000119102 | Autoimmune disorder of left inner ear |
| SNOMED CT | 1090601000119102 | Acute allergic otitis media of right middle ear |
| SNOMED CT | 1090931000119104 | Autoimmune disorder of right inner ear |
| SNOMED CT | 15691241000119101 | Bilateral seronegative rheumatoid arthritis of knees |
| SNOMED CT | 15691321000119101 | Bilateral seronegative rheumatoid arthritis of wrists |
| SNOMED CT | 15691881000119107 | Bilateral seropositive rheumatoid arthritis of knees |
| SNOMED CT | 16050071000119108 | Bilateral seropositive rheumatoid arthritis of feet |
| SNOMED CT | 16318061000119108 | Hypogammaglobulinemia due to monoclonal gammopathy of undetermined significance |
| SNOMED CT | 16606721000119107 | Seropositive rheumatoid arthritis in remission |
| SNOMED CT | 16894671000119102 | Immunoglobulin G deficiency due to monoclonal gammopathy of undetermined significance |
| SNOMED CT | 16894711000119103 | Immunoglobulin G deficiency due to multiple myeloma |
| SNOMED CT | 453720571000119100 | Acute ulcerative rectosigmoiditis |
| SNOMED CT | 63541000119109 | Corneal epithelial defect following penetrating keratoplasty |
| SNOMED CT | 890143000 | Allergic contact dermatitis caused by poison primrose |
| SNOMED CT | 890154005 | Allergic contact dermatitis caused by urushiol from Eastern poison ivy |
| SNOMED CT | 890169006 | Allergic contact dermatitis caused by urushiol from poison sumac |
| SNOMED CT | 890170007 | Allergic contact dermatitis caused by ragweed |
| SNOMED CT | 890283004 | Allergic contact dermatitis caused by oil |
| SNOMED CT | 890284005 | Allergic contact dermatitis caused by grease |
| SNOMED CT | 890331009 | Allergic contact dermatitis caused by soap |
| SNOMED CT | 966011731000119103 | Acute ulcerative pancolitis |
| SNOMED CT | 969688801000119108 | Acute left-sided ulcerative colitis |
| SNOMED CT | 1073601000119101 | Right foot seropositive rheumatoid arthritis |
| SNOMED CT | 1073611000119103 | Left foot seropositive rheumatoid arthritis |
| SNOMED CT | 1142045004 | Primary human immunodeficiency virus infection |
| SNOMED CT | 1142055000 | HIV modified skin disease |
| SNOMED CT | 1142207008 | Transient neonatal epidermolysis bullosa acquisita |
| SNOMED CT | 1144355009 | Allergic contact stomatitis caused by denture |
| SNOMED CT | 1144805008 | Microscopic polyangiitis |

Continued on next page

**Table 7 – continued from previous page**

| CodeSystem | ConceptCode | ConceptName |
|---|---|---|
| SNOMED CT | 1144907001 | Late acute graft-versus-host disease |
| SNOMED CT | 1144908006 | Late chronic graft-versus-host disease |
| SNOMED CT | 1144923001 | Disorder of immune function due to systemic lupus erythematosus |
| SNOMED CT | 1144925008 | Diffuse systemic sclerosis of childhood |
| SNOMED CT | 1144929002 | Secondary agranulocytosis |
| SNOMED CT | 1144941009 | Chronic tubulointerstitial nephritis following renal transplantation |
| SNOMED CT | 1144963000 | Acute on chronic graft-versus-host disease |
| SNOMED CT | 1144974009 | Antisynthetase syndrome due to polymyositis |
| SNOMED CT | 1144980001 | Food-dependent exercise-induced anaphylaxis |
| SNOMED CT | 1148594002 | Chronic rheumatic arthritis |
| SNOMED CT | 1148595001 | Subacute rheumatic arthritis |
| SNOMED CT | 1148597009 | Rheumatic arthritis |
| SNOMED CT | 1148598004 | Rheumatic arthritis of temporomandibular joint |
| SNOMED CT | 1148696006 | Leukopenia caused by drug |
| SNOMED CT | 1148763004 | Acute rheumatic carditis |
| SNOMED CT | 1148765006 | Chronic rheumatic carditis |
| SNOMED CT | 1148845007 | Follicular lymphoma grade 3b |
| SNOMED CT | 1148847004 | Follicular non-Hodgkin's lymphoma in situ |
| SNOMED CT | 1148851002 | Follicular lymphoma grade 3a |
| SNOMED CT | 1148890003 | Hemophagocytic lymphohistiocytosis due to malignant disease |
| SNOMED CT | 1148891004 | Bullous diffuse cutaneous mastocytosis |
| SNOMED CT | 1148906001 | Acute myeloid leukemia with t(8;21)(q22;q22) RUNX1-RUNX1T1 |
| SNOMED CT | 1149066008 | Chronic rheumatic pancarditis |
| SNOMED CT | 1149218009 | Isolated nailfold rheumatoid vasculitis |
| SNOMED CT | 1149219001 | Systemic rheumatoid vasculitis |
| SNOMED CT | 1149481006 | Sepsis caused by Coliform |
| SNOMED CT | 1149482004 | Sepsis caused by Clostridium |
| SNOMED CT | 1149486001 | Sepsis caused by Proteus |
| SNOMED CT | 1149494008 | Sepsis caused by Erysipelothrix rhusiopathiae |
| SNOMED CT | 1153336009 | Pediatric nodal marginal zone B cell lymphoma |
| SNOMED CT | 1153341001 | Myeloproliferative hypereosinophilic syndrome |
| SNOMED CT | 1153344009 | Pseudoxanthomatous nodular cutaneous mastocytosis |
| SNOMED CT | 1153345005 | Myelodysplastic syndrome with single lineage dysplasia |
| SNOMED CT | 1153346006 | Primary non-Hodgkin malignant lymphoma of uveal tract |
| SNOMED CT | 1153347002 | Primary non-Hodgkin malignant lymphoma of vitreoretinal tract |
| SNOMED CT | 1153348007 | Primary effusion lymphoma due to human immune deficiency virus infection |

Continued on next page

**Table 7 – continued from previous page**

| CodeSystem | ConceptCode | ConceptName |
|---|---|---|
| SNOMED CT | 1153349004 | Primary familial polycythemia due to erythropoietin receptor mutation |
| SNOMED CT | 1153354008 | Malignant lymphoma of duodenum |
| SNOMED CT | 1153355009 | Malignant lymphoma of esophagus |
| SNOMED CT | 1153356005 | Lymphoma of anus |
| SNOMED CT | 1153357001 | Lymphoma of appendix |
| SNOMED CT | 1153358006 | Lymphocyte passenger syndrome following organ transplantation |
| SNOMED CT | 1153374001 | Myelofibrosis caused by drug |
| SNOMED CT | 1153377008 | Mixed phenotype acute leukemia with t(9;22) (q34;q11.2); BCR-ABL1 |
| SNOMED CT | 1153379006 | Mixed phenotype acute leukemia with T-cell and myeloid lineage |
| SNOMED CT | 1153381008 | Mixed phenotype acute leukemia with myeloid and B-cell lymphoid phenotypes |
| SNOMED CT | 1153382001 | Lymphoma of lacrimal gland |
| SNOMED CT | 1153383006 | Lymphoma of liver |
| SNOMED CT | 1153397003 | Hydroa vacciniforme-like cutaneous T-cell lymphoma |
| SNOMED CT | 1153417000 | Secondary hemophagocytic lymphohistiocytosis |
| SNOMED CT | 1153534006 | Sepsis caused by vancomycin resistant Enterococcus |
| SNOMED CT | 1155919006 | Acute rheumatic aortitis |
| SNOMED CT | 1155943009 | Allergic contact dermatitis caused by metal and/or metal compound |
| SNOMED CT | 1155956002 | Follicular lymphoma grade 3a |
| SNOMED CT | 1155957006 | Follicular lymphoma grade 3b |
| SNOMED CT | 1156197003 | Complication of embryo transfer |
| SNOMED CT | 1156250009 | Hypereosinophilic syndrome |
| SNOMED CT | 1156296001 | Acquired neutropenia |
| SNOMED CT | 1156300000 | Benign ethnic neutropenia |
| SNOMED CT | 1156403002 | Composite Hodgkin and non-Hodgkin lymphoma |
| SNOMED CT | 1156495004 | Dendritic cell sarcoma |
| SNOMED CT | 1156631003 | Dendritic cell sarcoma |
| SNOMED CT | 1156746003 | Vaccine-induced prothrombotic immune thrombocytopenia |
| SNOMED CT | 1156765006 | Persistent onset antinuclear antibody negative juvenile idiopathic oligoarthritis |
| SNOMED CT | 1156766007 | Persistent onset antinuclear antibody positive juvenile idiopathic oligoarthritis |
| SNOMED CT | 1156774008 | Rheumatoid factor positive and anti-citrullinated protein antibody positive juvenile polyarthritis |
| SNOMED CT | 1156777001 | Rheumatoid factor positive and anti-citrullinated protein antibody negative juvenile polyarthritis |
| SNOMED CT | 1156778006 | Rheumatoid factor negative and anti-citrullinated protein antibody negative juvenile polyarthritis |

Continued on next page

**Table 7 – continued from previous page**

| CodeSystem | ConceptCode | ConceptName |
|---|---|---|
| SNOMED CT | 1156779003 | Rheumatoid factor negative and anti-citrullinated protein antibody positive juvenile polyarthritis |
| SNOMED CT | 1156801007 | Macrophage activation syndrome due to juvenile systemic onset arthritis |
| SNOMED CT | 1156817001 | Extended oligoarticular onset antinuclear antibody positive juvenile idiopathic arthritis |
| SNOMED CT | 1156819003 | Extended oligoarticular onset antinuclear antibody negative juvenile idiopathic arthritis |
| SNOMED CT | 1157157006 | Acute myeloid leukemia with 11q23 abnormality |
| SNOMED CT | 1157158001 | Acute myeloid leukemia with t(9;11)(p22;q23); MLLT3-MLL |
| SNOMED CT | 1157162007 | Intravascular large B-cell lymphoma |
| SNOMED CT | 15691081000119100 | Bilateral seronegative rheumatoid arthritis of feet |
| SNOMED CT | 15691121000119103 | Bilateral seronegative rheumatoid arthritis of elbow joints |
| SNOMED CT | 15691841000119102 | Bilateral seropositive rheumatoid arthritis of shoulder regions |
| SNOMED CT | 15691921000119100 | Bilateral seropositive rheumatoid arthritis of hip joints |
| SNOMED CT | 15691961000119105 | Bilateral seropositive rheumatoid arthritis of ankle joints |
| SNOMED CT | 15692001000119101 | Bilateral seropositive rheumatoid arthritis of joint of elbows |
| SNOMED CT | 15692041000119104 | Bilateral seropositive rheumatoid arthritis of wrist joint regions |
| SNOMED CT | 16606681000119101 | Seronegative rheumatoid arthritis in remission |
| SNOMED CT | 16697831000119103 | Idiopathic mast cell activation syndrome |
| SNOMED CT | 16775311000119107 | Mycosis fungoides of facial lymph node |
| SNOMED CT | 318841000119100 | Left ankle seropositive rheumatoid arthritis |
| SNOMED CT | 318851000119103 | Left elbow seropositive rheumatoid arthritis |
| SNOMED CT | 318891000119108 | Seropositive rheumatoid arthritis of joint of left shoulder region |
| SNOMED CT | 318901000119107 | Left wrist region seropositive rheumatoid arthritis |
| SNOMED CT | 318911000119105 | Right ankle seropositive rheumatoid arthritis |
| SNOMED CT | 318921000119103 | Right elbow seropositive rheumatoid arthritis |
| SNOMED CT | 318961000119108 | Seropositive rheumatoid arthritis of joint of right shoulder region |
| SNOMED CT | 318971000119102 | Right wrist region seropositive rheumatoid arthritis |
| SNOMED CT | 319011000119103 | Seronegative rheumatoid arthritis of joint of left ankle |
| SNOMED CT | 319041000119104 | Left hip joint seronegative rheumatoid arthritis |
| SNOMED CT | 319051000119102 | Seronegative rheumatoid arthritis of left knee joint |
| SNOMED CT | 319061000119100 | Left shoulder region seronegative rheumatoid arthritis |
| SNOMED CT | 319071000119106 | Left wrist region seronegative rheumatoid arthritis |
| SNOMED CT | 319091000119107 | Seronegative rheumatoid arthritis of joint of right ankle |
| SNOMED CT | 319121000119106 | Right hip joint seronegative rheumatoid arthritis |
| SNOMED CT | 319131000119109 | Seronegative rheumatoid arthritis of right knee joint |
| SNOMED CT | 319141000119100 | Right shoulder region seronegative rheumatoid arthritis |

Continued on next page

**Table 7 – continued from previous page**

| CodeSystem | ConceptCode | ConceptName |
|---|---|---|
| SNOMED CT | 319151000119103 | Right wrist region seronegative rheumatoid arthritis |
| SNOMED CT | 328371000119107 | Pancytopenia caused by antithyroid drug |
| SNOMED CT | 328381000119105 | Pancytopenia caused by anticonvulsant |
| SNOMED CT | 350691000119103 | Agranulocytosis due to and following administration of antineoplastic agent |
| SNOMED CT | 351251000119103 | Acute myelofibrosis in remission |
| SNOMED CT | 471141000124102 | Anaphylaxis caused by vaccine product containing Streptococcus pneumoniae antigen |
| SNOMED CT | 471311000124103 | Anaphylaxis caused by vaccine product containing Hepatitis A virus antigen |
| SNOMED CT | 471321000124106 | Anaphylaxis caused by vaccine product containing human poliovirus antigen |
| SNOMED CT | 471331000124109 | Anaphylaxis caused by vaccine product containing Measles morbillivirus and Mumps orthorubulavirus and Rubella virus antigens |
| SNOMED CT | 471341000124104 | Anaphylaxis caused by vaccine containing Human alphaherpesvirus 3 antigen |
| SNOMED CT | 471351000124102 | Anaphylaxis caused by vaccine product |
| SNOMED CT | 471361000124100 | Anaphylaxis caused by vaccine product containing Influenza virus antigen |
| SNOMED CT | 471371000124107 | Anaphylaxis caused by vaccine product containing human alphaherpesvirus 3 recombinant surface glycoprotein E antigen |
| SNOMED CT | 471381000124105 | Anaphylaxis caused by vaccine product containing live attenuated human alphaherpesvirus 3 antigen |
| SNOMED CT | 686981000119108 | Mycosis fungoides of inguinal lymph node |
| SNOMED CT | 686991000119106 | Mycosis fungoides of lower limb lymph node |
| SNOMED CT | 84821000119100 | Primary follicular dendritic cell sarcoma of spleen |
| SNOMED CT | 987840791000119102 | Adenosine deaminase 2 deficiency |
| Truveta | 1215935 | COVID-19 in immunocompromised patient |
| HCPCS | G0267 | BONE MARROW OR PERIPHERAL STEM CELL HARVEST ELIMINATE CELL TYPE(S) (E.G. T-CELLS |
| HCPCS | S2140 | Cord blood harvesting for transplantation, allogeneic |
| HCPCS | S2142 | Cord blood-derived stem-cell transplantation, allogeneic |
| SNOMED CT | 1163005009 | Hodgkin lymphoma |
| SNOMED CT | 1162677006 | Rheumatoid arthritis with rheumatoid lung disease |
| SNOMED CT | 1217476001 | Anaphylactic reaction due to component of vaccine product |
| SNOMED CT | 139841000119108 | Anaphylaxis caused by allergy skin test |
| SNOMED CT | 15919571000119107 | Anaphylaxis caused by antibacterial agent |
| SNOMED CT | 1228843008 | Anti-p200 pemphigoid |
| SNOMED CT | 1162336001 | Anti-citrullinated protein antibody positive erosive rheumatoid arthritis |

Continued on next page

**Table 7 – continued from previous page**

| CodeSystem | ConceptCode | ConceptName |
|---|---|---|
| SNOMED CT | 1177173001 | Congenital progressive bone marrow failure, B-cell immunodeficiency, skeletal dysplasia syndrome |
| SNOMED CT | 1186855002 | Autoimmune hypopituitarism |
| SNOMED CT | 1186859008 | Autoimmune male infertility |
| SNOMED CT | 1162928000 | Acute myeloid leukemia |
| SNOMED CT | 1162531007 | Acute myeloid leukemia in complete remission |
| SNOMED CT | 1172892009 | Autosomal recessive mendelian susceptibility to mycobacterial disease due to complete RORgamma receptor mutation |
| SNOMED CT | 1179288008 | Combined immunodeficiency due to TFRC deficiency |
| SNOMED CT | 1162660006 | B lymphoblastic leukemia lymphoma |
| SNOMED CT | 1186720006 | CEBPE-associated autoinflammation, immunodeficiency, neutrophil dysfunction syndrome |
| SNOMED CT | 1179300002 | BENTA disease |
| SNOMED CT | 1172730009 | Breast implant-associated anaplastic large cell lymphoma |
| SNOMED CT | 1177120001 | Bronchiolitis obliterans syndrome due to and following allogeneic stem cell transplant |
| SNOMED CT | 1186712009 | Combined immunodeficiency due to CARMIL2 deficiency |
| SNOMED CT | 1186715006 | Combined immunodeficiency due to CD70 deficiency |
| SNOMED CT | 1179286007 | Combined immunodeficiency due to GINS1 deficiency |
| SNOMED CT | 1186714005 | Combined immunodeficiency due to ITK deficiency |
| SNOMED CT | 1179285006 | Combined immunodeficiency due to moesin deficiency |
| SNOMED CT | 1197732001 | Colorectal Crohn disease |
| SNOMED CT | 1197205005 | Combined immunodeficiency due to DOCK8 deficiency |
| SNOMED CT | 1197477000 | Combined immunodeficiency due to LRBA deficiency |
| SNOMED CT | 1197428008 | Combined immunodeficiency, enteropathy spectrum |
| SNOMED CT | 515623061000119105 | Crohn disease of colon in remission |
| SNOMED CT | 527788091000119106 | Crohn disease of small intestine and large intestine in remission |
| SNOMED CT | 253855721000119107 | Crohn disease of small intestine in remission |
| SNOMED CT | 1197727002 | Crohn's disease of parastomal skin |
| SNOMED CT | 1230290005 | Cytokine release syndrome due to chimeric antigen receptor T-cell immunotherapy |
| SNOMED CT | 1231239003 | Dacryoadenitis due to Graves' disease |
| SNOMED CT | 1197479002 | DOCK2 deficiency |
| SNOMED CT | 1204463009 | Encephalopathy with sepsis |
| SNOMED CT | 1222550009 | Enteropathy-associated T-cell lymphoma |
| SNOMED CT | 1220590003 | Familial chilblain lupus erythematosus |
| SNOMED CT | 1204466001 | Gastritis with sepsis |
| SNOMED CT | 1204464003 | Gastrointestinal hemorrhage with sepsis |
| SNOMED CT | 1204461006 | Gastrointestinal lesion with sepsis |
| SNOMED CT | 1204467005 | Gastrointestinal ulceration due to sepsis |
| SNOMED CT | 1230342001 | Hepatitis B reinfection following liver transplantation |
| SNOMED CT | 1197733006 | Ileojejunal Crohn disease |

Continued on next page

**Table 7 – continued from previous page**

| CodeSystem | ConceptCode | ConceptName |
|---|---|---|
| SNOMED CT | 1230414002 | Immune effector cell-associated neurotoxicity syndrome |
| SNOMED CT | 145781000119106 | Infection of transplanted kidney |
| SNOMED CT | 1204462004 | Left ventricular failure with sepsis |
| SNOMED CT | 1204459002 | Lung injury with sepsis |
| SNOMED CT | 15988711000119101 | Perennial allergic conjunctivitis of left eye |
| SNOMED CT | 15988751000119100 | Perennial allergic conjunctivitis of right eye |
| SNOMED CT | 1230291009 | Primary biliary cholangitis and/or primary sclerosing cholangitis and autoimmune hepatitis overlap syndrome |
| SNOMED CT | 1228842003 | Primary cutaneous plasmacytosis |
| SNOMED CT | 1197478005 | Primary immunodeficiency with multifaceted aberrant lymphoid immunity |
| SNOMED CT | 1197150002 | Recurrent hepatitis C virus induced liver disease following liver transplant |
| SNOMED CT | 1204468000 | Right ventricular failure with sepsis |
| SNOMED CT | 1201760007 | Sepsis caused by Bacteroides |
| SNOMED CT | 1290841000000104 | Sepsis caused by monkeypox virus |
| SNOMED CT | 1204037006 | Sepsis following induced termination of pregnancy |
| SNOMED CT | 1230323000 | Sepsis of premature infant |
| SNOMED CT | 1229942009 | Severe combined immunodeficiency due to CORO1A deficiency |
| SNOMED CT | 1197415001 | Susceptibility to infection due to TYK2 deficiency |
| SNOMED CT | 1197482007 | Susceptibility to localized juvenile periodontitis |
| SNOMED CT | 1197701002 | Type 1 autoimmune pancreatitis with extrapancreatic involvement |
| SNOMED CT | 1197704005 | Autoantibody negative autoimmune hepatitis |
| SNOMED CT | 1208833001 | Autoimmune generalized polymyoclonus |
| SNOMED CT | 1222681008 | Autoimmune hemolytic anemia, autoimmune thrombocytopenia, primary immunodeficiency syndrome |
| SNOMED CT | 1208830003 | Autoimmune interstitial keratitis |
| SNOMED CT | 1222679006 | Autoimmune interstitial lung disease, arthritis syndrome |
| SNOMED CT | 1197361002 | Autoimmune lymphoproliferative syndrome due to CTLA4 haploinsufficiency |
| SNOMED CT | 1197740007 | Autoimmune pancreatitis type 2 |
| SNOMED CT | 1208829008 | Autoimmune retinitis |
| SNOMED CT | 1197366007 | Autosomal recessive primary immunodeficiency with defective spontaneous natural killer cell cytotoxicity |
| SNOMED CT | 1229941002 | Autosomal recessive T-cell negative, B-cell positive severe combined immunodeficiency due to CD45 deficiency |
| SNOMED CT | 1229940001 | Autosomal recessive T-cell negative, B-cell positive severe combined immunodeficiency due to IL-7Ralpha deficiency |
| SNOMED CT | 1196900003 | B cell lymphoma of small intestine |
| SNOMED CT | 1230295000 | B-cell immunodeficiency, limb anomaly, urogenital malformation syndrome |

Continued on next page

**Table 7 – continued from previous page**

| CodeSystem | ConceptCode | ConceptName |
|---|---|---|
| SNOMED CT | 1172709000 | Lymphoid leukemia |
| SNOMED CT | 1162263002 | Complement component 8 deficiency |
| SNOMED CT | 1163528001 | Erythema multiforme due to bacterial infection |
| SNOMED CT | 1187129009 | Primary cutaneous B-cell lymphoma |
| SNOMED CT | 1187190002 | Primary cutaneous diffuse large B-cell lymphoma leg type |
| SNOMED CT | 1187135009 | Primary cutaneous large T-cell lymphoma |
| SNOMED CT | 1187136005 | Primary cutaneous T-cell lymphoma |
| SNOMED CT | 1163590002 | Myelomonocytic leukemia |
| SNOMED CT | 1163439000 | Myeloid leukemia |
| SNOMED CT | 16058751000119103 | Bilateral transplant failure of corneas |
| SNOMED CT | 1162588009 | Chronic myeloid leukemia |
| SNOMED CT | 1163404000 | Peripheral T-cell lymphoma |
| SNOMED CT | 1173999006 | IL21-related infantile inflammatory bowel disease |
| SNOMED CT | 1162973007 | Cutaneous T-cell lymphoma |
| SNOMED CT | 1162993004 | Acute eosinophilic leukemia |
| SNOMED CT | 1186652002 | Inflammatory bowel disease, recurrent sinopulmonary infection syndrome |
| SNOMED CT | 1187123005 | Mixed phenotype acute leukemia |
| SNOMED CT | 1172704005 | High grade B-cell lymphoma with MYC and BCL2 and/or BCL6 rearrangements |
| SNOMED CT | 1172516002 | High-dose chemotherapy with stem cell transplant |
| SNOMED CT | 1186654001 | Immune dysregulation, inflammatory bowel disease, arthritis, recurrent infection, lymphopenia syndrome |
| SNOMED CT | 1187508009 | Immunoglobulin G4 related aortitis |
| SNOMED CT | 1186834004 | Immunoglobulin G4 related hypophysitis |
| SNOMED CT | 1187507004 | Immunoglobulin G4 related kidney disease |
| SNOMED CT | 1187510006 | Immunoglobulin G4 related ophthalmic disease |
| SNOMED CT | 1186721005 | Infantile inflammatory bowel disease with neurological involvement |
| SNOMED CT | 1187000005 | Infectious mononucleosis caused by human immunodeficiency virus |
| SNOMED CT | 1187001009 | Infectious mononucleosis caused by human immunodeficiency virus type I |
| SNOMED CT | 1162768007 | Leukemia |
| SNOMED CT | 1187233008 | Leukocyte adhesion deficiency |
| SNOMED CT | 1169361002 | Limbic encephalitis with neurexin-3 antibodies |
| SNOMED CT | 1186646003 | Malignant histiocytic neoplasm |
| SNOMED CT | 1163043007 | Malignant lymphoma |
| SNOMED CT | 1162264008 | Malignant lymphoma of uveal tract |
| SNOMED CT | 1172895006 | Mendelian susceptibility to mycobacterial disease |
| SNOMED CT | 1172592001 | Non-Hodgkin lymphoma |
| SNOMED CT | 1186728004 | Pediatric multiple sclerosis |

Continued on next page

**Table 7 – continued from previous page**

| CodeSystem | ConceptCode | ConceptName |
|---|---|---|
| SNOMED CT | 1187623009 | Phosphoglucomutase 3-related congenital disorder of glycosylation |
| SNOMED CT | 1162576007 | Plasma cell myeloma |
| SNOMED CT | 1187107005 | Plasmacytoma |
| SNOMED CT | 1186719000 | Predisposition to invasive fungal disease due to CARD9 deficiency |
| SNOMED CT | 1172686000 | Protein contact dermatitis |
| SNOMED CT | 1162319009 | Rheumatoid factor and anti-citrullinated protein antibody positive erosive rheumatoid arthritis |
| SNOMED CT | 1162313005 | Rheumatoid factor and anti-citrullinated protein antibody positive rheumatoid arthritis |
| SNOMED CT | 1162303002 | Seronegative erosive rheumatoid arthritis |
| SNOMED CT | 1179284005 | Severe combined immunodeficiency due to LAT deficiency |
| SNOMED CT | 1177175008 | Skeletal dysplasia, T-cell immunodeficiency, developmental delay syndrome |
| SNOMED CT | 1186725001 | Warts, immunodeficiency, lymphedema, anogenital dysplasia syndrome |
| SNOMED CT | 1162828001 | X-linked lymphoproliferative disease due to SH2D1A deficiency |
| SNOMED CT | 1162830004 | X-linked lymphoproliferative disease due to XIAP deficiency |
| SNOMED CT | 1187193000 | Hairy cell leukemia variant |
| SNOMED CT | 1162505005 | Agranulocytosis caused by antithyroid agent |
| SNOMED CT | 1172729004 | ALK negative anaplastic large cell lymphoma |
| SNOMED CT | 15919911000119108 | Allergic reaction caused by analgesic |
| SNOMED CT | 15919991000119104 | Allergic reaction caused by anesthetic |
| SNOMED CT | 15920641000119103 | Allergic reaction caused by food additive |
| SNOMED CT | 15920241000119101 | Allergic reaction caused by fruit |
| SNOMED CT | 15920441000119100 | Allergic reaction caused by seed |
| SNOMED CT | 1172702009 | Anaplastic diffuse large B-cell lymphoma |
| SNOMED CT | 1172695008 | Diffuse large B cell lymphoma |
| SNOMED CT | 1172701002 | Facial diplegia with paresthesia |
| SNOMED CT | 1162362005 | Felty syndrome with seronegative erosive rheumatoid arthritis |
| SNOMED CT | 1162364006 | Felty syndrome with seronegative rheumatoid arthritis |
| SNOMED CT | 1197476009 | Infantile-onset pulmonary alveolar proteinosis, hypogammaglobulinemia |
| SNOMED CT | 1197594000 | Periodic fever, infantile enterocolitis, autoinflammatory syndrome |
| SNOMED CT | 1197344000 | T-cell lymphoma of small intestine |
| SNOMED CT | 1204420006 | Chronic nonbacterial osteomyelitis/chronic recurrent multifocal osteomyelitis |
| SNOMED CT | 1217669004 | Chronic papillary vernal conjunctivitis |
| SNOMED CT | 1197362009 | STAT3-related early-onset multisystem autoimmune disease |

Continued on next page

**Table 7 – continued from previous page**

| CodeSystem | ConceptCode | ConceptName |
|---|---|---|
| SNOMED CT | 353211000119103 | Extramedullary plasmacytoma in remission |
| SNOMED CT | 16058791000119108 | Bilateral corneal transplant rejection |

### 4 Counts of patients by comorbidity and outcome type

#### 4.1 Breakthrough COVID-19 infection or not

| ckd | diabetes | immunocompromised | lung | covid | count |
|---|---|---|---|---|---|
| 0 | 0 | 0 | 0 | 0 | 745949 |
| 0 | 0 | 0 | 0 | 1 | 10229 |
| 0 | 0 | 0 | 1 | 0 | 42900 |
| 0 | 0 | 0 | 1 | 1 | 1221 |
| 0 | 0 | 1 | 0 | 0 | 165684 |
| 0 | 0 | 1 | 0 | 1 | 4096 |
| 0 | 0 | 1 | 1 | 0 | 49639 |
| 0 | 0 | 1 | 1 | 1 | 1888 |
| 0 | 1 | 0 | 0 | 0 | 51687 |
| 0 | 1 | 0 | 0 | 1 | 1081 |
| 0 | 1 | 0 | 1 | 0 | 8092 |
| 0 | 1 | 0 | 1 | 1 | 253 |
| 0 | 1 | 1 | 0 | 0 | 27421 |
| 0 | 1 | 1 | 0 | 1 | 835 |
| 0 | 1 | 1 | 1 | 0 | 12394 |
| 0 | 1 | 1 | 1 | 1 | 548 |
| 1 | 0 | 0 | 0 | 0 | 12110 |
| 1 | 0 | 0 | 0 | 1 | 204 |
| 1 | 0 | 0 | 1 | 0 | 3093 |
| 1 | 0 | 0 | 1 | 1 | 69 |
| 1 | 0 | 1 | 0 | 0 | 13377 |
| 1 | 0 | 1 | 0 | 1 | 391 |
| 1 | 0 | 1 | 1 | 0 | 7162 |
| 1 | 0 | 1 | 1 | 1 | 294 |
| 1 | 1 | 0 | 0 | 0 | 9496 |
| 1 | 1 | 0 | 0 | 1 | 175 |
| 1 | 1 | 0 | 1 | 0 | 3001 |
| 1 | 1 | 0 | 1 | 1 | 106 |
| 1 | 1 | 1 | 0 | 0 | 10312 |
| 1 | 1 | 1 | 0 | 1 | 336 |
| 1 | 1 | 1 | 1 | 0 | 7731 |
| 1 | 1 | 1 | 1 | 1 | 361 |

Table 8: Counts of patients by comorbidity and breakthrough COVID-19 infection outcome status. 0 indicates absence of comorbidity or no breakthrough infection. 1 indicates present of comorbidity or breakthrough infeciton.

#### 4.2 Hospitalized following breakthrough COVID-19 infection or not

| ckd | diabetes | immunocompromised | lung | hospital | count |
|---|---|---|---|---|---|
| 0 | 0 | 0 | 0 | 0 | 14424 |
| 0 | 0 | 0 | 0 | 1 | 1778 |
| 0 | 0 | 0 | 1 | 0 | 1351 |
| 0 | 0 | 0 | 1 | 1 | 161 |
| 0 | 0 | 1 | 0 | 0 | 403 |
| 0 | 0 | 1 | 0 | 1 | 140 |
| 0 | 0 | 1 | 1 | 0 | 81 |
| 0 | 0 | 1 | 1 | 1 | 27 |
| 0 | 1 | 0 | 0 | 0 | 1819 |
| 0 | 1 | 0 | 0 | 1 | 462 |
| 0 | 1 | 0 | 1 | 0 | 303 |
| 0 | 1 | 0 | 1 | 1 | 73 |
| 0 | 1 | 1 | 0 | 0 | 89 |
| 0 | 1 | 1 | 0 | 1 | 44 |
| 0 | 1 | 1 | 1 | 0 | 19 |
| 0 | 1 | 1 | 1 | 1 | 15 |
| 1 | 0 | 0 | 0 | 0 | 451 |
| 1 | 0 | 0 | 0 | 1 | 238 |
| 1 | 0 | 0 | 1 | 0 | 74 |
| 1 | 0 | 0 | 1 | 1 | 64 |
| 1 | 0 | 1 | 0 | 0 | 63 |
| 1 | 0 | 1 | 0 | 1 | 56 |
| 1 | 0 | 1 | 1 | 0 | 23 |
| 1 | 0 | 1 | 1 | 1 | 15 |
| 1 | 1 | 0 | 0 | 0 | 344 |
| 1 | 1 | 0 | 0 | 1 | 330 |
| 1 | 1 | 0 | 1 | 0 | 84 |
| 1 | 1 | 0 | 1 | 1 | 57 |
| 1 | 1 | 1 | 0 | 0 | 54 |
| 1 | 1 | 1 | 0 | 1 | 94 |
| 1 | 1 | 1 | 1 | 0 | 15 |
| 1 | 1 | 1 | 1 | 1 | 23 |

Table 9: Counts of patients by comorbidity and hospitalization following breakthrough COVID-19 infection outcome status. 0 indicates absence of comorbidity or not hospitalized. 1 indicates present of comorbidity or hospitalized.

### 5 Model parameter estimates

#### 5.1 Cox regression model of time till breakthrough COVID-19 infection

| term | estimate | std.error | statistic | p.value | conf.low | conf.high |
|---|---|---|---|---|---|---|
| ckd | 1.57 | 0.07 | 6.49 | 0.00 | 1.37 | 1.80 |
| diabetes | 1.67 | 0.03 | 15.89 | 0.00 | 1.57 | 1.78 |
| immunocompromised | 1.86 | 0.02 | 33.13 | 0.00 | 1.80 | 1.93 |
| lung | 1.92 | 0.03 | 21.53 | 0.00 | 1.81 | 2.04 |
| ns(age_years, df = 5)1 | 0.89 | 0.04 | -3.22 | 0.00 | 0.83 | 0.96 |
| ns(age_years, df = 5)2 | 0.81 | 0.05 | -4.55 | 0.00 | 0.75 | 0.89 |
| ns(age_years, df = 5)3 | 0.51 | 0.05 | -13.58 | 0.00 | 0.47 | 0.56 |
| ns(age_years, df = 5)4 | 0.70 | 0.09 | -4.06 | 0.00 | 0.59 | 0.83 |
| ns(age_years, df = 5)5 | 0.54 | 0.09 | -7.05 | 0.00 | 0.45 | 0.64 |
| raceUnknown | 0.88 | 0.02 | -5.04 | 0.00 | 0.84 | 0.93 |
| raceBlack or African American | 0.99 | 0.02 | -0.60 | 0.55 | 0.95 | 1.03 |
| raceAsian | 0.77 | 0.04 | -7.03 | 0.00 | 0.72 | 0.83 |
| raceAmerican Indian or Alaska Native | 1.07 | 0.09 | 0.75 | 0.45 | 0.89 | 1.29 |
| raceNative Hawaiian or Other Pacific Islander | 0.91 | 0.12 | -0.77 | 0.44 | 0.72 | 1.16 |
| ethnicityHispanic or Latino | 1.33 | 0.02 | 12.33 | 0.00 | 1.27 | 1.39 |
| ethnicityUnknown | 0.83 | 0.04 | -5.11 | 0.00 | 0.77 | 0.89 |
| sexMale | 0.96 | 0.01 | -3.00 | 0.00 | 0.93 | 0.99 |
| month_year2021-02-01 | 0.87 | 0.08 | -1.80 | 0.07 | 0.75 | 1.01 |
| month_year2021-03-01 | 0.77 | 0.07 | -3.54 | 0.00 | 0.67 | 0.89 |
| month_year2021-04-01 | 0.92 | 0.07 | -1.20 | 0.23 | 0.80 | 1.06 |
| month_year2021-05-01 | 0.84 | 0.07 | -2.46 | 0.01 | 0.73 | 0.96 |
| month_year2021-06-01 | 0.93 | 0.08 | -0.95 | 0.34 | 0.80 | 1.08 |
| month_year2021-07-01 | 2.00 | 0.08 | 9.04 | 0.00 | 1.72 | 2.32 |
| month_year2021-08-01 | 3.14 | 0.08 | 14.95 | 0.00 | 2.71 | 3.65 |
| month_year2021-09-01 | 3.61 | 0.07 | 17.35 | 0.00 | 3.12 | 4.17 |
| month_year2021-10-01 | 3.46 | 0.07 | 16.68 | 0.00 | 2.99 | 4.01 |
| month_year2021-11-01 | 3.19 | 0.08 | 14.64 | 0.00 | 2.73 | 3.73 |
| month_year2021-12-01 | 3.21 | 0.08 | 13.93 | 0.00 | 2.73 | 3.79 |
| month_year2022-01-01 | 2.46 | 0.09 | 9.65 | 0.00 | 2.05 | 2.96 |
| month_year2022-02-01 | 1.93 | 0.11 | 6.07 | 0.00 | 1.56 | 2.38 |
| month_year2022-03-01 | 2.59 | 0.11 | 8.43 | 0.00 | 2.08 | 3.24 |
| ckd:diabetes | 0.73 | 0.10 | -3.27 | 0.00 | 0.61 | 0.88 |
| ckd:immunocompromised | 1.03 | 0.08 | 0.39 | 0.70 | 0.88 | 1.21 |
| ckd:lung | 0.85 | 0.11 | -1.42 | 0.16 | 0.69 | 1.06 |
| diabetes:immunocompromised | 0.84 | 0.05 | -3.60 | 0.00 | 0.76 | 0.92 |
| diabetes:lung | 0.81 | 0.07 | -3.02 | 0.00 | 0.70 | 0.93 |
| immunocompromised:lung | 0.78 | 0.04 | -6.13 | 0.00 | 0.72 | 0.84 |
| ckd:diabetes:immunocompromised | 1.02 | 0.11 | 0.15 | 0.88 | 0.82 | 1.26 |
| ckd:diabetes:lung | 1.24 | 0.11 | 2.03 | 0.04 | 1.01 | 1.52 |
| ckd:immunocompromised:lung | 1.02 | 0.12 | 0.20 | 0.84 | 0.81 | 1.29 |
| diabetes:immunocompromised:lung | 1.10 | 0.09 | 1.12 | 0.26 | 0.93 | 1.31 |

Table 10: Summary of the selected Cox proportaional hazards model of time till breakthrough infection showing estimates for all of the regression coefficients. Estimates are on the hazard scale (i.e. exponentiated).

#### 5.2 Logistic regression model of hospitalization following breakthrough COVID-19 infection

| term | estimate | std.error | statistic | p.value | conf.low | conf.high |
|---|---|---|---|---|---|---|
| (Intercept) | 0.04 | 0.28 | -11.47 | 0.00 | 0.02 | 0.07 |
| ns(age_years, df = 5)1 | 0.79 | 0.15 | -1.54 | 0.12 | 0.59 | 1.07 |
| ns(age_years, df = 5)2 | 2.32 | 0.17 | 4.93 | 0.00 | 1.67 | 3.25 |
| ns(age_years, df = 5)3 | 2.04 | 0.13 | 5.42 | 0.00 | 1.58 | 2.64 |
| ns(age_years, df = 5)4 | 30.22 | 0.34 | 10.16 | 0.00 | 15.81 | 58.94 |
| ns(age_years, df = 5)5 | 13.63 | 0.18 | 14.58 | 0.00 | 9.60 | 19.39 |
| ckd | 2.41 | 0.06 | 14.99 | 0.00 | 2.15 | 2.71 |
| diabetes | 1.67 | 0.05 | 10.27 | 0.00 | 1.51 | 1.84 |
| immunocompromised | 1.59 | 0.04 | 10.61 | 0.00 | 1.46 | 1.74 |
| lung | 1.70 | 0.05 | 11.55 | 0.00 | 1.55 | 1.86 |
| raceUnknown | 1.16 | 0.08 | 1.91 | 0.06 | 1.00 | 1.35 |
| raceBlack or African American | 1.06 | 0.07 | 0.89 | 0.37 | 0.93 | 1.20 |
| raceAsian | 1.01 | 0.12 | 0.06 | 0.96 | 0.79 | 1.27 |
| raceAmerican Indian or Alaska Native | 1.63 | 0.26 | 1.90 | 0.06 | 0.96 | 2.64 |
| raceNative Hawaiian or Other Pacific Islander | 0.62 | 0.48 | -0.99 | 0.32 | 0.21 | 1.44 |
| ethnicityHispanic or Latino | 1.18 | 0.07 | 2.33 | 0.02 | 1.03 | 1.36 |
| ethnicityUnknown | 0.54 | 0.14 | -4.19 | 0.00 | 0.41 | 0.72 |
| sexMale | 1.04 | 0.04 | 0.90 | 0.37 | 0.96 | 1.13 |
| month_year2021-02-01 | 1.12 | 0.26 | 0.42 | 0.67 | 0.69 | 1.91 |
| month_year2021-03-01 | 0.84 | 0.25 | -0.70 | 0.48 | 0.52 | 1.42 |
| month_year2021-04-01 | 0.88 | 0.25 | -0.49 | 0.62 | 0.56 | 1.49 |
| month_year2021-05-01 | 0.90 | 0.25 | -0.43 | 0.66 | 0.56 | 1.51 |
| month_year2021-06-01 | 1.16 | 0.26 | 0.57 | 0.57 | 0.72 | 1.98 |
| month_year2021-07-01 | 0.95 | 0.26 | -0.20 | 0.84 | 0.58 | 1.63 |
| month_year2021-08-01 | 0.96 | 0.26 | -0.15 | 0.88 | 0.59 | 1.66 |
| month_year2021-09-01 | 0.95 | 0.26 | -0.20 | 0.84 | 0.59 | 1.62 |
| month_year2021-10-01 | 1.19 | 0.26 | 0.67 | 0.50 | 0.74 | 2.02 |
| month_year2021-11-01 | 1.15 | 0.27 | 0.51 | 0.61 | 0.69 | 1.99 |
| month_year2021-12-01 | 1.16 | 0.28 | 0.53 | 0.60 | 0.68 | 2.07 |
| month_year2022-01-01 | 1.04 | 0.31 | 0.14 | 0.89 | 0.57 | 1.95 |
| month_year2022-02-01 | 1.11 | 0.35 | 0.30 | 0.77 | 0.56 | 2.21 |
| month_year2022-03-01 | 1.03 | 0.35 | 0.08 | 0.94 | 0.52 | 2.07 |

Table 11: Summary of the selected logistic regression model of hospitalization following breakthrough infection showing estimates for all of the regression coefficients. Estimates are on the odds scale (i.e. exponentiated).
